# Independent Polygenic Component Scores Link Multivariate Brain Imaging Genetics to Diverse Phenotypes

**DOI:** 10.1101/2025.06.06.25329112

**Authors:** Lennart M. Oblong, Sourena Soheili-Nezhad, Nicolò Trevisan, Yingjie Shi, Christian F. Beckmann, Emma Sprooten

**Affiliations:** Department of Medical Neuroscience, Donders Institute for Brain, Cognition and Behaviour, Radboud University Medical Centre, Nijmegen, The Netherlands; Department of Human Genetics, Donders Institute for Brain, Cognition and Behaviour, Radboud University Medical Centre, Nijmegen, The Netherlands; Centre for Cognitive Neuroimaging, Donders Institute for Brain, Cognition and Behaviour, Radboud University, Nijmegen, The Netherlands

**Author notes:** Corresponding authors: Lennart M. Oblong & Emma Sprooten.

**Keywords:** Genomic ICA, Independent Component Analysis, MELODIC, neuroimaging, GWAS, Multivariate Genomics, Statistical Genetics, Neuroimaging Genetics, Polygenic Scores

## Abstract

Complex traits are influenced by many, often overlapping genetic influences, which makes it difficult to separate shared from trait-specific genetic mechanisms. We previously introduced Genomic Independent Component Analysis (genomICA), a novel, data-driven, multivariate method to disentangle genetic effects across thousands of univariate GWAS statistics into statistically independent genomic components (i.e. latent factors). Each component captures patterns of genetic covariance across traits. Our goals in this paper are threefold: To validate the capability of this new method to reliably recover genetic effects across thousands of complex phenotypes; to map shared and distinct genetic architectures across brain features and their relations with other complex traits; and to provide a new framework for polygenic scoring that improves out-of-sample prediction, interpretability, and stratification of individuals along independent genetic dimensions.

Using the SNP-loadings from our previously derived genomICA components, derived from thousands of brain imaging GWASs, we generated polygenic component scores (PCS) in an independent cohort. We first evaluated mutual independence among PCS, as a condition for stratification utility. Next, we tested their out-of-sample predictive performance and specificity for 1269 neuroimaging and 858 behavioural, clinical, and lifestyle phenotypes. Additionally, we tested whether association patterns with brain traits recapitulated the original trait-loadings, as a cross-validation.

Correlations between PCSs were low (|*r*| < 0.05), confirming mutual independence. PCSs explained substantial variance in brain traits (*R*^2^*_max_* =0.12). For behavioural/clinical/lifestyle phenotypes (not included in the discovery data) explained variance was also significant, but much lower (*R*^2^*_max_* =0.02). Individual PCSs were associated with distinct groups of neuroimaging categories and brain tissues. PCSs showed meaningful patterns of associations with clinical/behavioural/lifestyle phenotypes. For instance, PCS 8 captured associations with lifestyle, behavior, diet, and socioeconomic factors, while PCS 15 was linked to cardiovascular health outcomes. The dominant genomic influences captured here predicted physical, cognitive, lifestyle and environmental phenotypes, but surprisingly not mental health or neurological diagnoses.

In the present study we present a novel polygenic scoring approach based on multivariate independent genomICA components. Each PCS reflects a reliable, distinct individual disposition towards a combination of brain-features. Individual PCSs align with distinct phenotypic domains. Overall, our results indicate that PCSs aid prediction and stratification utility of high-dimensional GWAS statistics.

## Introduction

Many complex human traits are heritable ^1–11^ and shaped by pleiotropic and polygenic effects ^10,12–15^. Genome-wide association studies (GWAS) show that individual effects of common single nucleotide polymorphisms (SNPs) are weak, and even after aggregation across hundreds of thousands of SNPs, they explain a relatively small portion of trait variance (0.019 < *R*^2^ < 0.147) ^16–22^. This complexity compounds the difficulty of mechanistic interpretation of GWASes^10,13,15,23^. A common way to predict genetic predisposition to multifactorial traits is via polygenic scores (PGS) ^24–28^. Polygenicity and pleiotropy propagate to PGS analyses because many different biological processes underlying univariate SNP associations are aggregated into a single score. The unresolved mixture of pleiotropic effects means that standard PGS may have reduced specificity for biologically adjacent phenotypes and for describing etiological heterogeneity (e.g. biological subtypes of patients). Ideally, a PGS-like score that accounts for pleiotropy would be desirable to uncover specific genetic profiles that predispose to various subtypes of a trait, and their subserving genetic correlates ^29^. The present paper seeks to address this issue by validating a method that exploits the shared underlying genetic signal across many traits and recovers shared and distinct genetic sources.

By leveraging pleiotropy across traits, the genetic covariance structure across many traits can be estimated and used to model latent factors reflecting these shared genetic associations across traits. Using GWAS data, several multivariate methods have been proposed to achieve this goal, including genomic structural equation modeling (SEM) ^30^, genomic Principal Component Analysis (PCA) ^31^, the Multivariate Omnibus Statistical Test (MOSTest) ^32^, and Genetic Unmixing by Independent Decomposition (GUIDE) ^33^. Genomic SEM requires manual specification of model order (i.e. number of latent factors) and pre-specifying which traits map onto which latent factors, and the computational cost of genomic SEM becomes prohibitive as more traits are explored ^30^. MOSTest combines SNP statistics across traits in a single multi-trait genome-wide association, which limits the method’s power to recover multiple distinct genomic patterns across traits. Genomic PCA is entirely data-driven and designed to decompose genetic covariance matrices into components (i.e. latent factors) that capture uncorrelated axes explaining maximum variance ^31^. However, multiple dominant and correlated biological signals may be aggregated into the first component that captures the most data variance. Furthermore, all subsequent components are enforced to be orthogonal in PCA, which does not necessarily reflect how biological processes interact.

Statistically *independent* components offer an interpretable, data-driven approach to investigate complex genetic architecture of multifactorial phenotypes ^33,34^. We developed *genomic Independent Component Analysis* (genomICA) to untangle genetic associations (i.e. independent *signals*) from GWAS summary statistics across thousands of traits into few genomic components ^34^. This method treats GWAS output as a ‘mixture’ of biological signal sources within and across GWASs, reflecting how SNPs may be involved in the same or in different biological pathways. Multivariate ICA approaches untangle these mixed signals into distinct, statistically *independent* genetic signal sources while simultaneously filtering out random noise ^35^. Each component (i.e. factor) is associated with two vectors, one containing the SNP-loadings and the other containing the trait-loadings. The SNP-loadings constitute the independent genomic sources and can be used as a more disentangled “pseudo-GWAS summary statistic” linked to brain phenotypes encoded in trait-loadings.

In a large set of GWASs we can recover multiple independent genetic components, each representing a distinct pattern of shared SNP effects across traits. High trait loadings into the same component indicate shared genetic influences, revealing groups of traits linked by common underlying biological pathways. This addresses a central limitation of classical PGS that collapses genome-wide effects into a single score for each trait, obscuring pleiotropy. Each genomICA component captures a uniquely polygenic axis of pleiotropy, allowing for the calculation of pairwise independent polygenic component scores (PCS). PCS enable genetic architectures of many traits to be partitioned into multiple, more interpretable axes of genetic predisposition.

In the present exploratory study, we used the genomic components as the summary-statistics-like discovery data to calculate a PCS for each out-of-sample participant. The components we previously generated based on neuroimaging phenotypes may reflect patterns of pleiotropy that are not limited to brain traits alone ^34,36,37^. For example, these components may reflect patterns linked to brain-relevant traits outside of the brain and even in the environment. We seek to test the potential of genomICA components to capture individual-level phenotypic variance out-of-sample, and to yield interpretable patterns of association across a wide range of phenotypes indicating their future potential for stratification of heterogeneous samples. First, we tested out-of-sample PCS’s predictive power for neuroimaging phenotypes as validation. Then, we tested PCSs on hundreds of non-neuroimaging phenotypes that include general physical attributes, nutritional questionnaire data, physical health, mental health, general well-being, lifestyle, behaviour, socio-economic, and demographic metrics. In this way, we investigate the potential for GenomICA to increase sensitivity of polygenic prediction and provide insights into shared biological pathways and their links to environmental influences.

## Results

### Polygenic Component Scores of multivariate genomic independent and principal components are orthogonal

Pearson-correlation between all PCSs showed that PCSs derived respectively from genomic ICs and PCs were largely uncorrelated, validating that genomic patterns subserving their calculation are likewise independent at the individual-level out-of-sample (PGS-PC1 from genomic IC 1-16: *r_min_* = −0.04, *r_mean_* = 4e-4, *r_max_* = 0.05; PGS-PC 1 from genomic PC1-16: *r_min_* = −0.04, *r_mean_* = 5e-4, *r_max_* = 0.04; figure S1). Correlations between the IC-PCSs and PC-PCSs were high, which is expected given that the ICs are a rotation of the PCs and both sets of components capture variance in the same data space. Given this low collinearity, we performed all subsequent analyses using models containing all 16 PCSs simultaneously as predictors for each trait.

### Neuroimaging-derived polygenic component scores are associations with neuroimaging phenotypes in an independent sample

We assessed the performance of PCSs on the same brain imaging phenotypes that were included in the GenomICA pipeline, in the independent target sample. GenomICA derived PCSs captured statistically significant variance in N = 1256 imaging derived phenotypes post-FDR (*p_FDR_* < 0.05). Polygenic variance explained varied across neuroimaging categories (figure S2). Neuroimaging QC metrics (*R*^2^*_mean_* = 0.09), dMRI TBSS metrics (*R*^2^*_mean_* = 0.07), and dMRI ProbtrackX metrics (*R*^2^*_mean_* = 0.07) showed the highest average phenotypic variance explained. For individual neuroimaging traits, the highest (conventional) polygenic variance was captured in global brain features such as *T1-imaging scan to standard-space warping*, a proxy for intracranial volume (ICV), large white-matter tract *orientational discrepancy* (OD) *of the posterior limb of the internal capsule in both hemispheres*, and *global normalized cerebrospinal fluid* (CSF) volume derived from T1-weighted SIENAX tissue segmentation (table 1). Overall, the highest *R*^2^-values were achieved in metrics related to large white matter tracts with half of the top 20 IDPs related to diffusion neuroimaging values which generally reflect tissue density (table 1).

**Table 1:** Table showing the top 20 imaging derived phenotypes predicted by PCSs in concert, sorted by Nagelkerke’s pseudo-*R*^2^. Full.*R*^2^ refers to the variance explained by the full model. Null.R refers to the variance explained by the null model. PCS.*R*^2^ indicates the variance explained by the PCSs only, while PCS.*R*^2^.adj denotes the proportional or adjusted variance explained (see methods section). The p.value columns shows the FDR-corrected p-value. Abbreviations: CSF – cerebrospinal fluid, dMRI – diffusion magnetic resonance imaging, FA – fractional anisotropy, ICVF – intracellular volume fraction, ifo – inferior fronto-occipital fasciculus, ISOVF – isotropic volume fraction, L1-L3 – diffusion directions of the 3D diffusion tensor, MD – mean diffusivity, OD – orientation dispersion, Probtrackx – probabilistic tractography, slf – superior longitudinal fasciculus, TBSS – tract-based spatial statistics, unc – uncinate fasciculus, QC – quality control, L or l & R or r – left and right brain hemisphere.

| UKB.ID | Trait | Full-$R^2$ | Null-$R^2$ | PCS-$R^2$ | PCS-$R^2$.adj | p.value | Modality |
| --- | --- | --- | --- | --- | --- | --- | --- |
| 25733 | T1-to-standard nonlinear alignment warping | 0.27 | 0.149 | 0.12 | 0.141 | 2.54e-72 | QC |
| 25411 | OD Posterior limb of internal capsule L | 0.168 | 0.0517 | 0.116 | 0.122 | 1.28e-60 | dMRI-TBSS |
| 25003 | CSF normalised volume | 0.406 | 0.292 | 0.114 | 0.161 | 1.55e-84 | T1-SIENAX-Volume |
| 25410 | OD Posterior limb of internal capsule R | 0.169 | 0.0556 | 0.114 | 0.121 | 7.02e-60 | dMRI-TBSS |
| 25365 | ICVF Retrolenticular part of internal capsule L | 0.207 | 0.097 | 0.11 | 0.122 | 1.85e-60 | dMRI-TBSS |
| 25481 | ISOVF Superior longitudinal fasciculus L | 0.213 | 0.105 | 0.108 | 0.121 | 7.02e-60 | dMRI-TBSS |
| 25671 | ICVF slf l | 0.238 | 0.13 | 0.108 | 0.124 | 1.02e-61 | dMRI-Probtrackx |
| 25581 | L1 ifo l | 0.216 | 0.11 | 0.107 | 0.12 | 1.96e-59 | dMRI-Probtrackx |
| 25337 | L3 Superior longitudinal fasciculus L | 0.248 | 0.142 | 0.107 | 0.124 | 1.02e-61 | dMRI-TBSS |
| 25662 | ICVF ifo l | 0.221 | 0.115 | 0.105 | 0.119 | 4.52e-59 | dMRI-Probtrackx |
| 25725 | ISOVF slf l | 0.207 | 0.102 | 0.105 | 0.117 | 1.09e-57 | dMRI-Probtrackx |
| 25408 | OD Anterior limb of internal capsule R | 0.162 | 0.0579 | 0.104 | 0.11 | 4.52e-54 | dMRI-TBSS |
| 25385 | ICVF Superior longitudinal fasciculus L | 0.23 | 0.127 | 0.103 | 0.118 | 1.93e-58 | dMRI-TBSS |
| 25672 | ICVF slf r | 0.223 | 0.121 | 0.102 | 0.116 | 1.87e-57 | dMRI-Probtrackx |
| 25500 | FA ifo l | 0.182 | 0.0799 | 0.102 | 0.111 | 2.71e-54 | dMRI-Probtrackx |
| 25527 | MD ifo l | 0.229 | 0.128 | 0.101 | 0.116 | 3.69e-57 | dMRI-Probtrackx |
| 25417 | OD Superior corona radiata L | 0.18 | 0.079 | 0.101 | 0.109 | 1.97e-53 | dMRI-TBSS |
| 25145 | MD Superior longitudinal fasciculus L | 0.269 | 0.169 | 0.1 | 0.121 | 7.02e-60 | dMRI-TBSS |
| 25364 | ICVF Retrolenticular part of internal capsule R | 0.239 | 0.139 | 0.1 | 0.116 | 2.6e-57 | dMRI-TBSS |
| 25730 | ISOVF unc r | 0.162 | 0.0622 | 0.0998 | 0.106 | 8.8e-52 | dMRI-Probtrackx |

As a further confirmation of the out-of-sample reproducibility of the method, we found that PCS effect sizes on neuroimaging phenotypes in the target sample were highly correlated with the neuroimaging trait-loadings in the discovery sample (0.58 < *r* < 0.92; table S1).

Since PCS-*R*^2^ is bounded by SNP-*h*^2^, we also calculated the *R*^2^*_conventional_* as a fraction of the SNP-heritability (*h^2^_SNP_*). For some phenotypes, such as the *T1-weighted image to standard-space warping* PCSs collectively explain a large fraction of the heritability (*R*^2^*_SNP_* / *h*^2^*_SNP_* = 0.82). Across all neuroimaging phenotypes, the average *h*^2^-fraction explained by PCS was 0.17. Similar results for other neuroimaging phenotypes are presented in full in table S2 and figure S5.

Results for the genomic PCA derived PCS analysis were similar in *R*^2^-magnitude as those for the genomic ICA PCS and can be found in the supplement (figure S6 & S7; table S3).

### Neuroimaging-based polygenic component scores associated with non-neuroimaging phenotypes

Overall, many non-neuroimaging phenotypes were significantly associated with neuroimaging-derived PCSs, but effect sizes (*R*^2^) were much lower than for the neuroimaging phenotypes themselves.

Looking at broader domain categories, the largest variance explained were in phenotypes in the domains of *physical health, lifestyle and behavior*, and numeric memory in the domain of cognitive performance (figure S3). Individual binary phenotypes most strongly associated with PCS were *self-reported Nephritis, and self-reported Malabsorption/Coeliac disease*, followed by *frequency of memory loss due to drinking alcohol in the past year* and frequency of feeling remorse after drinking alcohol in the past year (table 2). Weaker in effect size but also associated with significance were measures of physical attributes, such as the comparative height at age 10, and physical health measures related to cardiovascular health such as *self-reported high blood-pressure, diagnosed high blood pressure, and no diagnosis of any cardiovascular health problems*. The full results are shown in the supplement (table S4).

**Table 2:** Table showing the top 20 binary/binarized non-imaging phenotypes predicted by PCSs in concert, sorted by Nagelkerke’s *R*^2^. Phenotype indicates the binarized phenotype that was tested in the model, while base.Phenotype denotes the phenotype that the binarized phenotype was derived from. Full.*R*^2^ refers to the variance explained by the full model. Null.*R*^2^ refers to the variance explained by the null model. PCS.Nag.*R*^2^ indicates the variance explained by the PCSs only. The p.value column shows the FDR-corrected p-value.

| *Phenotype* | *base.Phenotype* | *PCS.Nag.R<sup>2</sup>* | *full.R<sup>2</sup>* | *Null.R<sup>2</sup>* | *p.value* |
| --- | --- | --- | --- | --- | --- |
| self-reported Nephritis | Non-cancer illness code, self-reported | 0.022 | 0.113 | 0.09 | 0.014 |
| self-reported Malabsorption/Coeliac disease | Non-cancer illness code, self-reported | 0.011 | 0.04 | 0.029 | 1.2e-27 |
| Frequency of memory loss due to drinking alcohol in last year 2 vs 3 | Frequency of memory loss due to drinking alcohol in last year | 0.0096 | 0.021 | 0.012 | 0.046 |
| Frequency of feeling guilt or remorse after drinking alcohol in last year 2 vs 4 | Frequency of feeling guilt or remorse after drinking alcohol in last year | 0.0077 | 0.022 | 0.015 | 0.042 |
| Numeric memory test - Maximum digits remembered correctly 5 vs 10 | Numeric memory test - Maximum digits remembered correctly | 0.0073 | 0.08 | 0.073 | 0.034 |
| Numeric memory test - Maximum digits remembered correctly 5 vs 9 | Numeric memory test - Maximum digits remembered correctly | 0.0051 | 0.089 | 0.084 | 0.0041 |
| Financial situation satisfaction 1 vs 6 | Financial situation satisfaction | 0.005 | 0.224 | 0.219 | 0.04 |
| Frequency of memory loss due to drinking alcohol in last year 1 vs 3 | Frequency of memory loss due to drinking alcohol in last year | 0.0045 | 0.026 | 0.021 | 0.035 |
| Happiness.0 1 vs 4 | Happiness | 0.0044 | 0.091 | 0.087 | 0.044 |
| Comparative height size at age 10 1 vs 3 | Comparative height size at age 10 | 0.004 | 0.013 | 0.0086 | 9.5e-84 |
| Numeric memory test - Maximum digits remembered correctly 5 vs 8 | Numeric memory test - Maximum digits remembered correctly | 0.0033 | 0.051 | 0.048 | 0.0021 |
| Financial situation satisfaction 1 vs 4 | Financial situation satisfaction | 0.003 | 0.125 | 0.122 | 0.0042 |
| Numeric memory test - Maximum digits remembered correctly 6 vs 9 | Numeric memory test - Maximum digits remembered correctly | 0.0028 | 0.042 | 0.039 | 0.0088 |
| Oily fish intake 0 vs 4 | Oily fish intake | 0.0028 | 0.051 | 0.048 | 0.018 |
| Happiness and subjective well-being - General happiness with own health 1 vs 4 | Happiness and subjective well-being - General happiness with own health | 0.0026 | 0.124 | 0.121 | 0.023 |
| Amount of alcohol drunk on a typical drinking day.0 1 vs 5 | Amount of alcohol drunk on a typical drinking day | 0.002 | 0.213 | 0.211 | 0.043 |
| Traumatic events - Able to pay rent/mortgage as an adult 2 vs 4 | Traumatic events - Able to pay rent/mortgage as an adult | 0.0019 | 0.039 | 0.037 | 0.022 |
| Skin colour 1 vs 4 | Skin colour | 0.0018 | 0.086 | 0.084 | 0.047 |
| self-reported Hypertension | Non-cancer illness code, self-reported | 0.0017 | 0.138 | 0.136 | 1,00E-62 |
| Relative age of first facial hair male 1 vs 3 | Relative age of first facial hair male | 0.0017 | 0.061 | 0.06 | 0.027 |

In continuous phenotypes the strongest associations were likewise found in physical health and physical attributes, including *height, weight, basal metabolic rate*, and systolic- and diastolic blood-pressure (figure S4; table S6).

We calculated the *h*^2^-fraction explained by PCSs for all tested phenotypes and found lower ratios than those in neuroimaging phenotypes, with an average *h*^2^-fraction of 0.01 (*h*^2^-fraction*_max_* = 0.28) in binary and binarized phenotypes (table S4), and 0.004 in continuous phenotypes (*h*^2^-fraction = 0.02) (table S6).

In the analysis of genomic PCA derived PCSs we found similar associations in that the strongest domains of association are those of physical health, lifestyle and behaviour, cognitive performance and physical attributes. In the following sections we focus on the results of genomic ICA PCSs considering constraints on manuscript length. The full results of this genomic PCA analysis are presented in the supplement (table S5 & S7; figure S7 & S8).

### Overall patterns of phenotypic associations emerge across polygenic component scores

Next, we interrogated which individual PCSs drove the associations with individual phenotypes. We found striking patterns of different phenotypes driven by few PCSs in the model, such that some phenotypes were associated with specific genetic patterns captured by distinct genomic ICs. Other phenotypes associated with multiple PCSs suggesting a widespread polygenic influence linked to genetically distinct brain features. The full results of genomic ICs (figures S13 – S44 & S77 – S86) and PCs (figures S45 – S76 & S87 – S96) are presented in the supplement and are published on a publicly available website (genomica.info). Given the large number of phenotypes, each paired with 16 ICs and 16 PCs, we only highlight several global observations.

Many PCSs are significantly associated with a wide range of neuroimaging features while fewer PCSs clearly drive the associations with specific white matter tracts or regions of interest. Diffusion-weighted white matter metrics are significantly predicted by all PCSs (figure S2), but different components were associated with different white matter tracts, sometimes very distinctly. For example, PCS7 was strongly associated with the Fornix and superior *fronto-occipital fasciculu*s (figure S83), while PCS2 with the anterior thalamic radiation (figure S83). Subcortical volumes were similarly predicted with a high degree of specificity by PCS 10 markedly driving associations with whole brainstem volume, Pons, Midbrain and Medulla (figure S80); while PCS 7 predicted multiple specific thalamic nuclei to the exclusion of other PCSs (figure S80). Notably, the entire category of cortical thickness phenotypes was mainly associated with two specific components: PCS 3 and PCS 8 (figure S78). Overall, our analyses showed that individual PCSs captured mutually independent genetic signals, with a degree of specificity for tissues, following known neuroanatomical boundaries. All results can be viewed in the supplement (figure S77 – S86).

In non-neuroimaging space, associations of PCSs with physical attributes such as *height, weight, fat-mass, fat-free mass*, and metabolic rate were pervasive across PCSs. Widespread associations with cardiovascular health metrics likewise stood out. Hypertension in particular was significantly associated with 13 out of 16 PCSs Most hypertension-associated PCSs are additionally, and reliably in the same direction, associated with other cardiovascular conditions such as Angina, heart attack or stroke (1, 5, 6, 7, 9, 11, 15, 16), and with heart disease of mother, father or siblings (4, 5, 6, 7, 9, 10, 12, 15, 16). Also, markedly stable across components is *high Cholesterol*, which is only significantly associated in PCSs also associated with cardiovascular health outcomes, and reliably in the same direction (1, 5, 7, 11, 12, 14, 15). Please refer to the supplement table S4 and figures S13 – S44 for full statistics and figures.

Several other multifactorial clinical diagnoses are significantly predicted by PCSs. These diagnoses are Nephritis (11, 12, 16), Osteoporosis (4, 9, 12), Osteoarthritis (1, 2, 3), Malabsorption/Coeliac disease (5, 6, 8, 14, 15), Hypothyroidism/Myxoedema (2, 5, 6, 11, 13), Asthma (4, 6, 13), chronic Bronchitis (4, 12, 13), and Diabetes (4, 5, 10).

Further, multiple PCSs link to lifestyle and behavior, including dietary metrics that reliably co-associate with cardiovascular health outcomes. For instance, PCS (3, 4, 5, 6, 7, 11, 12, 13) show inverse patterns of correlations between cardiovascular health risk and the intake of healthy fats, as oily fish intake. Other lifestyle metrics, including smoking, alcohol intake and physical activity, co-associate across several component PCSs with cardiovascular health outcomes in the expected direction (e.g. less smoking, less alcohol intake, more activity correlated with better cardiovascular health) (1, 6, 9, 10, 11, 13, 14). Few PCSs show unexpected associations, where associations with e.g. positive cardiovascular health outcomes co-associate with increased levels of smoking or increased alcohol intake (4, 12, 15).

PCSs further significantly associate with several socioeconomic measures such as average household income (1, 2, 4, 7, 8, 9, 11, 12, 13, 15), financial satisfaction (1, 2, 11, 12, 13, 14), owning or renting accommodations (8, 11, 14), and the number of vehicles owned (8). As expected, components associated with higher average household income also co-associate with *higher financial satisfaction, owning rather* than renting a house/apartment, and the number of vehicles owned. PCSs displaying an association with higher average household income consistently co-associate with an increased overall health rating (1, 8, 9, 11, 13).

In the category of well-being most of the significant associations were identified with overall health rating (1, 5, 6, 8, 9, 11, 13, 14, 15, 16), followed by general happiness with own health (1, 5, 9, 14). Happiness was significantly predicted by PCS 2 and 13 but did not reflect a clear pattern with other health ratings, nor a clear pattern with other metrics in this category. Other mental health outcome metrics that were significantly associated with PCSs are seen *general practitioner* (GP) for nerves, anxiety, tension or depression (2, 5, 6), and seen psychiatrist for nerves, anxiety, tension or depression (2, 5, 8). No significant associations were found for mental health or neurological diagnostic variables included in the analysis, such as Dementia, Anxiety, Depression, or Alzheimer’s Disease.

### Representative examples of selected individual genomic component polygenic scores

Here we present more detailed results for 2 components for illustration. We made our selection based on interpretability and consistency of the component PGS associations across phenotypes, which does not discount the interpretability and level of insight gained from other components. Please refer to the supplementary material for the full results of the other components (table S2 – S7) or consult our website at genomica.info for an interactive format.

PCS 15 displayed a clear pattern of association with cardiovascular features (figure 1). This PCS was positively highly significantly associated with not being diagnosed with cardiovascular problems and negatively associated with diagnosed high blood pressure. Furthermore, PCS 15 significantly predicted angina, stroke, heart attack, diabetes, and high cholesterol, and with their occurrence in family members of participants. Positive associations that fit this general theme were found in an increased overall health rating predicted by this PCS. This trend was also reflected in the analyses of continuous non-neuroimaging phenotypes where the PCS was negatively associated with traits in the domains of physical health and physical attributes (figure S9).

**Figure 1:**
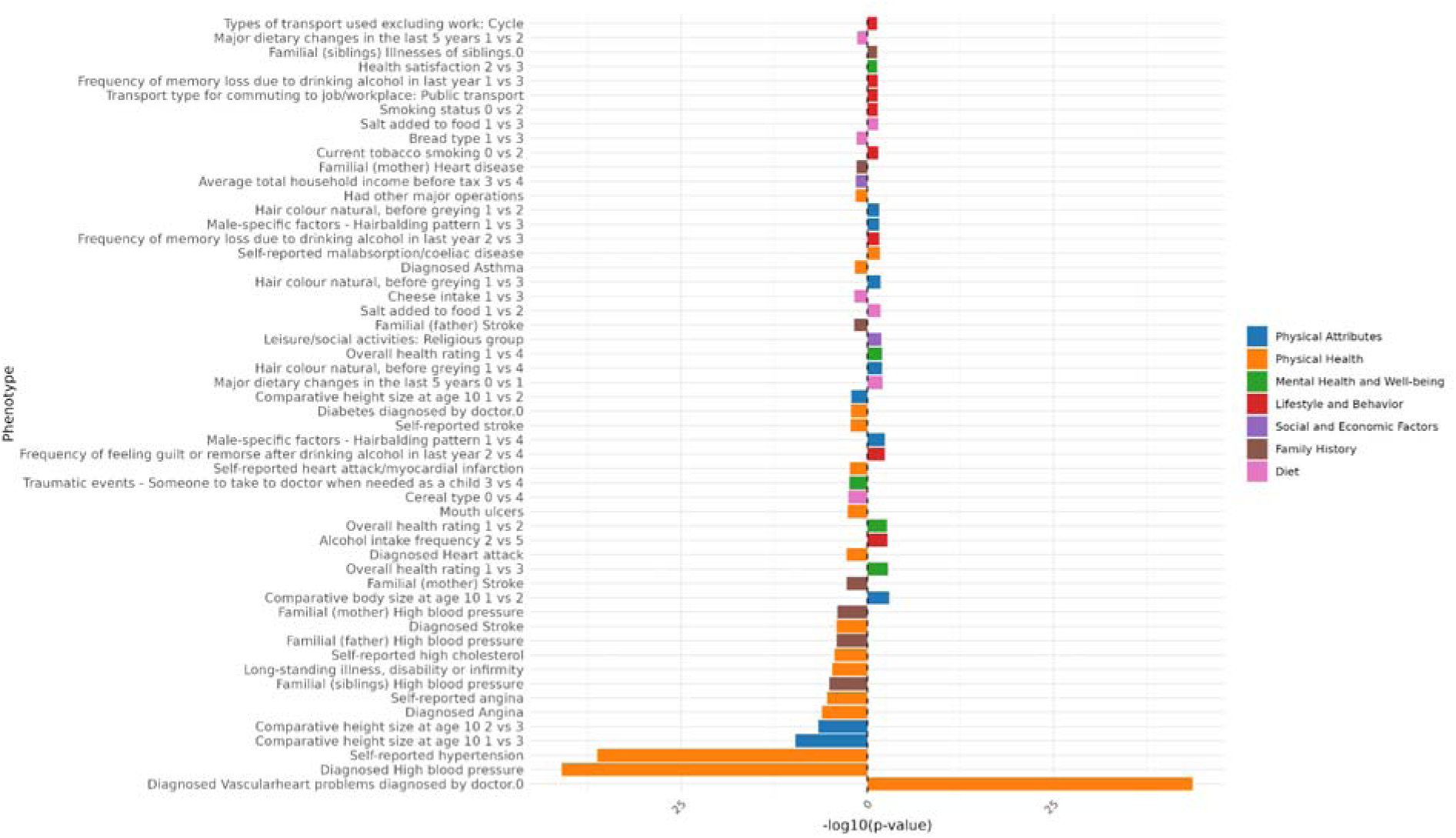
Overview of PCS15 significant associations with binary/binarized non-imaging phenotypes. The x-axis shows the -log(10) p-value. The direction of the bar indicates the direction of association, in this case the sign of the log-odds ratio. Right facing bars show a positive association, left facing bars a negative association. In binarized ordinal metrics the direction of the bar indicates the same, but the contrast is constructed in such a way that the lower number (e.g. 1 vs 3) is the reference. Phenotypes are binned into domains which are indicated in a color scheme contained in the legend on the right-hand side of the figure. “not being diagnosed with cardiovascular problems” is indicated by “Diagnosed Vascularheart problems diagnosed by doctor.0”.

In neuroimaging space PCS 15 drives associations with multiple dMRI derived metrics of isotropic water fraction (ISOVF) around the superior and inferior longitudinal fasciculi (figure S10, table S2). PCS15 uniquely captured variance in this neuroimaging phenotype with high significance, contrasting with other PCSs that drive associations with different dMRI phenotypes and white matter tracts (such as PCS7 driving associations in the Fornix).

Another interesting PCS to highlight is PCS 8, which was not associated with any cardiovascular health metrics in *UKB participants*, but was significantly associated with several behaviour and lifestyle, and social and environmental phenotypes. We found that this PCS co-associated with a wide range of dietary choices. PCS 8 showed significant associations with eating no sugar, tending towards whole grain bread and cereal, less salt added to food, and indicating a major dietary shift within the last 5 years (figure 2). Furthermore, we found significant association with less smoking across multiple metrics, and an increased usual walking pace. This is combined with associations indicating an increased average household income, increased financial satisfaction, and a higher overall health rating. Conversely, this PCS also associated with increased levels of alcohol intake in both frequency and past alcohol consumption, and les deliberate physical activity in the last four weeks. Interestingly, we also found a positive association with maternal smoking around birth, and a negative association with being breastfed as a baby (figure 2). For continuous non-imaging metrics, the bulk of significant associations was identified in lower height, weight and several impedance measures, but also an increased impedance of the whole body and increased body fat percentage (figure S11). Furthermore, there were significant associations with spending more time watching television, but also with spending more time outside in both summer and winter. Overall, this PCS drive dietary and lifestyle associations to the exclusion of other metrics of physical health, indicating a pleiotropic neuroimaging-based pattern of genetic effects that act in this domain.

**Figure 2:**
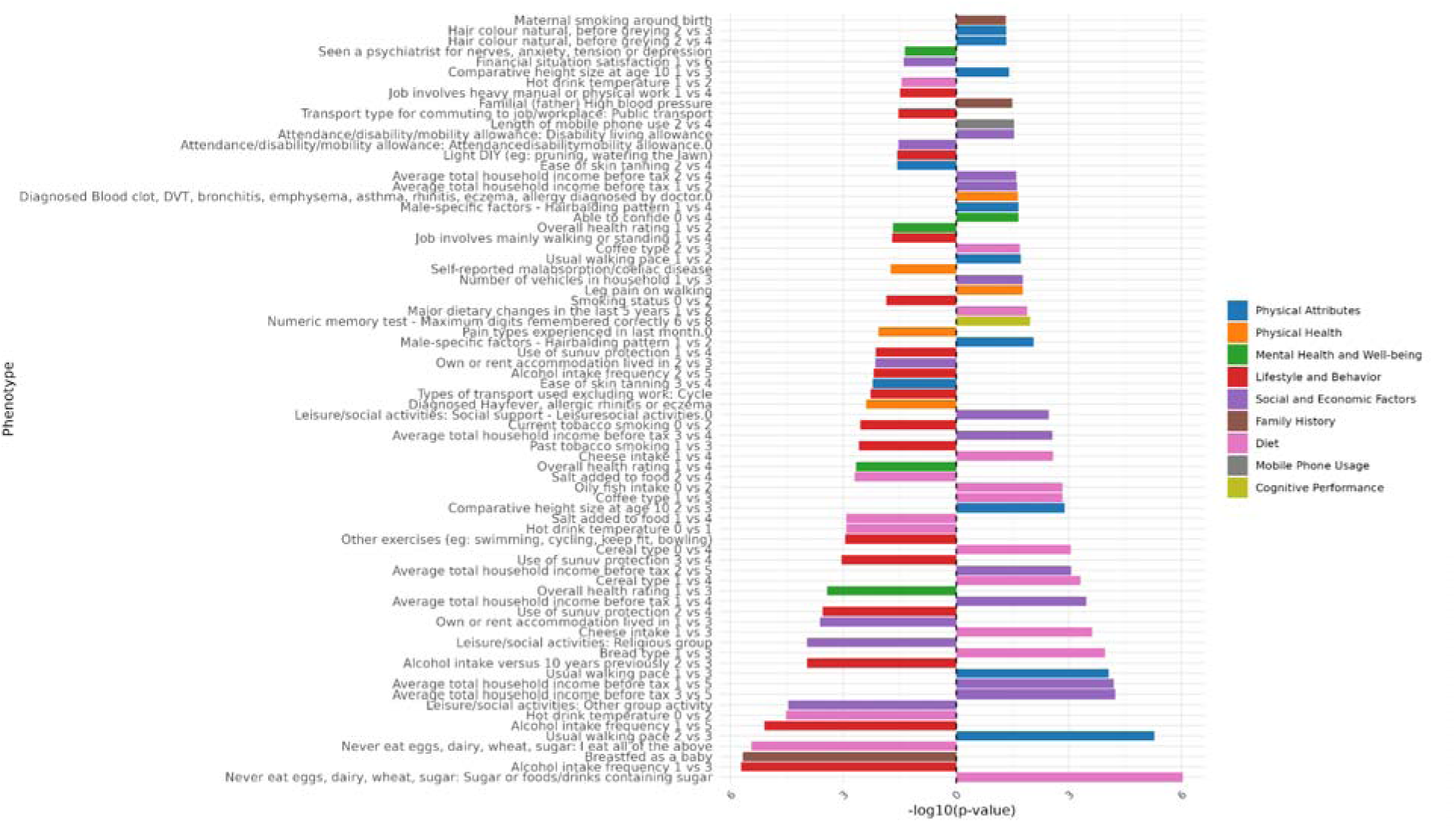
Overview of PCS8 significant associations with binary/binarized non-imaging phenotypes. The x-axis shows the -log(10) p-value. The direction of the bar indicates the direction of association, in this case the sign of the log-odds ratio. Right facing bars show a positive association, left facing bars a negative association. In binarized ordinal metrics the direction of the bar indicates the same, but the contrast is constructed in such a way that the lower number (e.g. 1 vs 3) is the reference. Phenotypes are binned into domains which are indicated in a color scheme contained in the legend on the right-hand side of the figure.

In neuroimaging space PCS 8 captured phenotypic variance in the domain of cortical thicknes and drives associations across a wide range of brain imaging phenotypes across hemisphere together with PCS 3 (figure S78). In other neuroimaging categories this PCS is notably inconspicuous, except for cortical and subcortical volumes, and intensity, where it contributed to, but is not the main contributor of them (figure S12, figures S79-S81). Taken together, this suggests that IC 8 captured polygenic effects chiefly related with cortical thickness that seem to link to many *dietary, lifestyle and behaviour*, and environmental variables.

## Discussion

In this study we demonstrate the utility of genomICA components as a basis for polygenic scores in an independent sample. We decomposed genome-wide brain-wide GWAS summary statistics into 16 components that represent latent patterns of genetic associations with a diverse range of brain phenotypes. We calculated component-based polygenic scores in each individual of an independent sample and used them to predict a range of neuroimaging and other phenotypes.

PCSs were not correlated with each other, confirming that polygenic scores derived from genomICA reflect independent and multivariate genetic effects. PCSs predicted out-of-sample neuroimaging phenotypes with substantial phenotypic variance explained, up to 12%. In many phenotypes this constituted a large portion of the SNP-heritability, up to 82%. As cross-validation, our PCS effects on brain traits correlated very highly with the trait-loadings of the decomposition in the discovery sample, reflecting out-of-sample reliability. Many PCSs showed high specificity for distinct white matter tracts, brain regions, and tissues.

Neuroimaging phenotype-based PCSs significantly predicted a wide range of phenotypes across non-neuroimaging variables albeit with much smaller effect sizes compared to brain traits. Many PCSs displayed internally consistent patterns of domain-specific associations, thus providing construct validity of our genomICA approach. One of these domains includes physical attributes such as height, weight, and impedance measures. Further, several PCSs showed cross-domain co-associations fitting with a common, interpretable pleiotropic pattern (e.g. *cardiovascular health* and *cholesterol*). This suggests that genomICA components capture independent genetic signals, grouping individual variant effects in meaningful ways. This points to potential future applications that identify how distinct PCS profiles relate to variation in phenotypic outcomes across individuals

Compelling co-associations between cardiovascular health outcomes, cholesterol, and socio-economic and lifestyle metrics were dominant across PCSs. High cholesterol is a known risk factor for heart disease ^38^, and the presented PCSs show stable, directly proportional associations with cardiovascular health outcomes. This suggests a common, heritable genetic link between cholesterol levels and heart disease risk. This is illustrated by the inverse correlation with oily fish intake, which is rich in unsaturated fats, and lowers cholesterol and cardiovascular risk ^39^. Across PCSs, we find consistent inverse correlations between oily fish intake, high cholesterol, and cardiovascular health outcomes. Cholesterol is critical for white matter myelination ^40^ and the brain contains 15% - 25% of body cholesterol ^41^. The PCSs associated with high cholesterol also show strong associations with white matter features, where hyperintensities have been linked to stroke ^42^. Additionally, hypertension, a risk factor for white matter lesions ^43^, further highlights a link between cardiovascular health and white matter integrity. This is compounded by co-association patterns with smoking behaviour, which is a risk factor for cardiovascular health problems and was inversely associated across several PCSs ^44^. Thus, genomICA applied to brain measures captures common genetic variation related to cardiovascular health. This could be a reflection for the presence of cardiovascular risk factors and their impact on the brain, in the middle-aged UK Biobank population, while more subtle or rarer gene-brain covariation patterns are not picked up by our analysis in this sample.

In the domain of social and environmental variables PCS associations also displayed marked internal consistency. Multiple socioeconomic variables were consistently co-associated (e.g. average household income, financial satisfaction, owning rather than renting homes, an increased number of vehicles), and they were linked with well-being measures of overall health ratings, a connection that has been widely accepted in social study literature ^45–47^. This suggests that brain-based pleiotropic patterns captured across components consistently reflect an environmental link between genetic signal associated with brain phenotypes, socioeconomic metrics and overall health rating.

PCS8 stood out through its unique association pattern, which did not include cardiovascular features and was instead driven by diet, lifestyle and behaviour, and social and environmental variables. The high consistency towards metrics from these domains, combined with the wide range of associations in the imaging space, suggests that this component captures strong gene-environment correlations that act on brain function and structure^23^.

In separate work, our group applied bioinformatic analyses using FUMA ^48^ and MAGMA ^49^ to the same components ^50^. This work indicated that genomICA components capture an array of biological processes. IC8 showed enrichment for zinc/calcium signaling (SLC39A8, DOC2A) and redox balance (MSRB3), combined with developmental regulation (PPP4C, YPEL3). This is compounded by IC8 trait associations matching genetic variants previously associated with diet, alcohol intake and smoking ^50^. IC15 captured genetic signal related to white matter organisation through immune and lipid metabolic pathways ^50^. Particularly a dominant locus in the HLA region stood out among other loci involved in immune functioning/neuroinflammation, and myelination, which have been implicated to impact brain white matter phenotypes ^51^ and cardiovascular health outcomes ^52^ respectively. Furthermore, matching trait associations of the original IC15 include healthy fat intake, cholesterol metrics, diabetes type 2, and blood pressure metrics ^50^ (genomica.info). Both examples show a consistency in our interpretations, whether derived from bioinformatics ^50^, or from out-of-sample PCS associations shown in this paper.

When comparing outcomes of PCSs derived from genomic ICs and genomic PCs, differences are minor. Both approaches explain similar magnitudes of variance across neuroimaging and non-neuroimaging domains, and each yields components driving associations with specific neuroimaging categories (table S2 & S3). While genomic ICs capture maximally independent genetic patterns, genomic PCs capture orthogonal axes of maximum variance. Theoretically, higher genetic specificity of ICA is expected, because maximizing non-Gaussianity typically increases kurtosis, hence a more concentrated signal. Qualitatively, for example ICA indeed identified a separate component of lifestyle and sociodemographic that did not include cardiovascular signal (IC8), while PCA did not separate cardiovascular and dietary and sociodemographic factors in terms of their genetic underpinnings.

The dominant genetic influences on individual variation in brain features appear to relate mostly to physical characteristics—particularly body shape, height, and cardiovascular traits— and, to a lesser extent, to environmental and lifestyle factors. In contrast, genetic overlap with neurological or psychiatric diagnoses, as well as with broader mental health traits, appears minimal. This suggests that departing from neuroimaging genomics may not be the optimal strategy for uncovering the genetic mechanisms underlying mental or *neurological phenotypes*. One notable exception was “*ever visited a GP for nerves, anxiety, tension or depression*”, previously included in assessing “*broad depression*”^22^, which did show some associations. By expanding the decomposition to include - beyond neuroimaging traits - environmental, sociodemographic, lifestyle, and mental health related discovery GWASs in the future, we hope to improve the sensitivity of PCS analysis to capture genetic covariance between brain phenotypes, environment and mental health outcomes simultaneously.

There are some limitations to our work. Our approach is limited by GWAS summary statistic discovery sample size. The present data allows for reproducible decomposition of ∼40% of the variance, into 16 stable, multivariate ICs^34^. The remaining ∼61% of signal remains unaccounted for in components, as previously shown it was too noisy to capture reliably with the present data^34^. Furthermore, even within the 39% explained variance, the multitude of genetically driven mechanisms influencing brain phenotypes is unlikely to be captured in only 16 dimensions. Each component is likely to reflect a combination of different underlying mechanistic sources. In the future, with increasing (neuroimaging) GWAS samples sizes, a larger number of components may be reliably extracted. Another limitation is that the input GWAS summary statistics themselves are observational data that inherently limit causal inferences. With these limitations in mind, the interpretation of the presented multivariate PCS analysis is tentative. Nevertheless, the presented PCS associations show clear patterns of co-association within and across components that are supported by existing literature and gene-enrichment analysis of the components ^50^.

In conclusion, our novel GWAS summary statistic decomposition using genomICA has yielded data-driven, mutually uncorrelated polygenic scores that reliably associate with individual differences in a wide range of physical and behavioural phenotypes out-of-sample. From the present analysis clear patterns of associations emerge that are supported by literature, particularly between cardiovascular health outcomes and lifestyle, and socioeconomic factors and overall health ratings. Overall, these results strengthen the idea of gene-environment correlations that jointly act on brain structure and function. These results are promising for future applications of genomICA and subsequent polygenic component scores. Genomic components are uncorrelated and unmix intermediate genetic processes. Therefore, they may help better understand and stratify the (etiological) heterogeneity of complex traits in both clinical and general population cohorts.

## Methods

### Methods Overview Flowchart

**Figure 3:**
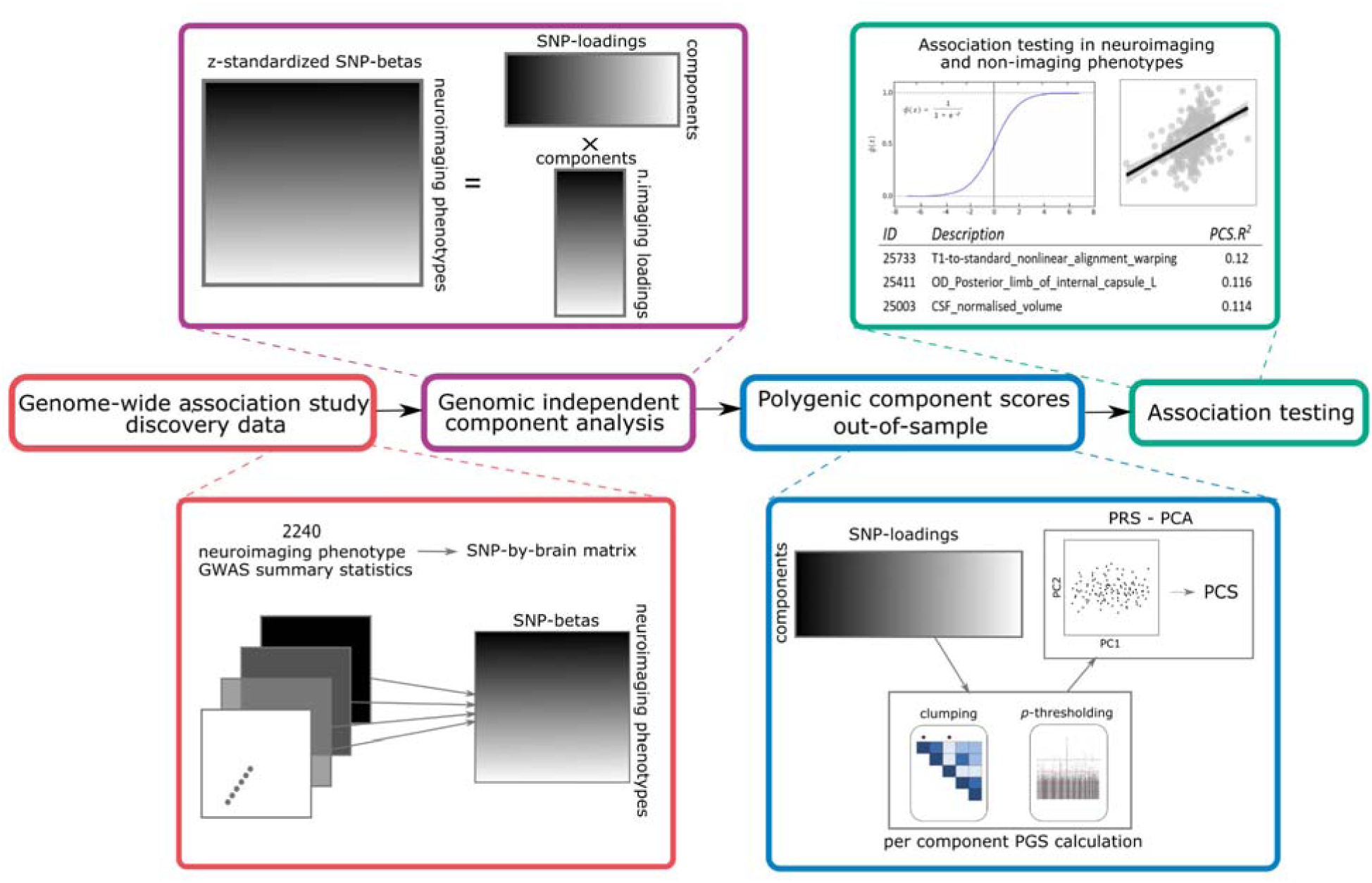
Schematic overview of the genomic independent component analysis with subsequent polygenic scoring analysis in four steps. 1. GWAS summary statistics are concatenated into a SNP-by-trait matrix. Then, betas across summary statistics are adjusted for their own standard error (z-standardized). 2. GenomICA is applied to this matrix to create a set of principal components and a set of independent components. 3. SNP-loadings of each component are used to compute polygenic component scores in an independent sample with PRS-PCA. 4. Polygenic component scores are used in general linear models and logistic regression models to predict phenotypes out-of-sample.

### Data

The analysis required multiple data sets. First, a discovery dataset of SNP-by-brain phenotype GWAS summary statistics for the calculation of latent genomic components. Second, two out-of-sample target datasets for calculation of PCSs: one with neuroimaging data (N = 3,506), and one without neuroimaging data (N = 338,429). As the discovery dataset (N = 33,224), we used the Oxford Brain Imaging Genetics (BIG-40) database containing the results of over 4000 GWASs of MRI based derived brain phenotypes in the UK Biobank (UKB) ^8,53^. The neuroimaging traits were grouped into categories: T1-weighted MRI, diffusion-weighted MRI (dMRI), susceptibility-weighted imaging (SWI), fluid-attenuated inversion recovery (FLAIR), task-based functional MRI (tfMRI), resting-state functional MRI (rsfMRI), as well as a quality control (QC) category. Metrics of rsfMRI with relatively high heritability were the amplitudes of functional signal fluctuations and six ICA-derived ‘*global*’ measures of functional connectivity and are therefore included in the analysis ^1,8^. Excluded brain phenotypes with low heritability were node-to-node rsfMRI metrics ^1^. Specifics on the UKBB GWAS pipeline can be found in the main publications ^1,8^ and the BIG-40 website (https://open.win.ox.ac.uk/ukbiobank/big40/). This set of N = 2,240 neuroimaging GWAS summary statistics were used to extract latent genomic components using genomICA, described in a subsection below.

To compute PCSs based on genomICA derived principal and independent components we used an out-of-sample set of UKB participants (i.e. not included in genomICA decomposition). We were granted access to the full UKBB genotype data of N = 377,663 participants aged between 40 and 69 years. These data were genotyped using the Affymetrix UK BiLEVE Axiom array or the UK Biobank Axiom array. Of the 377,663 participants originally available, 33,224 were excluded due to their inclusion in the discovery sample used to derive the genomICA components. This left a final genotype dataset of 344,439 participants and 10,203,392 SNPs (post-QC) available for polygenic scoring prior to clumping. Among these, 3,506 participants had neuroimaging data and were added to the UK Biobank after 2021, ensuring no overlap with the discovery sample. These 3,506 individuals constituted the sample for the first analysis and were removed from the dataset for the second analysis. For the second analysis of non-neuroimaging phenotypes the paper of Watanabe and colleagues (2019) was consulted to select non-neuroimaging phenotypes with clean data and large sample size available in UKB. Based on the data availability in our UKB data the second independent target dataset for non-neuroimaging traits consisted of N = 338,429 participants across non-imaging phenotypes.

### Data quality control of target samples

The initial target sample genetic dataset contained 93,095,623 SNPs ^54^. The data was quality controlled for sex mismatches, sample missingness > 0.05, heterozygosity outside of 3 standard deviations (SD), and participants of non-European descent (self-report) were removed. Principal components (PC) to account for population stratification during association testing were derived. Further, relatedness was accounted for by calculating the KING kinship coefficients and creating family clusters for participants with KING > 0.0884 (first – and second-degree relatives). Then, the sample was reduced until only one participant was left per family cluster ^55^. SNPs were removed if they had Hardy-Weinberg equilibrium (HWE) < 1e-6, genotype missing rate > 0.05, minor allele frequency (MAF) < 0.005, or imputation quality INFO score < 0.8. After quality control the final SNP count was N = 10,203,392 ^55^.

The neuroimaging target dataset of 3,506 participants was quality controlled by checking if the field values were consistent with the neuroimaging-phenotype they represent via the UKB catalogue. Then, the number of missing values per phenotype was assessed to ensure that phenotypes had sufficient samples for regression analysis (N > 2,500). After QC, neuroimaging-phenotypes (N = 2,240) were filtered based on their loadings in the neuroimaging-space of independent and principal genomic components (|z| > 2). This brought the number of neuroimaging-phenotypes down to N = 1,269 for genomic ICs and N = 1,330 for genomic PCs (tables S2 & S3).

The non-imaging target dataset consisted of 152 continuous and 758 binary, categorical or ordinal phenotypes from the UKB data repository^56^. The data consisted of phenotypes from multiple domains: physical attributes, physical health, reproductive health, mental health and well-being, lifestyle and behaviour, social and economic factors, cognitive performance, diet, family history, and mobile phone usage. A full list of phenotypes used in the analysis is shown in supplement (table S4 & S6). We altered the data coding for some variables based on the recommendations of Watanabe and colleagues (^56^ - supplementary table 1). In the binary, categorical and ordinal phenotypes we also removed all instances where participants answered, “do not know”, “do not recall”, “prefer not to answer”, or “less than one [item] a week” across all phenotypes. Lastly, we only retained those phenotypes that had >50,000 data points, as in ^56^, to ensure sufficient power. Ordinal phenotypes were binarized in a pair-wise manner so that the full range of answer contrasts could be analysed. Categorical multiple-choice answers were binarized. For continuous phenotypes with multiple measurements at the same visit (e.g. systolic blood pressure) the mean across measurements was calculated.

### Clumping

Prior to the decomposition with genomICA the GWAS summary statistic SNPs (N = 17,103,079) of 2240 neuroimaging phenotypes were clumped at LD-threshold *R*^2^ = 0.5 and significance threshold p = 0.001 with a 250kb range using plink2.0^57^. This resulted in N = 1,032,967 lead-SNPs in the genome-dimension included in the decomposition. The data was clumped in such a way that SNPs in the European 1000G reference panel are prioritized and retained to facilitate overlap with other GWAS summary statistics where the 1000G reference panel is often used. During post-hoc QC, 440 SNPs were identified as duplicates and therefore removed, bringing the final number of SNPs in the SNP x brain data to N = 1,032,527.

### GenomICA components

GenomICA was applied as previously described in ^34^. The GWAS summary statistics were concatenated into a m x n matrix, representing a genome-wide matrix of MRI-derived neuroimaging phenotypes (m = 2240) and common genetic variants (n = 1,032,527 clumped SNPs). Multivariate decomposition was applied to the z-standardized GWAS betas, such that SNPs are normalized for their own standard errors (SE). The matrix was decomposed using v3.15 of Multivariate Exploratory Linear Optimised decomposition into independent components (MELODIC), a probabilistic ICA algorithm^58^. We applied the mask of clumped SNPs to the m x n matrix such that only lead-SNPs remain. The ensuing m x n matrix of trait-by-SNP values was decomposed using MELODIC (V. 3.1.5)^58^, part of the FSL toolbox^59^. This produces a predefined number of PCs as an intermediate measure, because we did not utilize the available option to infer the optimal number of components with Bayesian inference. The PCs are then rotated to optimize non-Gaussianity across components, thereby creating the same number of independent components (IC). Each component is represented by two vectors: one vector of SNP-loadings and a vector of neuroimaging-phenotype-loadings. This feature of genomICA decomposition allows the linking between patterns of genetic effect sizes and corresponding patterns of brain features. In our previous work we determined that decomposing into 10 components was optimal for reproducibility for N = ∼160,000 SNPs clumped at *R*^2^ < 0.1. For this highly clumped dataset 10 components captured ∼39% of SNP effect variance. Here, with less stringent clumping 16 PCs and ICs (Dimension 16) captured the same (reproducible) ∼39% SNP effect variance. For this reason, we here chose to decompose into 16 components, to capture the same, previously determined reproducible, variance.

The final output of genomICA is therefore 16 PCs and 16 ICs, each of which is a vector of SNP-loadings and trait-loadings. Both PCs and ICs are used in subsequent analyses.

### Polygenic Score calculation

To compute polygenic component scores (PCSs), we applied PRS-PCA^60^, using SNP-loadings from genomic components as weights. As previously described, polygenic scores were first calculated using PRSice 2.3.5^25^ across a range of p-value thresholds (0.001, 0.05, 0.1, 0.2, 0.3, 0.4, 0.5, and 1). SNPs were re-clumped using an LD threshold of *r*² < 0.1 within a 250 kb window, and a sliding window approach, reducing the set to N = 290,660 lead SNPs. Then, PCA was applied to identify the principal axis of variance across thresholds. The first principal component effectively captured the shared variance across thresholds for all components (table S8 & S9). This component was used for all downstream analyses and is referred to as the PCS of a given component. We opted not to use methods like PRS-CS or LDpred^61,62^, which rely on continuous shrinkage and LD-informed priors, due to insufficient LD structure in our genomic components.

### Polygenic Component Score association testing

PCSs of all ICs and PCs were used as predictors in general linear regression models (for continuous phenotypes) and logistic regression models (for binary/binarized phenotypes). To calculate the phenotypic variance explained by PCSs we constructed a full-model with all 16 PCSs and covariates, and a null-model, with only covariates as predictors. For all models we calculate the conventional and proportional variance explained (*R*^2^). Although both are routinely reported in polygenic scoring they can differ substantially depending on the variance explained by the covariates, so for comparison with other literature, we report both here. Conventional (*R*^2^*_conventional_*) is the variance explained by the null model subtracted from the variance explained by the full model.

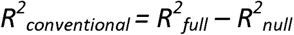

Proportional variance explained is the proportion of the variance that the full model explains which is not captured by the null model.

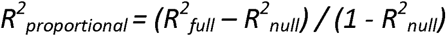

For logistic regression models we calculated *Nagelkerke’s* pseudo-*R*^2^, as well as *McFadden’s* pseudo-*R*^2^ as a comparative metric, and then calculated the proportion of phenotypic PCS-*R*^2^ in the same way as outlined above. To determine the significance of PCS-*R*^2^ we ran an ANOVA between the full and null general linear models, and a χ^2^-test for logistic binomial models. P-values were corrected for type I error with false-discovery rate (FDR) correction across outcome phenotypes tested. Finally, coefficients and p-values of individual PCSs were extracted from each model in turn to interrogate individual component associations. P-values within each model were likewise corrected for type I errors by applying FDR correction.

Covariates included in our analysis were age, sex, and body-mass-index, assessment location, genotyping batch and 40 population stratification PCs derived during data quality control. The latter provide an estimate for global genetic structure and are therefore useful for inclusion in statistical models. For the neuroimaging phenotype models we also included head-motion and head-location (x-, y-, z-axes) in the MRI-scanner as covariates.

## Data Availability

All data used to create the components used in the present study are available online at https://open.win.ox.ac.uk/ukbiobank/big40/; https://ctg.cncr.nl/software/summary_statistics; https://pgc.unc.edu/for-researchers/download-results/. The genomic components generated by this study as well as the outcome of association testing are available online under https://genomica.info/. Component vectors created by the current study are available upon reasonable request to the authors. The human questionnaire data used for association testing was obtained through an application with the UK Biobank consortium and can be obtained upon request (https://biobank.ndph.ox.ac.uk/ukb/catalogs.cgi).

## Acknowledgments

FAMILY is funded by the European Union, the Swiss State Secretariat for Education, Research and Innovation (SERI) and the UK Research and Innovation (UKRI) under the UK government’s Horizon Europe funding guarantee. ES is also funded by an ERC Consolidator Grant (Project 101170512) funded by the European Union. Views and opinions expressed are however those of the author(s) only and do not necessarily reflect those of the European Union, or the European Research Executive Agency (REA), the SERI or the UKRI. Neither the European Union nor the granting authorities can be held responsible for them. CFB gratefully acknowledges funding from the Wellcome Trust Collaborative Award in Science 215573/Z/19/Z and the Netherlands Organization for Scientific Research Vici Grant No. 17854.

This research has been conducted using data from UK Biobank (http://www.ukbiobank.ac.uk/), under application 23668. UK Biobank is supported by its founding funders the Wellcome Trust and UK Medical Research Council, as well as the Department of Health, Scottish Government, the Northwest Regional Development Agency, British Heart Foundation and Cancer Research UK.

This work used the Dutch national e-infrastructure with the support of the SURF Cooperative using grant no. EINF-10477.

## Supplemental Material

This file contains the visualized supplementary output of the genomICA based PRS-PCA analysis. CTRL+F search is encouraged for individual components or Neuroimaging modalities.

Results presented here can be viewed on genomica.info, an interactive platform that presents these results as well as the outcome of gene-enrichment analyses on the original components.

### Results of full models

**Figure 1:**
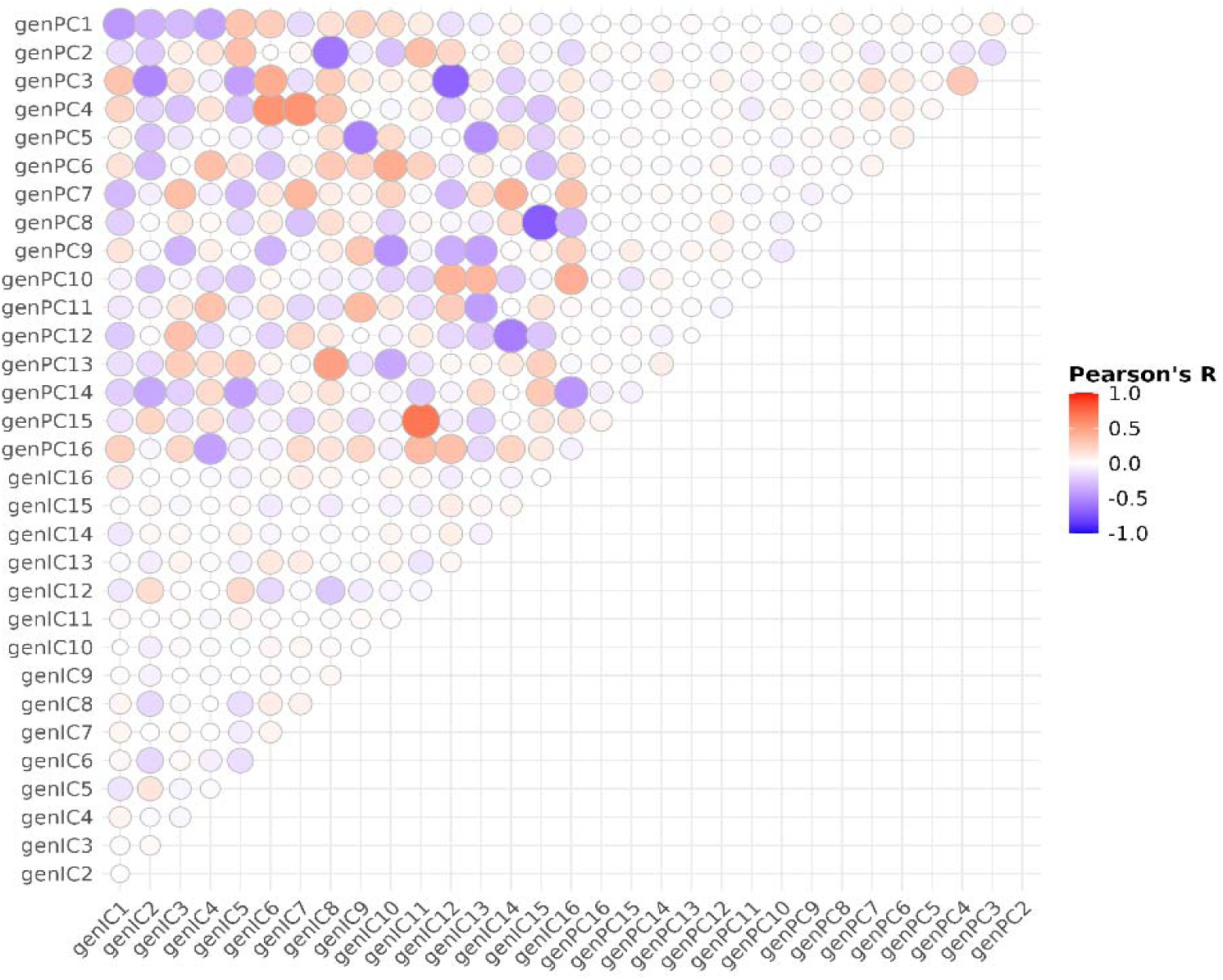
Correlation heatmap resulting from the correlation of the first polygenic score PC derived from all 16 genomic IC and genomic PC. The upper-left 16-by-16 square indicated the correlations of genomic PC derived polygenic PCs with the genomic IC derived polygenic PCs. The upper-right and lower-left triangles indicate the correlations between genomic PC derived polygenic PCs and genomic IC derived polygenic PCs, respectively. IC – independent component; PC – principal component; PRS - polygenic risk score

### Genomic Independent Polygenic Component Scores

**Figure 2:**
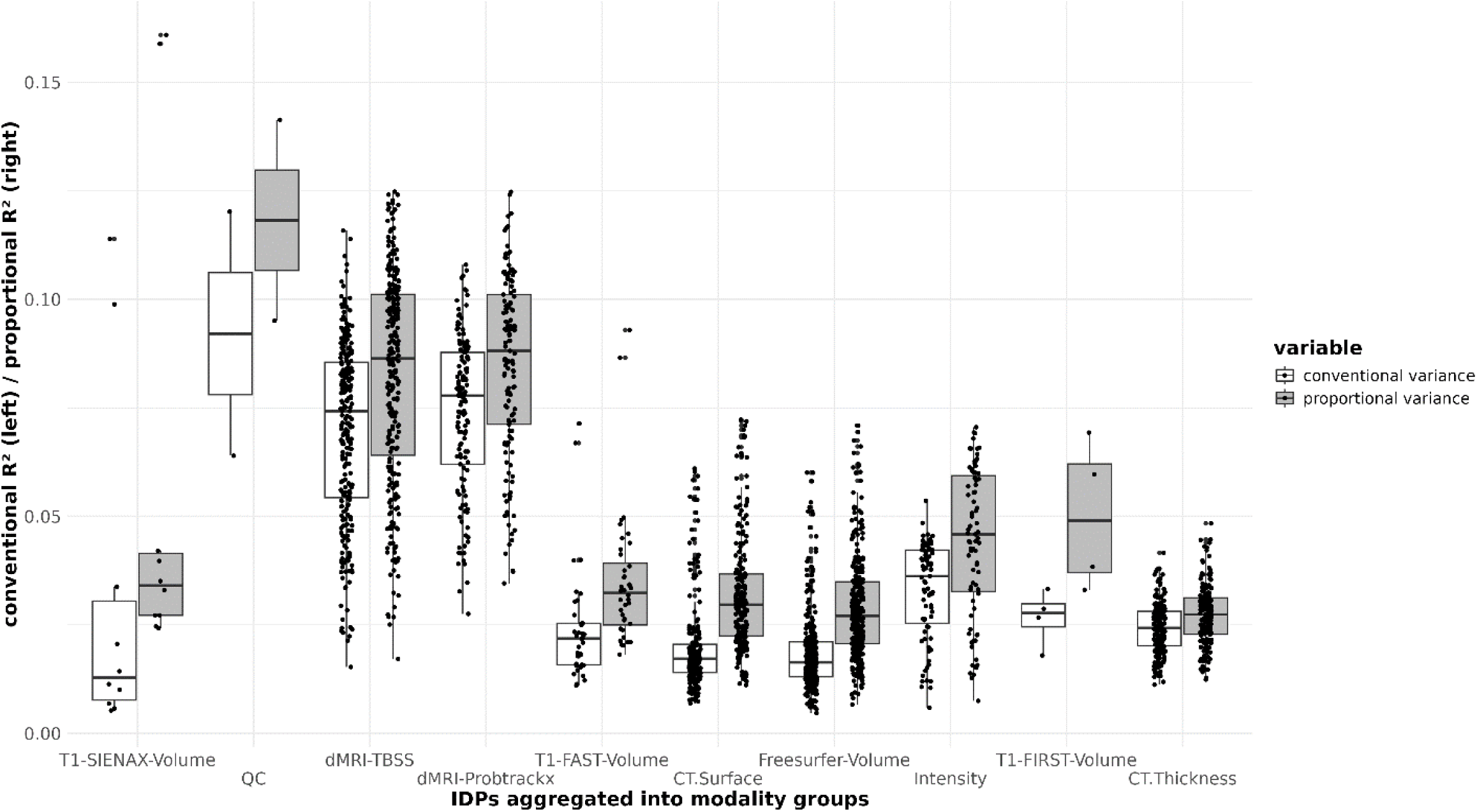
Overview of neuroimaging phenotypic variance explained by genomic IC PCSs significant after FDR-correction and aggregated into categories, indicated on the x-axis. Shown is the conventional variance explained (white), and proportional variance explained (grey). The box-plot central line indicates the mean within the category and the outer borders the 1^st^ and 3^rd^ quartiles. CT – cortical; IC – independent component; IDP – imaging derived phenotype; ProbtrackX – probabilistic tractography; TBSS – tract-based spatial statistics.

**Figure 3:**
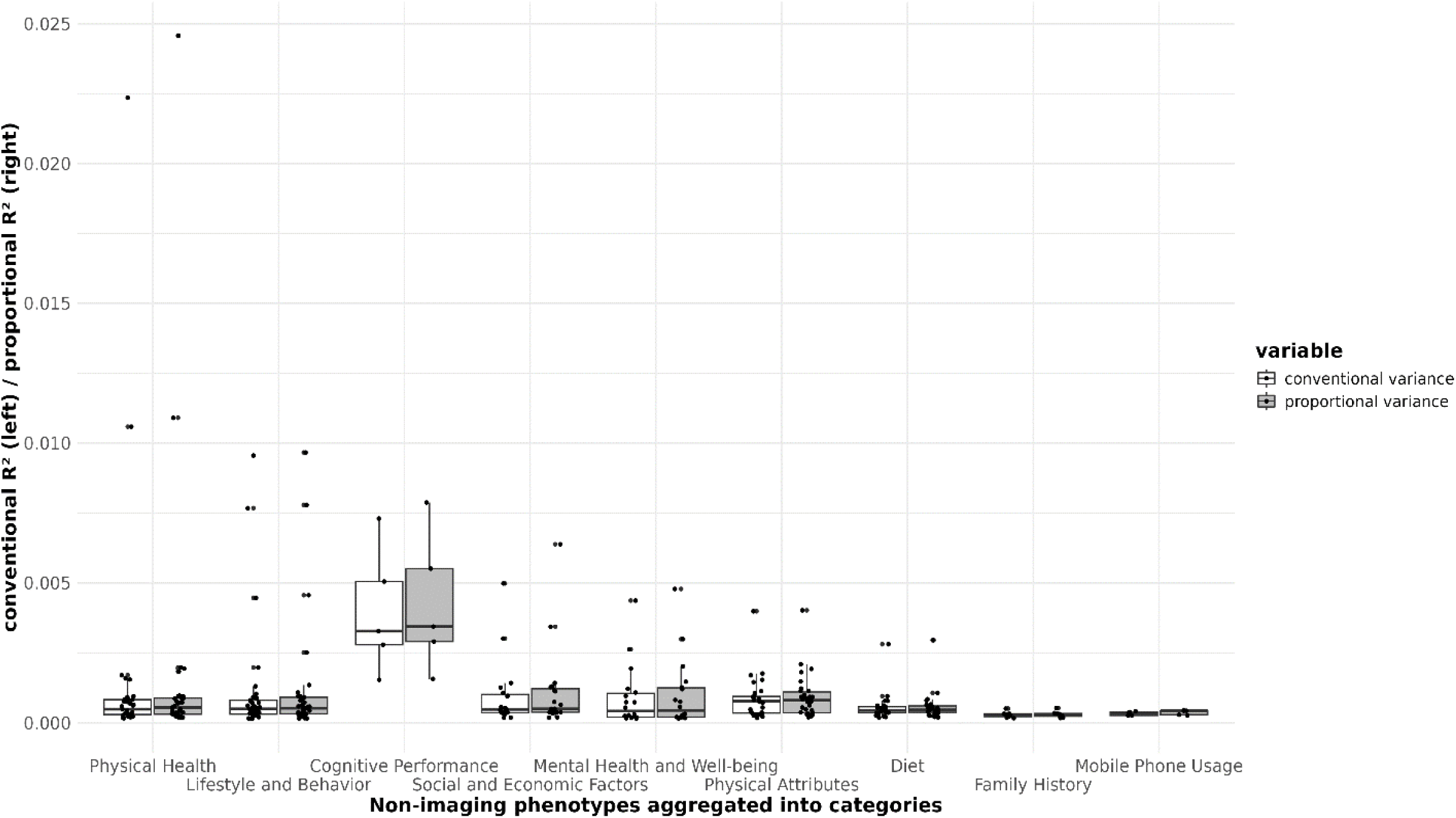
Overview of binary/categorical phenotypic variance explained by genomic IC PCSs significant after FDR-correction and aggregated into domains, indicated on the x-axis. Shown is the conventional Nagelkerke’s variance explained (white), and proportional Nagelkerke’s variance explained (grey). The box-plot central line indicates the mean within the category and the outer borders the 1^st^ and 3^rd^ quartiles.

**Figure 4:**
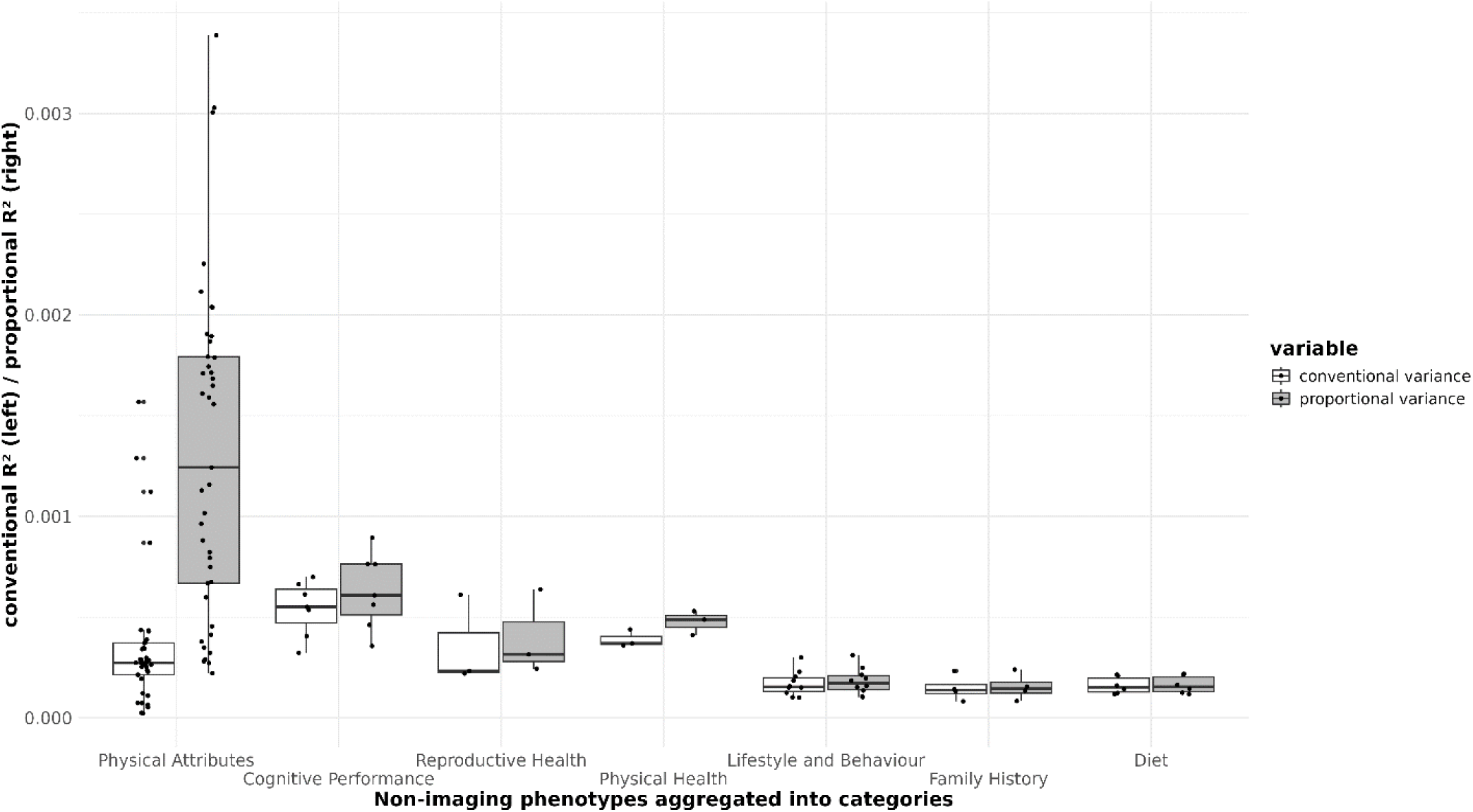
Overview of continuous phenotypic variance explained by genomic IC PCSs significant after FDR-correction and aggregated into domains, indicated on the x-axis. Shown is the conventional Nagelkerke’s variance explained (white), and proportional Nagelkerke’s variance explained (grey). The box-plot central line indicates the mean within category and the outer borders the 1^st^ and 3^rd^ quantiles.

**Figure 5:**
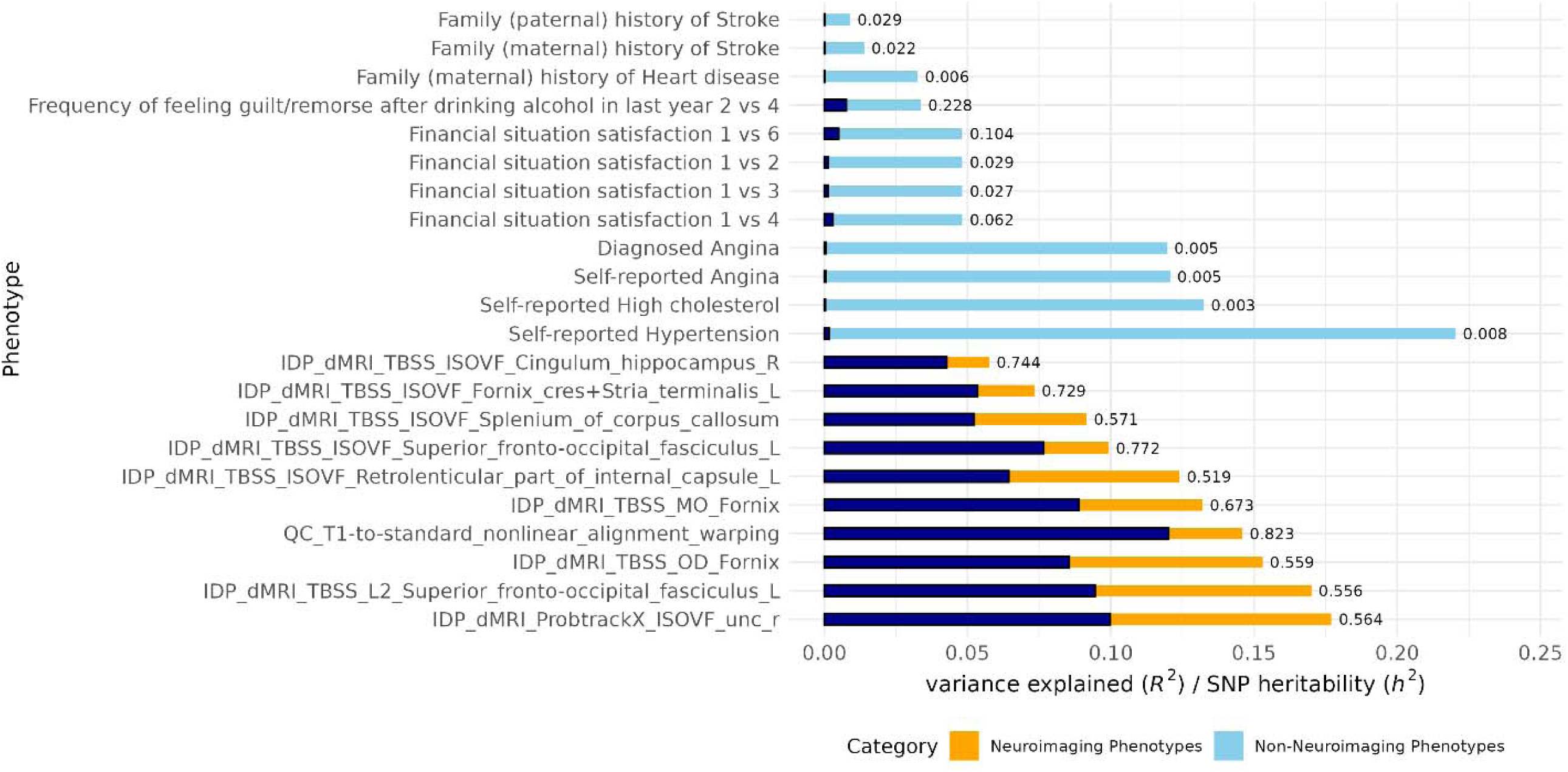
Bar-plot of phenotypic variance explained jointly by genomic IC PCSs and the SNP-heritability of select phenotypes. Phenotypes (y-axis) are selected to represent SNP-heritability samples from non-neuroimaging space (light-blue) and neuroimaging phenotypes (orange). The corresponding phenotypic variance explained Is overlaid in dark-blue. The x-axis shows both the variance explained (dark-blue), and the SNP-heritability (light blue / orange). The number next to each bar represents the fraction of SNP-heritability captured as phenotypic variance explained.

**Figure 6:**
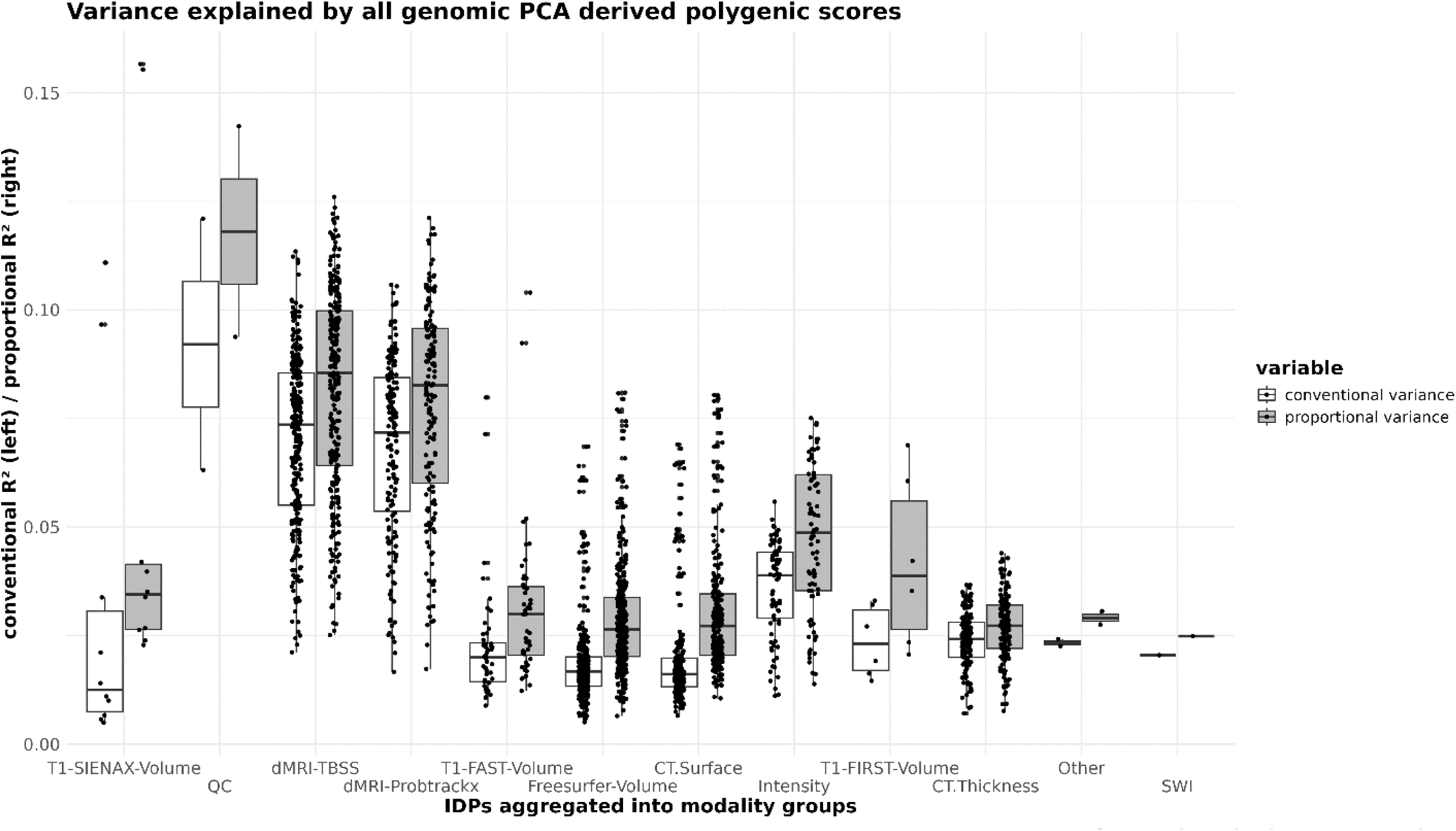
Figure showing the overview of variance explained of the full model that includes all genomic ICs. Shown are the imaging phenotypes aggregated into modalities, where each dot represents a single neuroimaging phenotype within each category. Here indicated are both the conventional variance explained (*R*^2^=*R*^2^full – *R*^2^null) and the proportional variance explained (*R*^2^= (*R*^2^_full_ – *R*^2^_null_) / (1 - *R*^2^_null_)), the latter of which calculates the proportion of the variance explained by the full model not explained by the null model.

### Genomic Principal Polygenic Component Scores

**Figure 7:**
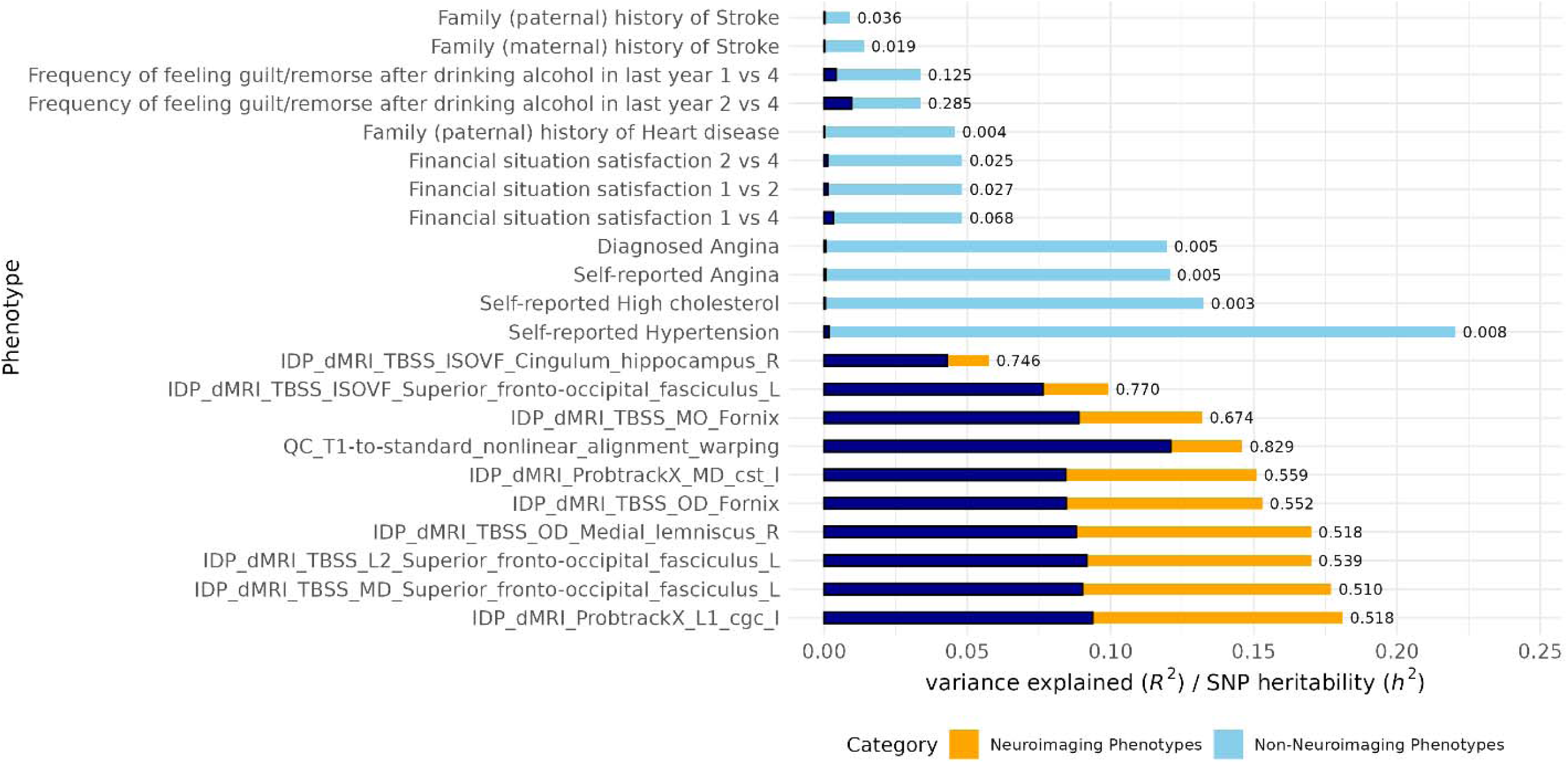
Bar-plot of phenotypic variance explained jointly by genomic PC PCSs and the SNP-heritability of select phenotypes. Phenotypes (y-axis) are selected to represent SNP-heritability samples from non-neuroimaging space (light-blue) and neuroimaging phenotypes (orange). The corresponding phenotypic variance explained Is overlaid in dark-blue. The number next to each bar represents the fraction of SNP-heritability captured as phenotypic variance explained.

**Figure 8:**
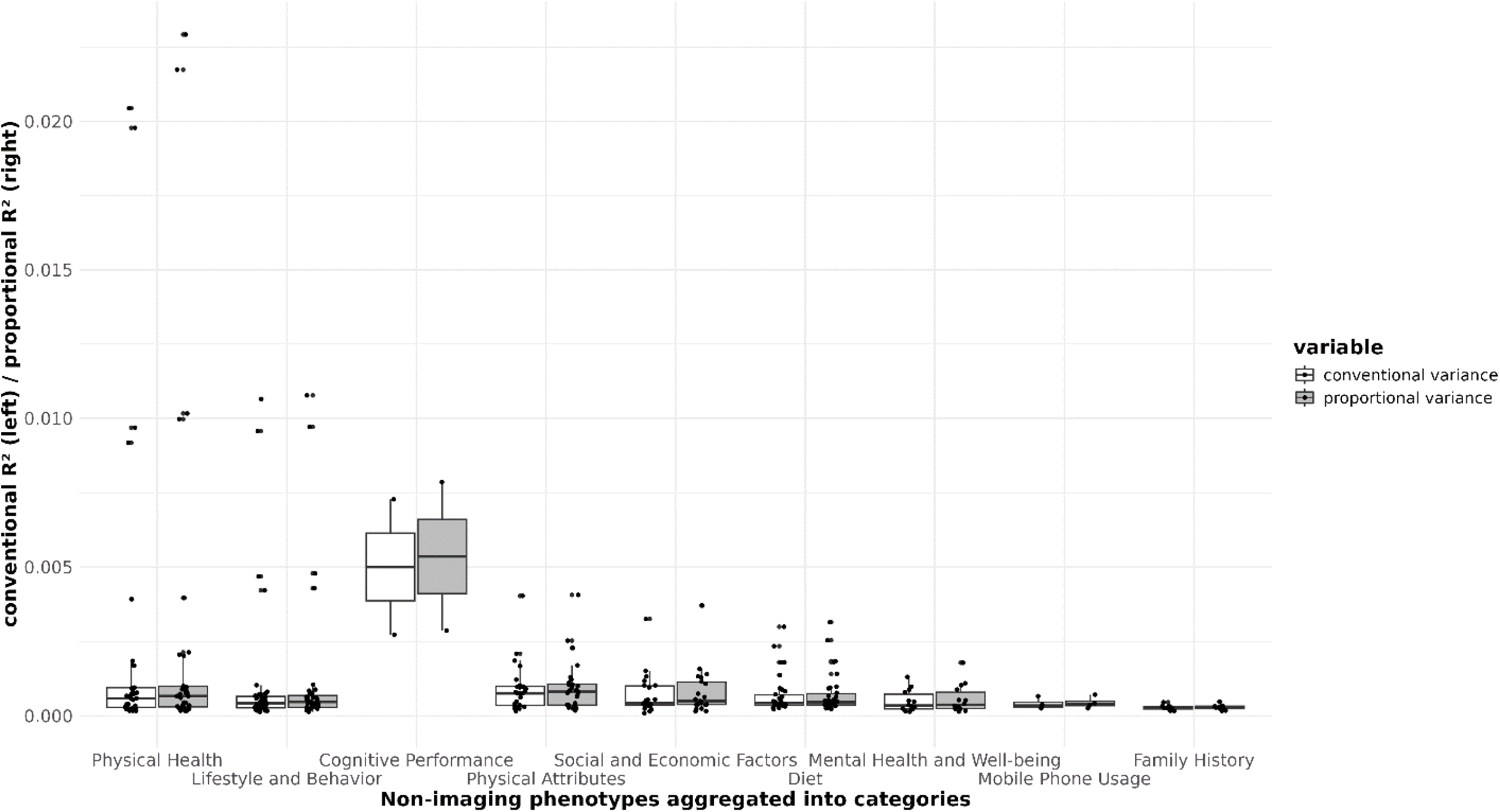
Figure showing the overview of variance explained of the full model that includes all genomic PCs. Shown are the binary non-Neuroimaging phenotypes aggregated int categories, where each dot represents a single non-neuroimaging phenotype within each category. Here indicated are both the conventional variance explained (*R*^2^=*R*^2^full – *R*^2^null) and the proportional variance explained (*R*^2^= (*R*^2^_full_ – *R*^2^_null_) / (1 - *R*^2^null)), the latter of which calculates the proportion of the variance explained by the full model not explained by the null model.

### Results of Genomic independent Component Scores in Non-Neuroimaging Phenotype Space highlighted in the text (PCS15 and PCS8)

**Figure 9:**
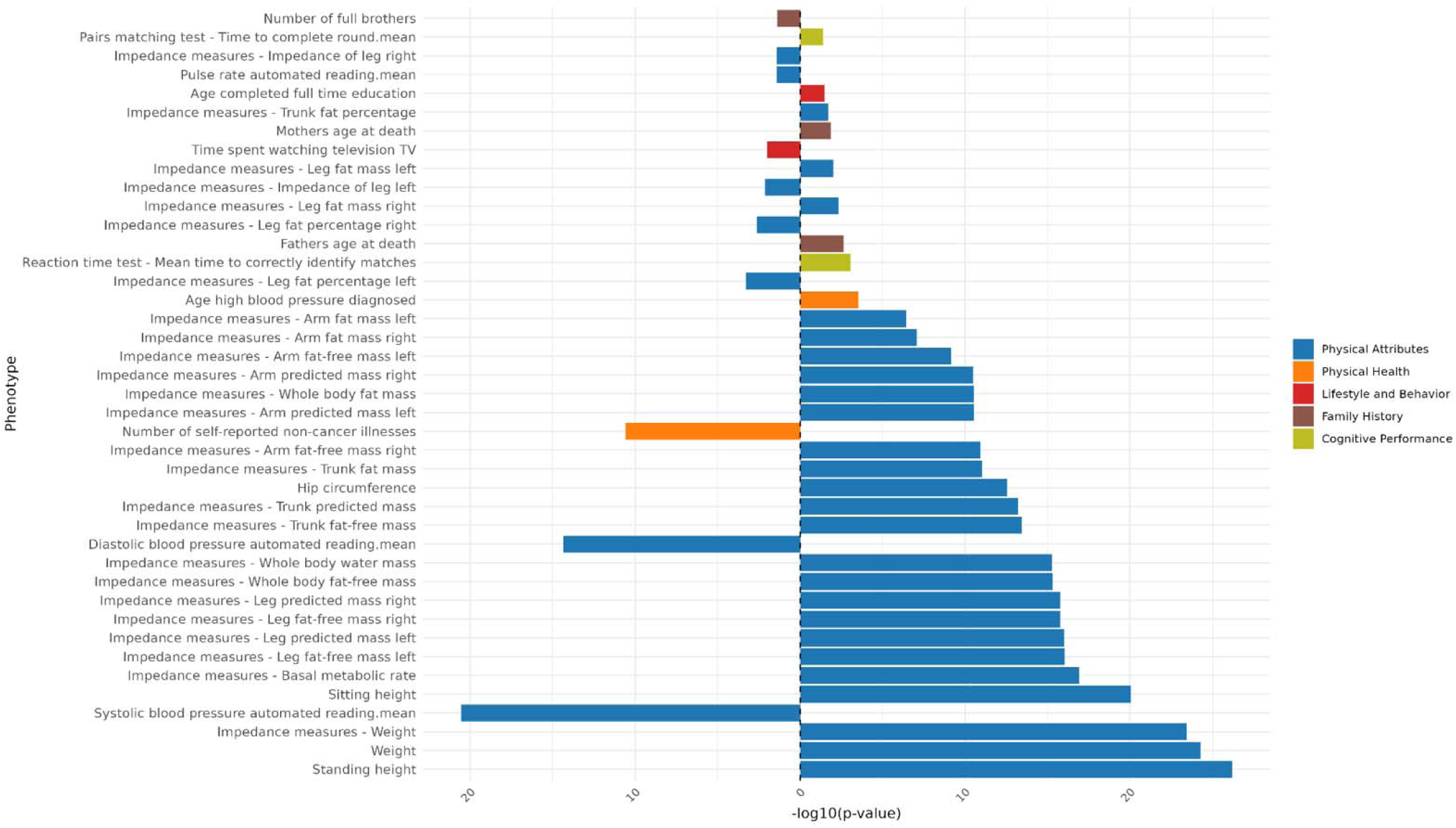
Overview of PCS15 significant associations with continuous non-imaging phenotypes. The x-axis shows the -log(10) p-value. The direction of the bar indicates the direction of association, in this case the sign of the beta. Right facing bars show a positive association, left facing bars a negative association. Phenotypes are binned into domains which are indicated in a color scheme contained in the legend on the right-hand side of the figure.

**Figure 10:**
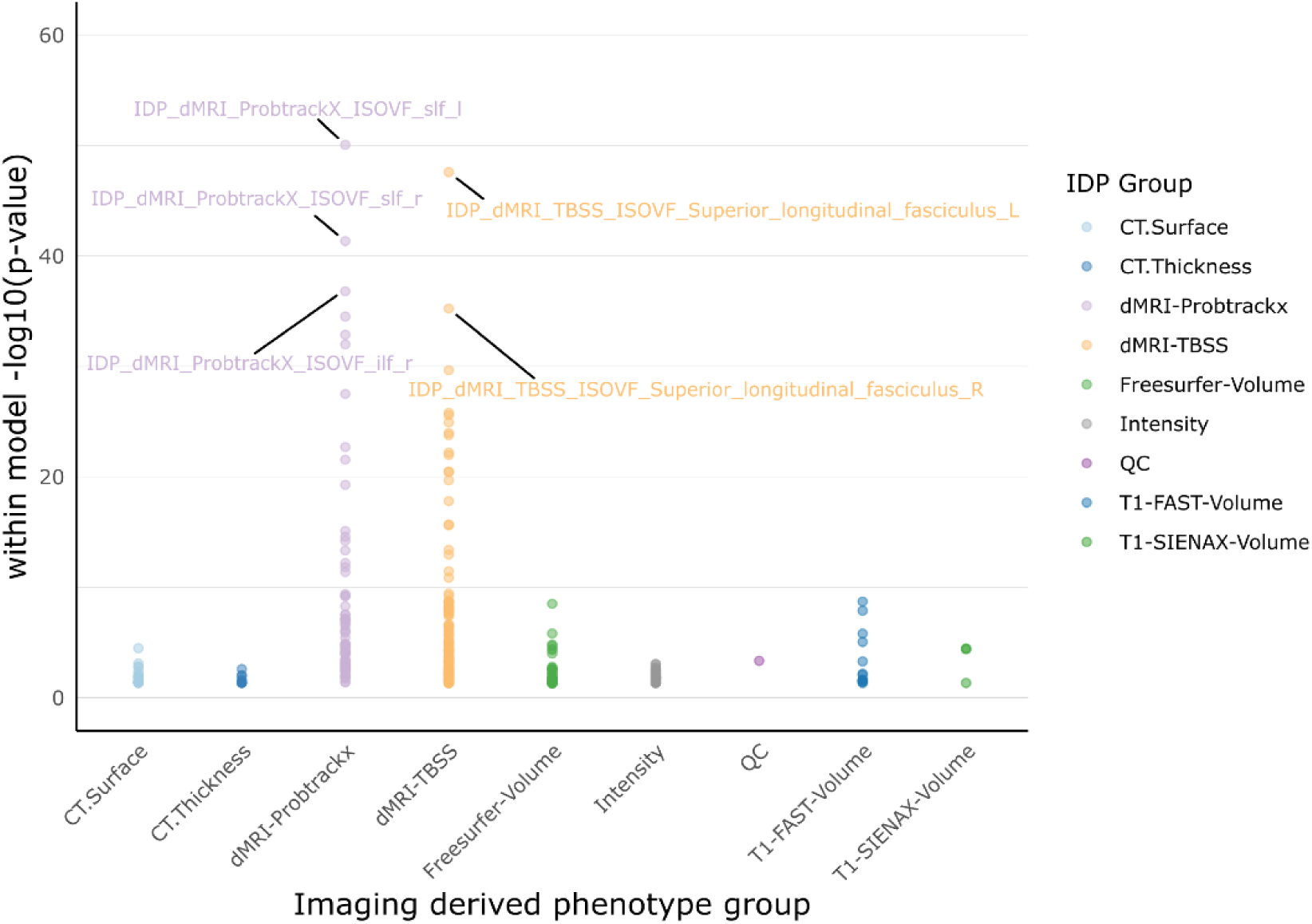
Dot-plot showing PCS15 associations with IDPs. The y-axis shows the -log(10) p-value, therefore the height of the dot in the plot is proportional to its significance. Each dot represents one IDP. IDPs were binned into categories shown in the legend on the right-hand side of the figure, indicated by a color scheme. Dmri – diffusion magnetic resonance imaging; IDP – imaging derived phenotype; ilf – inferior longitudinal fasciculus; ISOVF – Isotropic water diffusion; slf – superior longitudinal fasciculus

**Figure 11:**
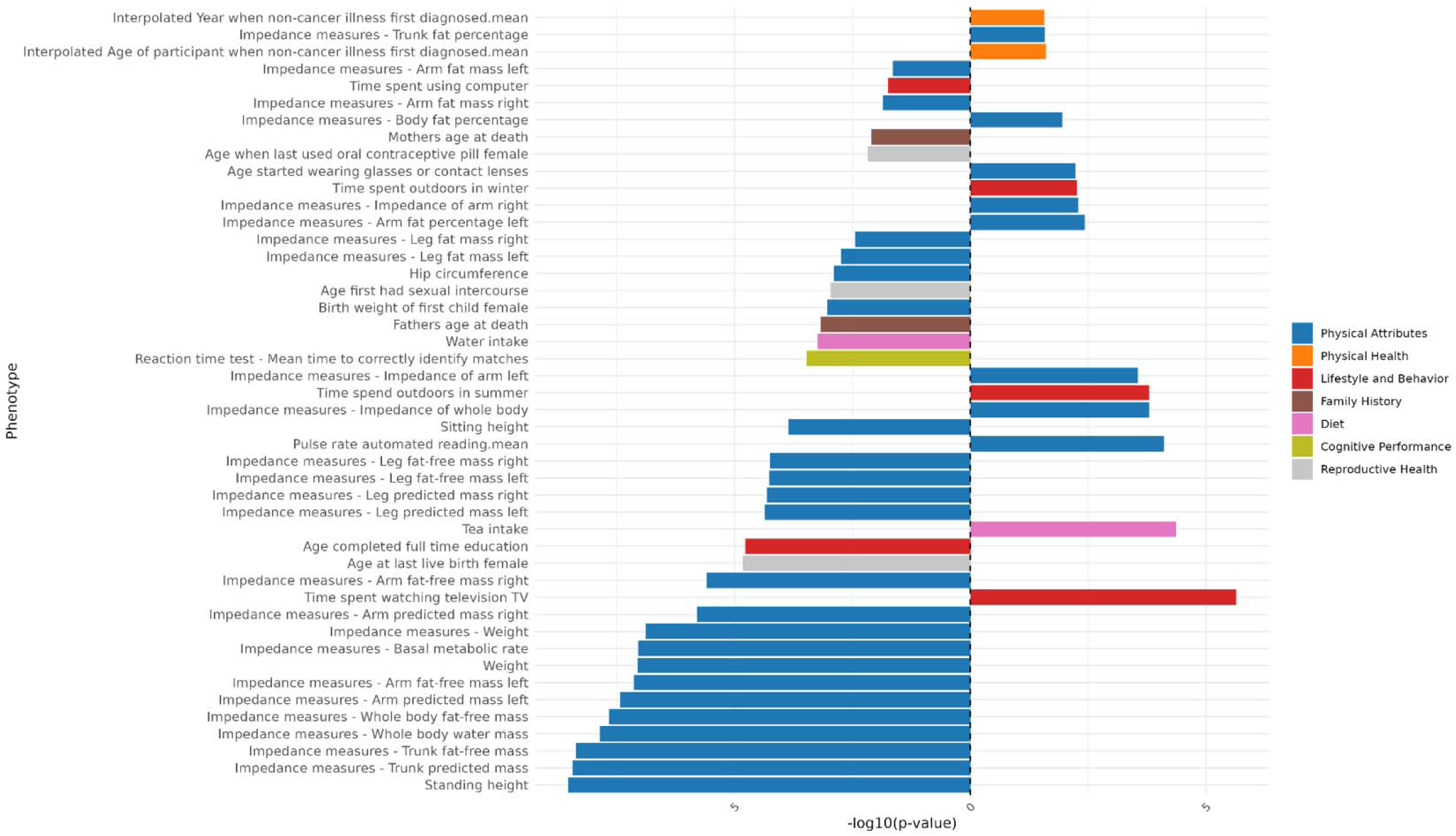
Overview of PCS8 significant associations with continuous non-imaging phenotypes. The x-axis shows the -log(10) p-value. The direction of the bar indicates the direction of association, in this case the sign of the beta. Right facing bars show a positive association, left facing bars a negative association. Phenotypes are binned into domains which are indicated in a color scheme contained in the legend on the right-hand side of the figure.

**Figure 12:**
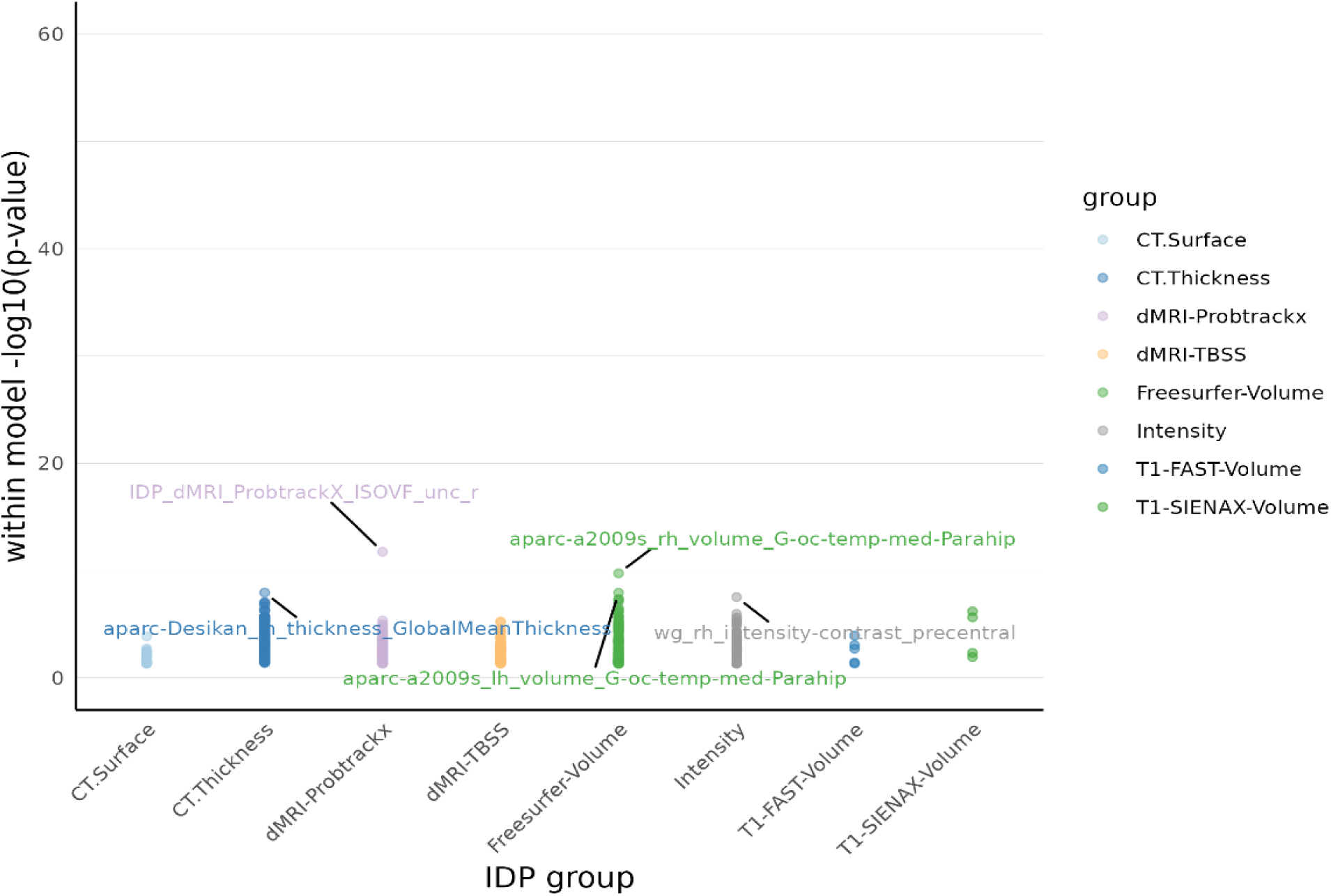
Dot-plot showing PCS8 associations with IDPs. The y-axis shows the -log(10) p-value, therefore the height of the dot in the plot is proportional to its significance. Each dot represents one IDP. IDPs were binned into categories shown in the legend on the right-hand side of the figure, indicated by a color scheme.

### Results of all other Genomic Independent Component Scores in Non-Neuroimaging Phenotype Space Independent Component 1

**Figure 13:**
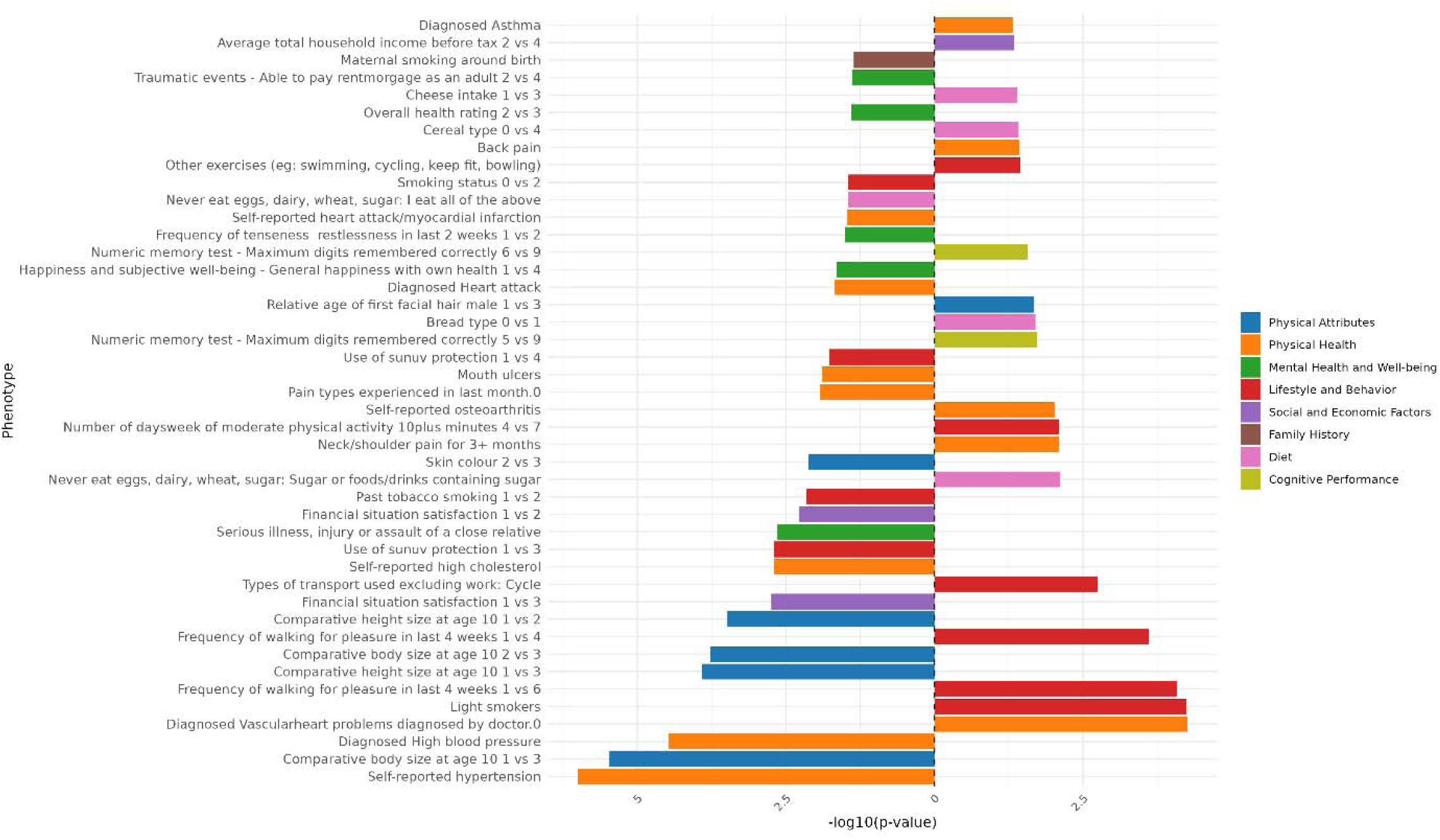
Genomic Independent Component 1 PCS association strength across binary/binarized non-neuroimaging phenotypes. Phenotypes (y-axis) are plotted against the -log(10) p-value (x-axis). Phenotypes shown are significant after FDR-correction. The direction of the bar indicates the direction of association, where a right-hand bar indicates a positive association, and a left-hand bar a negative association. In contrast analyses (e.g., comparing response 1 vs. response 3), the direction of the bar indicates the direction of the association in relation to the answers contrasted. Specifically, in a 1 vs. 3 contrast, a bar pointing to the right indicates an association with response 3, while a bar pointing to the left indicates an association with response 1. The phenotype “Diagnosed Vascular problems diagnosed by doctor.0” equates to “No Vascular problems diagnosed”. Phenotypes are color coded by phenotype category.

**Figure 14:**
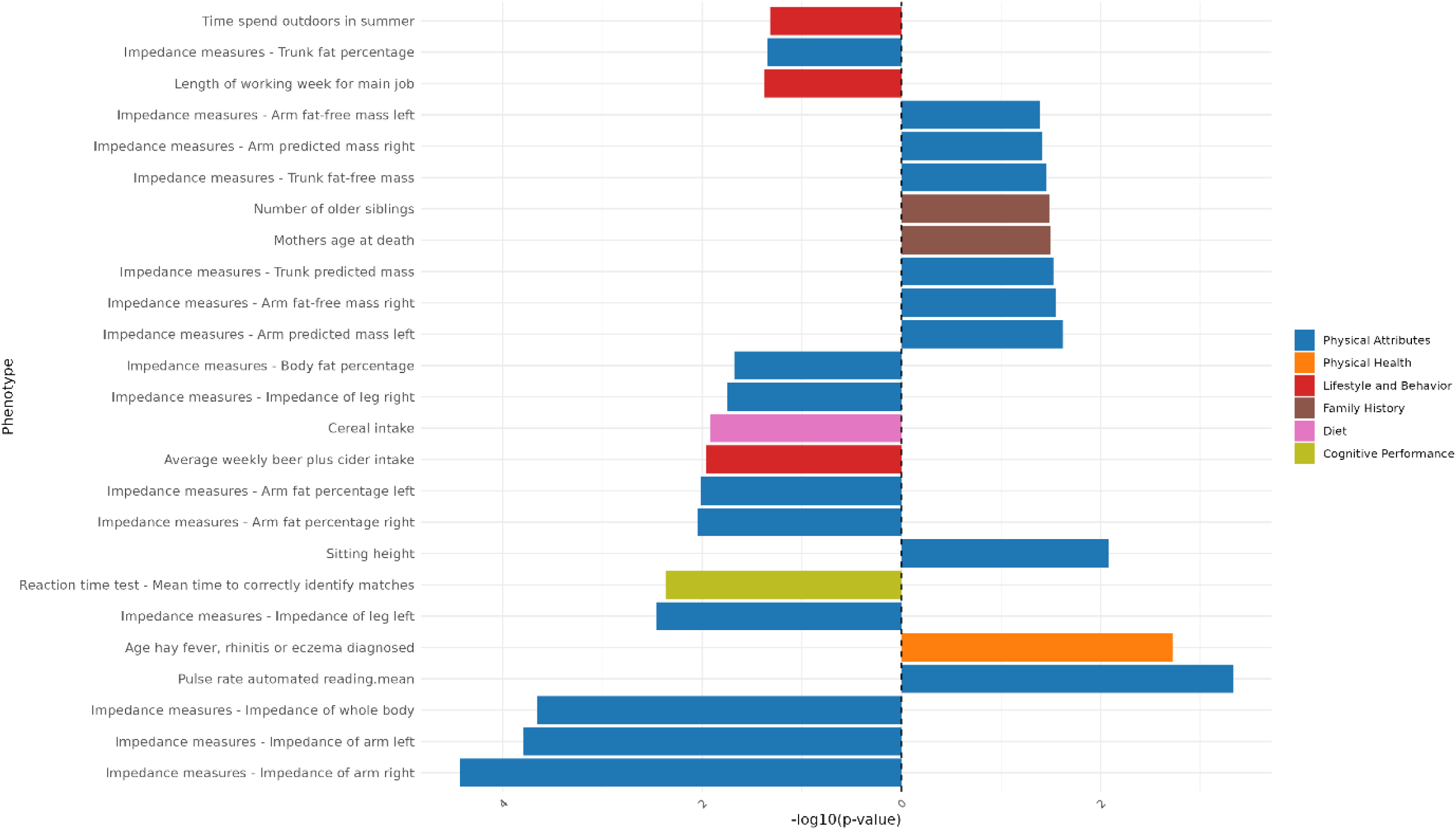
Genomic Independent Component 1 PCS association strength across continuous non-neuroimaging phenotypes. Phenotypes (y-axis) are plotted against the -log(10) p-value (x-axis). Phenotypes shown are significant after FDR-correction. The direction of the bar indicates the direction of association, where a right-hand bar indicates a positive association, and a left-hand bar a negative association. Phenotypes are color coded by phenotype category.

### Independent Component 2

**Figure 15:**
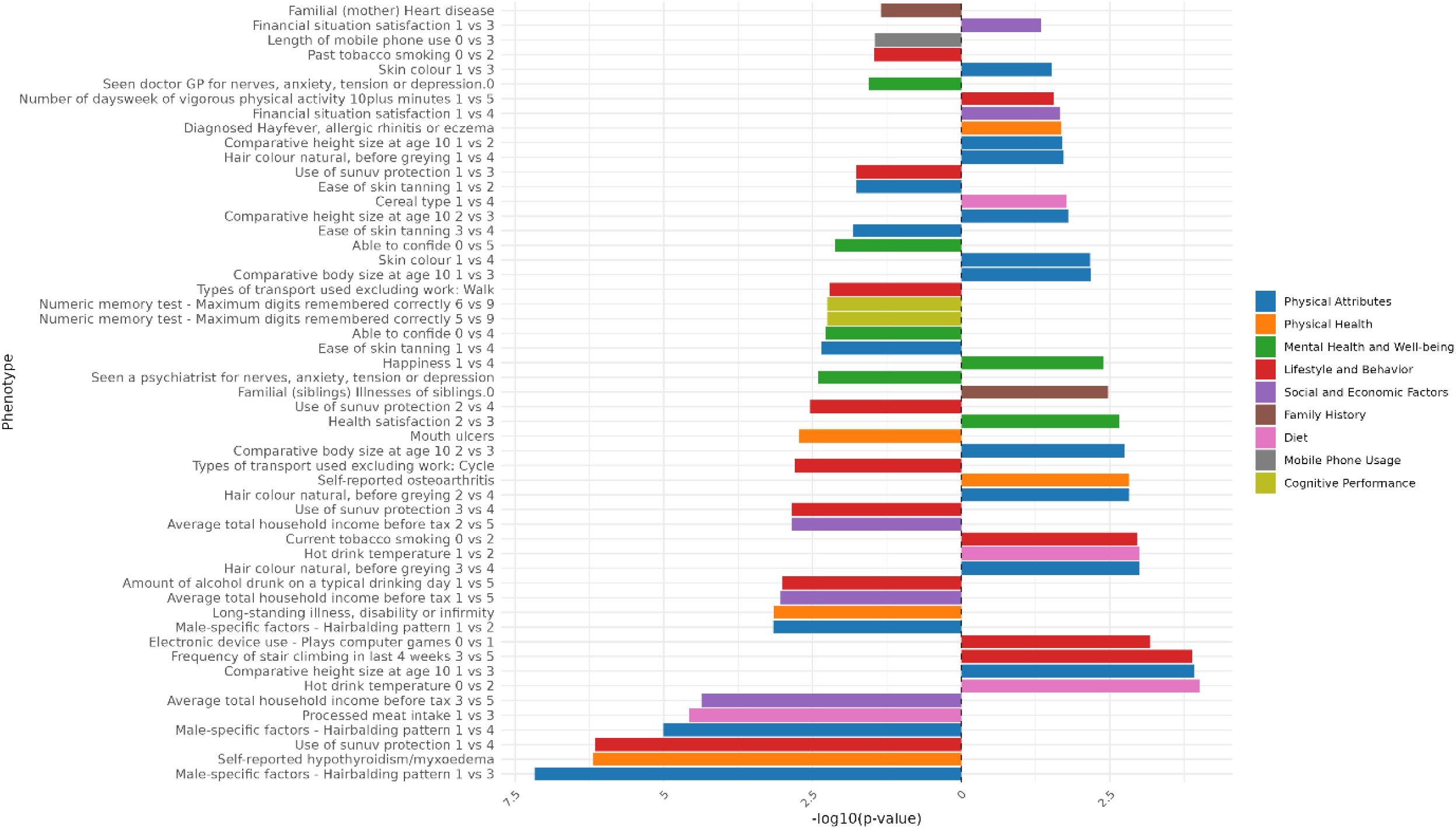
Genomic Independent Component 2 PCS association strength across binary/binarized non-neuroimaging phenotypes. Phenotypes (y-axis) are plotted against the -log(10) p-value (x-axis). Phenotypes shown are significant after FDR-correction. The direction of the bar indicates the direction of association, where a right-hand bar indicates a positive association, and a left-hand bar a negative association. In contrast analyses (e.g., comparing response 1 vs. response 3), the direction of the bar indicates the direction of the association in relation to the answers contrasted. Specifically, in a 1 vs. 3 contrast, a bar pointing to the right indicates an association with response 3, while a bar pointing to the left indicates an association with response 1. The phenotype “Diagnosed Vascular problems diagnosed by doctor.0” equates to “No Vascular problems diagnosed”. Phenotypes are color coded by phenotype category.

**Figure 16:**
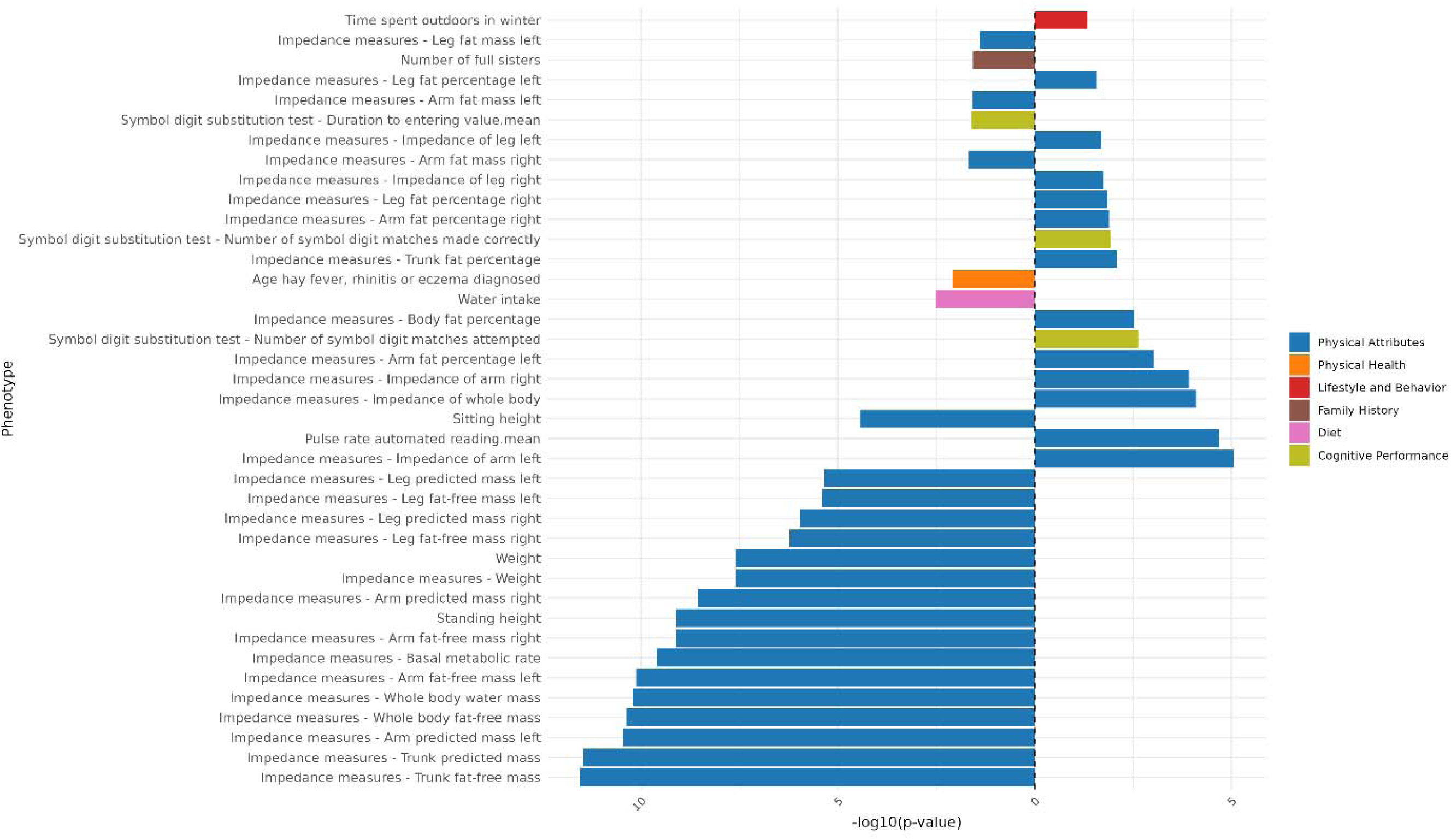
Genomic Independent Component 2 PCS association strength across continuous non-neuroimaging phenotypes. Phenotypes (y-axis) are plotted against the -log(10) p-value (x-axis). Phenotypes shown are significant after FDR-correction. The direction of the bar indicates the direction of association, where a right-hand bar indicates a positive association, and a left-hand bar a negative association. Phenotypes are color coded by phenotype category.

### Independent Component 3

**Figure 17:**
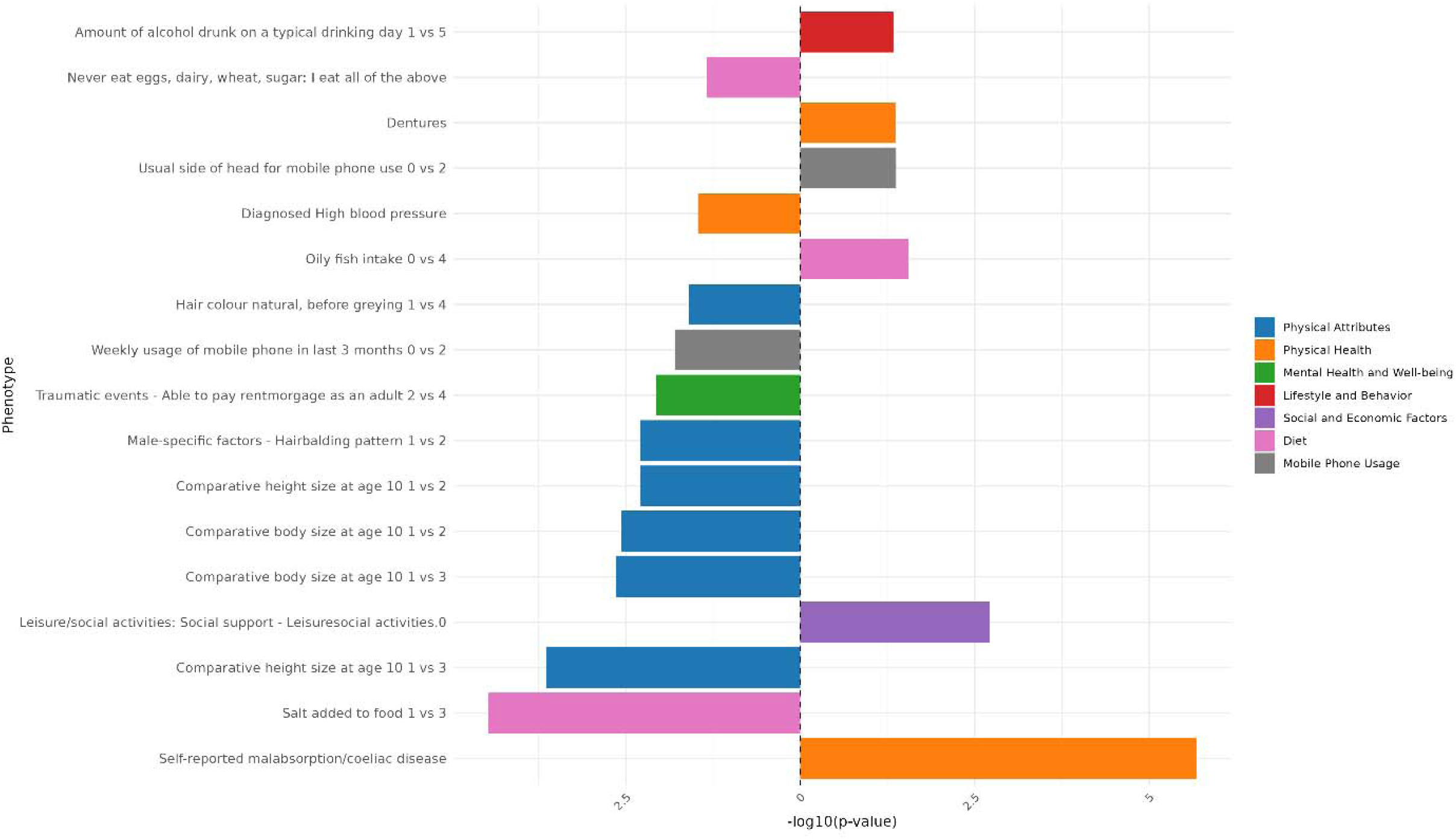
Genomic Independent Component 3 PCS association strength across binary/binarized non-neuroimaging phenotypes. Phenotypes (y-axis) are plotted against the -log(10) p-value (x-axis). Phenotypes shown are significant after FDR-correction. The direction of the bar indicates the direction of association, where a right-hand bar indicates a positive association, and a left-hand bar a negative association. In contrast analyses (e.g., comparing response 1 vs. response 3), the direction of the bar indicates the direction of the association in relation to the answers contrasted. Specifically, in a 1 vs. 3 contrast, a bar pointing to the right indicates an association with response 3, while a bar pointing to the left indicates an association with response 1. The phenotype “Diagnosed Vascular problems diagnosed by doctor.0” equates to “No Vascular problems diagnosed”. Phenotypes are color coded by phenotype category.

**Figure 18:**
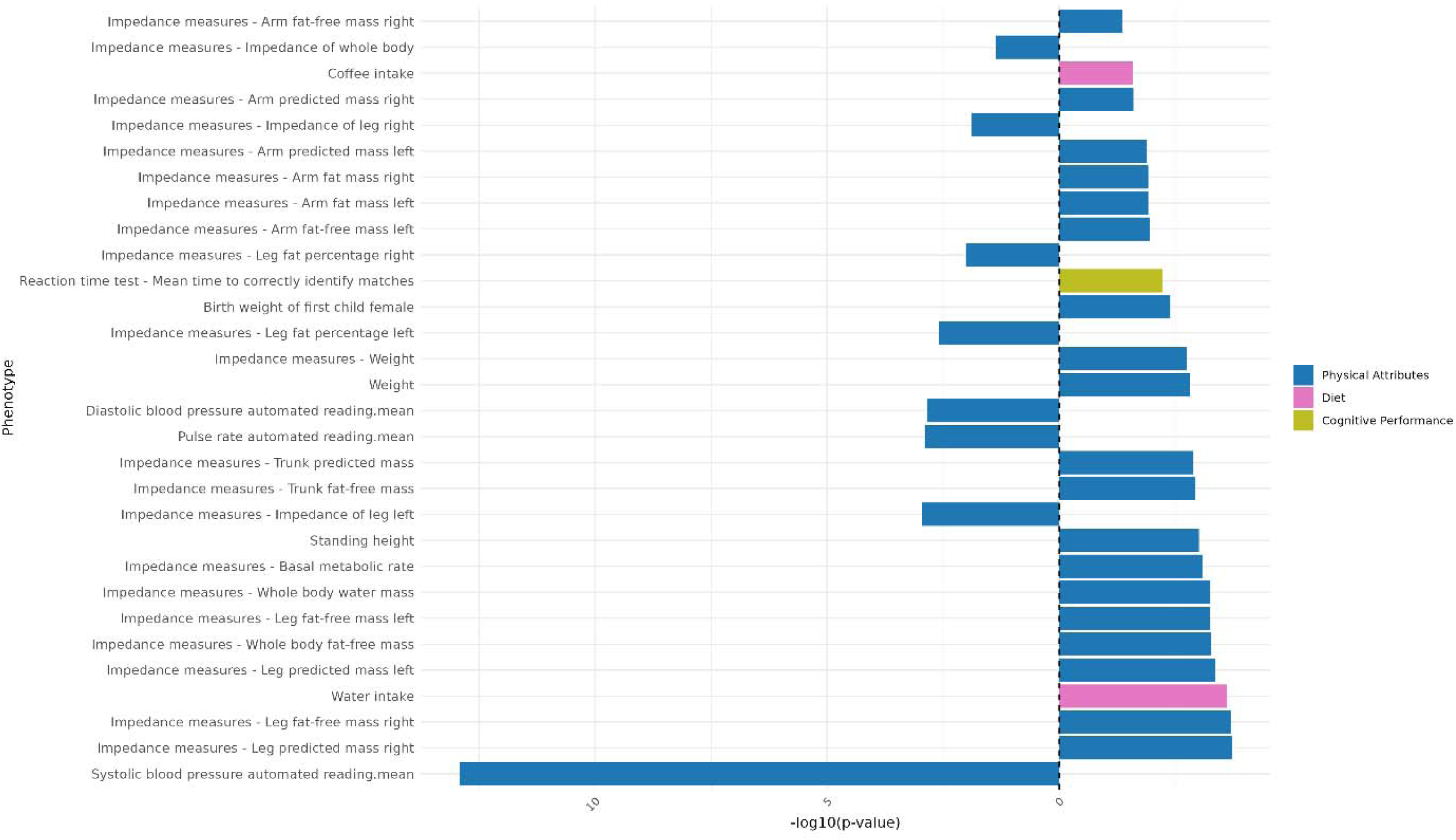
Genomic Independent Component 3 PCS association strength across continuous non-neuroimaging phenotypes. Phenotypes (y-axis) are plotted against the -log(10) p-value (x-axis). Phenotypes shown are significant after FDR-correction. The direction of the bar indicates the direction of association, where a right-hand bar indicates a positive association, and a left-hand bar a negative association. Phenotypes are color coded by phenotype category.

### Independent Component 4

**Figure 19:**
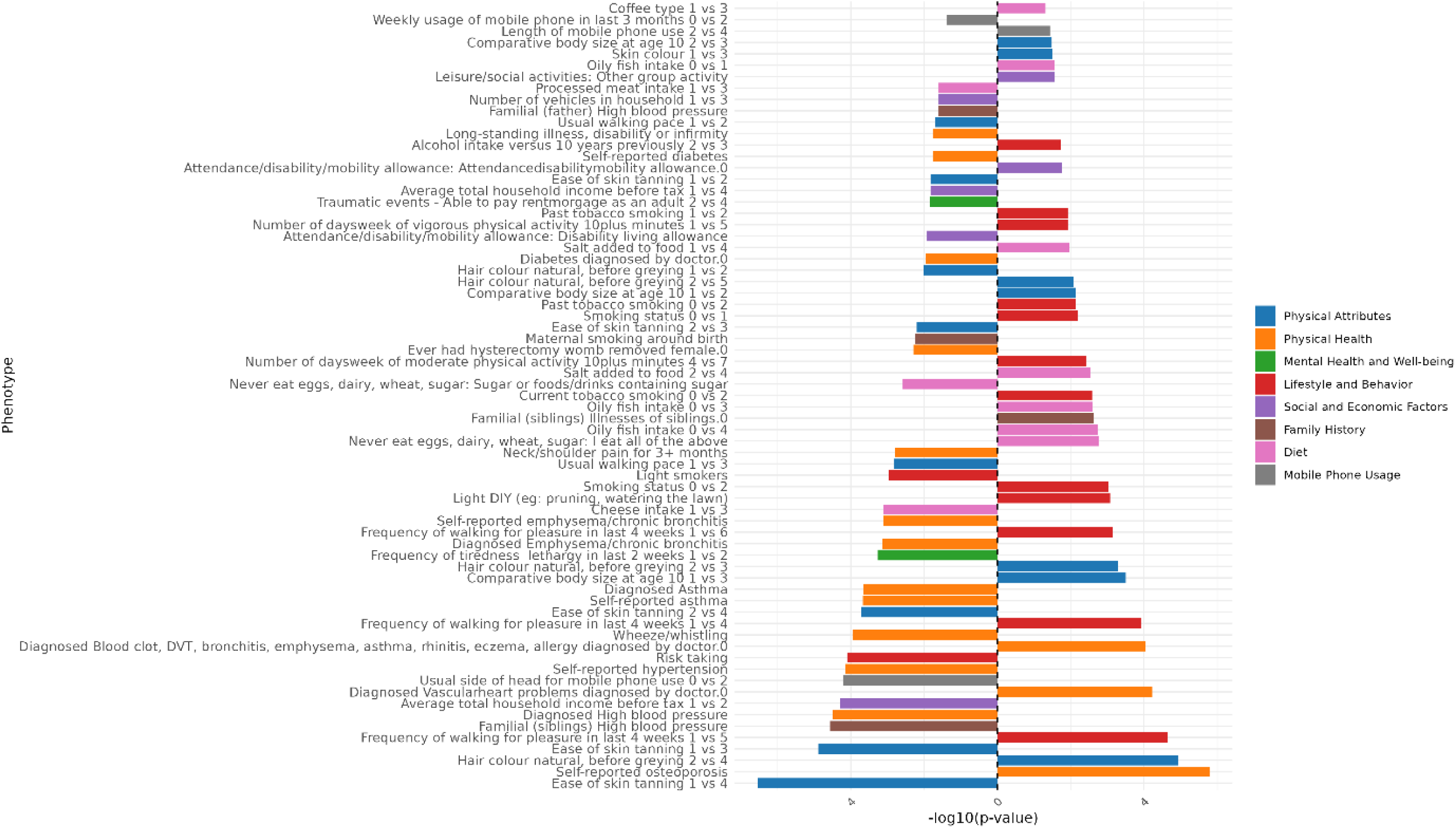
Genomic Independent Component 4 PCS association strength across binary/binarized non-neuroimaging phenotypes. Phenotypes (y-axis) are plotted against the -log(10) p-value (x-axis). Phenotypes shown are significant after FDR-correction. The direction of the bar indicates the direction of association, where a right-hand bar indicates a positive association, and a left-hand bar a negative association. In contrast analyses (e.g., comparing response 1 vs. response 3), the direction of the bar indicates the direction of the association in relation to the answers contrasted. Specifically, in a 1 vs. 3 contrast, a bar pointing to the right indicates an association with response 3, while a bar pointing to the left indicates an association with response 1. The phenotype “Diagnosed Vascular problems diagnosed by doctor.0” equates to “No Vascular problems diagnosed”. Phenotypes are color coded by phenotype category.

**Figure 20:**
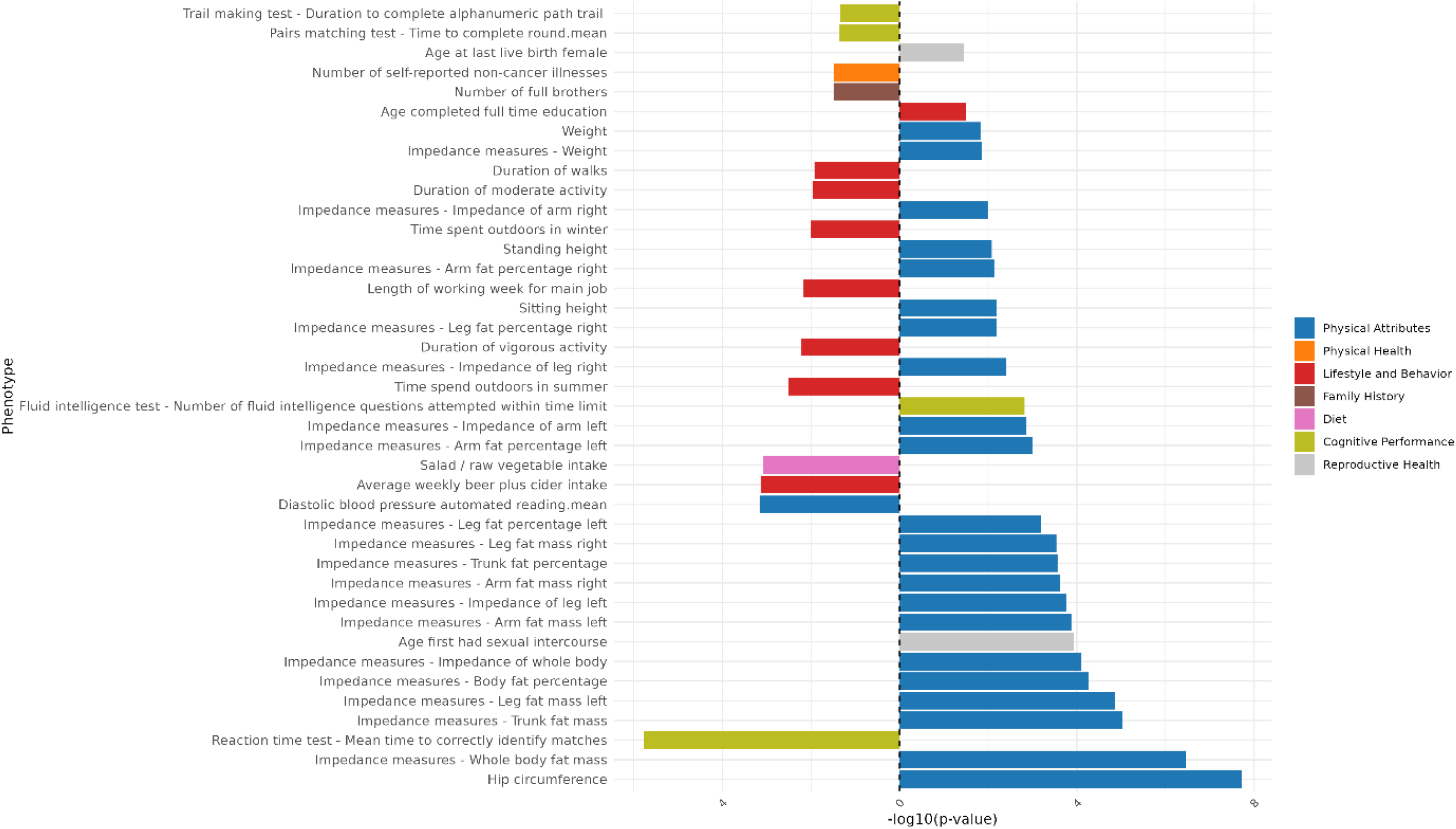
Genomic Independent Component 4 PCS association strength across continuous non-neuroimaging phenotypes. Phenotypes (y-axis) are plotted against the -log(10) p-value (x-axis). Phenotypes shown are significant after FDR-correction. The direction of the bar indicates the direction of association, where a right-hand bar indicates a positive association, and a left-hand bar a negative association. Phenotypes are color coded by phenotype category.

### Independent Component 5

**Figure 21:**
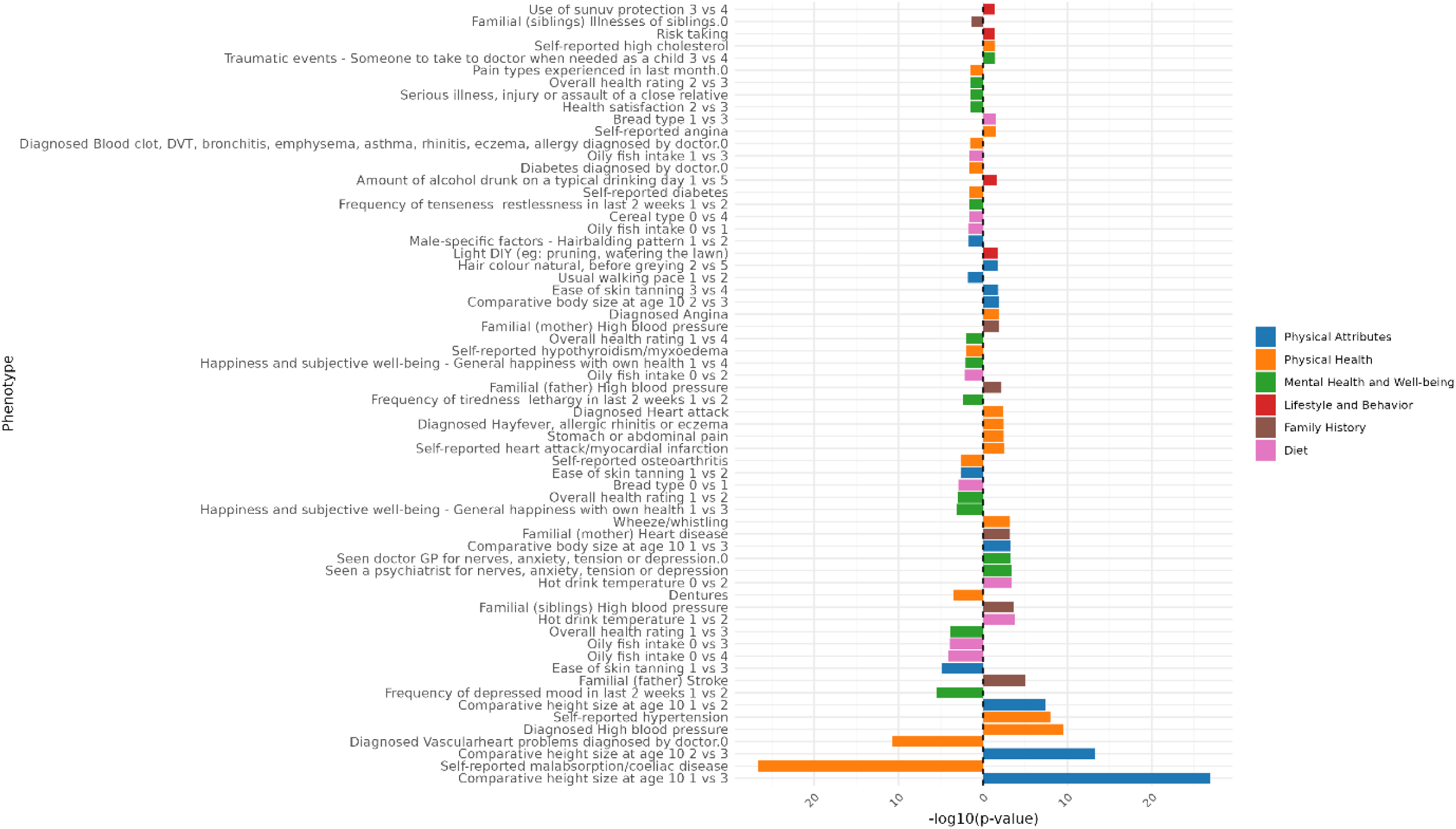
Genomic Independent Component 5 PCS association strength across binary/binarized non-neuroimaging phenotypes. Phenotypes (y-axis) are plotted against the -log(10) p-value (x-axis). Phenotypes shown are significant after FDR-correction. The direction of the bar indicates the direction of association, where a right-hand bar indicates a positive association, and a left-hand bar a negative association. In contrast analyses (e.g., comparing response 1 vs. response 3), the direction of the bar indicates the direction of the association in relation to the answers contrasted. Specifically, in a 1 vs. 3 contrast, a bar pointing to the right indicates an association with response 3, while a bar pointing to the left indicates an association with response 1. The phenotype “Diagnosed Vascular problems diagnosed by doctor.0” equates to “No Vascular problems diagnosed”. Phenotypes are color coded by phenotype category.

**Figure 22:**
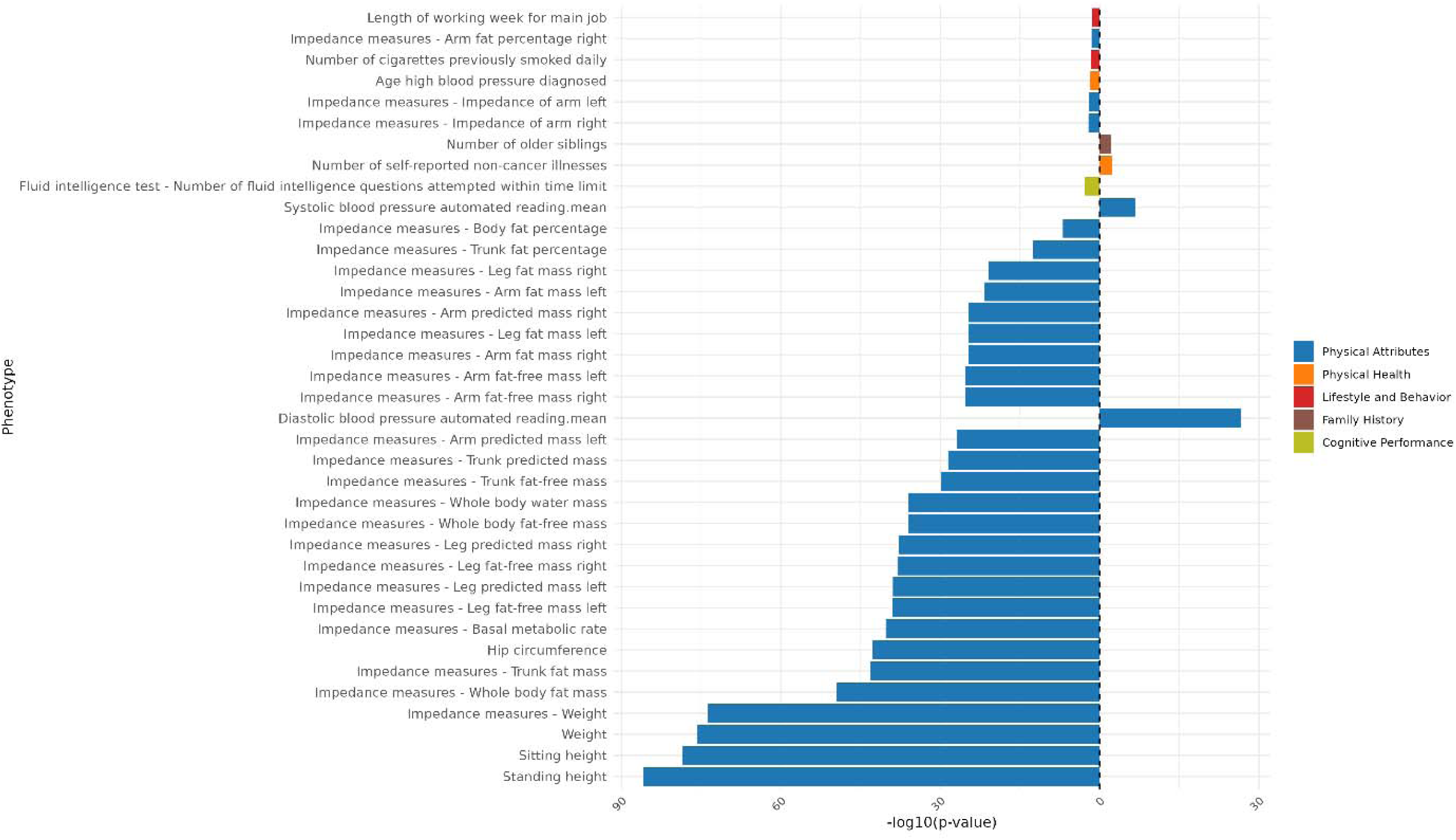
Genomic Independent Component 5 PCS association strength across continuous non-neuroimaging phenotypes. Phenotypes (y-axis) are plotted against the -log(10) p-value (x-axis). Phenotypes shown are significant after FDR-correction. The direction of the bar indicates the direction of association, where a right-hand bar indicates a positive association, and a left-hand bar a negative association. Phenotypes are color coded by phenotype category.

### Independent Component 6

**Figure 23:**
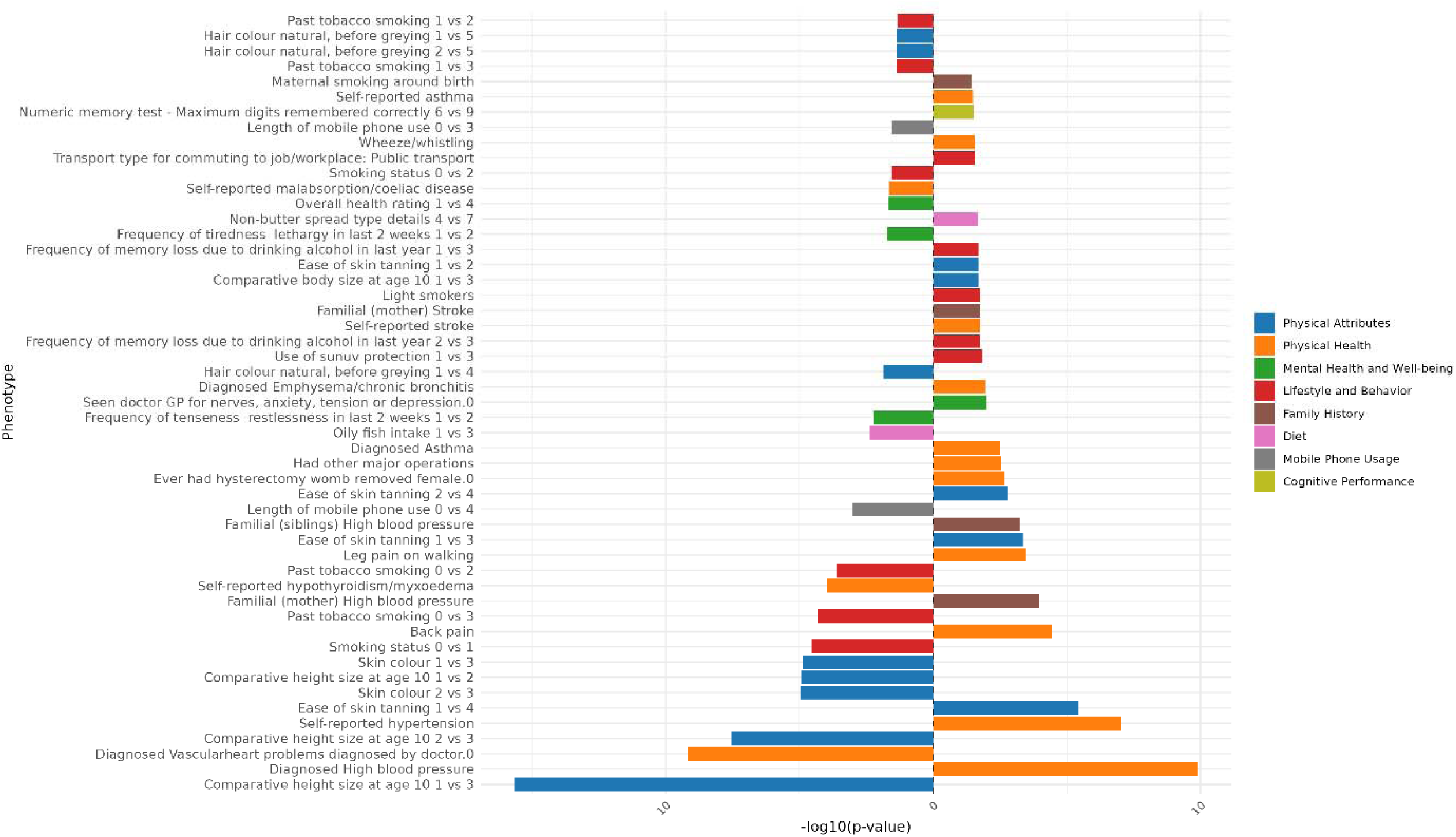
Genomic Independent Component 6 PCS association strength across binary/binarized non-neuroimaging phenotypes. Phenotypes (y-axis) are plotted against the -log(10) p-value (x-axis). Phenotypes shown are significant after FDR-correction. The direction of the bar indicates the direction of association, where a right-hand bar indicates a positive association, and a left-hand bar a negative association. In contrast analyses (e.g., comparing response 1 vs. response 3), the direction of the bar indicates the direction of the association in relation to the answers contrasted. Specifically, in a 1 vs. 3 contrast, a bar pointing to the right indicates an association with response 3, while a bar pointing to the left indicates an association with response 1. The phenotype “Diagnosed Vascular problems diagnosed by doctor.0” equates to “No Vascular problems diagnosed”. Phenotypes are color coded by phenotype category.

**Figure 24:**
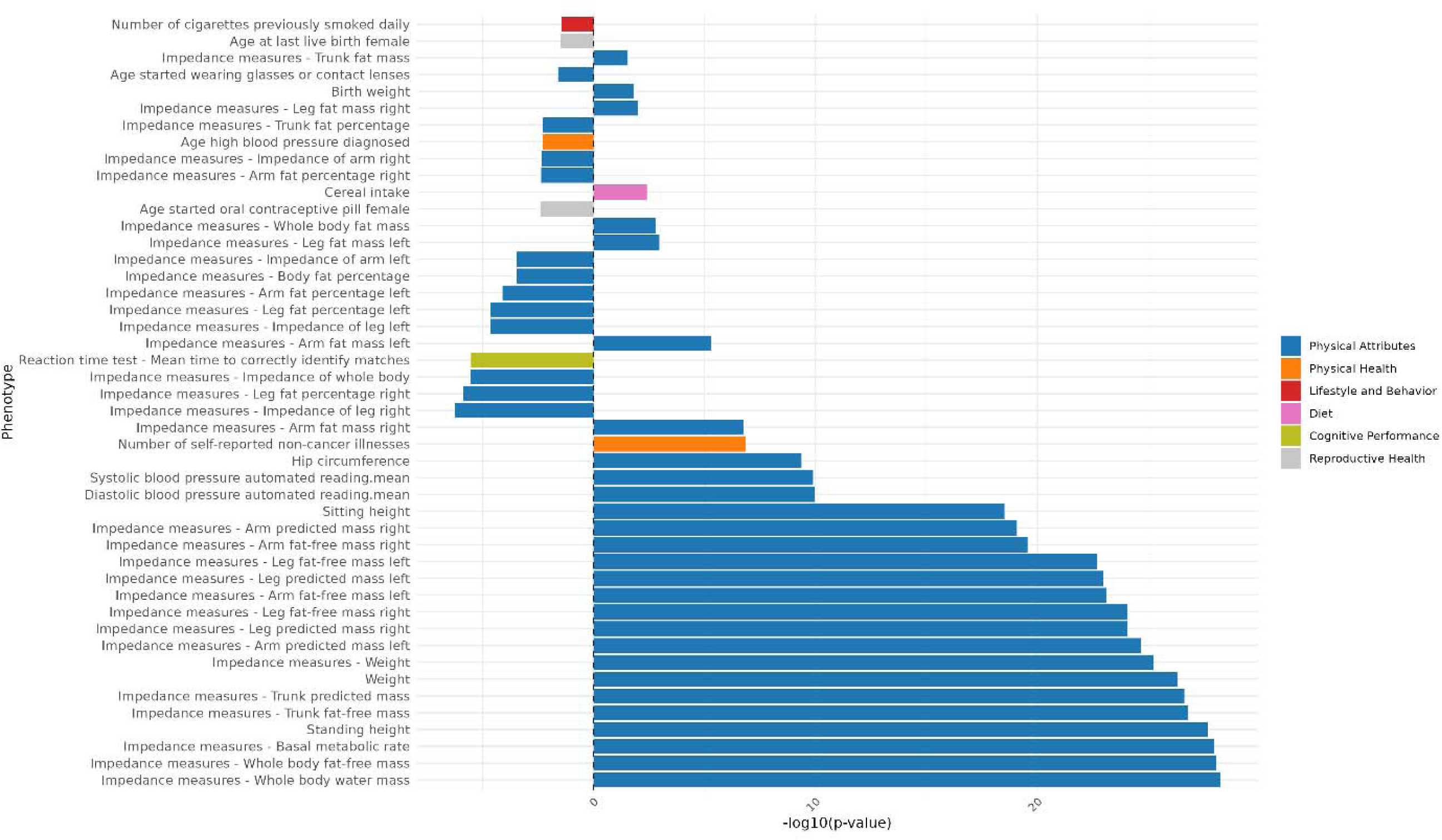
Genomic Independent Component 6 PCS association strength across continuous non-neuroimaging phenotypes. Phenotypes (y-axis) are plotted against the -log(10) p-value (x-axis). Phenotypes shown are significant after FDR-correction. The direction of the bar indicates the direction of association, where a right-hand bar indicates a positive association, and a left-hand bar a negative association. Phenotypes are color coded by phenotype category.

### Independent Component 7

**Figure 25:**
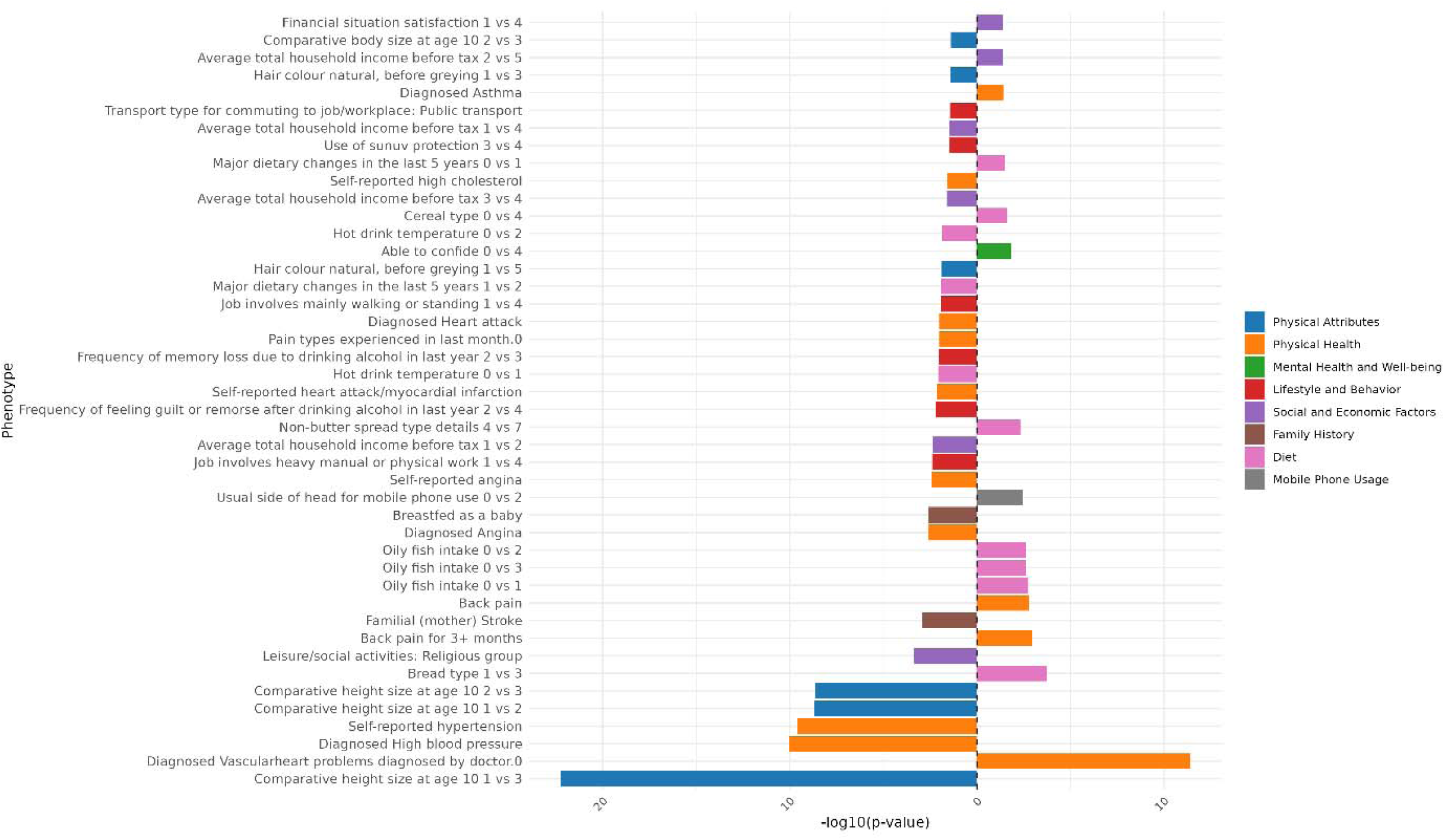
Genomic Independent Component 7 PCS association strength across binary/binarized non-neuroimaging phenotypes. Phenotypes (y-axis) are plotted against the -log(10) p-value (x-axis). Phenotypes shown are significant after FDR-correction. The direction of the bar indicates the direction of association, where a right-hand bar indicates a positive association, and a left-hand bar a negative association. In contrast analyses (e.g., comparing response 1 vs. response 3), the direction of the bar indicates the direction of the association in relation to the answers contrasted. Specifically, in a 1 vs. 3 contrast, a bar pointing to the right indicates an association with response 3, while a bar pointing to the left indicates an association with response 1. The phenotype “Diagnosed Vascular problems diagnosed by doctor.0” equates to “No Vascular problems diagnosed”. Phenotypes are color coded by phenotype category.

**Figure 26:**
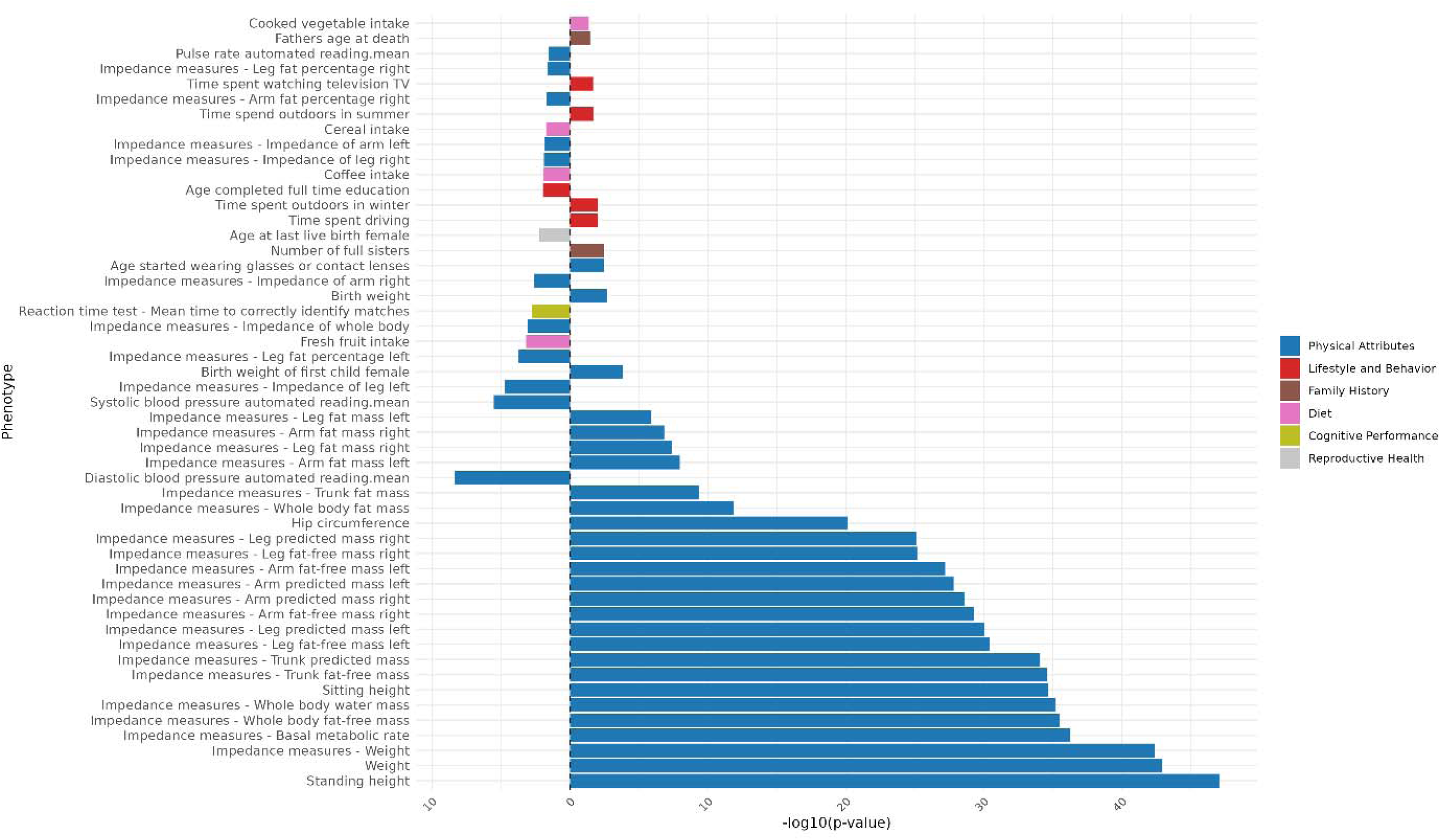
Genomic Independent Component 7 PCS association strength across continuous non-neuroimaging phenotypes. Phenotypes (y-axis) are plotted against the -log(10) p-value (x-axis). Phenotypes shown are significant after FDR-correction. The direction of the bar indicates the direction of association, where a right-hand bar indicates a positive association, and a left-hand bar a negative association. Phenotypes are color coded by phenotype category.

### Independent Component 8

**Figure 27:**
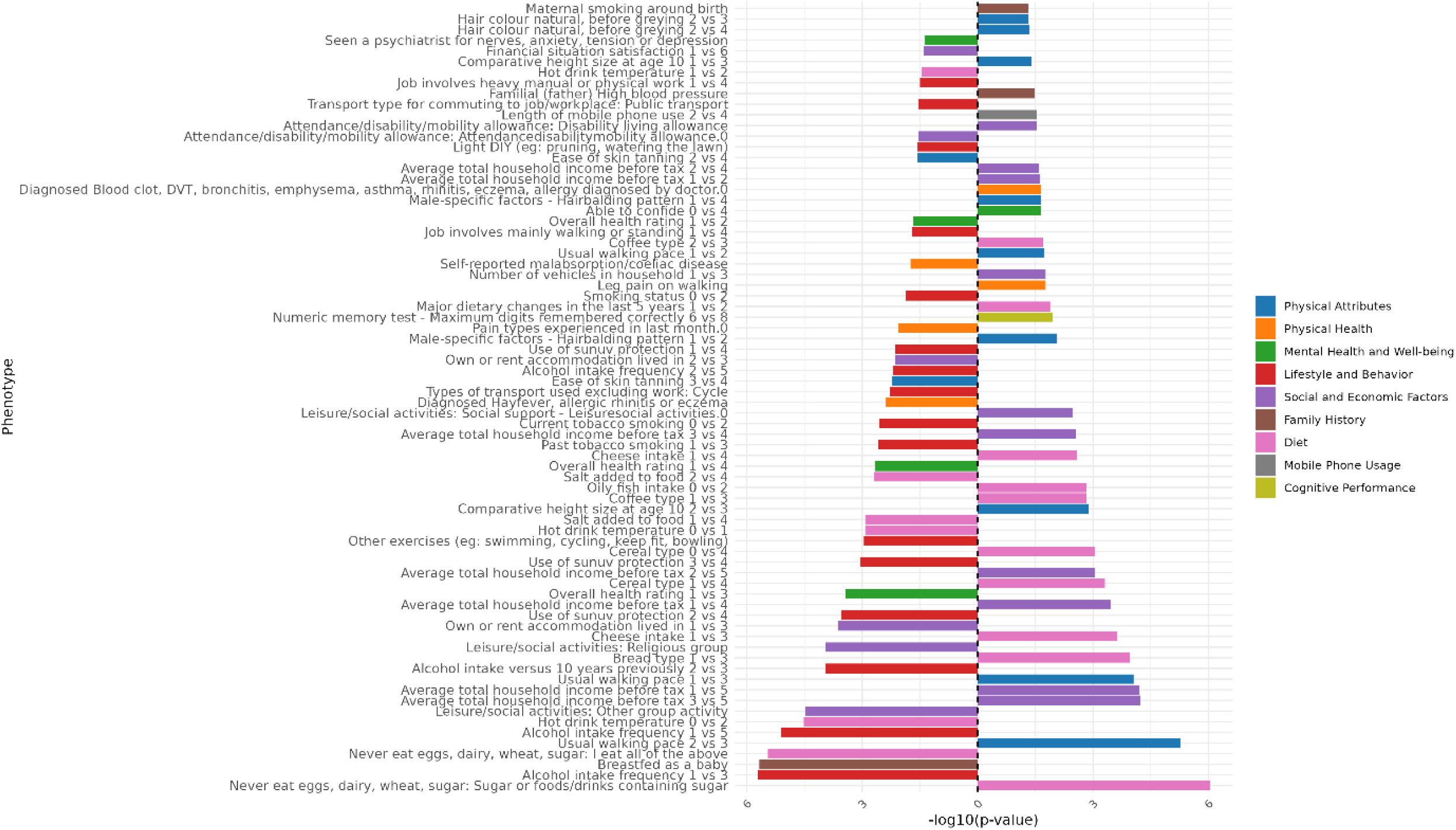
Genomic Independent Component 8 PCS association strength across binary/binarized non-neuroimaging phenotypes. Phenotypes (y-axis) are plotted against the -log(10) p-value (x-axis). Phenotypes shown are significant after FDR-correction. The direction of the bar indicates the direction of association, where a right-hand bar indicates a positive association, and a left-hand bar a negative association. In contrast analyses (e.g., comparing response 1 vs. response 3), the direction of the bar indicates the direction of the association in relation to the answers contrasted. Specifically, in a 1 vs. 3 contrast, a bar pointing to the right indicates an association with response 3, while a bar pointing to the left indicates an association with response 1. The phenotype “Diagnosed Vascular problems diagnosed by doctor.0” equates to “No Vascular problems diagnosed”. Phenotypes are color coded by phenotype category.

**Figure 28:**
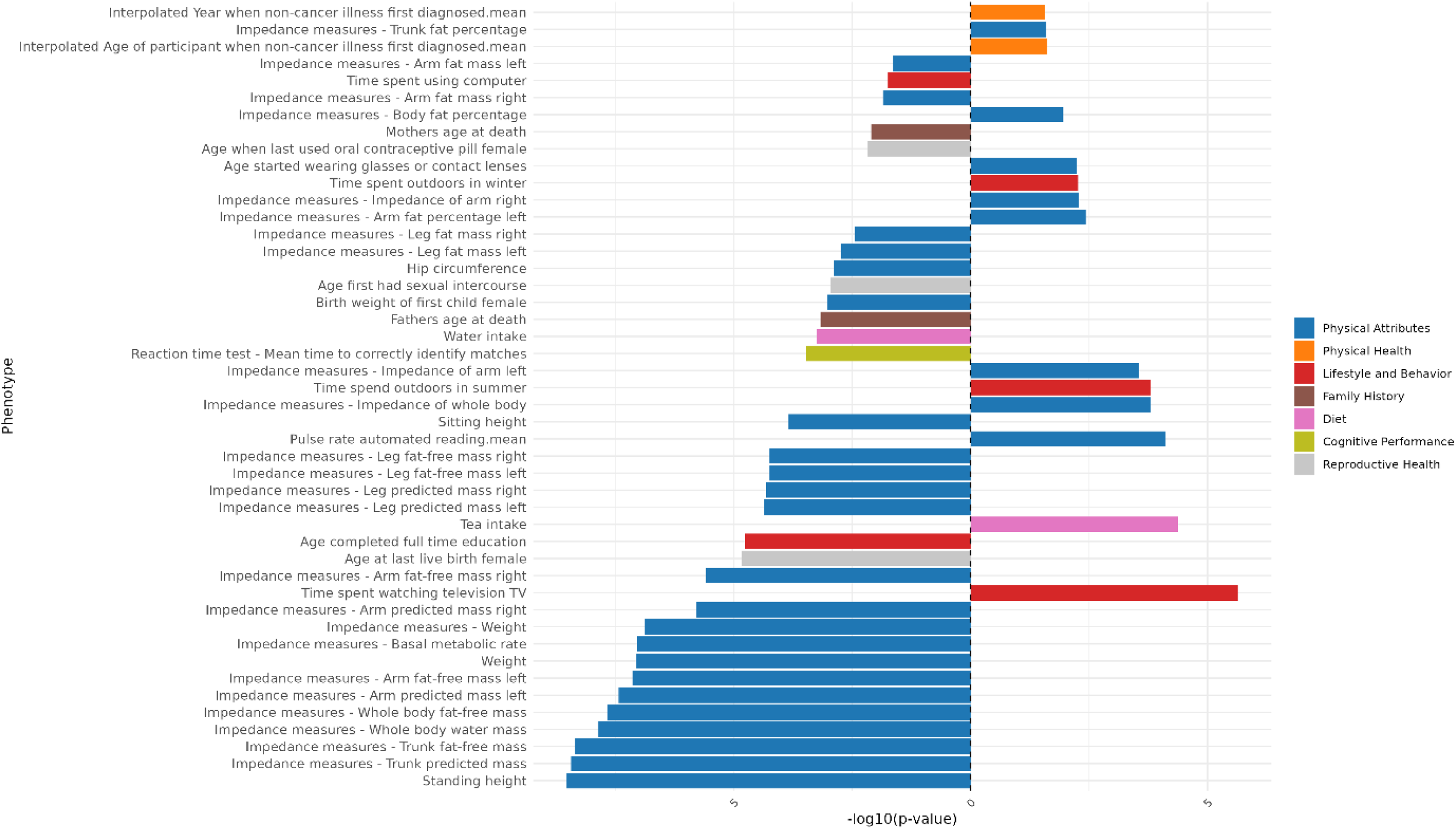
Genomic Independent Component 8 PCS association strength across continuous non-neuroimaging phenotypes. Phenotypes (y-axis) are plotted against the -log(10) p-value (x-axis). Phenotypes shown are significant after FDR-correction. The direction of the bar indicates the direction of association, where a right-hand bar indicates a positive association, and a left-hand bar a negative association. Phenotypes are color coded by phenotype category.

### Independent Component 9

**Figure 29:**
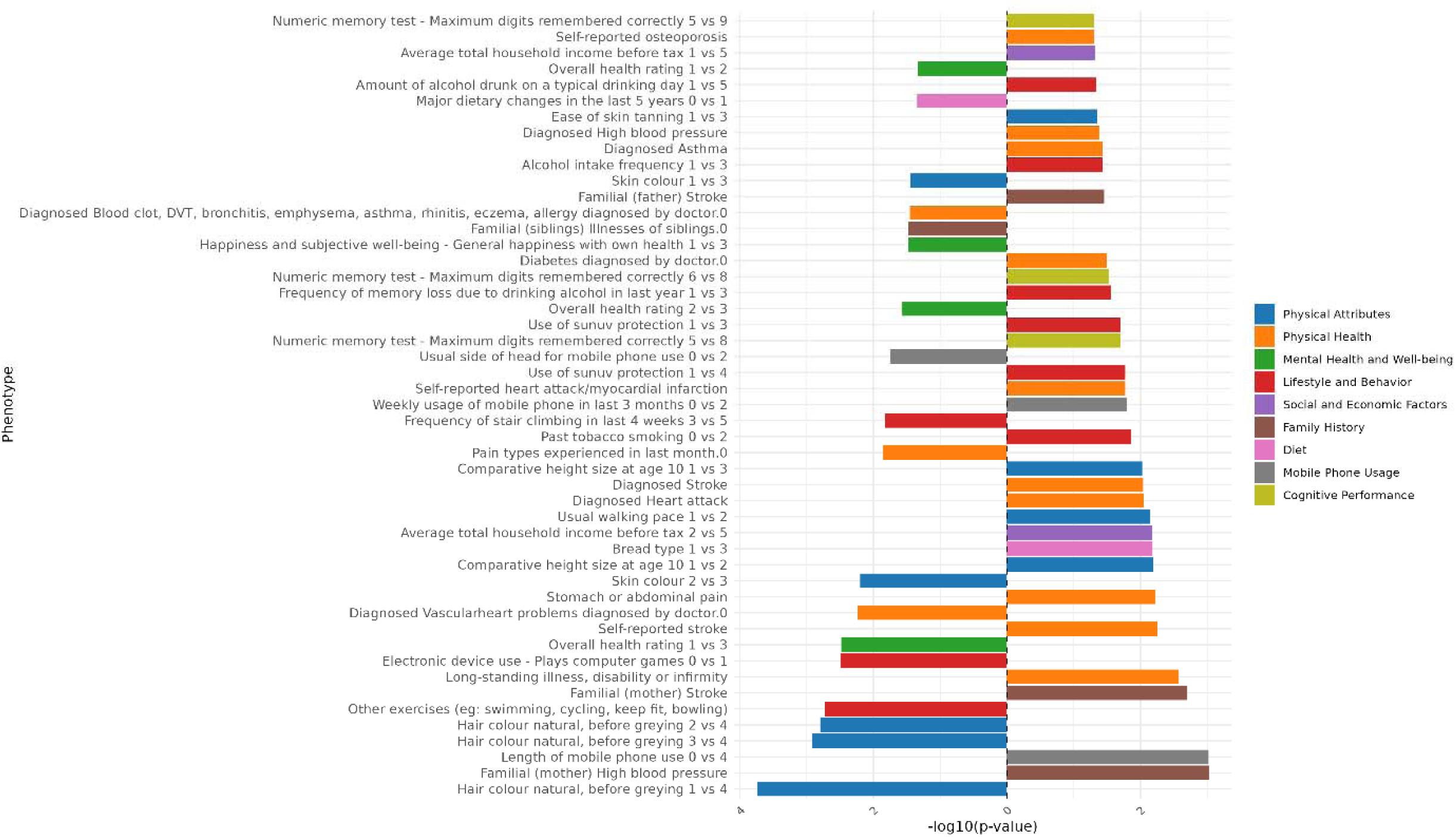
Genomic Independent Component 9 PCS association strength across binary/binarized non-neuroimaging phenotypes. Phenotypes (y-axis) are plotted against the -log(10) p-value (x-axis). Phenotypes shown are significant after FDR-correction. The direction of the bar indicates the direction of association, where a right-hand bar indicates a positive association, and a left-hand bar a negative association. In contrast analyses (e.g., comparing response 1 vs. response 3), the direction of the bar indicates the direction of the association in relation to the answers contrasted. Specifically, in a 1 vs. 3 contrast, a bar pointing to the right indicates an association with response 3, while a bar pointing to the left indicates an association with response 1. The phenotype “Diagnosed Vascular problems diagnosed by doctor.0” equates to “No Vascular problems diagnosed”. Phenotypes are color coded by phenotype category.

**Figure 30:**
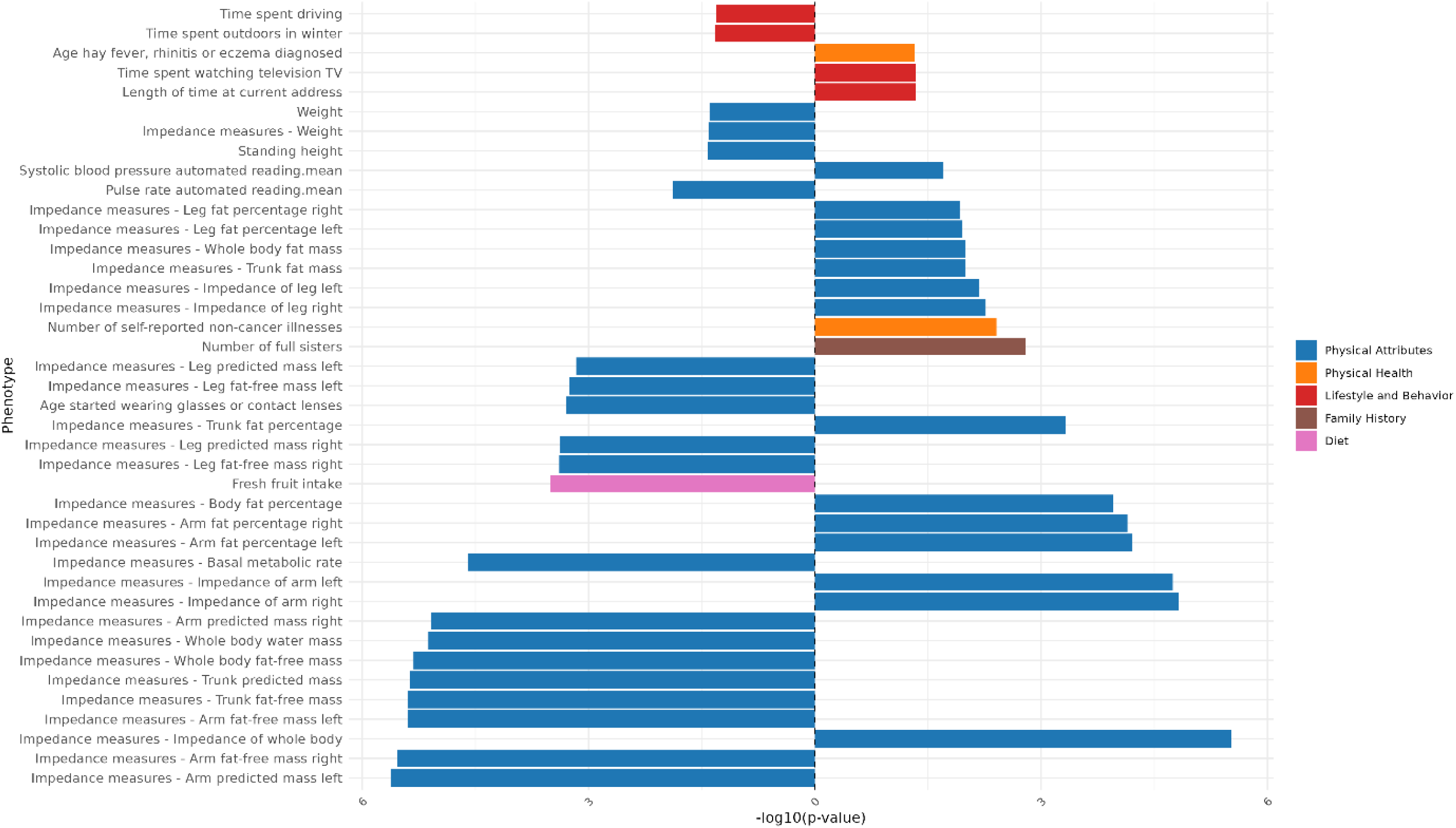
Genomic Independent Component 9 PCS association strength across continuous non-neuroimaging phenotypes. Phenotypes (y-axis) are plotted against the -log(10) p-value (x-axis). Phenotypes shown are significant after FDR-correction. The direction of the bar indicates the direction of association, where a right-hand bar indicates a positive association, and a left-hand bar a negative association. Phenotypes are color coded by phenotype category.

### Independent Component 10

**Figure 31:**
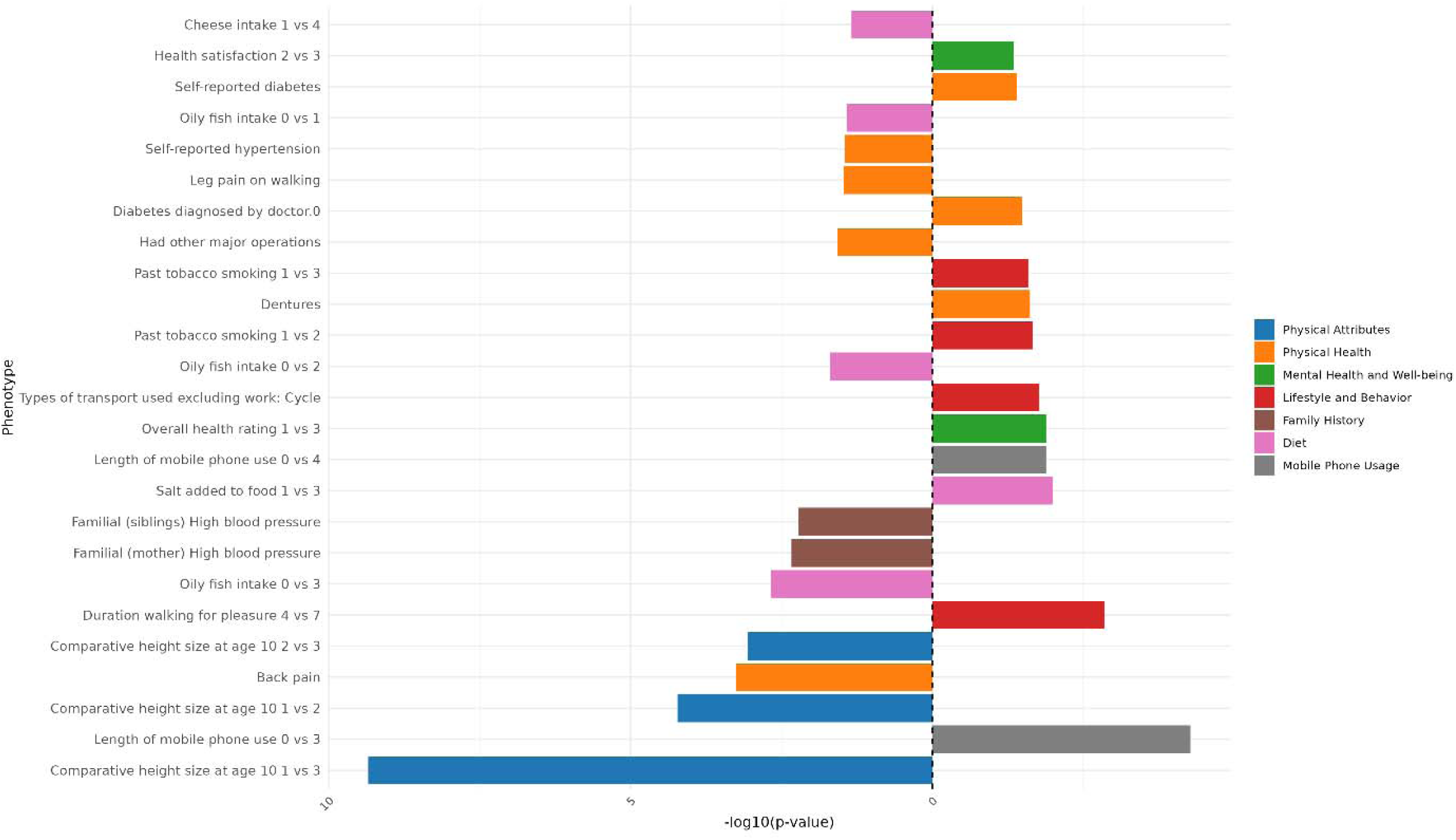
Genomic Independent Component 10 PCS association strength across binary/binarized non-neuroimaging phenotypes. Phenotypes (y-axis) are plotted against the -log(10) p-value (x-axis). Phenotypes shown are significant after FDR-correction. The direction of the bar indicates the direction of association, where a right-hand bar indicates a positive association, and a left-hand bar a negative association. In contrast analyses (e.g., comparing response 1 vs. response 3), the direction of the bar indicates the direction of the association in relation to the answers contrasted. Specifically, in a 1 vs. 3 contrast, a bar pointing to the right indicates an association with response 3, while a bar pointing to the left indicates an association with response 1. The phenotype “Diagnosed Vascular problems diagnosed by doctor.0” equates to “No Vascular problems diagnosed”. Phenotypes are color coded by phenotype category.

**Figure 32:**
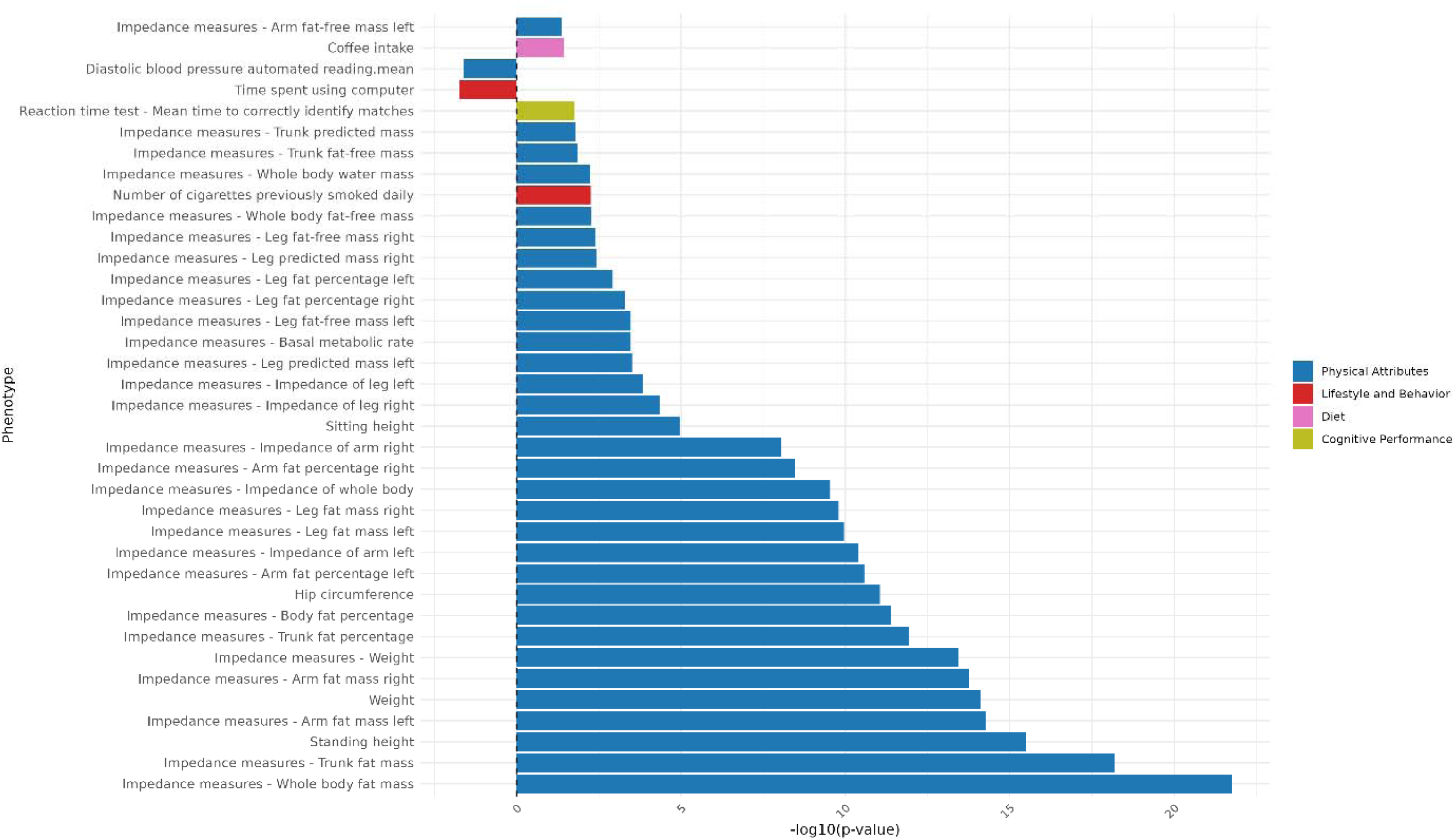
Genomic Independent Component 10 PCS association strength across continuous non-neuroimaging phenotypes. Phenotypes (y-axis) are plotted against the -log(10) p-value (x-axis). Phenotypes shown are significant after FDR-correction. The direction of the bar indicates the direction of association, where a right-hand bar indicates a positive association, and a left-hand bar a negative association. Phenotypes are color coded by phenotype category.

### Independent Component 11

**Figure 33:**
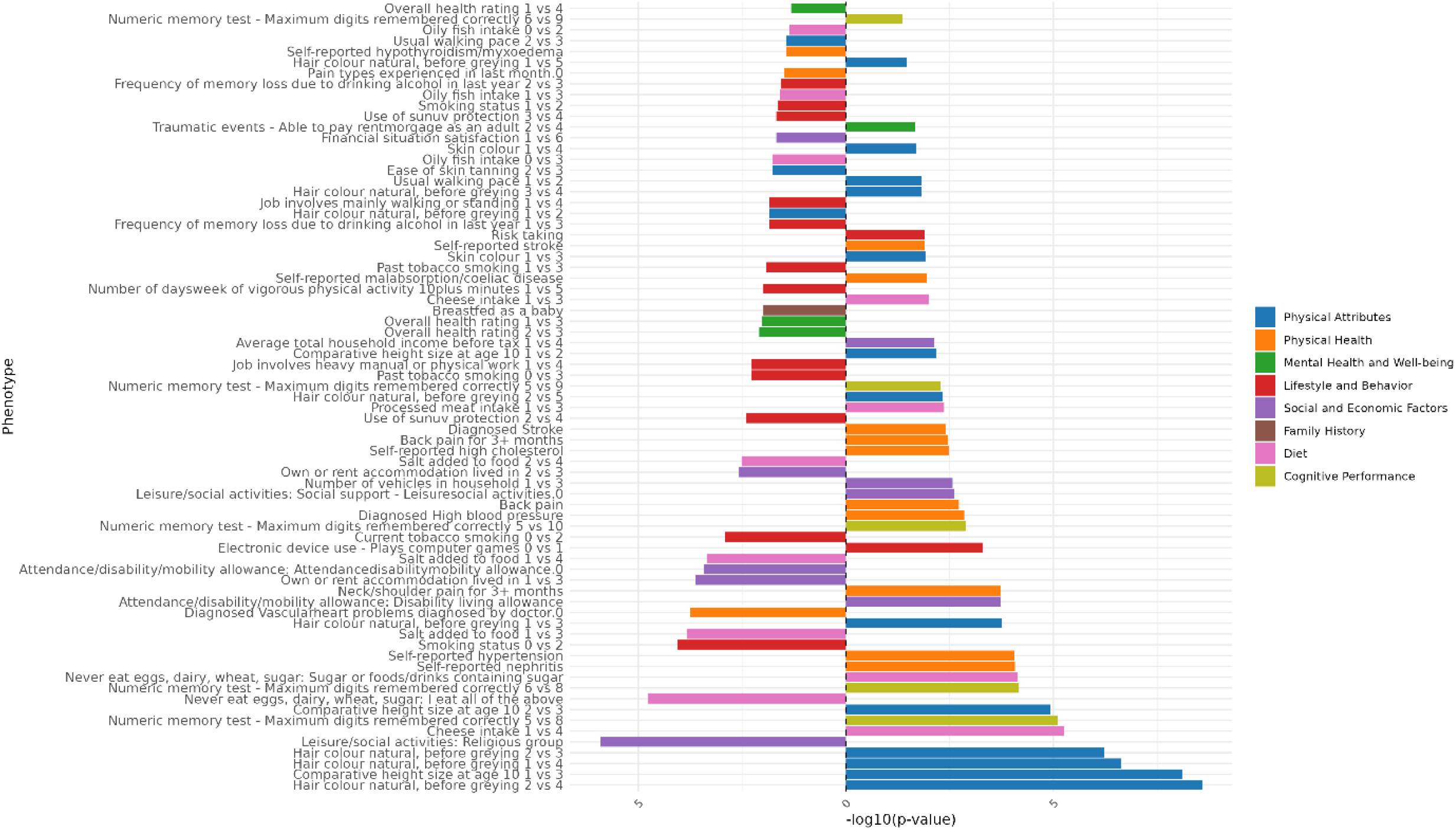
Genomic Independent Component 11 PCS association strength across binary/binarized non-neuroimaging phenotypes. Phenotypes (y-axis) are plotted against the -log(10) p-value (x-axis). Phenotypes shown are significant after FDR-correction. The direction of the bar indicates the direction of association, where a right-hand bar indicates a positive association, and a left-hand bar a negative association. In contrast analyses (e.g., comparing response 1 vs. response 3), the direction of the bar indicates the direction of the association in relation to the answers contrasted. Specifically, in a 1 vs. 3 contrast, a bar pointing to the right indicates an association with response 3, while a bar pointing to the left indicates an association with response 1. The phenotype “Diagnosed Vascular problems diagnosed by doctor.0” equates to “No Vascular problems diagnosed”. Phenotypes are color coded by phenotype category.

**Figure 34:**
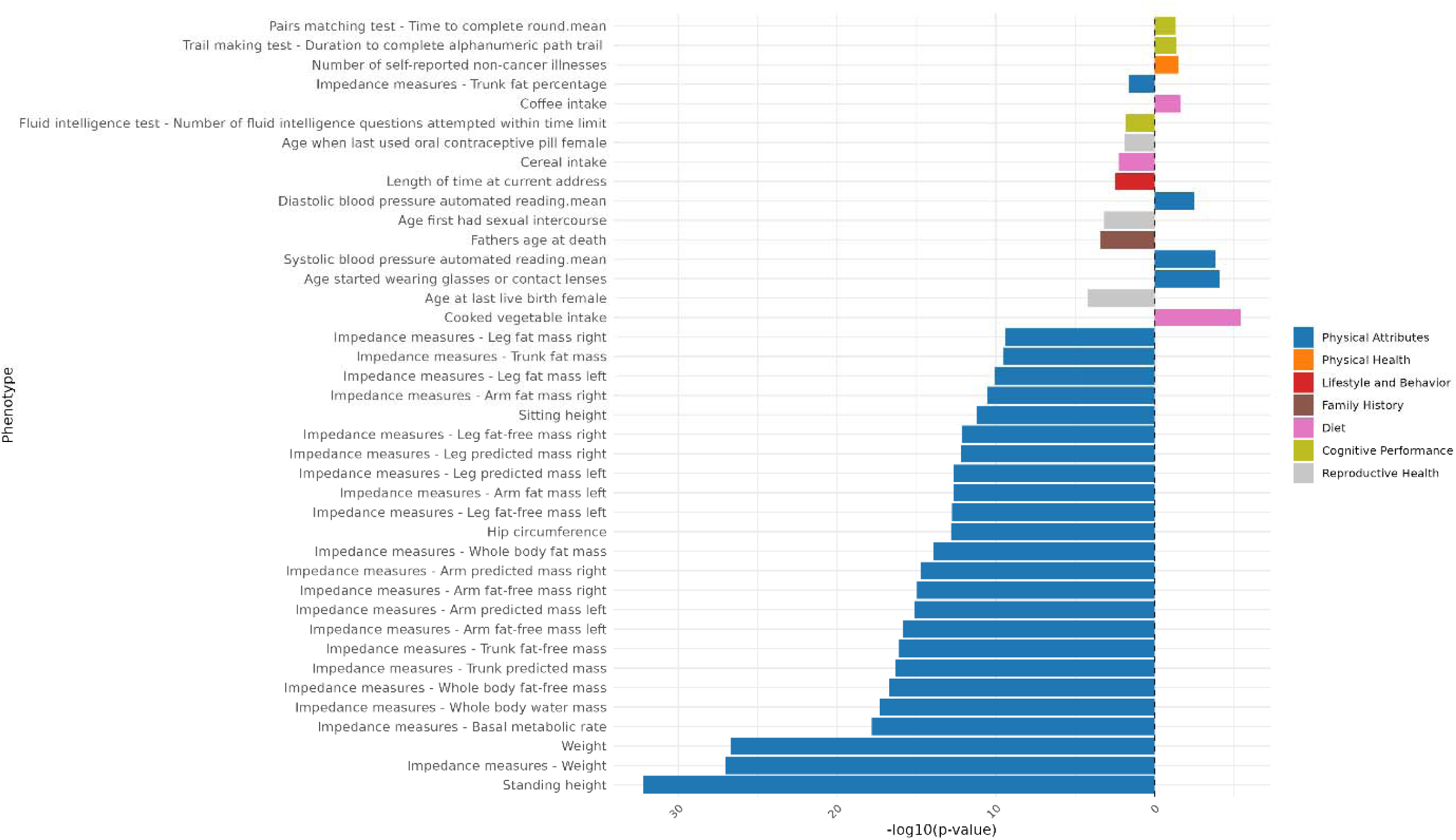
Genomic Independent Component 11 PCS association strength across continuous non-neuroimaging phenotypes. Phenotypes (y-axis) are plotted against the -log(10) p-value (x-axis). Phenotypes shown are significant after FDR-correction. The direction of the bar indicates the direction of association, where a right-hand bar indicates a positive association, and a left-hand bar a negative association. Phenotypes are color coded by phenotype category.

### Independent Component 12

**Figure 35:**
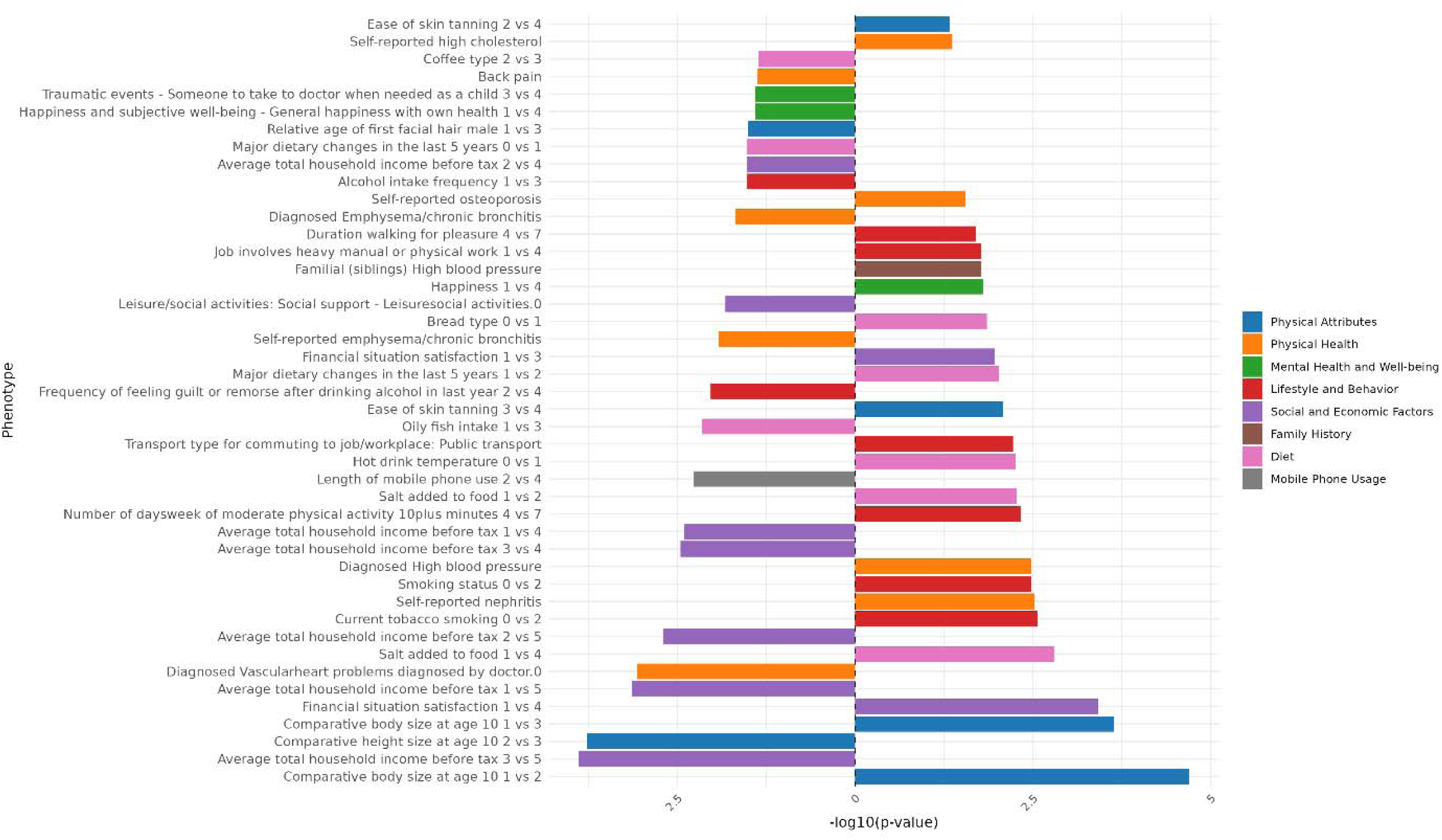
Genomic Independent Component 12 PCS association strength across binary/binarized non-neuroimaging phenotypes. Phenotypes (y-axis) are plotted against the -log(10) p-value (x-axis). Phenotypes shown are significant after FDR-correction. The direction of the bar indicates the direction of association, where a right-hand bar indicates a positive association, and a left-hand bar a negative association. In contrast analyses (e.g., comparing response 1 vs. response 3), the direction of the bar indicates the direction of the association in relation to the answers contrasted. Specifically, in a 1 vs. 3 contrast, a bar pointing to the right indicates an association with response 3, while a bar pointing to the left indicates an association with response 1. The phenotype “Diagnosed Vascular problems diagnosed by doctor.0” equates to “No Vascular problems diagnosed”. Phenotypes are color coded by phenotype category.

**Figure 36:**
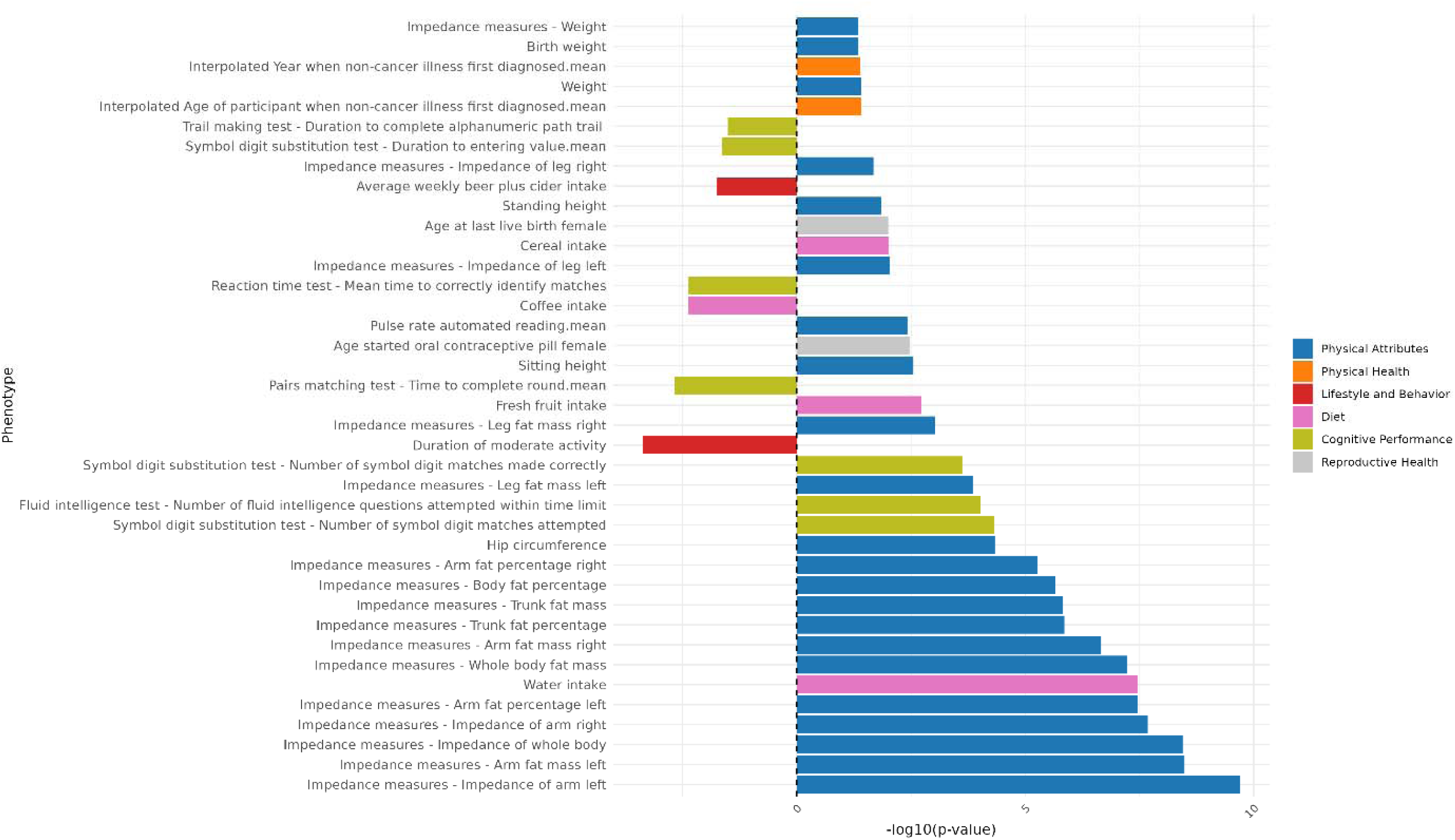
Genomic Independent Component 12 PCS association strength across continuous non-neuroimaging phenotypes. Phenotypes (y-axis) are plotted against the -log(10) p-value (x-axis). Phenotypes shown are significant after FDR-correction. The direction of the bar indicates the direction of association, where a right-hand bar indicates a positive association, and a left-hand bar a negative association. Phenotypes are color coded by phenotype category.

### Independent Component 13

**Figure 37:**
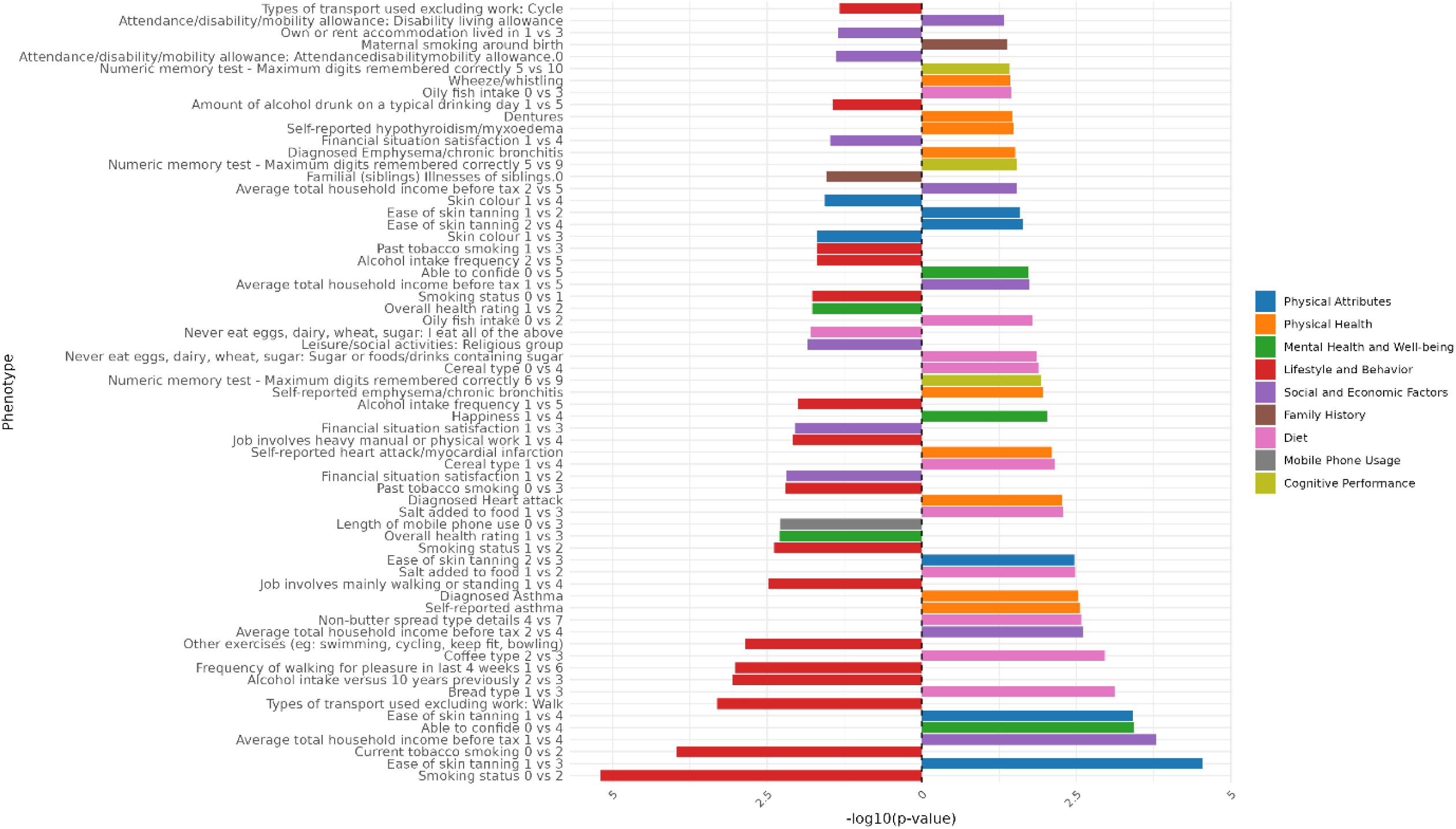
Genomic Independent Component 13 PCS association strength across binary/binarized non-neuroimaging phenotypes. Phenotypes (y-axis) are plotted against the -log(10) p-value (x-axis). Phenotypes shown are significant after FDR-correction. The direction of the bar indicates the direction of association, where a right-hand bar indicates a positive association, and a left-hand bar a negative association. In contrast analyses (e.g., comparing response 1 vs. response 3), the direction of the bar indicates the direction of the association in relation to the answers contrasted. Specifically, in a 1 vs. 3 contrast, a bar pointing to the right indicates an association with response 3, while a bar pointing to the left indicates an association with response 1. The phenotype “Diagnosed Vascular problems diagnosed by doctor.0” equates to “No Vascular problems diagnosed”. Phenotypes are color coded by phenotype category.

**Figure 38:**
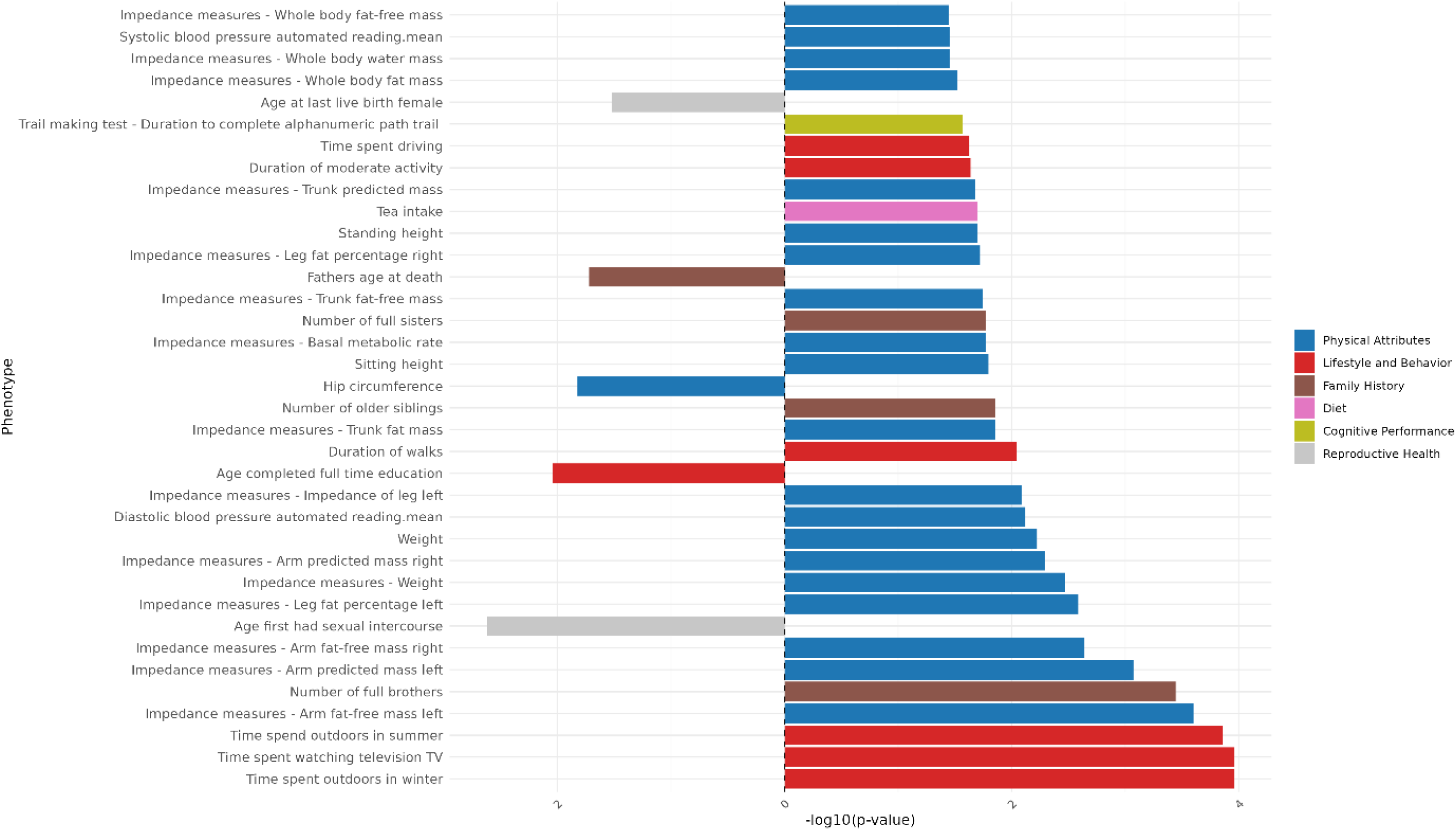
Genomic Independent Component 13 PCS association strength across continuous non-neuroimaging phenotypes. Phenotypes (y-axis) are plotted against the -log(10) p-value (x-axis). Phenotypes shown are significant after FDR-correction. The direction of the bar indicates the direction of association, where a right-hand bar indicates a positive association, and a left-hand bar a negative association. Phenotypes are color coded by phenotype category.

### Independent Component 14

**Figure 39:**
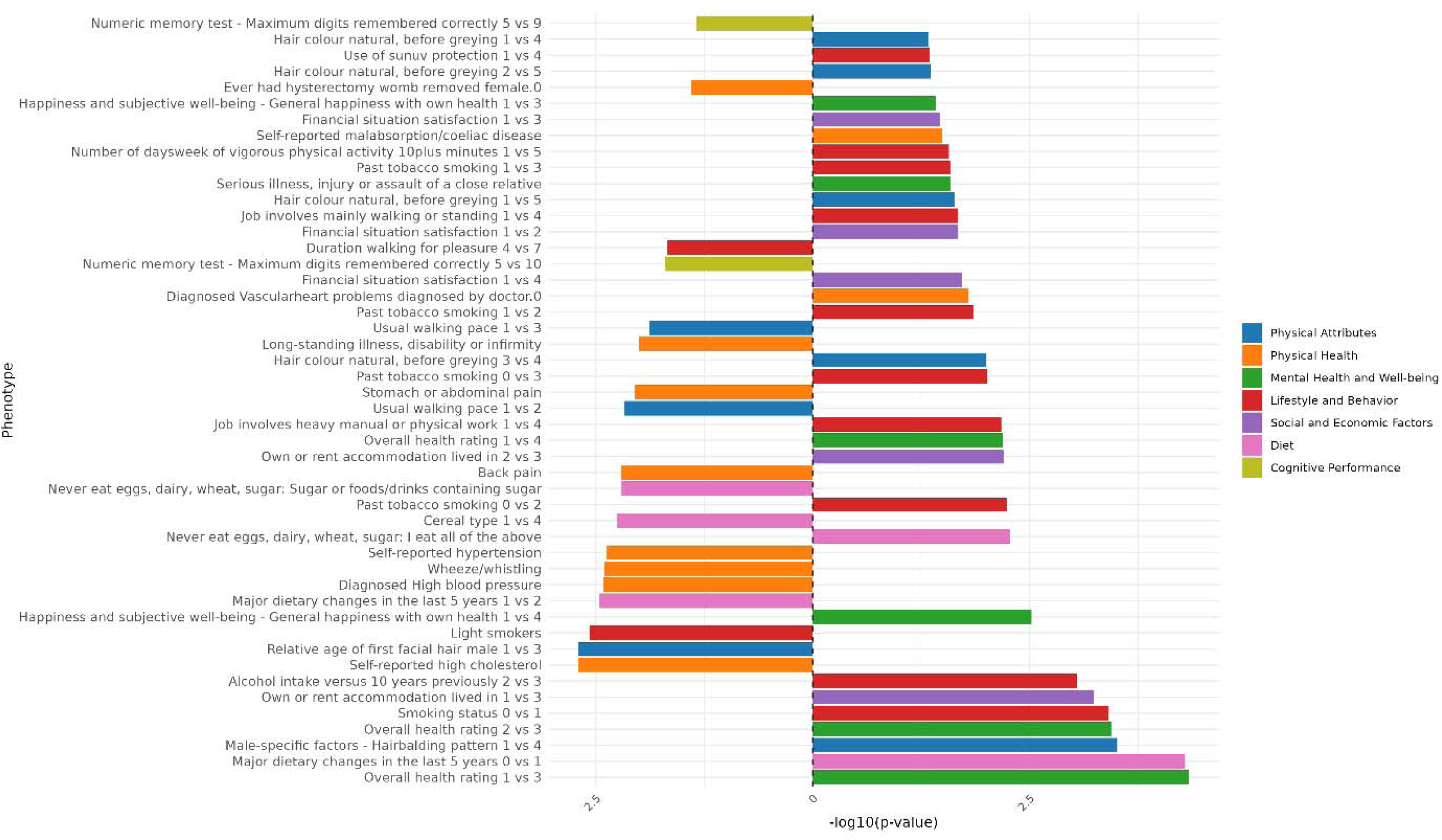
Genomic Independent Component 14 PCS association strength across binary/binarized non-neuroimaging phenotypes. Phenotypes (y-axis) are plotted against the -log(10) p-value (x-axis). Phenotypes shown are significant after FDR-correction. The direction of the bar indicates the direction of association, where a right-hand bar indicates a positive association, and a left-hand bar a negative association. In contrast analyses (e.g., comparing response 1 vs. response 3), the direction of the bar indicates the direction of the association in relation to the answers contrasted. Specifically, in a 1 vs. 3 contrast, a bar pointing to the right indicates an association with response 3, while a bar pointing to the left indicates an association with response 1. The phenotype “Diagnosed Vascular problems diagnosed by doctor.0” equates to “No Vascular problems diagnosed”. Phenotypes are color coded by phenotype category.

**Figure 40:**
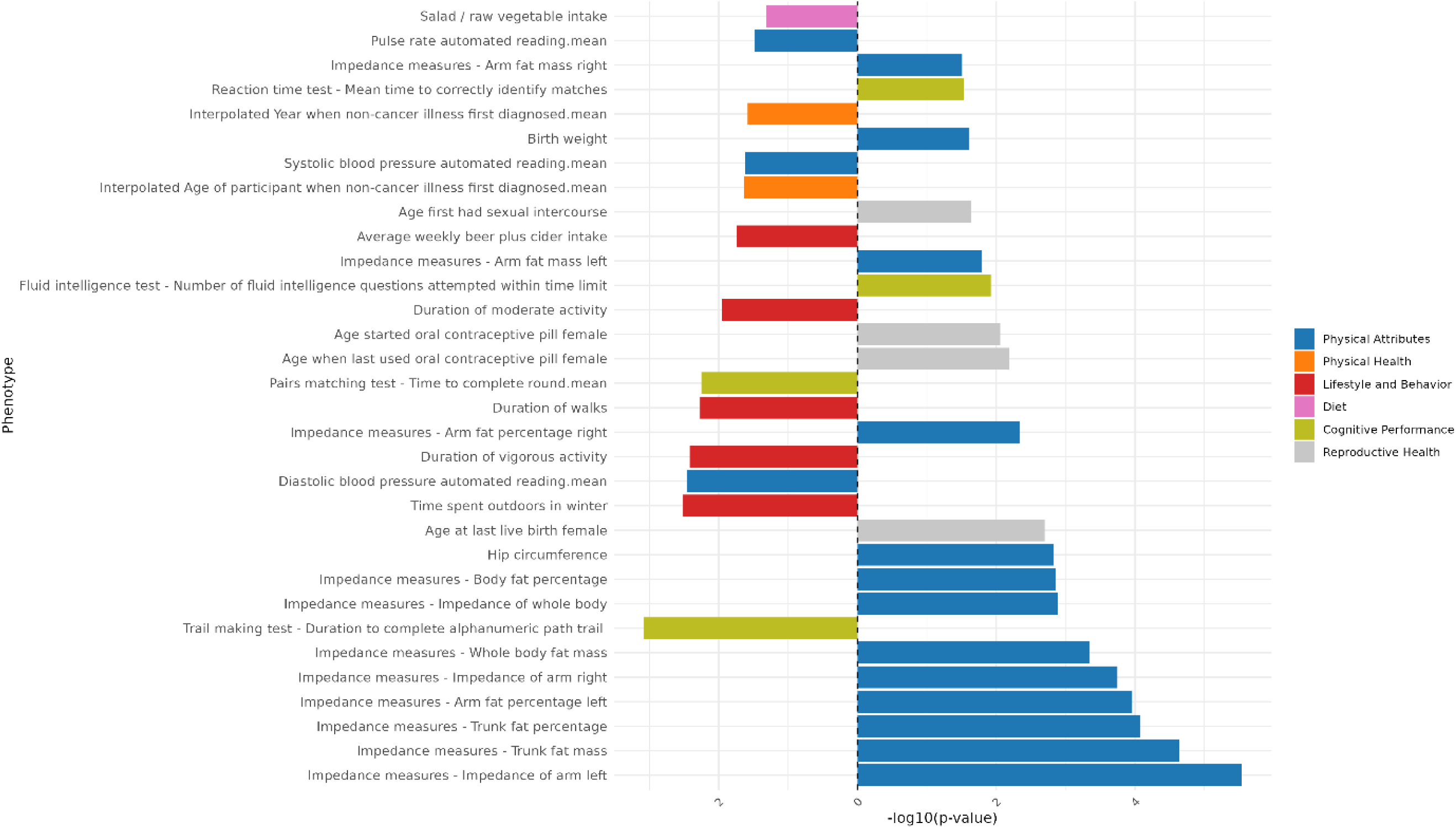
Genomic Independent Component 14 PCS association strength across continuous non-neuroimaging phenotypes. Phenotypes (y-axis) are plotted against the -log(10) p-value (x-axis). Phenotypes shown are significant after FDR-correction. The direction of the bar indicates the direction of association, where a right-hand bar indicates a positive association, and a left-hand bar a negative association. Phenotypes are color coded by phenotype category.

### Independent Component 15

**Figure 41:**
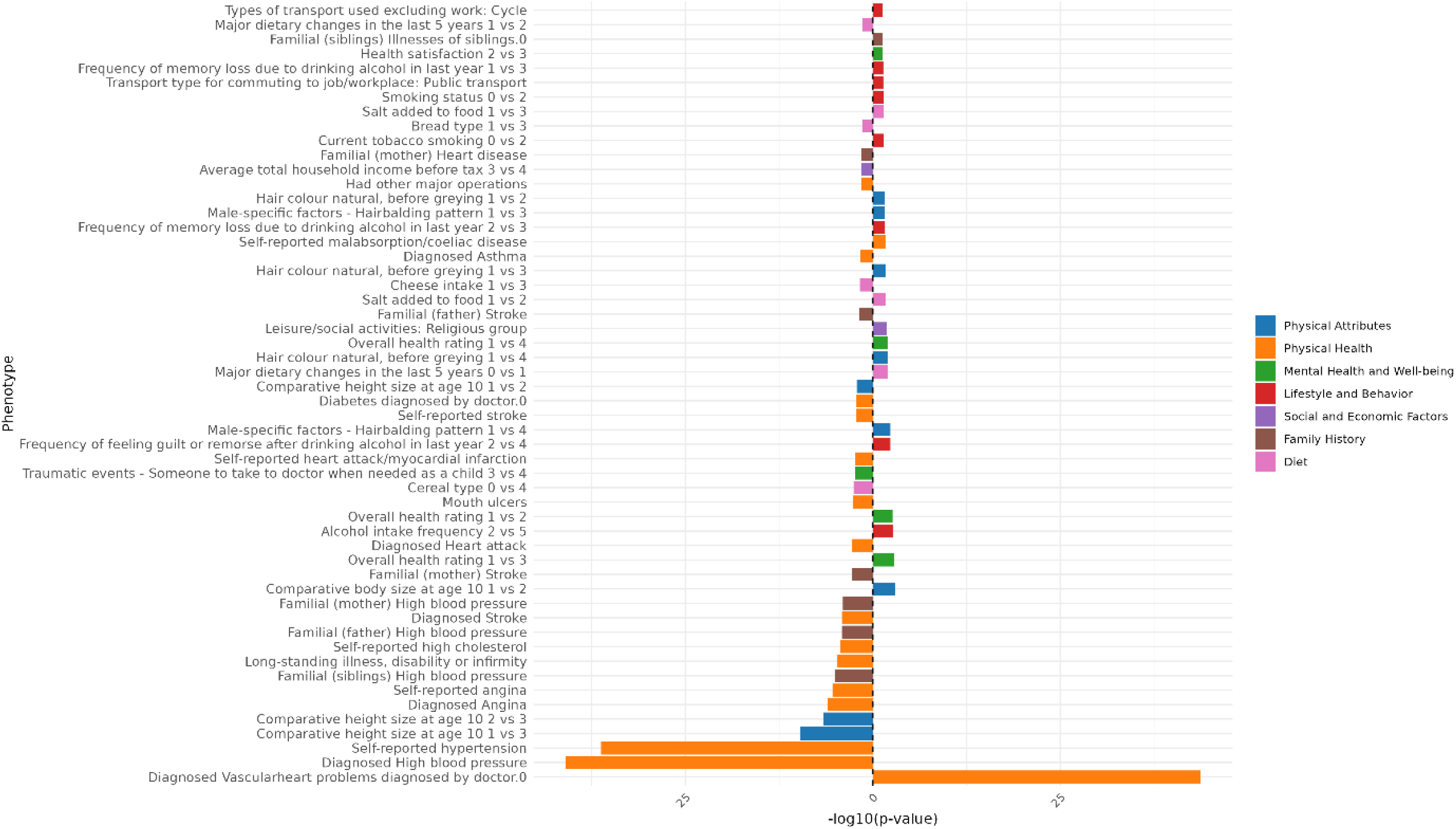
Genomic Independent Component 15 PCS association strength across binary/binarized non-neuroimaging phenotypes. Phenotypes (y-axis) are plotted against the -log(10) p-value (x-axis). Phenotypes shown are significant after FDR-correction. The direction of the bar indicates the direction of association, where a right-hand bar indicates a positive association, and a left-hand bar a negative association. In contrast analyses (e.g., comparing response 1 vs. response 3), the direction of the bar indicates the direction of the association in relation to the answers contrasted. Specifically, in a 1 vs. 3 contrast, a bar pointing to the right indicates an association with response 3, while a bar pointing to the left indicates an association with response 1. The phenotype “Diagnosed Vascular problems diagnosed by doctor.0” equates to “No Vascular problems diagnosed”. Phenotypes are color coded by phenotype category.

**Figure 42:**
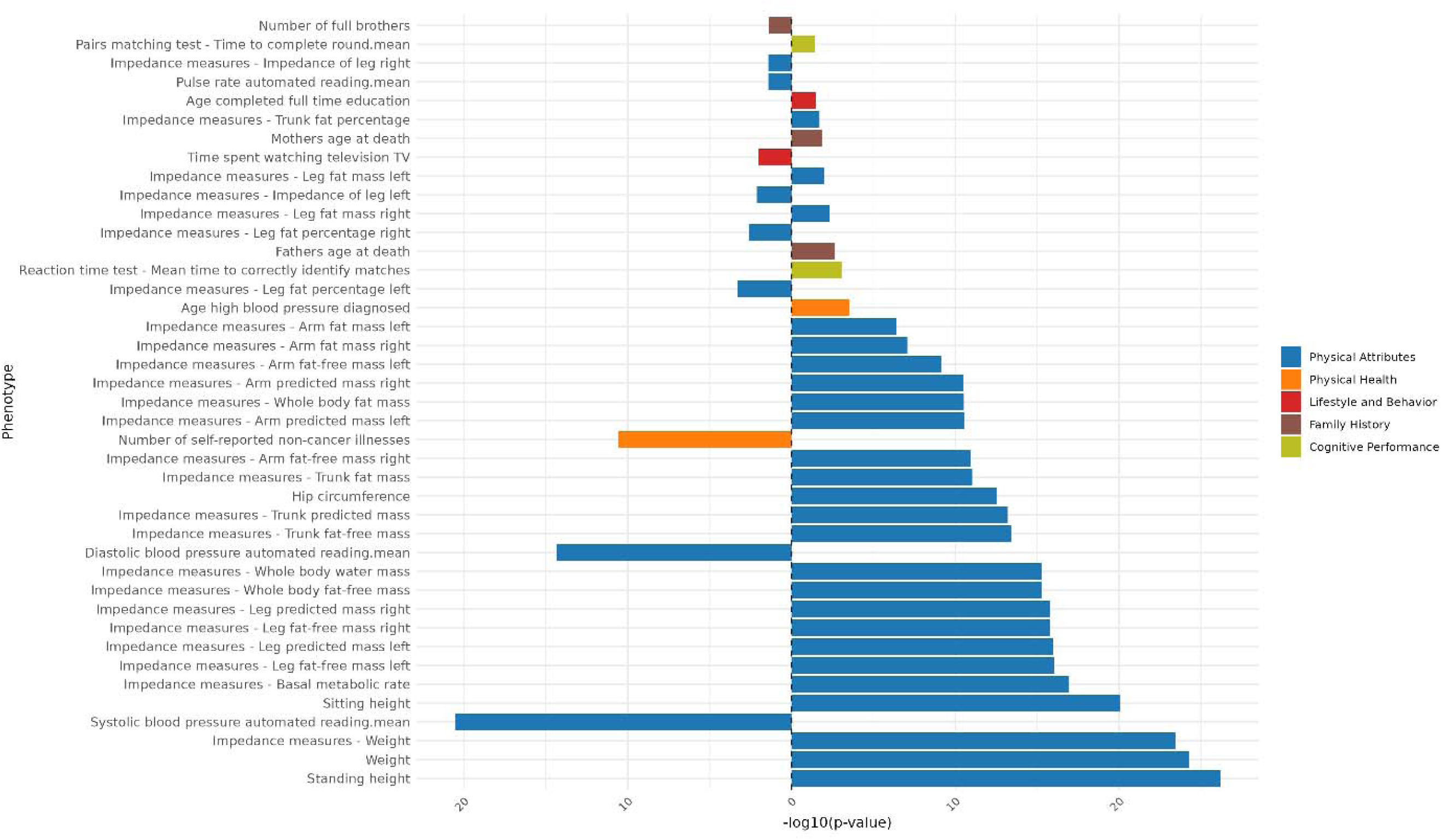
Genomic Independent Component 15 PCS association strength across continuous non-neuroimaging phenotypes. Phenotypes (y-axis) are plotted against the -log(10) p-value (x-axis). Phenotypes shown are significant after FDR-correction. The direction of the bar indicates the direction of association, where a right-hand bar indicates a positive association, and a left-hand bar a negative association. Phenotypes are color coded by phenotype category.

### Independent Component 16

**Figure 43:**
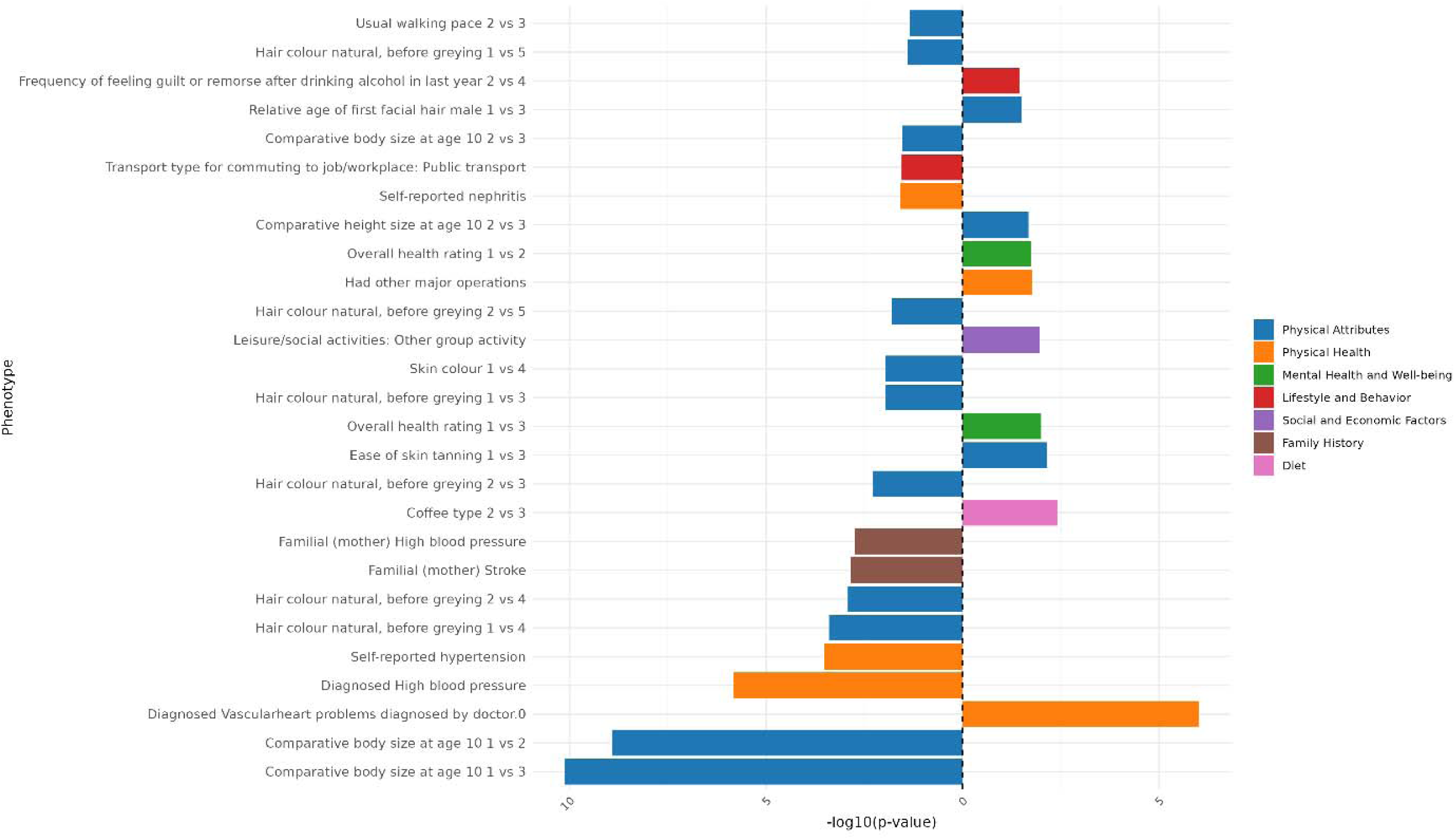
Genomic Independent Component 16 PCS association strength across binary/binarized non-neuroimaging phenotypes. Phenotypes (y-axis) are plotted against the -log(10) p-value (x-axis). Phenotypes shown are significant after FDR-correction. The direction of the bar indicates the direction of association, where a right-hand bar indicates a positive association, and a left-hand bar a negative association. In contrast analyses (e.g., comparing response 1 vs. response 3), the direction of the bar indicates the direction of the association in relation to the answers contrasted. Specifically, in a 1 vs. 3 contrast, a bar pointing to the right indicates an association with response 3, while a bar pointing to the left indicates an association with response 1. The phenotype “Diagnosed Vascular problems diagnosed by doctor.0” equates to “No Vascular problems diagnosed”. Phenotypes are color coded by phenotype category.

**Figure 44:**
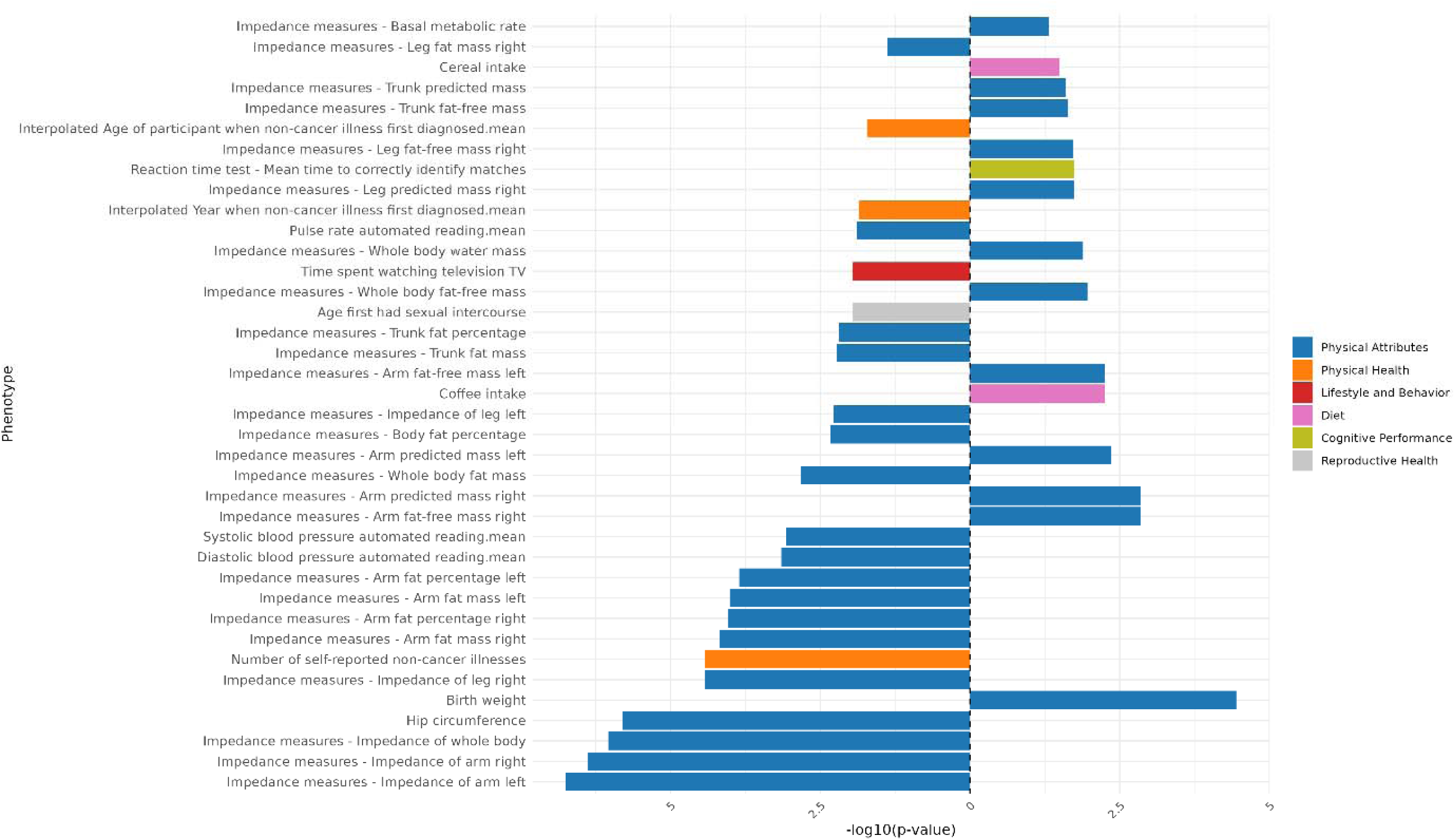
Genomic Independent Component 16 PCS association strength across continuous non-neuroimaging phenotypes. Phenotypes (y-axis) are plotted against the -log(10) p-value (x-axis). Phenotypes shown are significant after FDR-correction. The direction of the bar indicates the direction of association, where a right-hand bar indicates a positive association, and a left-hand bar a negative association. Phenotypes are color coded by phenotype category.

### Results of Genomic Principal Component Scores in Non-Neuroimaging space Principal Component 1

**Figure 45:**
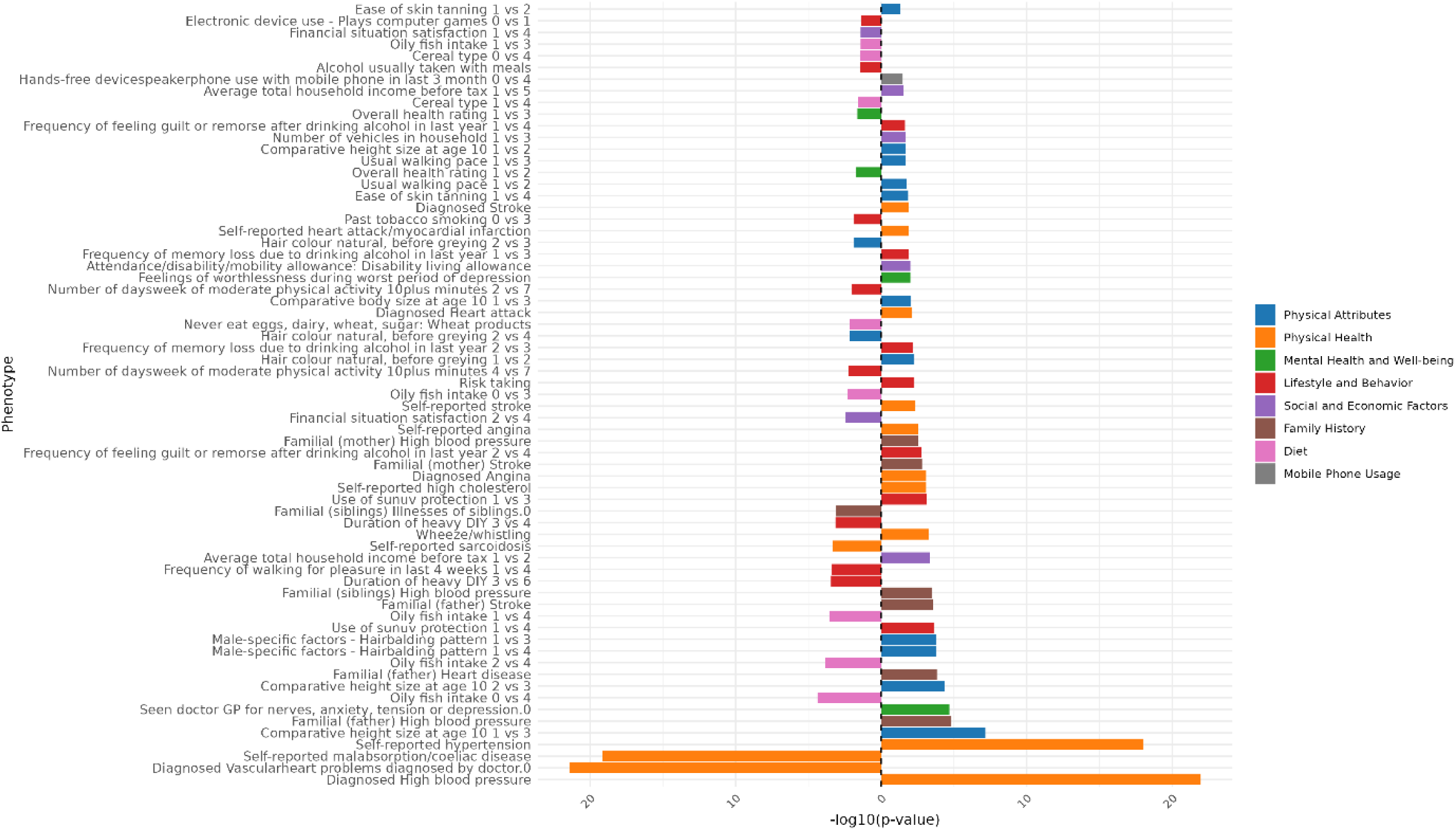
Genomic Principal Component 1 PCS association strength across binary/binarized non-neuroimaging phenotypes. Phenotypes (y-axis) are plotted against the -log(10) p-value (x-axis). Phenotypes shown are significant after FDR-correction. The direction of the bar indicates the direction of association, where a right-hand bar indicates a positive association, and a left-hand bar a negative association. In contrast analyses (e.g., comparing response 1 vs. response 3), the direction of the bar indicates the direction of the association in relation to the answers contrasted. Specifically, in a 1 vs. 3 contrast, a bar pointing to the right indicates an association with response 3, while a bar pointing to the left indicates an association with response 1. The phenotype “Diagnosed Vascular problems diagnosed by doctor.0” equates to “No Vascular problems diagnosed”. Phenotypes are color coded by phenotype category.

**Figure 46:**
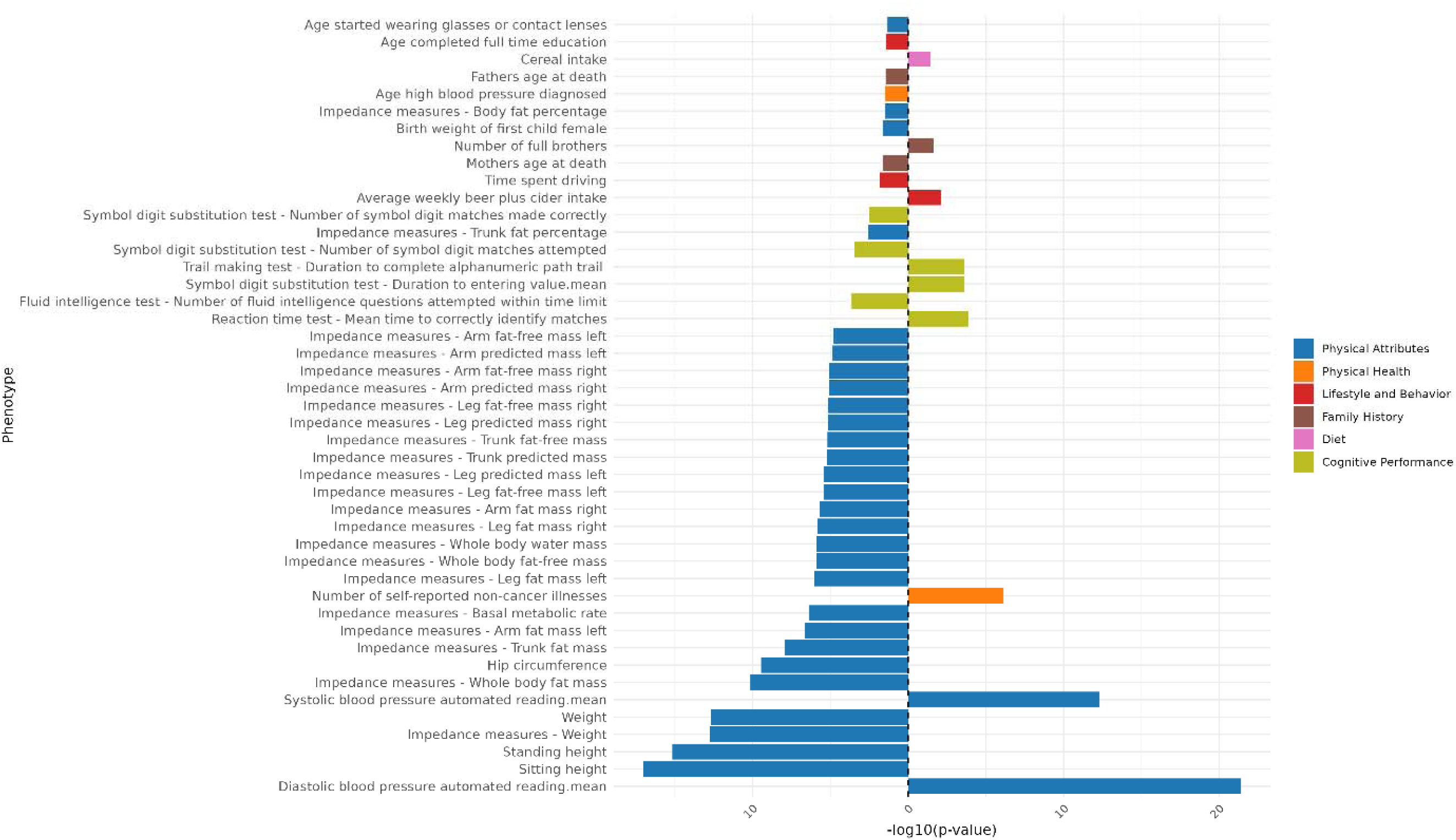
Genomic Principal Component 1 PCS association strength across continuous non-neuroimaging phenotypes. Phenotypes (y-axis) are plotted against the -log(10) p-value (x-axis). Phenotypes shown are significant after FDR-correction. The direction of the bar indicates the direction of association, where a right-hand bar indicates a positive association, and a left-hand bar a negative association. Phenotypes are color coded by phenotype category.

### Principal Component 2

**Figure 47:**
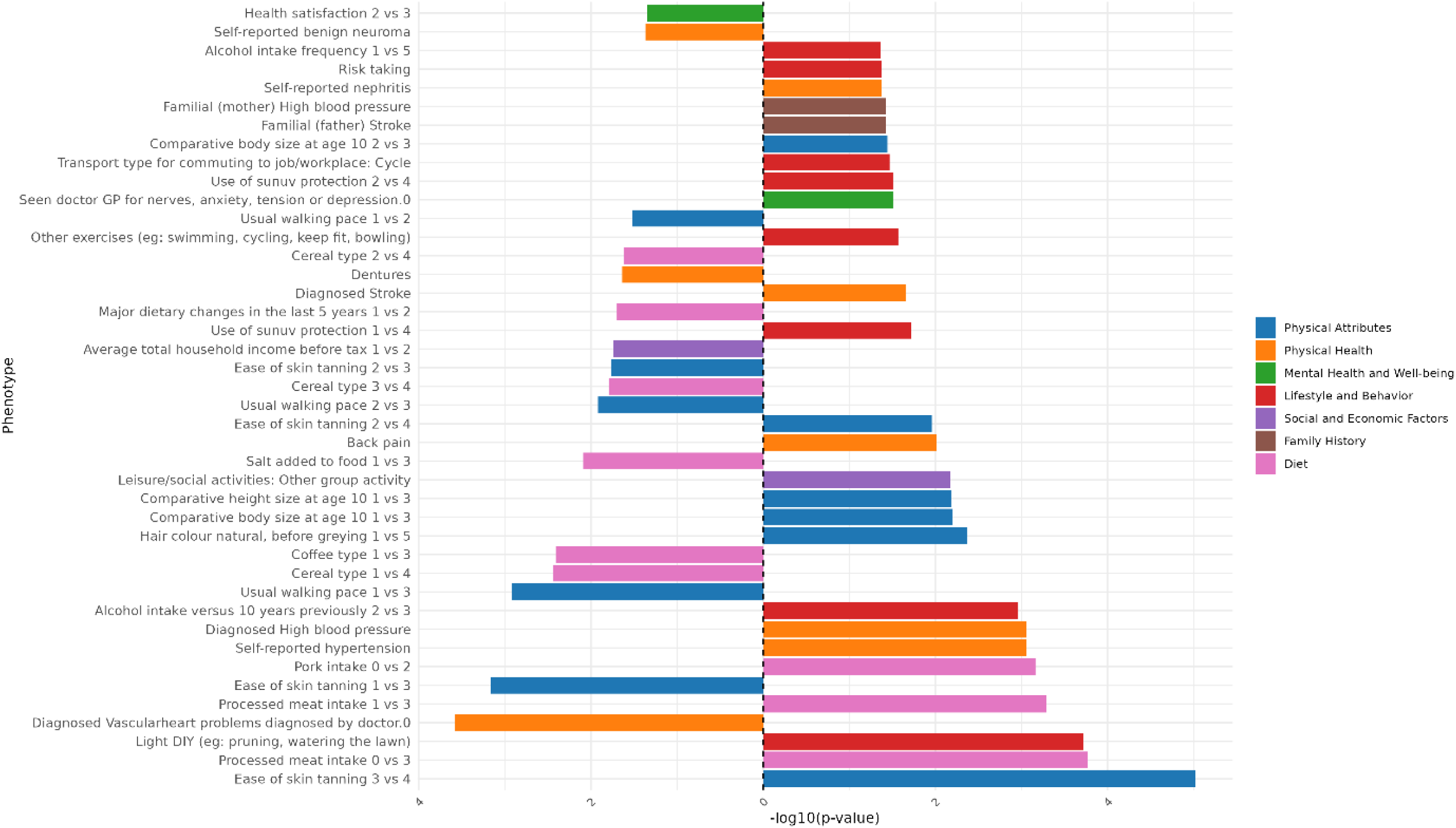
Genomic Principal Component 2 PCS association strength across binary/binarized non-neuroimaging phenotypes. Phenotypes (y-axis) are plotted against the -log(10) p-value (x-axis). Phenotypes shown are significant after FDR-correction. The direction of the bar indicates the direction of association, where a right-hand bar indicates a positive association, and a left-hand bar a negative association. In contrast analyses (e.g., comparing response 1 vs. response 3), the direction of the bar indicates the direction of the association in relation to the answers contrasted. Specifically, in a 1 vs. 3 contrast, a bar pointing to the right indicates an association with response 3, while a bar pointing to the left indicates an association with response 1. The phenotype “Diagnosed Vascular problems diagnosed by doctor.0” equates to “No Vascular problems diagnosed”. Phenotypes are color coded by phenotype category.

**Figure 48:**
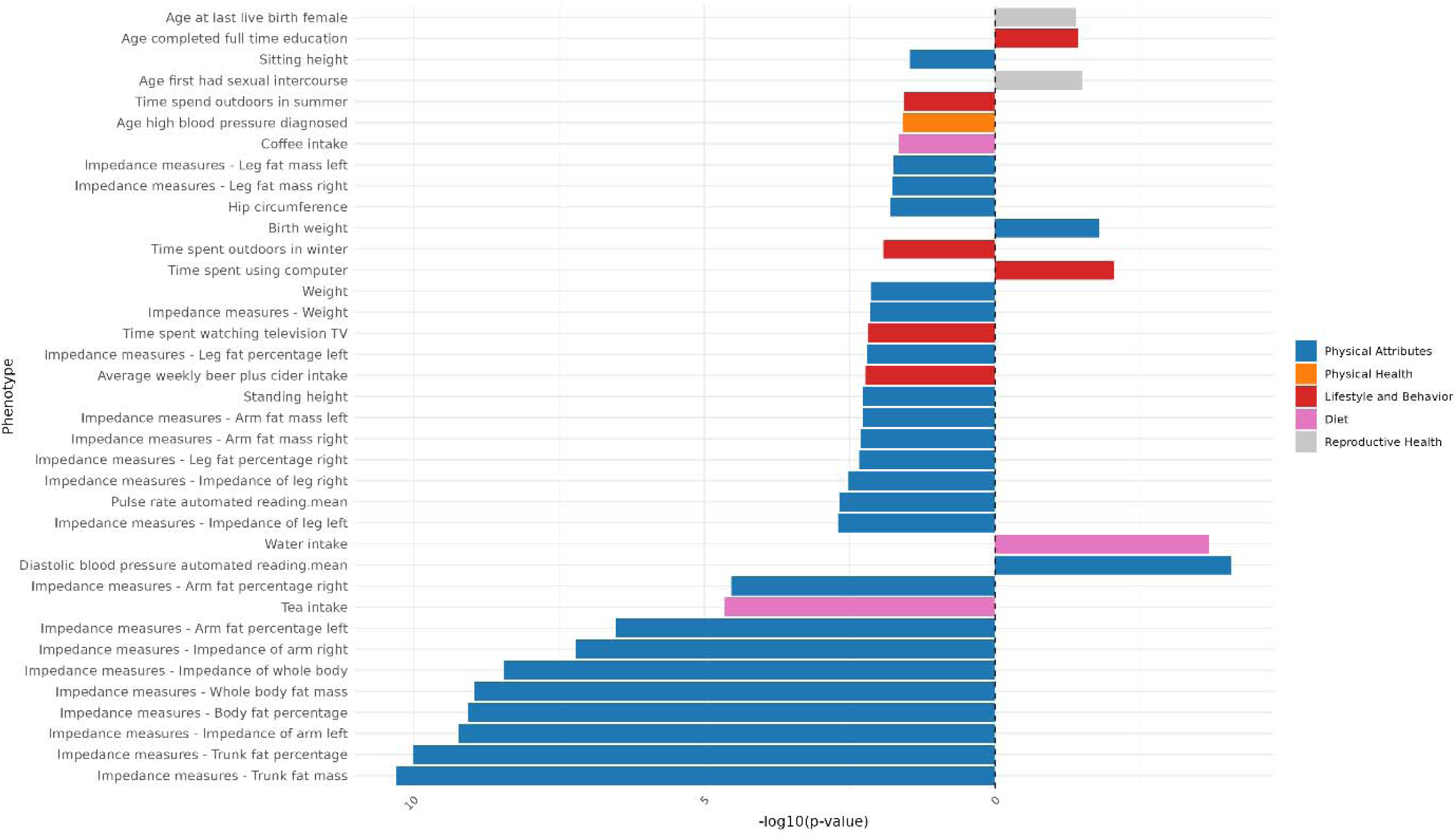
Genomic Principal Component 2 PCS association strength across continuous non-neuroimaging phenotypes. Phenotypes (y-axis) are plotted against the -log(10) p-value (x-axis). Phenotypes shown are significant after FDR-correction. The direction of the bar indicates the direction of association, where a right-hand bar indicates a positive association, and a left-hand bar a negative association. Phenotypes are color coded by phenotype category.

### Principal Component 3

**Figure 49:**
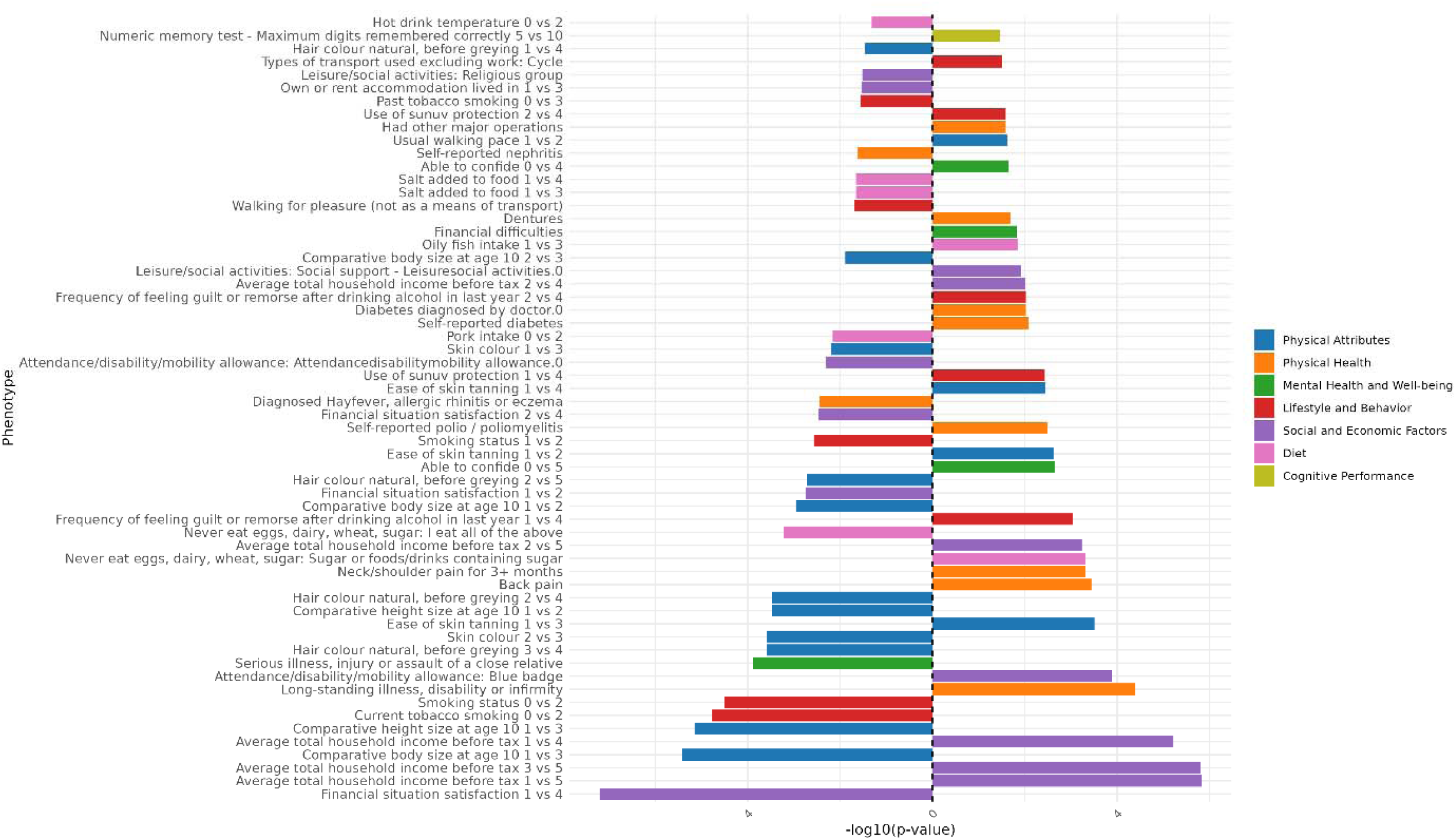
Genomic Principal Component 3 PCS association strength across binary/binarized non-neuroimaging phenotypes. Phenotypes (y-axis) are plotted against the -log(10) p-value (x-axis). Phenotypes shown are significant after FDR-correction. The direction of the bar indicates the direction of association, where a right-hand bar indicates a positive association, and a left-hand bar a negative association. In contrast analyses (e.g., comparing response 1 vs. response 3), the direction of the bar indicates the direction of the association in relation to the answers contrasted. Specifically, in a 1 vs. 3 contrast, a bar pointing to the right indicates an association with response 3, while a bar pointing to the left indicates an association with response 1. The phenotype “Diagnosed Vascular problems diagnosed by doctor.0” equates to “No Vascular problems diagnosed”. Phenotypes are color coded by phenotype category.

**Figure 50:**
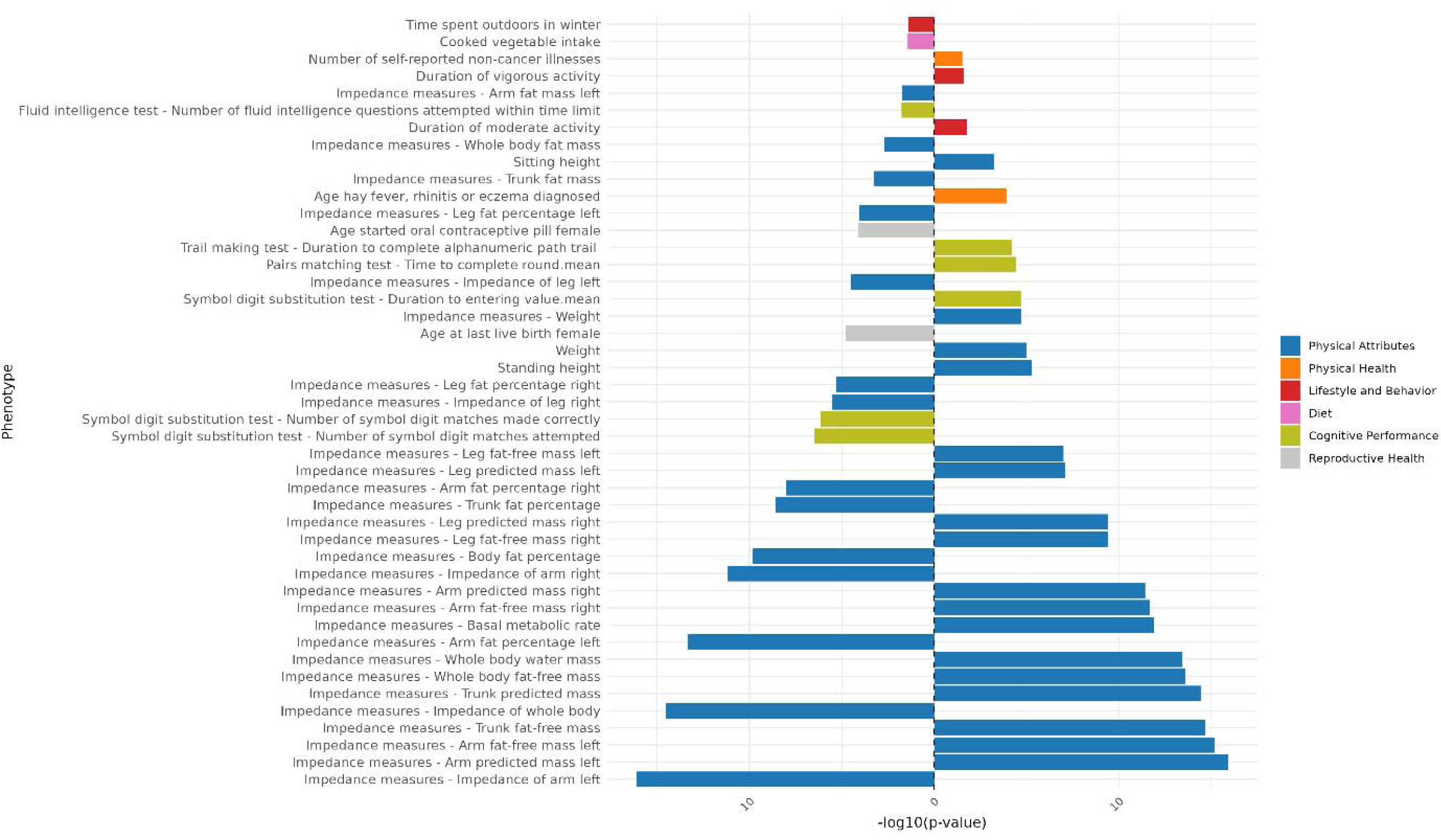
Genomic Principal Component 3 PCS association strength across continuous non-neuroimaging phenotypes. Phenotypes (y-axis) are plotted against the -log(10) p-value (x-axis). Phenotypes shown are significant after FDR-correction. The direction of the bar indicates the direction of association, where a right-hand bar indicates a positive association, and a left-hand bar a negative association. Phenotypes are color coded by phenotype category.

### Principal Component 4

**Figure 51:**
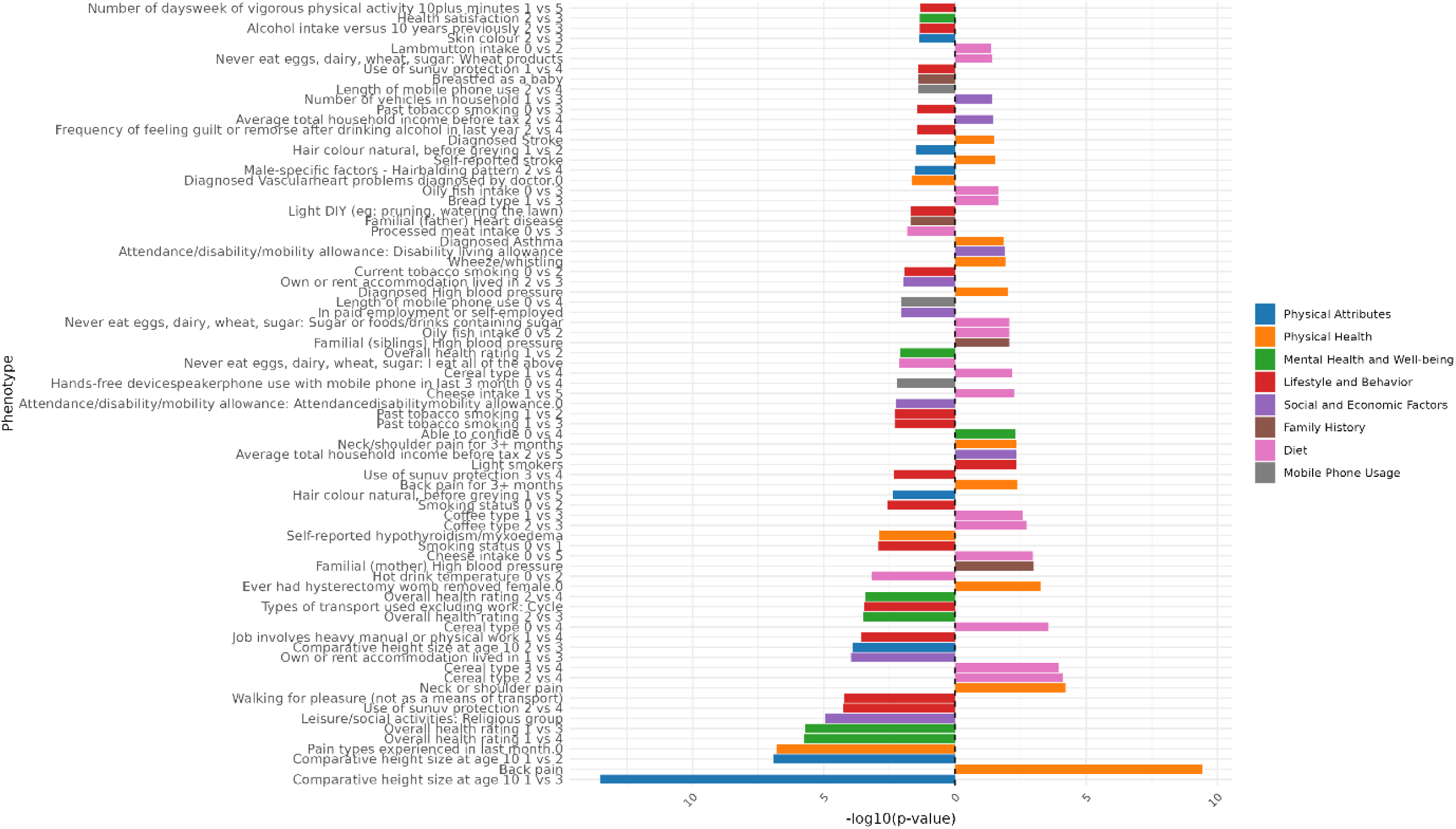
Genomic Principal Component 4 PCS association strength across binary/binarized non-neuroimaging phenotypes. Phenotypes (y-axis) are plotted against the -log(10) p-value (x-axis). Phenotypes shown are significant after FDR-correction. The direction of the bar indicates the direction of association, where a right-hand bar indicates a positive association, and a left-hand bar a negative association. In contrast analyses (e.g., comparing response 1 vs. response 3), the direction of the bar indicates the direction of the association in relation to the answers contrasted. Specifically, in a 1 vs. 3 contrast, a bar pointing to the right indicates an association with response 3, while a bar pointing to the left indicates an association with response 1. The phenotype “Diagnosed Vascular problems diagnosed by doctor.0” equates to “No Vascular problems diagnosed”. Phenotypes are color coded by phenotype category.

**Figure 52:**
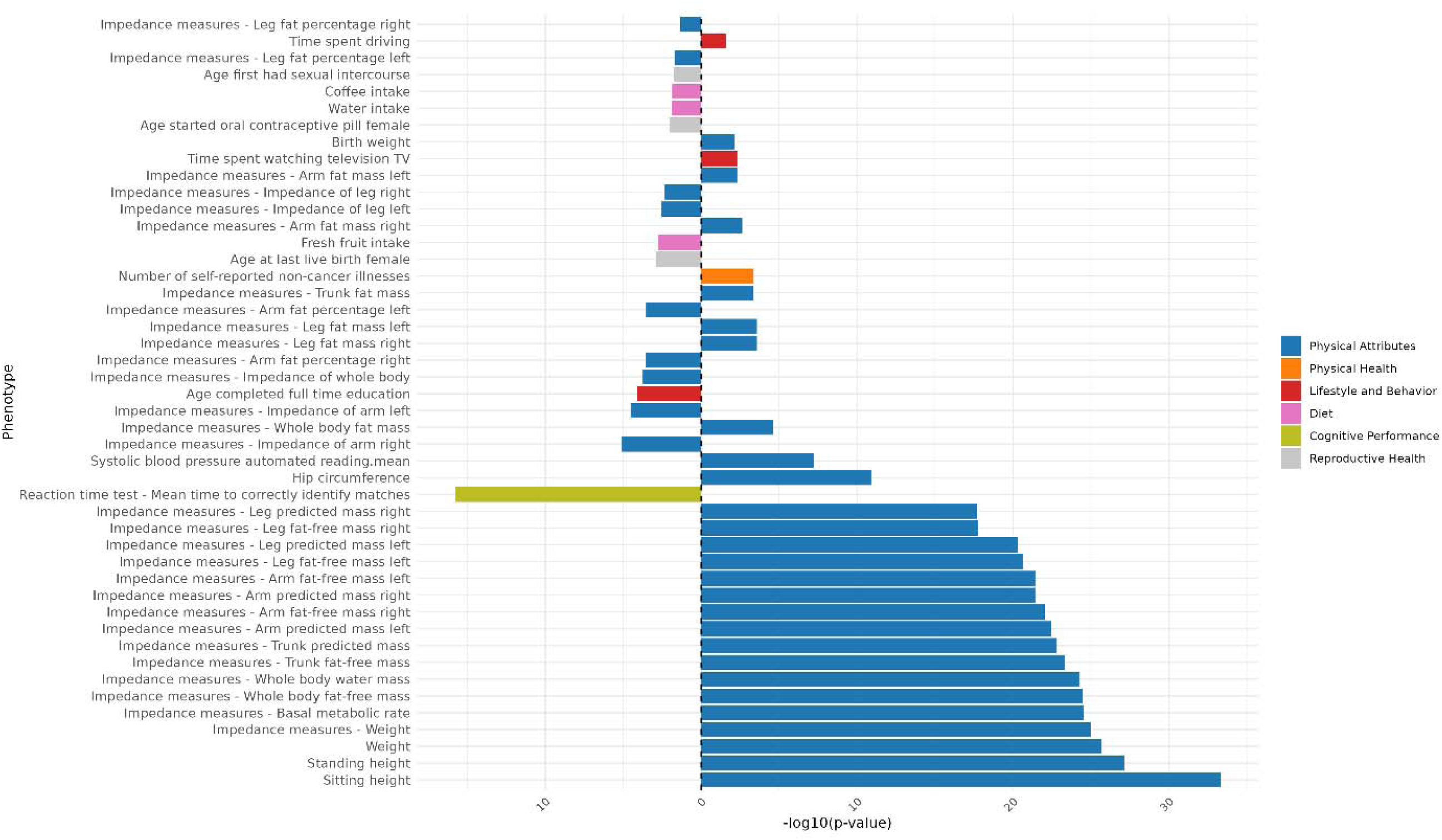
Genomic Principal Component 4 PCS association strength across continuous non-neuroimaging phenotypes. Phenotypes (y-axis) are plotted against the -log(10) p-value (x-axis). Phenotypes shown are significant after FDR-correction. The direction of the bar indicates the direction of association, where a right-hand bar indicates a positive association, and a left-hand bar a negative association. Phenotypes are color coded by phenotype category.

### Principal Component 5

**Figure 53:**
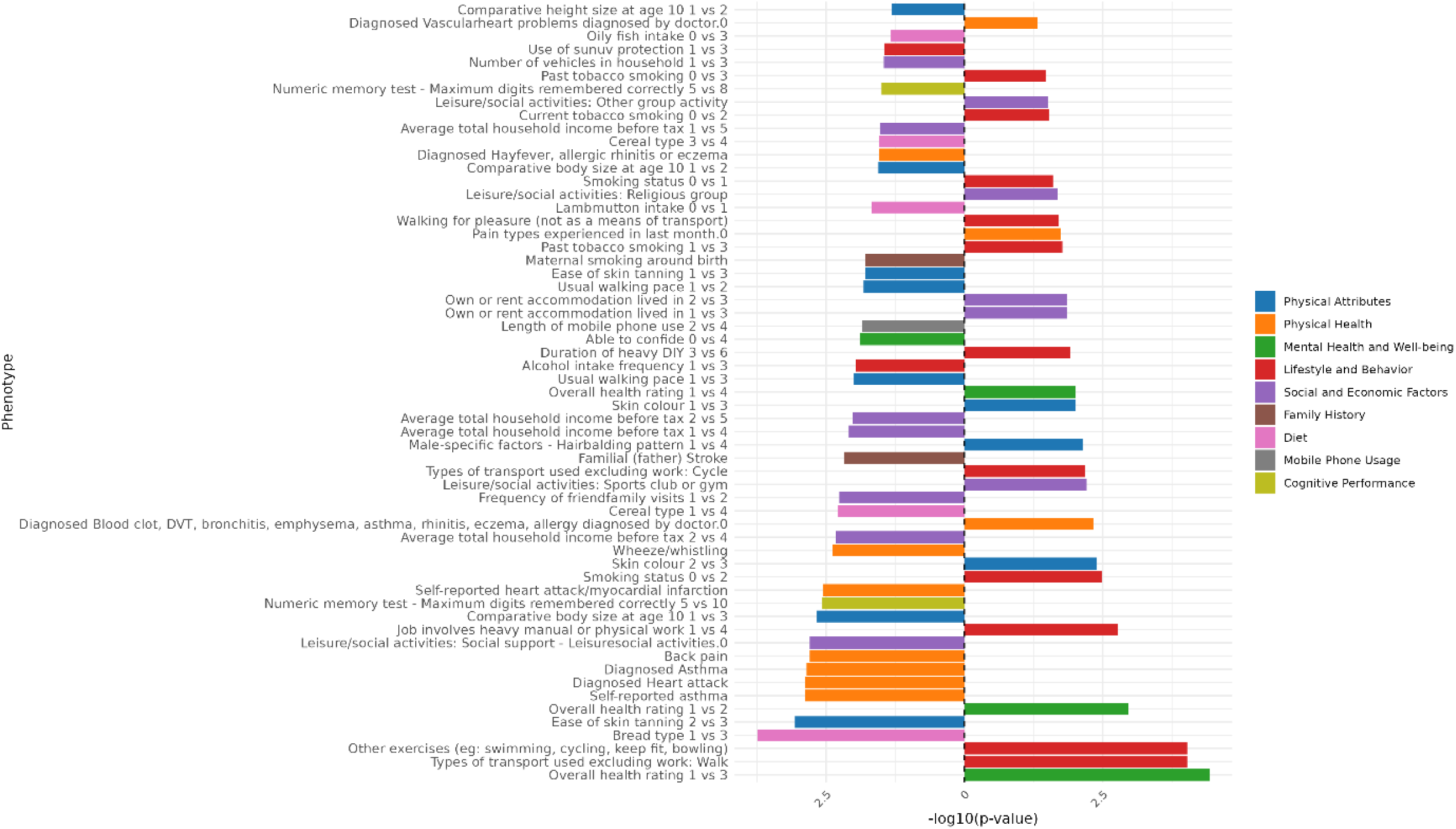
Genomic Principal Component 5 PCS association strength across binary/binarized non-neuroimaging phenotypes. Phenotypes (y-axis) are plotted against the -log(10) p-value (x-axis). Phenotypes shown are significant after FDR-correction. The direction of the bar indicates the direction of association, where a right-hand bar indicates a positive association, and a left-hand bar a negative association. In contrast analyses (e.g., comparing response 1 vs. response 3), the direction of the bar indicates the direction of the association in relation to the answers contrasted. Specifically, in a 1 vs. 3 contrast, a bar pointing to the right indicates an association with response 3, while a bar pointing to the left indicates an association with response 1. The phenotype “Diagnosed Vascular problems diagnosed by doctor.0” equates to “No Vascular problems diagnosed”. Phenotypes are color coded by phenotype category.

**Figure 54:**
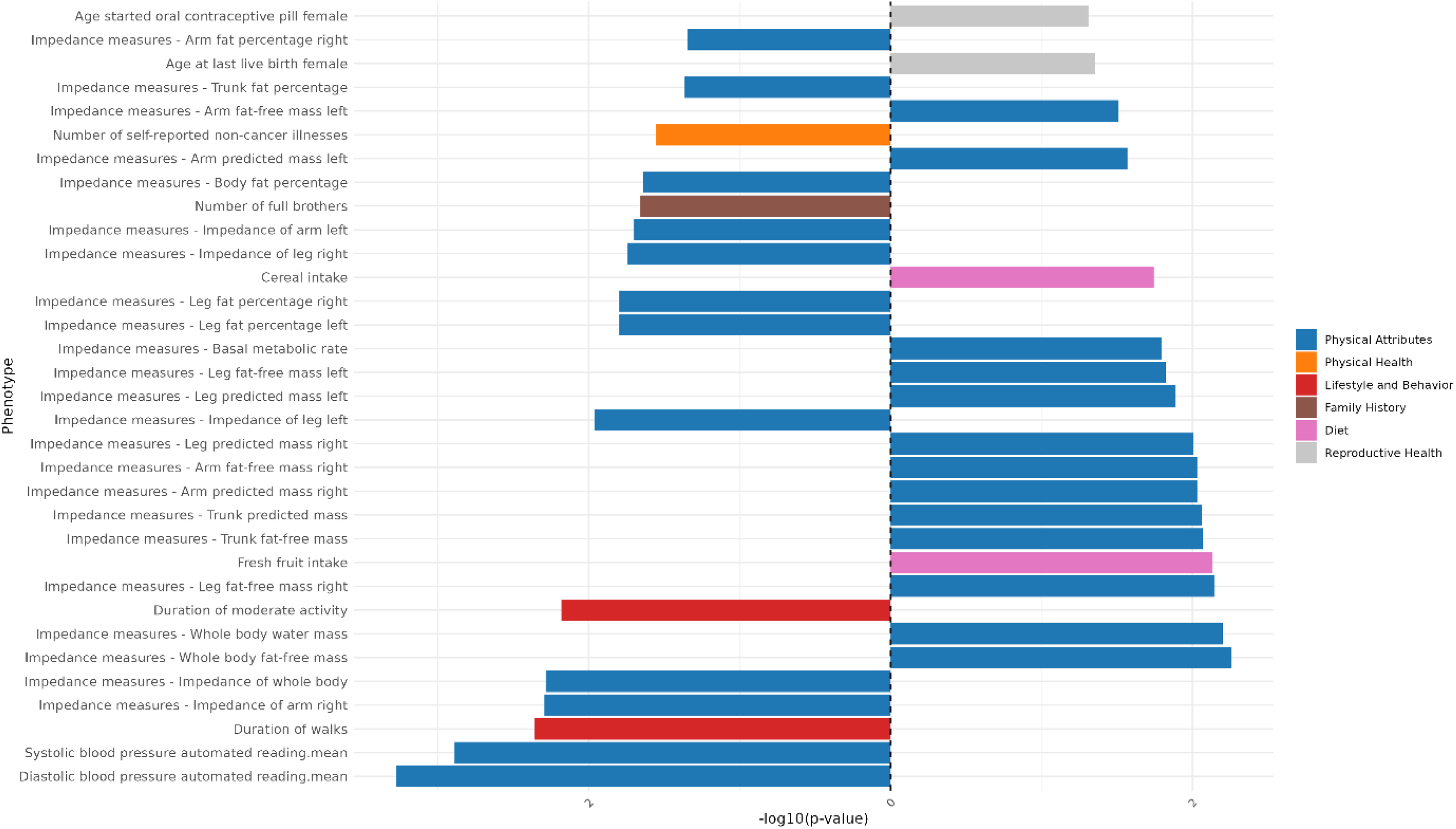
Genomic Principal Component 5 PCS association strength across continuous non-neuroimaging phenotypes. Phenotypes (y-axis) are plotted against the -log(10) p-value (x-axis). Phenotypes shown are significant after FDR-correction. The direction of the bar indicates the direction of association, where a right-hand bar indicates a positive association, and a left-hand bar a negative association. Phenotypes are color coded by phenotype category.

### Principal Component 6

**Figure 55:**
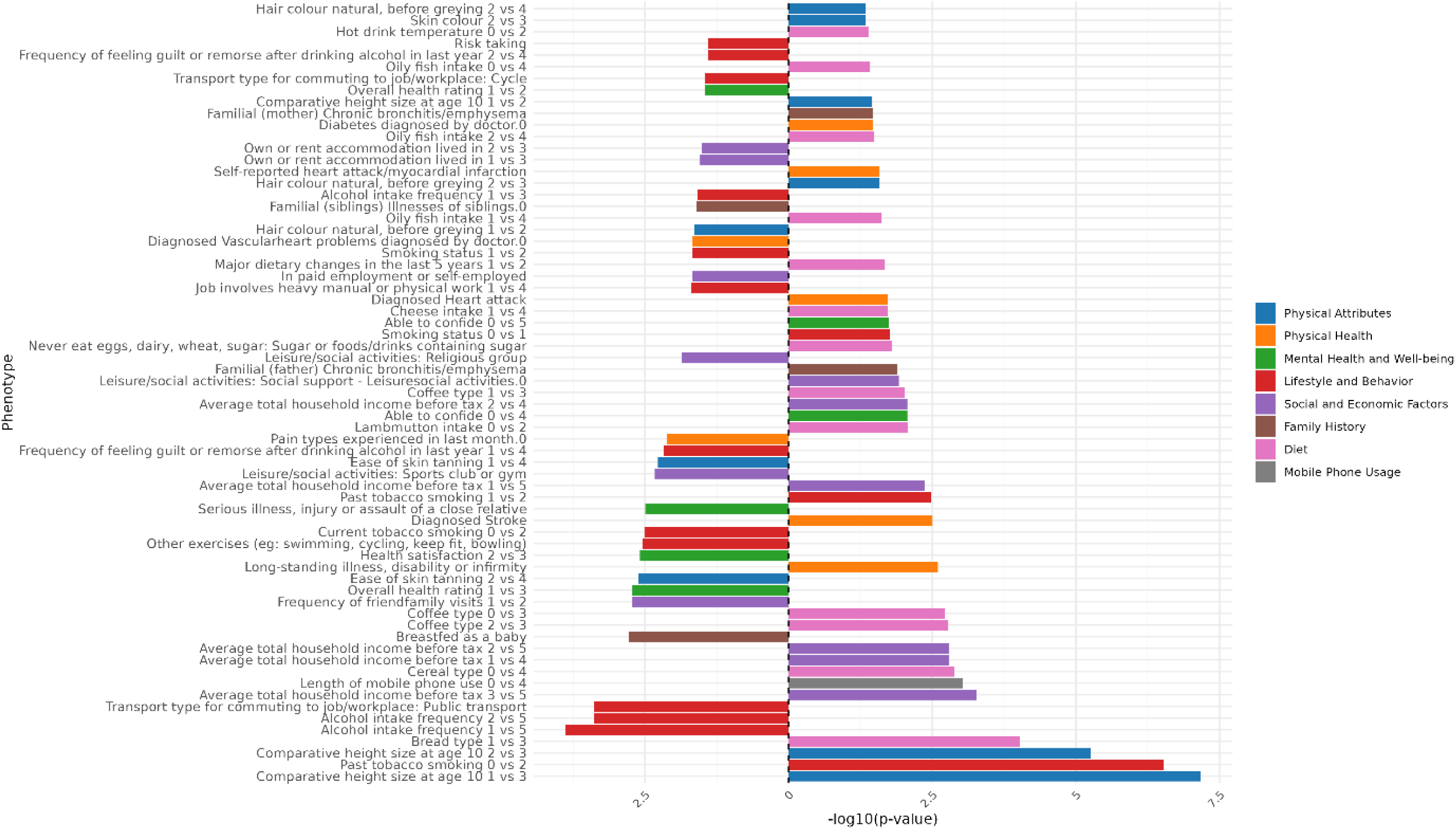
Genomic Principal Component 6 PCS association strength across binary/binarized non-neuroimaging phenotypes. Phenotypes (y-axis) are plotted against the -log(10) p-value (x-axis). Phenotypes shown are significant after FDR-correction. The direction of the bar indicates the direction of association, where a right-hand bar indicates a positive association, and a left-hand bar a negative association. In contrast analyses (e.g., comparing response 1 vs. response 3), the direction of the bar indicates the direction of the association in relation to the answers contrasted. Specifically, in a 1 vs. 3 contrast, a bar pointing to the right indicates an association with response 3, while a bar pointing to the left indicates an association with response 1. The phenotype “Diagnosed Vascular problems diagnosed by doctor.0” equates to “No Vascular problems diagnosed”. Phenotypes are color coded by phenotype category.

**Figure 56:**
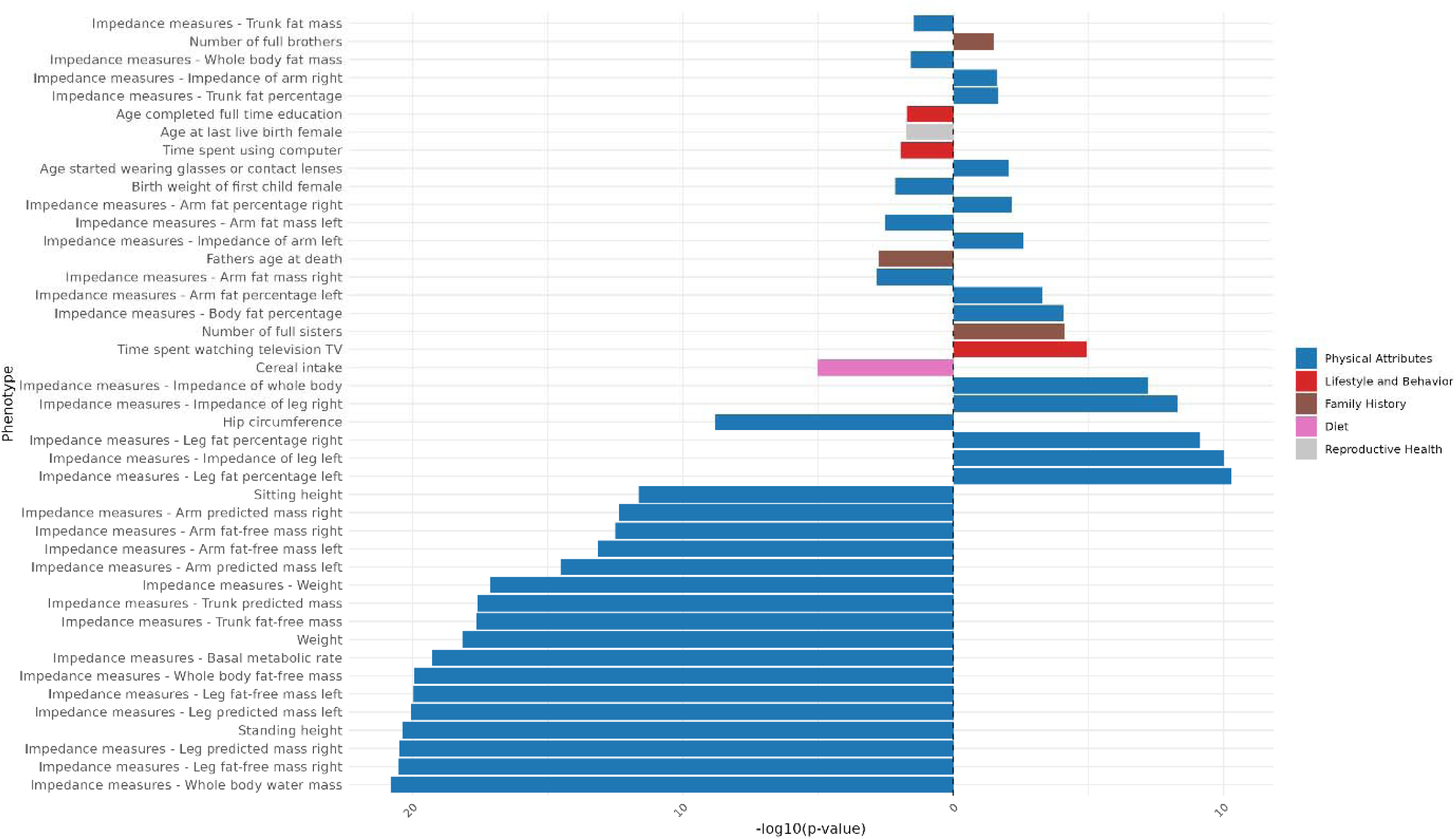
Genomic Principal Component 6 PCS association strength across continuous non-neuroimaging phenotypes. Phenotypes (y-axis) are plotted against the -log(10) p-value (x-axis). Phenotypes shown are significant after FDR-correction. The direction of the bar indicates the direction of association, where a right-hand bar indicates a positive association, and a left-hand bar a negative association. Phenotypes are color coded by phenotype category.

### Principal Component 7

**Figure 57:**
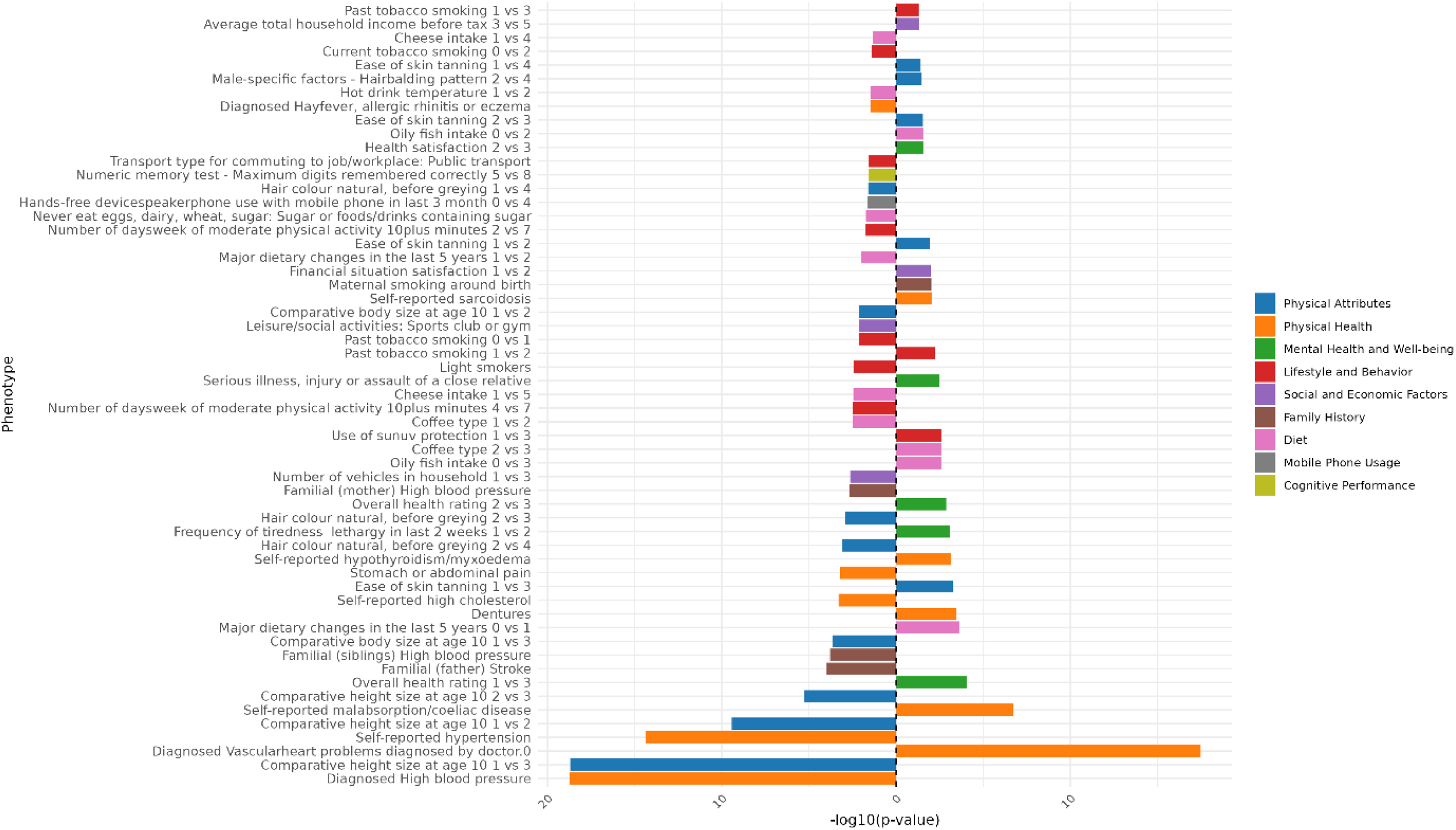
Genomic Principal Component 7 PCS association strength across binary/binarized non-neuroimaging phenotypes. Phenotypes (y-axis) are plotted against the -log(10) p-value (x-axis). Phenotypes shown are significant after FDR-correction. The direction of the bar indicates the direction of association, where a right-hand bar indicates a positive association, and a left-hand bar a negative association. In contrast analyses (e.g., comparing response 1 vs. response 3), the direction of the bar indicates the direction of the association in relation to the answers contrasted. Specifically, in a 1 vs. 3 contrast, a bar pointing to the right indicates an association with response 3, while a bar pointing to the left indicates an association with response 1. The phenotype “Diagnosed Vascular problems diagnosed by doctor.0” equates to “No Vascular problems diagnosed”. Phenotypes are color coded by phenotype category.

**Figure 58:**
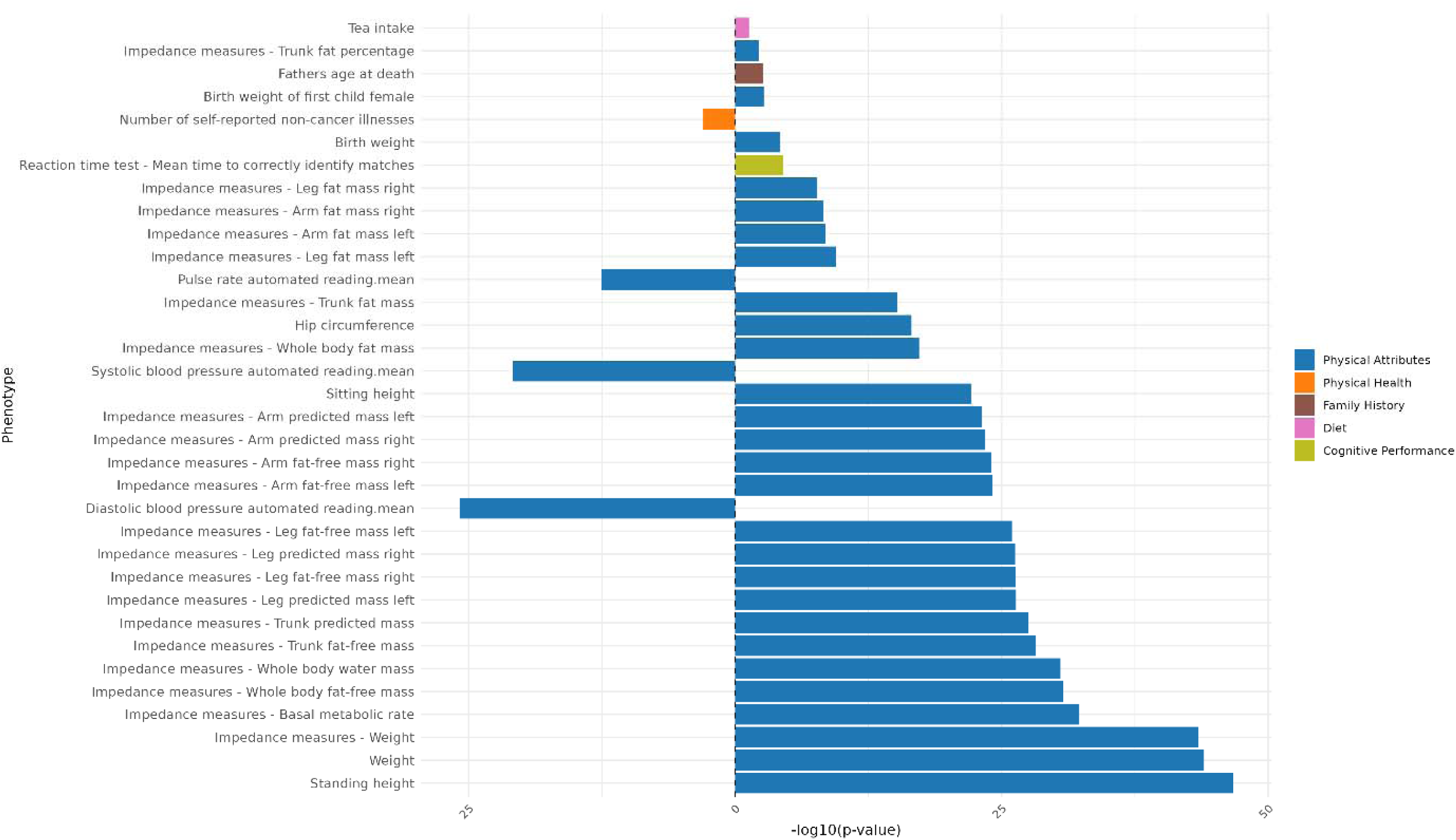
Genomic Principal Component 7 PCS association strength across continuous non-neuroimaging phenotypes. Phenotypes (y-axis) are plotted against the -log(10) p-value (x-axis). Phenotypes shown are significant after FDR-correction. The direction of the bar indicates the direction of association, where a right-hand bar indicates a positive association, and a left-hand bar a negative association. Phenotypes are color coded by phenotype category.

### Principal Component 8

**Figure 59:**
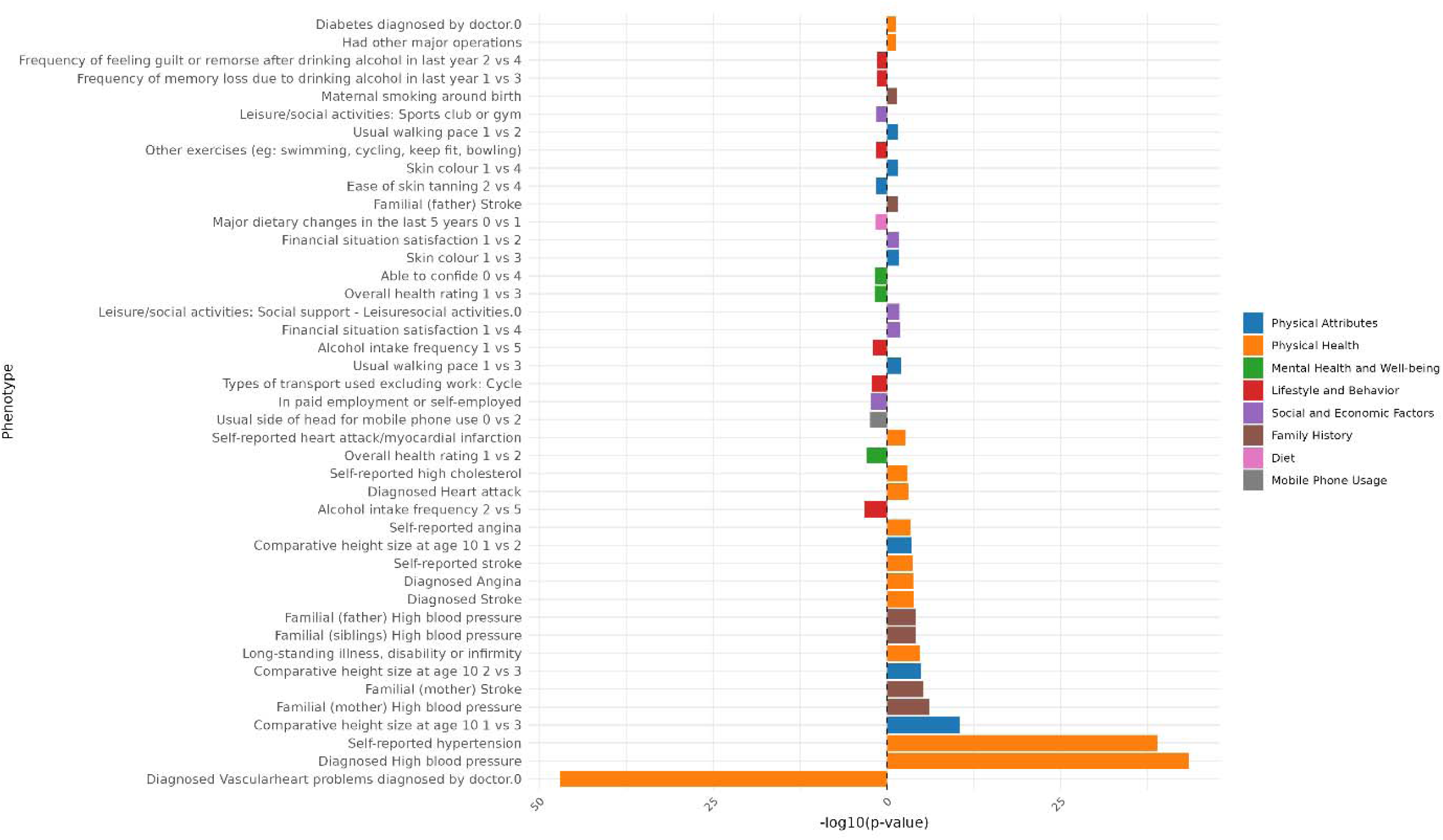
Genomic Principal Component 8 PCS association strength across binary/binarized non-neuroimaging phenotypes. Phenotypes (y-axis) are plotted against the -log(10) p-value (x-axis). Phenotypes shown are significant after FDR-correction. The direction of the bar indicates the direction of association, where a right-hand bar indicates a positive association, and a left-hand bar a negative association. In contrast analyses (e.g., comparing response 1 vs. response 3), the direction of the bar indicates the direction of the association in relation to the answers contrasted. Specifically, in a 1 vs. 3 contrast, a bar pointing to the right indicates an association with response 3, while a bar pointing to the left indicates an association with response 1. The phenotype “Diagnosed Vascular problems diagnosed by doctor.0” equates to “No Vascular problems diagnosed”. Phenotypes are color coded by phenotype category.

**Figure 60:**
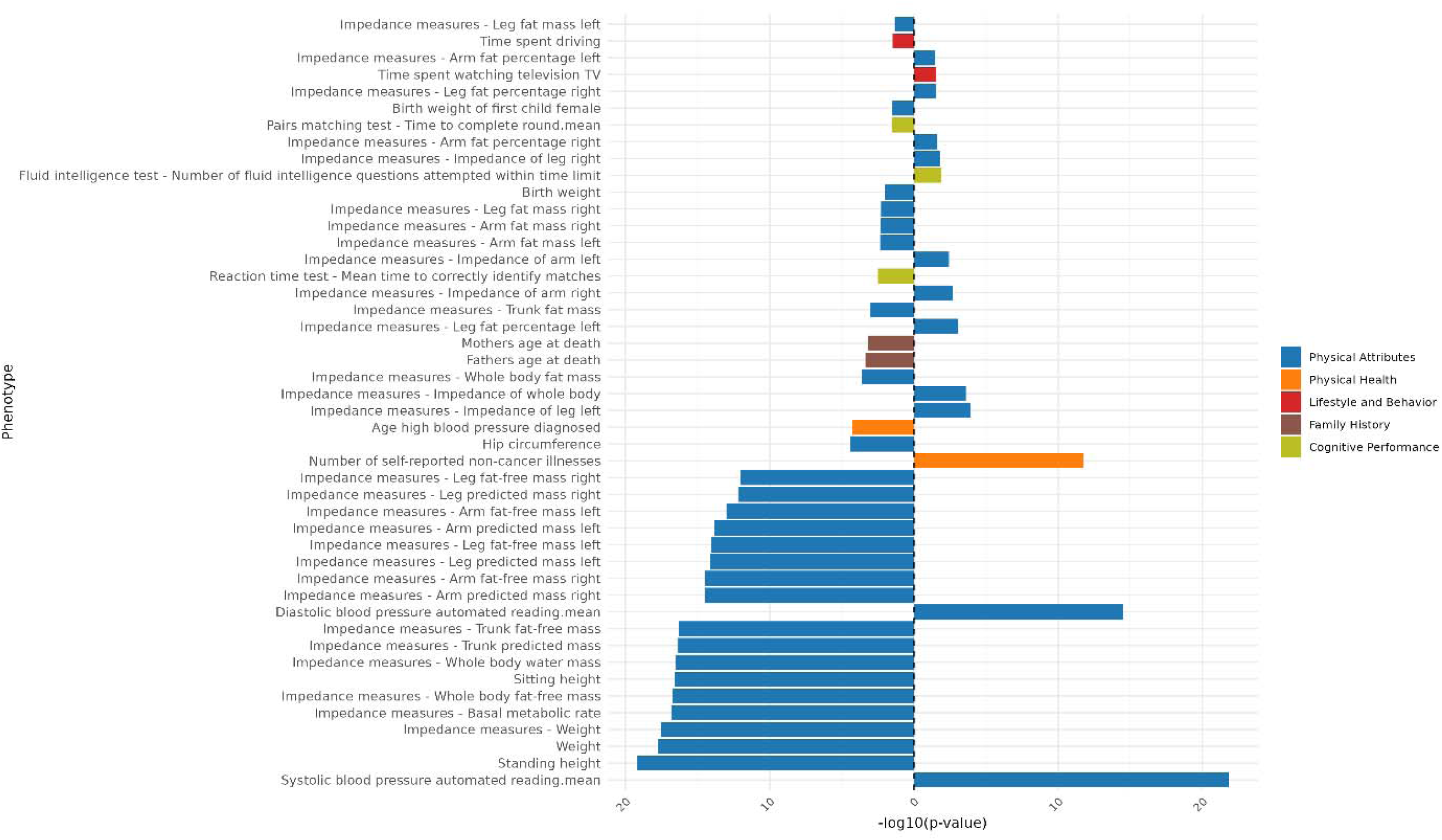
Genomic Principal Component 8 PCS association strength across continuous non-neuroimaging phenotypes. Phenotypes (y-axis) are plotted against the -log(10) p-value (x-axis). Phenotypes shown are significant after FDR-correction. The direction of the bar indicates the direction of association, where a right-hand bar indicates a positive association, and a left-hand bar a negative association. Phenotypes are color coded by phenotype category.

### Principal Component 9

**Figure 61:**
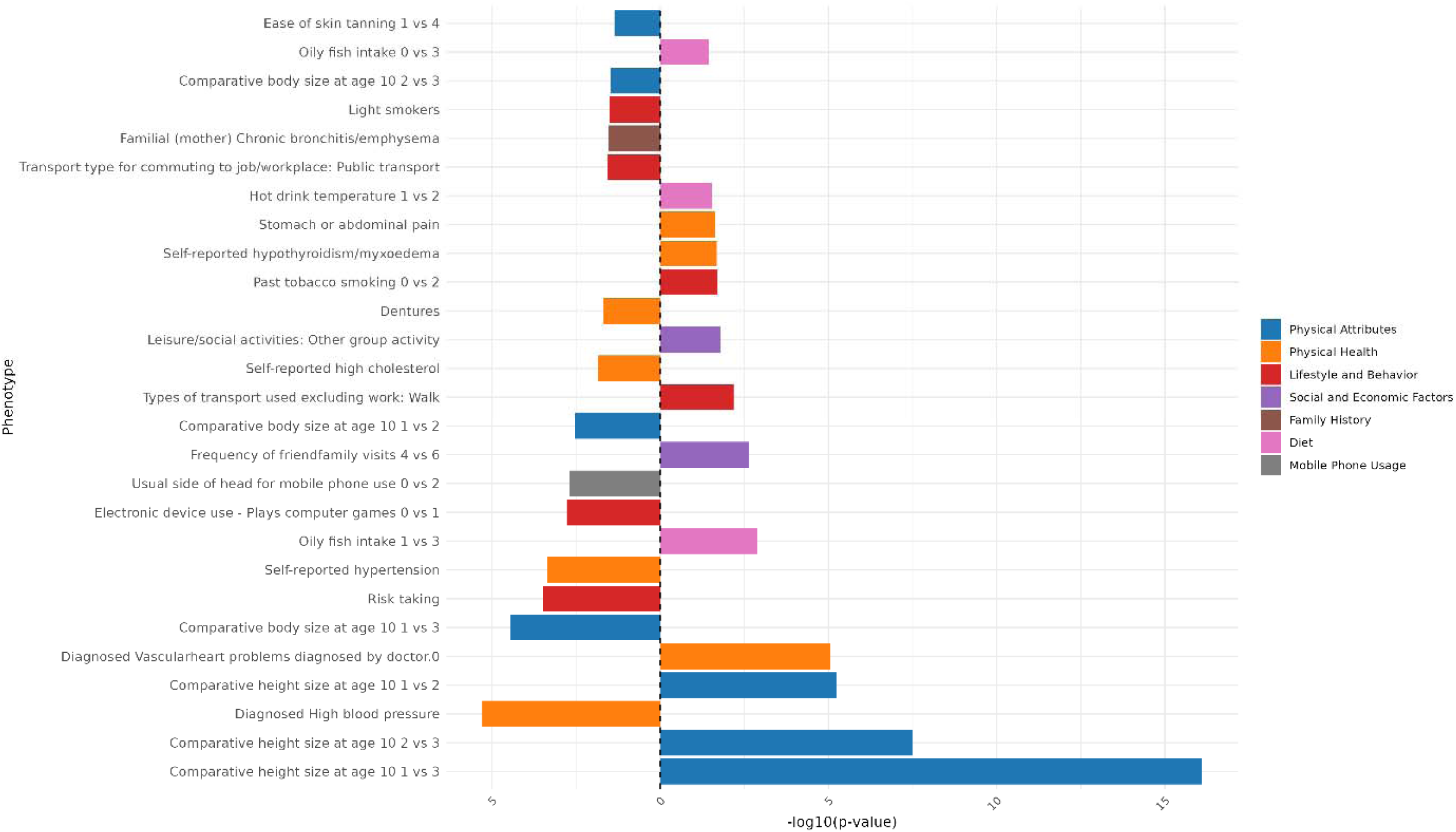
Genomic Principal Component 9 PCS association strength across binary/binarized non-neuroimaging phenotypes. Phenotypes (y-axis) are plotted against the -log(10) p-value (x-axis). Phenotypes shown are significant after FDR-correction. The direction of the bar indicates the direction of association, where a right-hand bar indicates a positive association, and a left-hand bar a negative association. In contrast analyses (e.g., comparing response 1 vs. response 3), the direction of the bar indicates the direction of the association in relation to the answers contrasted. Specifically, in a 1 vs. 3 contrast, a bar pointing to the right indicates an association with response 3, while a bar pointing to the left indicates an association with response 1. The phenotype “Diagnosed Vascular problems diagnosed by doctor.0” equates to “No Vascular problems diagnosed”. Phenotypes are color coded by phenotype category.

**Figure 62:**
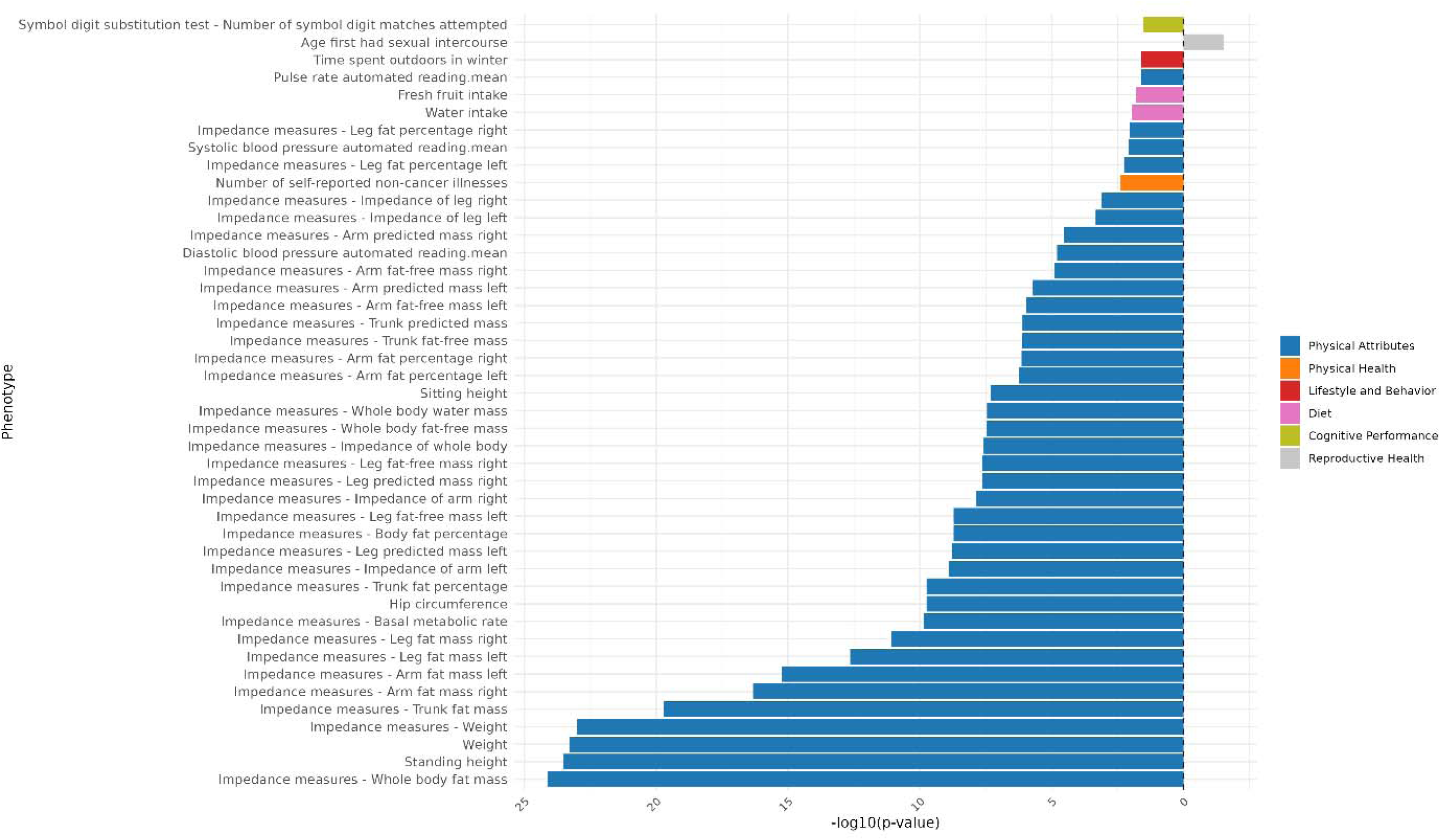
Genomic Principal Component 9 PCS association strength across continuous non-neuroimaging phenotypes. Phenotypes (y-axis) are plotted against the -log(10) p-value (x-axis). Phenotypes shown are significant after FDR-correction. The direction of the bar indicates the direction of association, where a right-hand bar indicates a positive association, and a left-hand bar a negative association. Phenotypes are color coded by phenotype category.

### Principal Component 10

**Figure 63:**
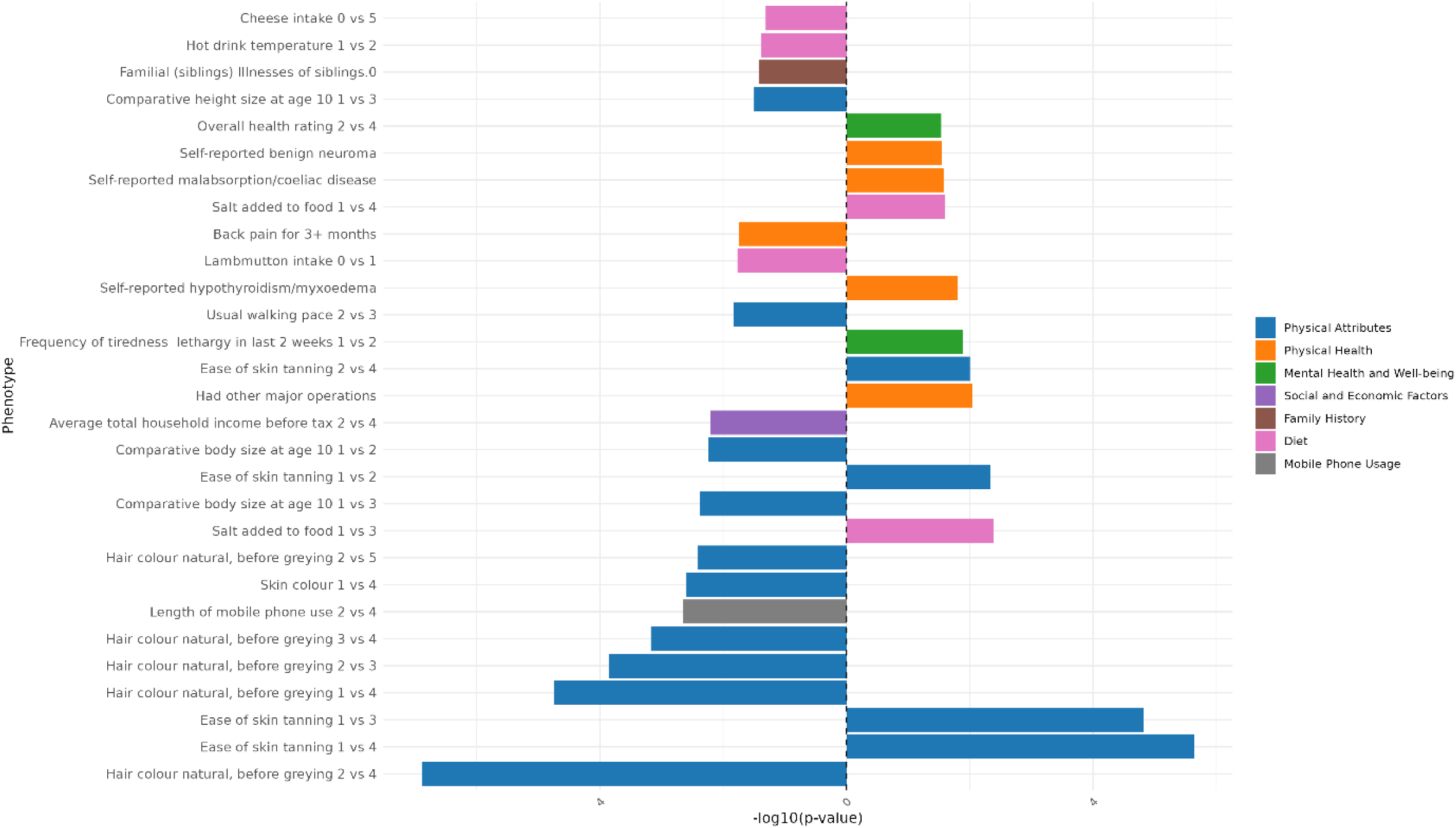
Genomic Principal Component 10 PCS association strength across binary/binarized non-neuroimaging phenotypes. Phenotypes (y-axis) are plotted against the -log(10) p-value (x-axis). Phenotypes shown are significant after FDR-correction. The direction of the bar indicates the direction of association, where a right-hand bar indicates a positive association, and a left-hand bar a negative association. In contrast analyses (e.g., comparing response 1 vs. response 3), the direction of the bar indicates the direction of the association in relation to the answers contrasted. Specifically, in a 1 vs. 3 contrast, a bar pointing to the right indicates an association with response 3, while a bar pointing to the left indicates an association with response 1. The phenotype “Diagnosed Vascular problems diagnosed by doctor.0” equates to “No Vascular problems diagnosed”. Phenotypes are color coded by phenotype category.

**Figure 64:**
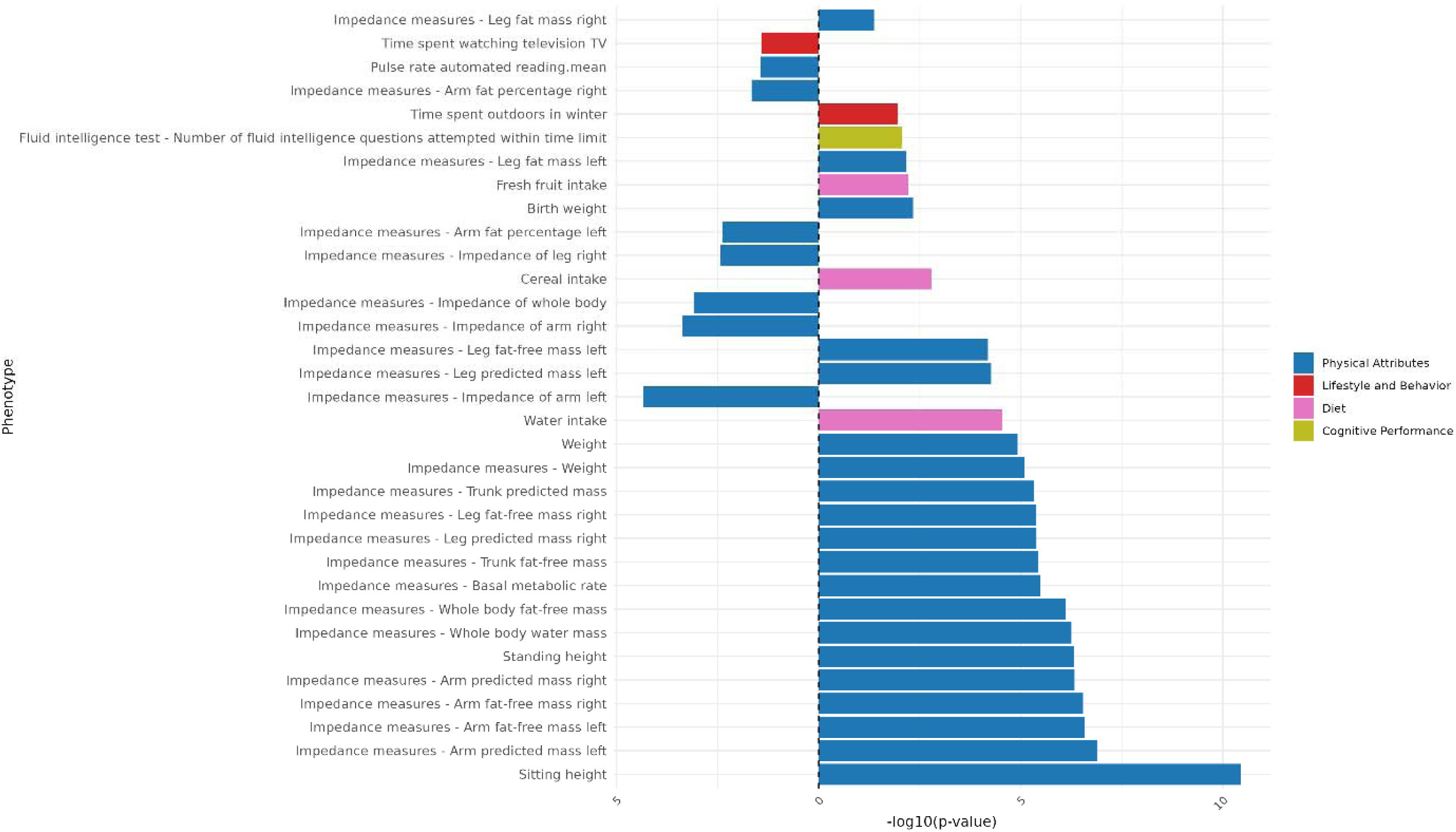
Genomic Principal Component 10 PCS association strength across continuous non-neuroimaging phenotypes. Phenotypes (y-axis) are plotted against the -log(10) p-value (x-axis). Phenotypes shown are significant after FDR-correction. The direction of the bar indicates the direction of association, where a right-hand bar indicates a positive association, and a left-hand bar a negative association. Phenotypes are color coded by phenotype category.

### Principal Component 11

**Figure 65:**
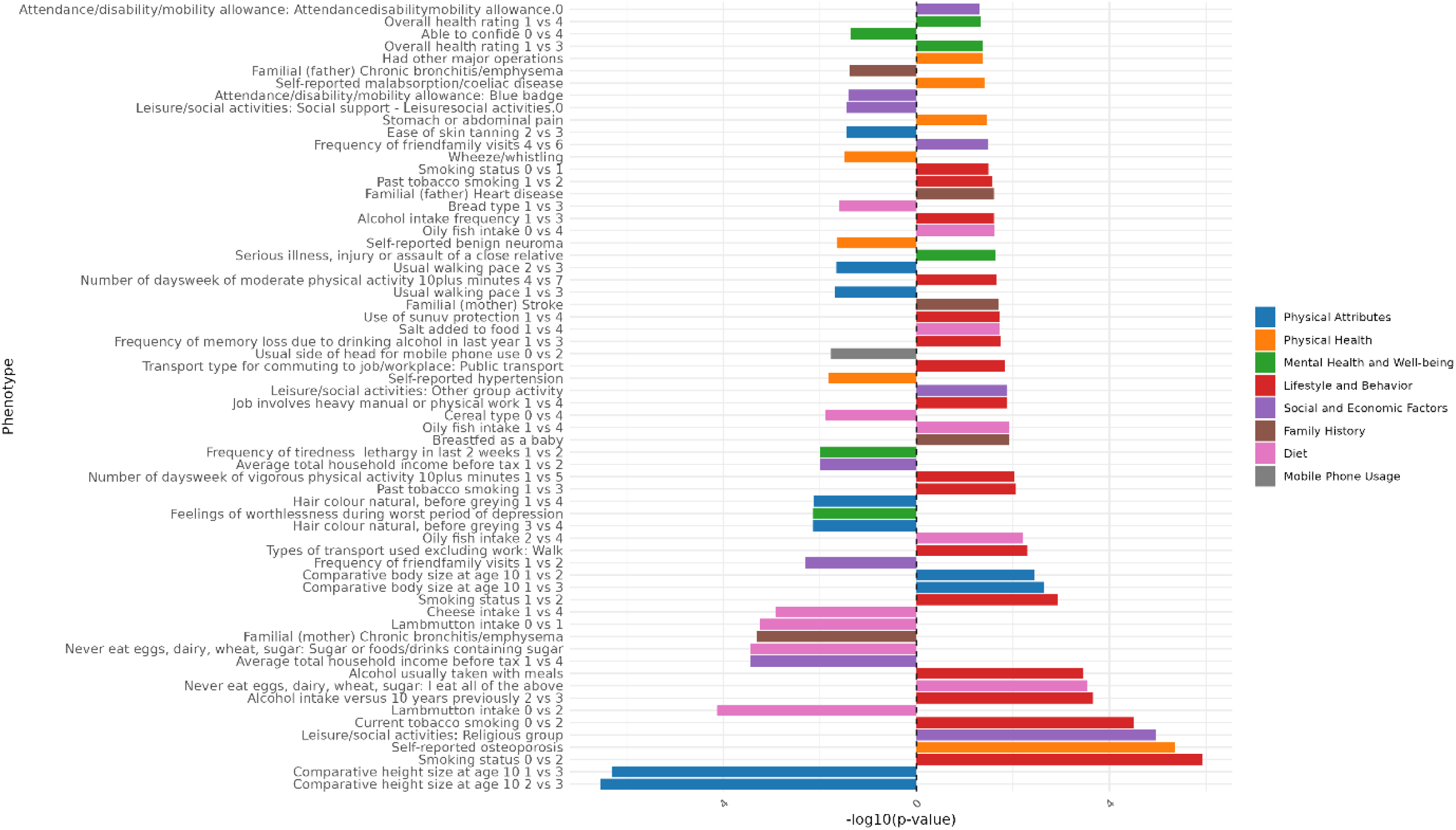
Genomic Principal Component 11 PCS association strength across binary/binarized non-neuroimaging phenotypes. Phenotypes (y-axis) are plotted against the -log(10) p-value (x-axis). Phenotypes shown are significant after FDR-correction. The direction of the bar indicates the direction of association, where a right-hand bar indicates a positive association, and a left-hand bar a negative association. In contrast analyses (e.g., comparing response 1 vs. response 3), the direction of the bar indicates the direction of the association in relation to the answers contrasted. Specifically, in a 1 vs. 3 contrast, a bar pointing to the right indicates an association with response 3, while a bar pointing to the left indicates an association with response 1. The phenotype “Diagnosed Vascular problems diagnosed by doctor.0” equates to “No Vascular problems diagnosed”. Phenotypes are color coded by phenotype category.

**Figure 66:**
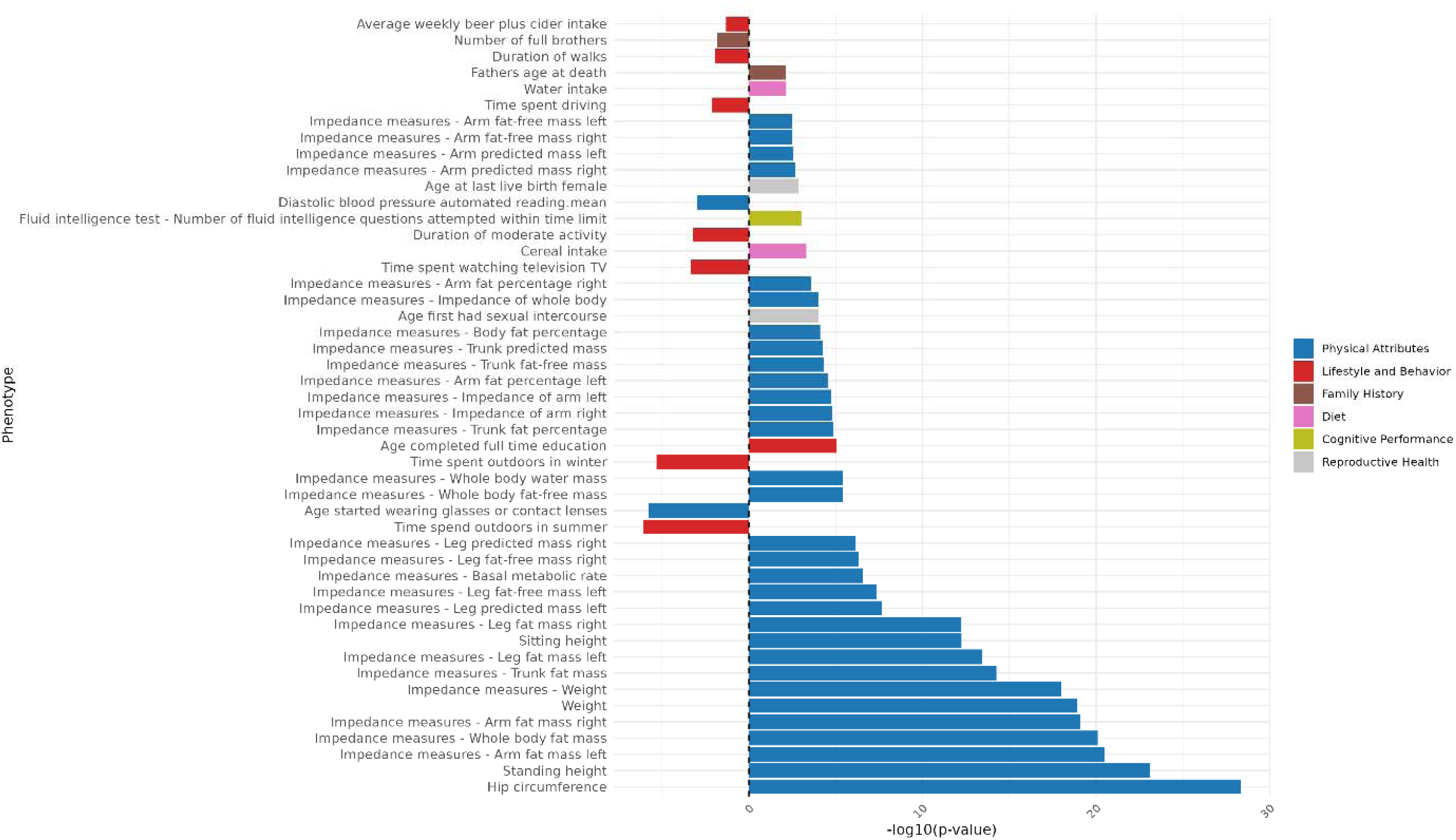
Genomic Principal Component 11 PCS association strength across continuous non-neuroimaging phenotypes. Phenotypes (y-axis) are plotted against the -log(10) p-value (x-axis). Phenotypes shown are significant after FDR-correction. The direction of the bar indicates the direction of association, where a right-hand bar indicates a positive association, and a left-hand bar a negative association. Phenotypes are color coded by phenotype category.

### Principal Component 12

**Figure 67:**
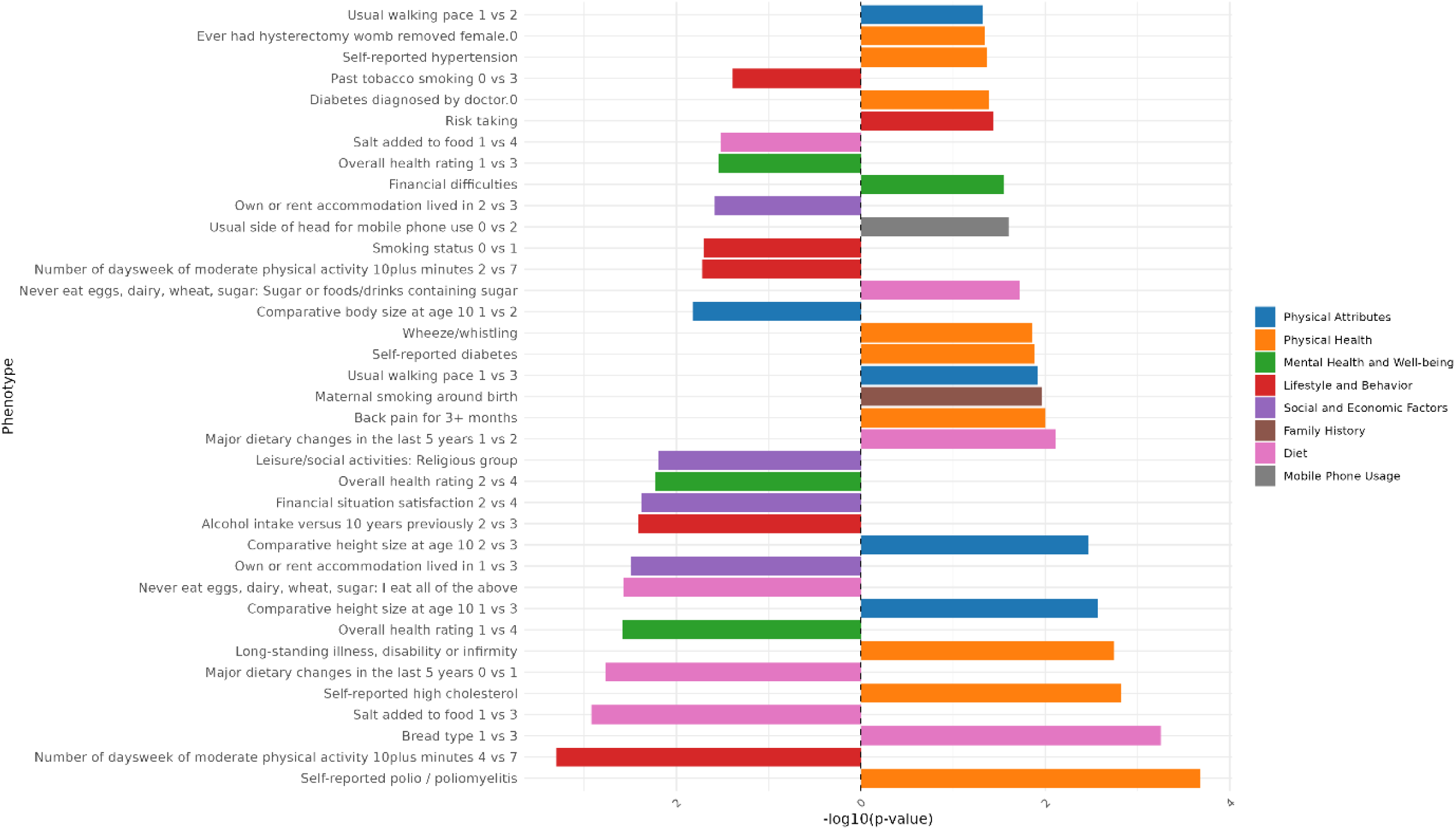
Genomic Principal Component 12 PCS association strength across binary/binarized non-neuroimaging phenotypes. Phenotypes (y-axis) are plotted against the -log(10) p-value (x-axis). Phenotypes shown are significant after FDR-correction. The direction of the bar indicates the direction of association, where a right-hand bar indicates a positive association, and a left-hand bar a negative association. In contrast analyses (e.g., comparing response 1 vs. response 3), the direction of the bar indicates the direction of the association in relation to the answers contrasted. Specifically, in a 1 vs. 3 contrast, a bar pointing to the right indicates an association with response 3, while a bar pointing to the left indicates an association with response 1. The phenotype “Diagnosed Vascular problems diagnosed by doctor.0” equates to “No Vascular problems diagnosed”. Phenotypes are color coded by phenotype category.

**Figure 68:**
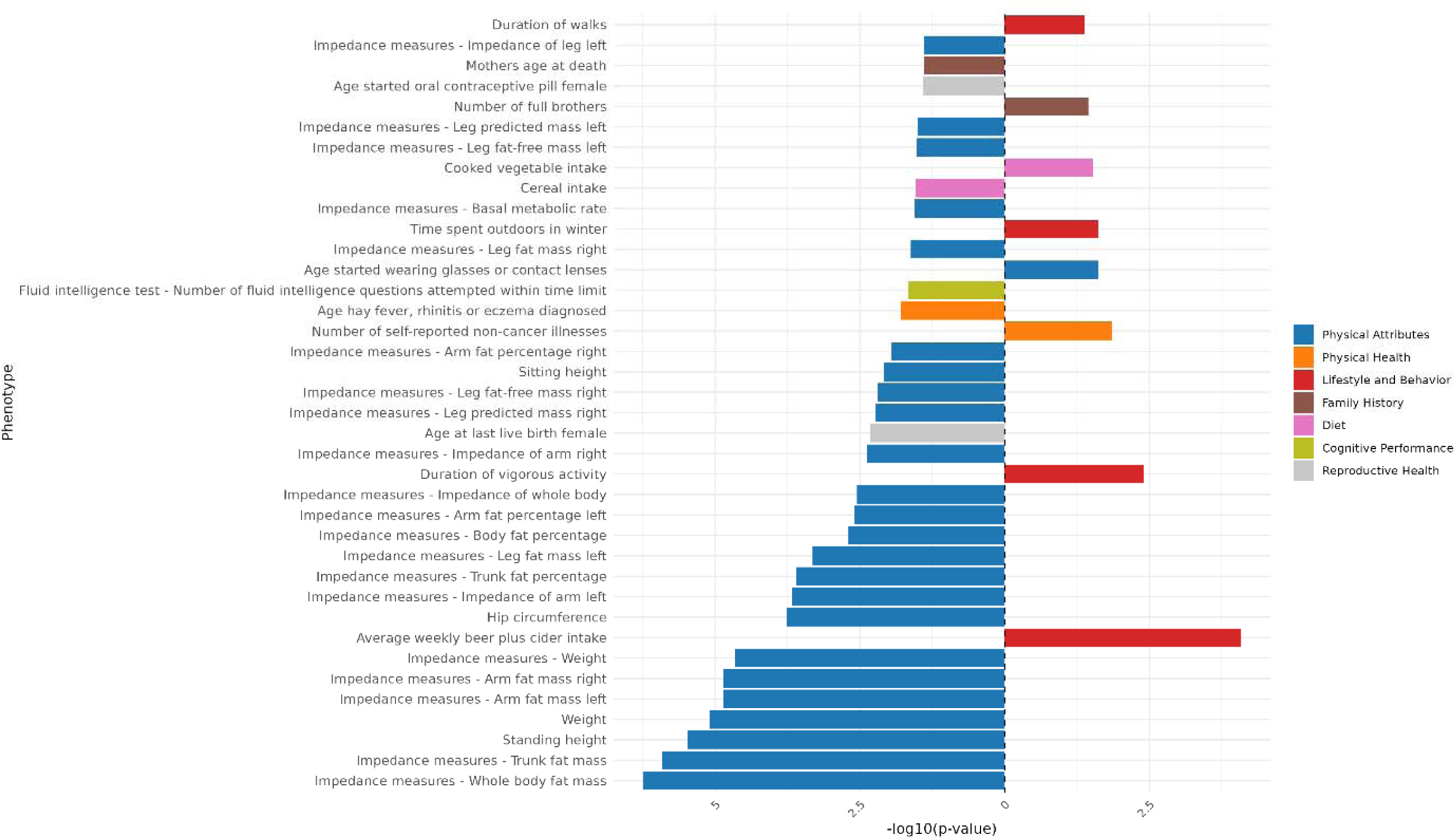
Genomic Principal Component 12 PCS association strength across continuous non-neuroimaging phenotypes. Phenotypes (y-axis) are plotted against the -log(10) p-value (x-axis). Phenotypes shown are significant after FDR-correction. The direction of the bar indicates the direction of association, where a right-hand bar indicates a positive association, and a left-hand bar a negative association. Phenotypes are color coded by phenotype category.

### Principal Component 13

**Figure 69:**
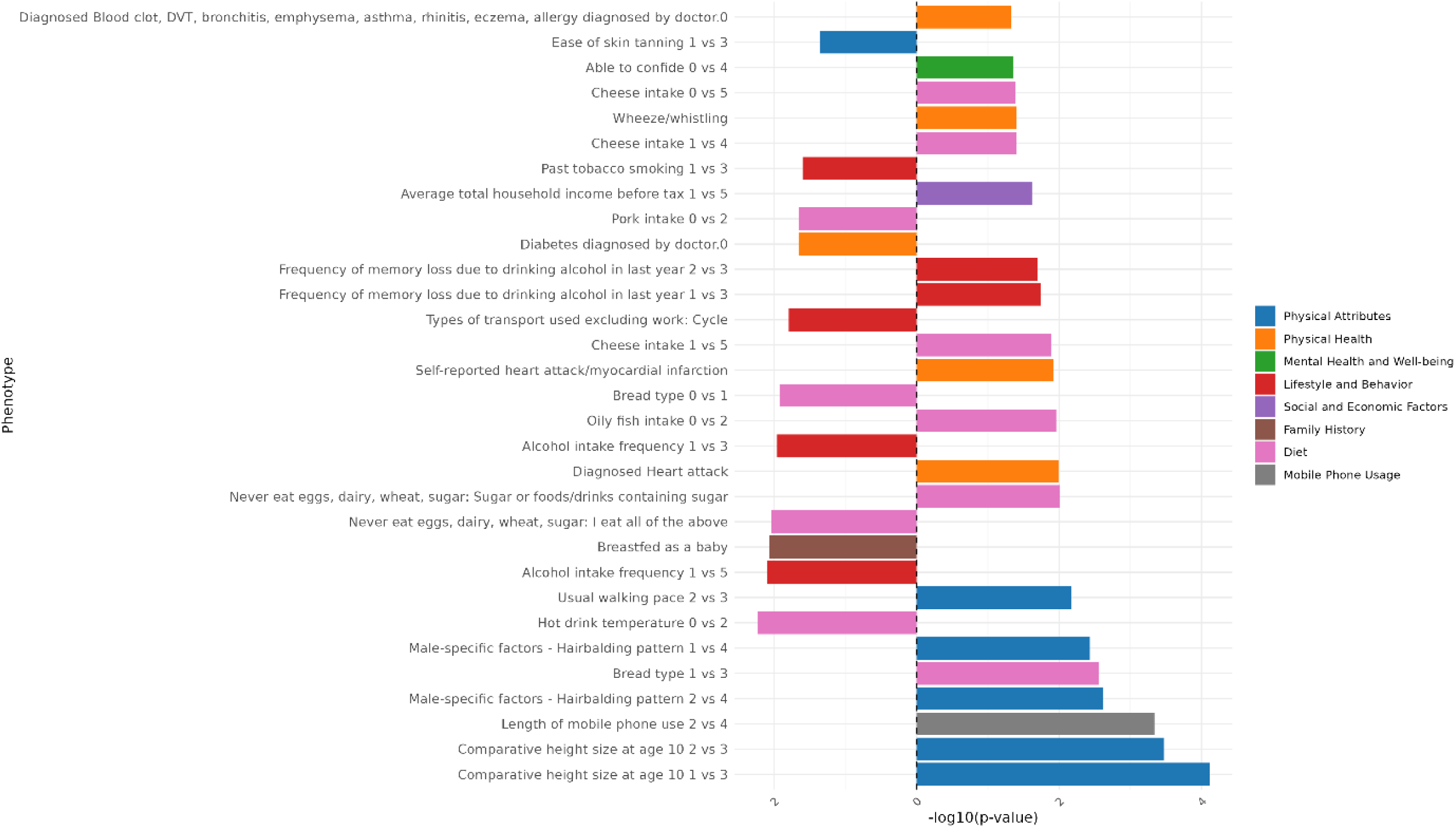
Genomic Principal Component 13 PCS association strength across binary/binarized non-neuroimaging phenotypes. Phenotypes (y-axis) are plotted against the -log(10) p-value (x-axis). Phenotypes shown are significant after FDR-correction. The direction of the bar indicates the direction of association, where a right-hand bar indicates a positive association, and a left-hand bar a negative association. In contrast analyses (e.g., comparing response 1 vs. response 3), the direction of the bar indicates the direction of the association in relation to the answers contrasted. Specifically, in a 1 vs. 3 contrast, a bar pointing to the right indicates an association with response 3, while a bar pointing to the left indicates an association with response 1. The phenotype “Diagnosed Vascular problems diagnosed by doctor.0” equates to “No Vascular problems diagnosed”. Phenotypes are color coded by phenotype category.

**Figure 70:**
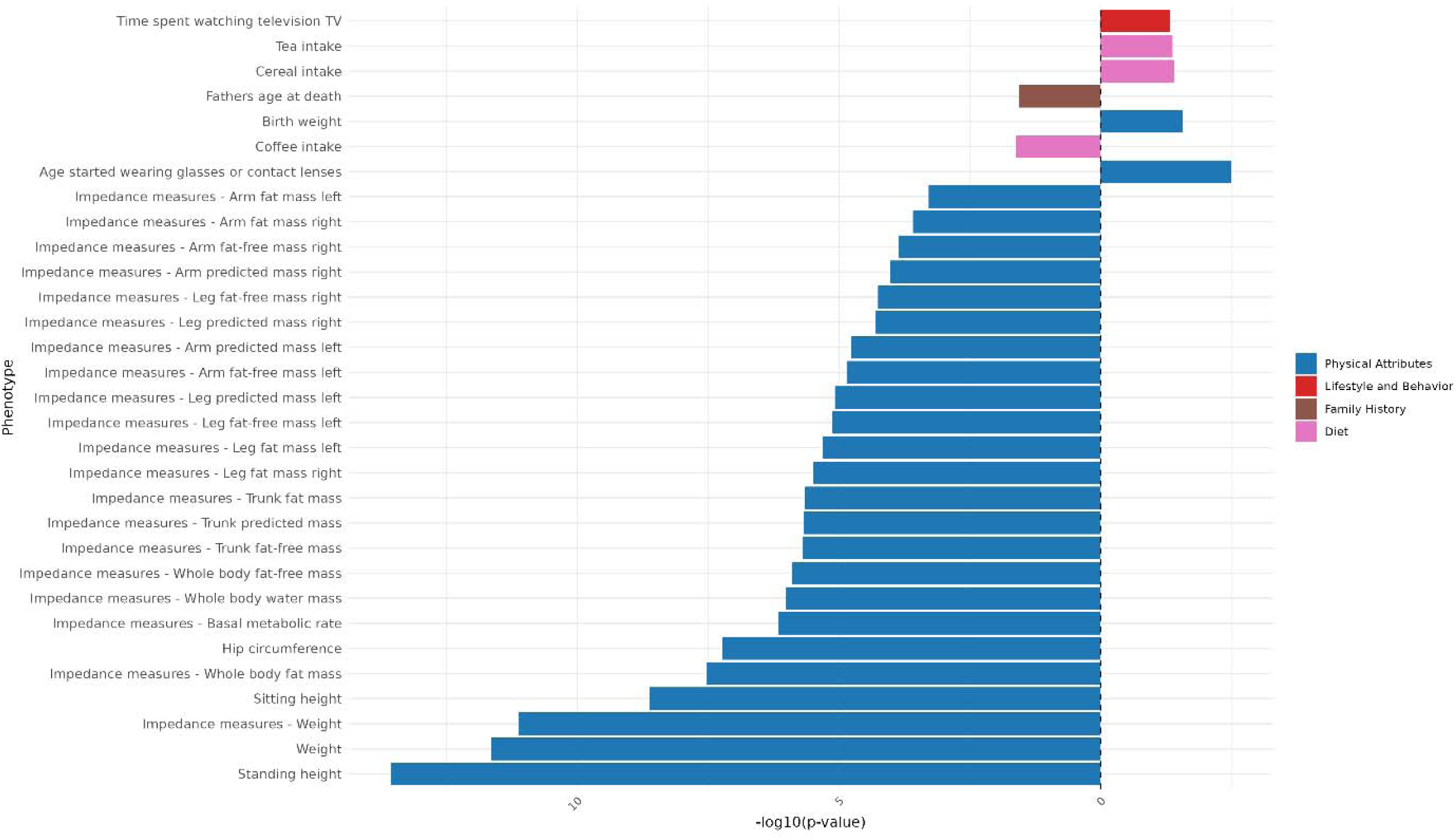
Genomic Principal Component 13 PCS association strength across continuous non-neuroimaging phenotypes. Phenotypes (y-axis) are plotted against the -log(10) p-value (x-axis). Phenotypes shown are significant after FDR-correction. The direction of the bar indicates the direction of association, where a right-hand bar indicates a positive association, and a left-hand bar a negative association. Phenotypes are color coded by phenotype category.

### Principal Component 14

**Figure 71:**
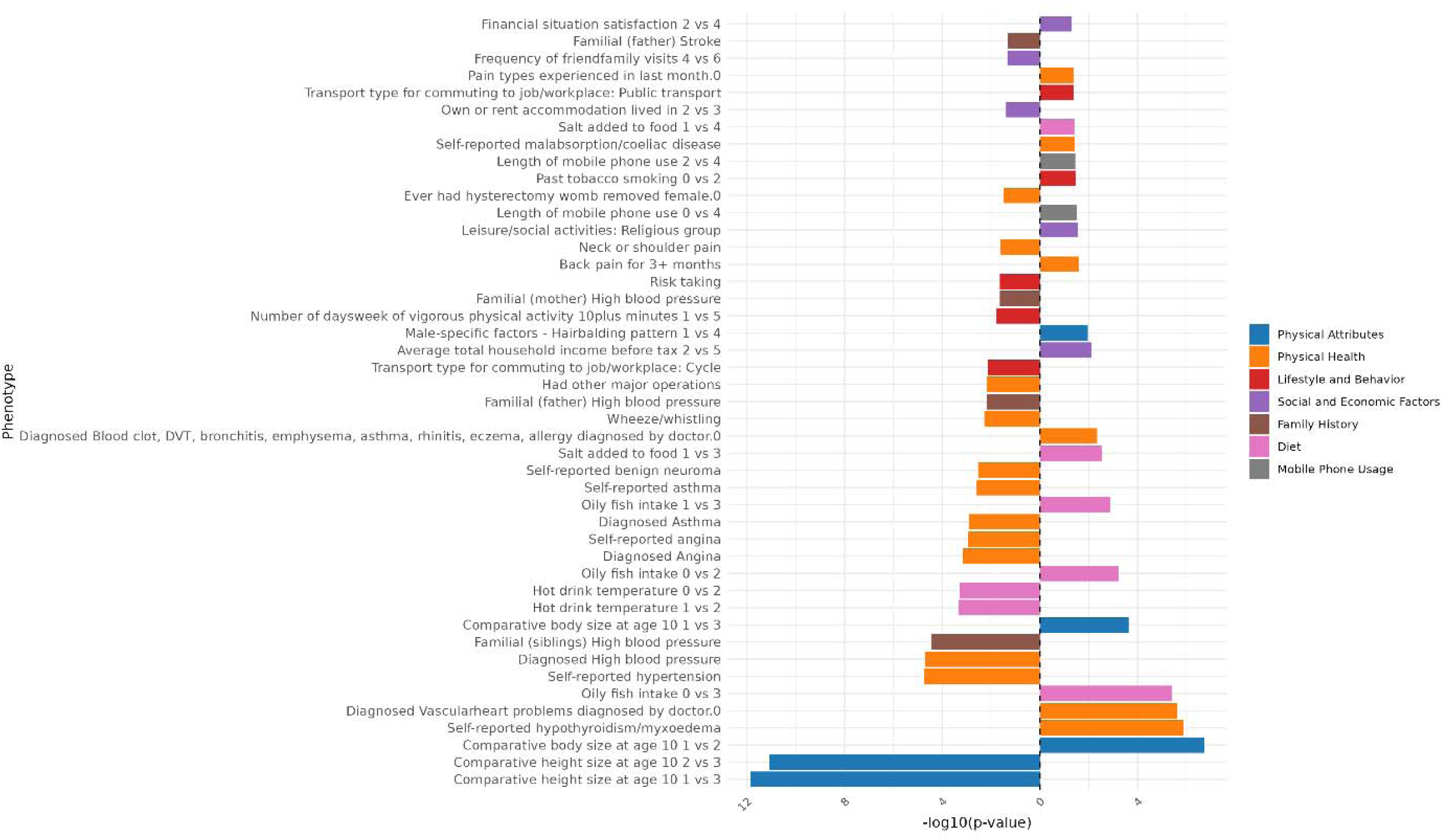
Genomic Principal Component 14 PCS association strength across binary/binarized non-neuroimaging phenotypes. Phenotypes (y-axis) are plotted against the -log(10) p-value (x-axis). Phenotypes shown are significant after FDR-correction. The direction of the bar indicates the direction of association, where a right-hand bar indicates a positive association, and a left-hand bar a negative association. In contrast analyses (e.g., comparing response 1 vs. response 3), the direction of the bar indicates the direction of the association in relation to the answers contrasted. Specifically, in a 1 vs. 3 contrast, a bar pointing to the right indicates an association with response 3, while a bar pointing to the left indicates an association with response 1. The phenotype “Diagnosed Vascular problems diagnosed by doctor.0” equates to “No Vascular problems diagnosed”. Phenotypes are color coded by phenotype category.

**Figure 72:**
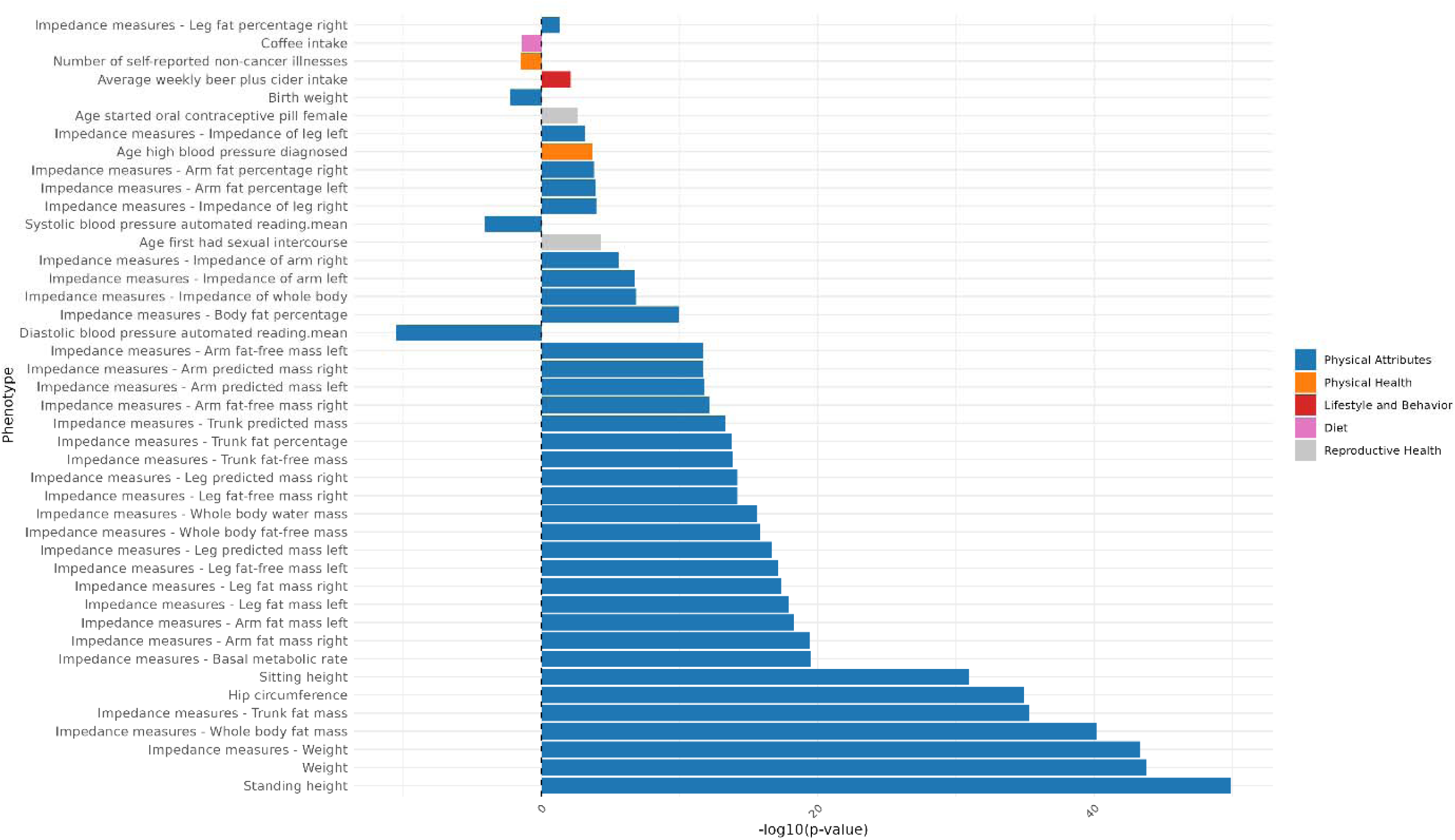
Genomic Principal Component 14 PCS association strength across continuous non-neuroimaging phenotypes. Phenotypes (y-axis) are plotted against the -log(10) p-value (x-axis). Phenotypes shown are significant after FDR-correction. The direction of the bar indicates the direction of association, where a right-hand bar indicates a positive association, and a left-hand bar a negative association. Phenotypes are color coded by phenotype category.

### Principal Component 15

**Figure 73:**
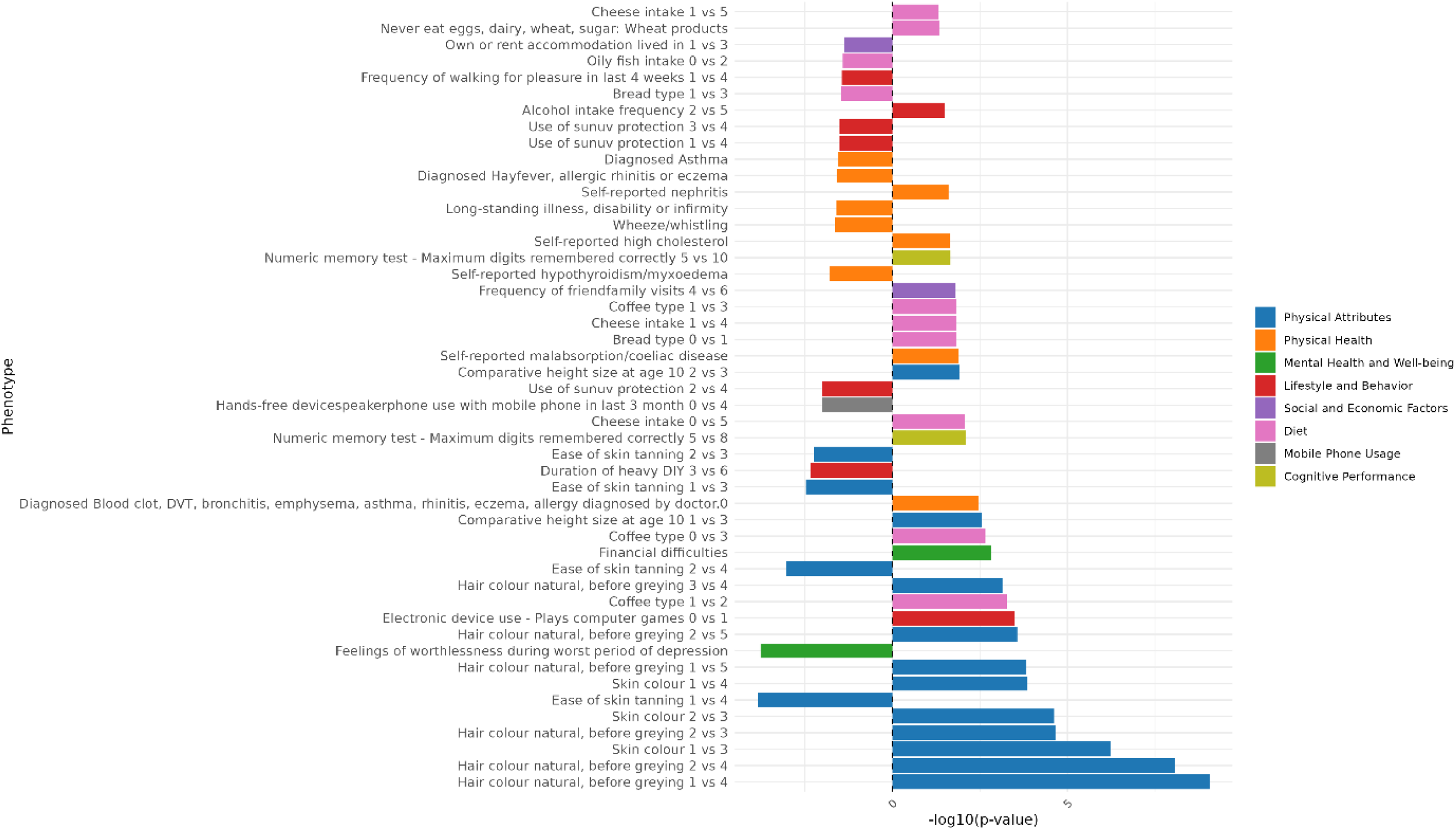
Genomic Principal Component 15 PCS association strength across binary/binarized non-neuroimaging phenotypes. Phenotypes (y-axis) are plotted against the -log(10) p-value (x-axis). Phenotypes shown are significant after FDR-correction. The direction of the bar indicates the direction of association, where a right-hand bar indicates a positive association, and a left-hand bar a negative association. In contrast analyses (e.g., comparing response 1 vs. response 3), the direction of the bar indicates the direction of the association in relation to the answers contrasted. Specifically, in a 1 vs. 3 contrast, a bar pointing to the right indicates an association with response 3, while a bar pointing to the left indicates an association with response 1. The phenotype “Diagnosed Vascular problems diagnosed by doctor.0” equates to “No Vascular problems diagnosed”. Phenotypes are color coded by phenotype category.

**Figure 74:**
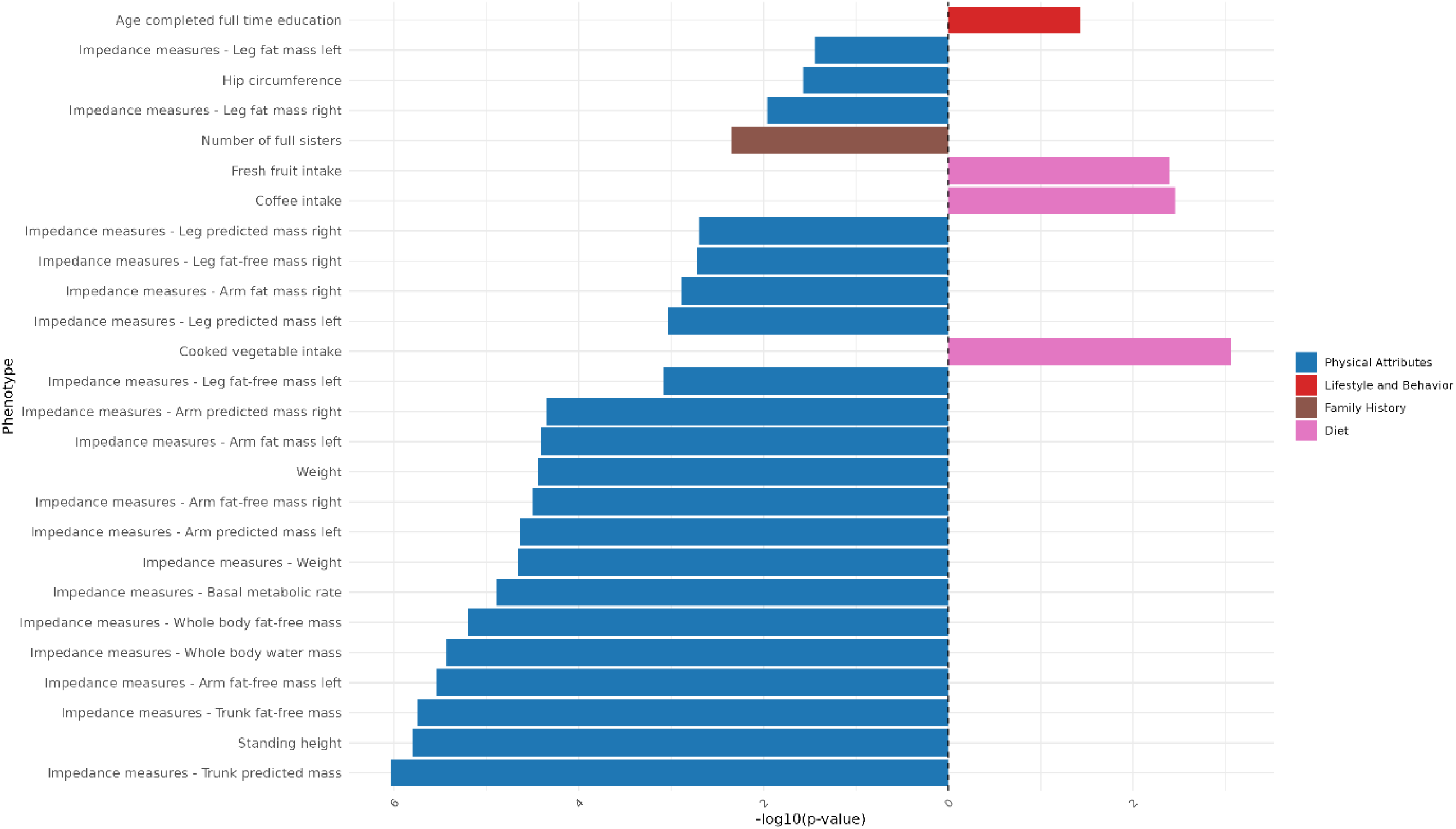
Genomic Principal Component 15 PCS association strength across continuous non-neuroimaging phenotypes. Phenotypes (y-axis) are plotted against the -log(10) p-value (x-axis). Phenotypes shown are significant after FDR-correction. The direction of the bar indicates the direction of association, where a right-hand bar indicates a positive association, and a left-hand bar a negative association. Phenotypes are color coded by phenotype category.

### Principal Component 16

**Figure 75:**
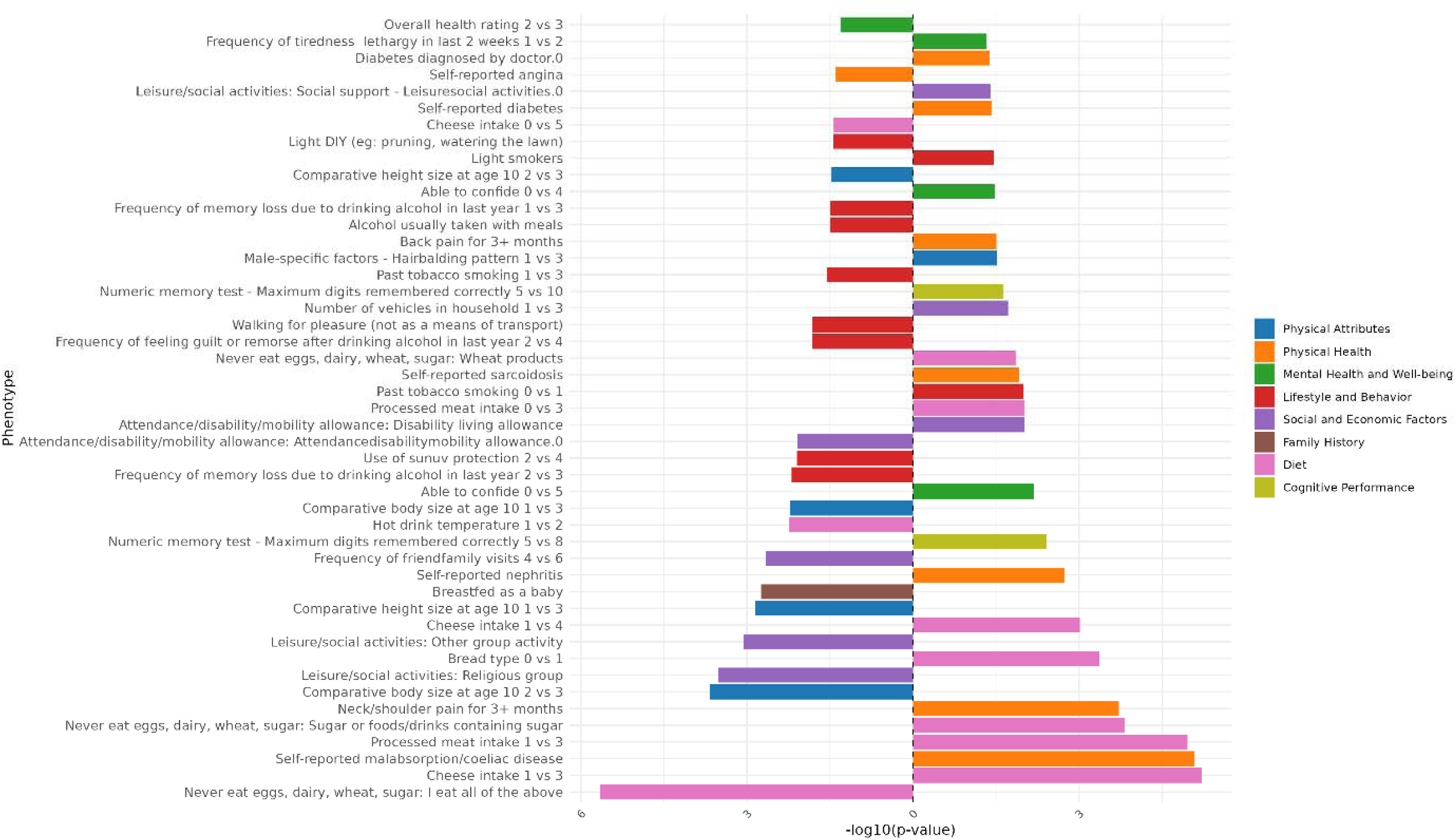
Genomic Principal Component 16 PCS association strength across binary/binarized non-neuroimaging phenotypes. Phenotypes (y-axis) are plotted against the -log(10) p-value (x-axis). Phenotypes shown are significant after FDR-correction. The direction of the bar indicates the direction of association, where a right-hand bar indicates a positive association, and a left-hand bar a negative association. In contrast analyses (e.g., comparing response 1 vs. response 3), the direction of the bar indicates the direction of the association in relation to the answers contrasted. Specifically, in a 1 vs. 3 contrast, a bar pointing to the right indicates an association with response 3, while a bar pointing to the left indicates an association with response 1. The phenotype “Diagnosed Vascular problems diagnosed by doctor.0” equates to “No Vascular problems diagnosed”. Phenotypes are color coded by phenotype category.

**Figure 76:**
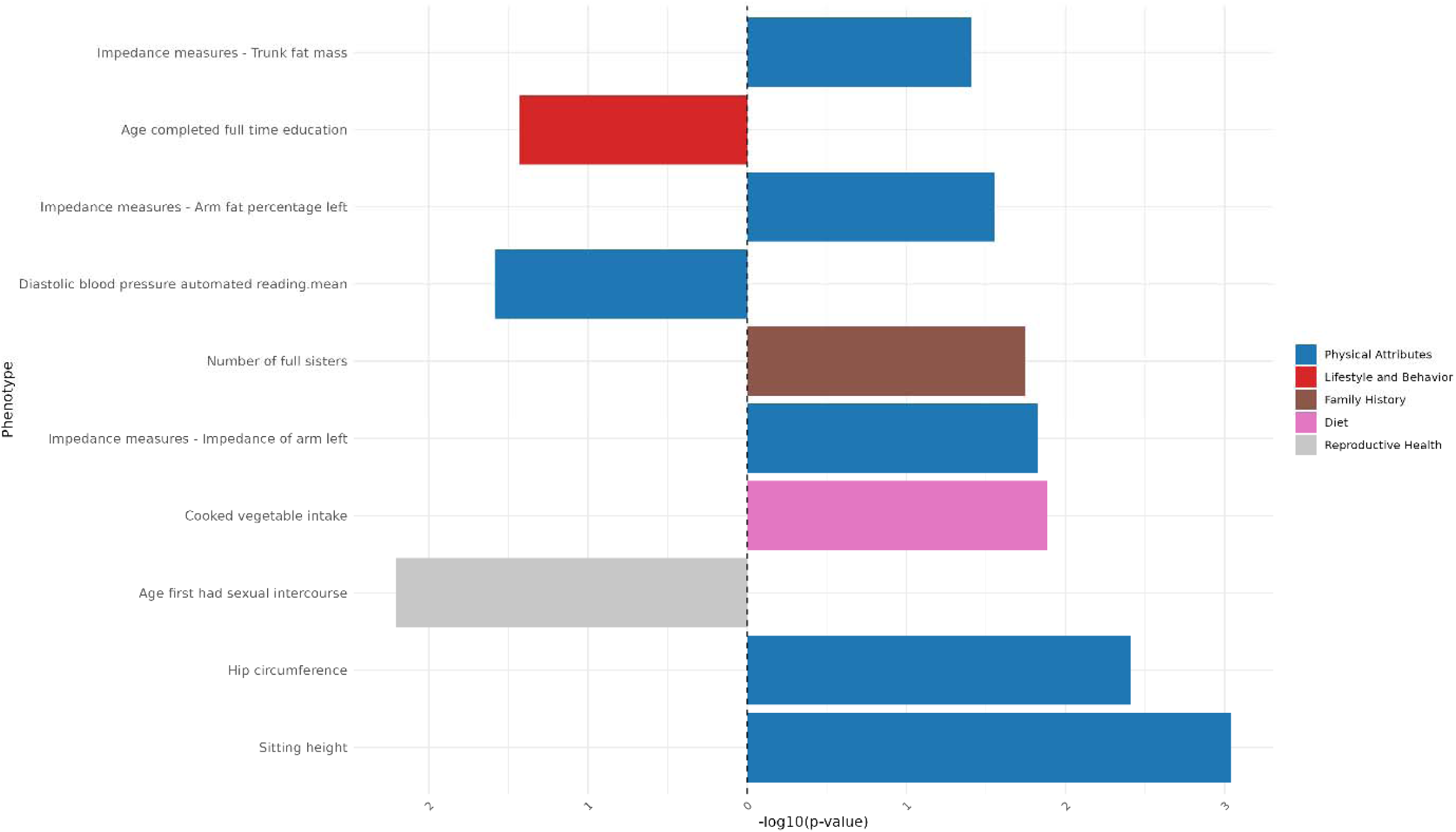
Genomic Principal Component 16 PCS association strength across continuous non-neuroimaging phenotypes. Phenotypes (y-axis) are plotted against the -log(10) p-value (x-axis). Phenotypes shown are significant after FDR-correction. The direction of the bar indicates the direction of association, where a right-hand bar indicates a positive association, and a left-hand bar a negative association. Phenotypes are color coded by phenotype category.

### Results of Genomic Independent Component Scores in Neuroimaging Phenotype space

**Figure 77:**
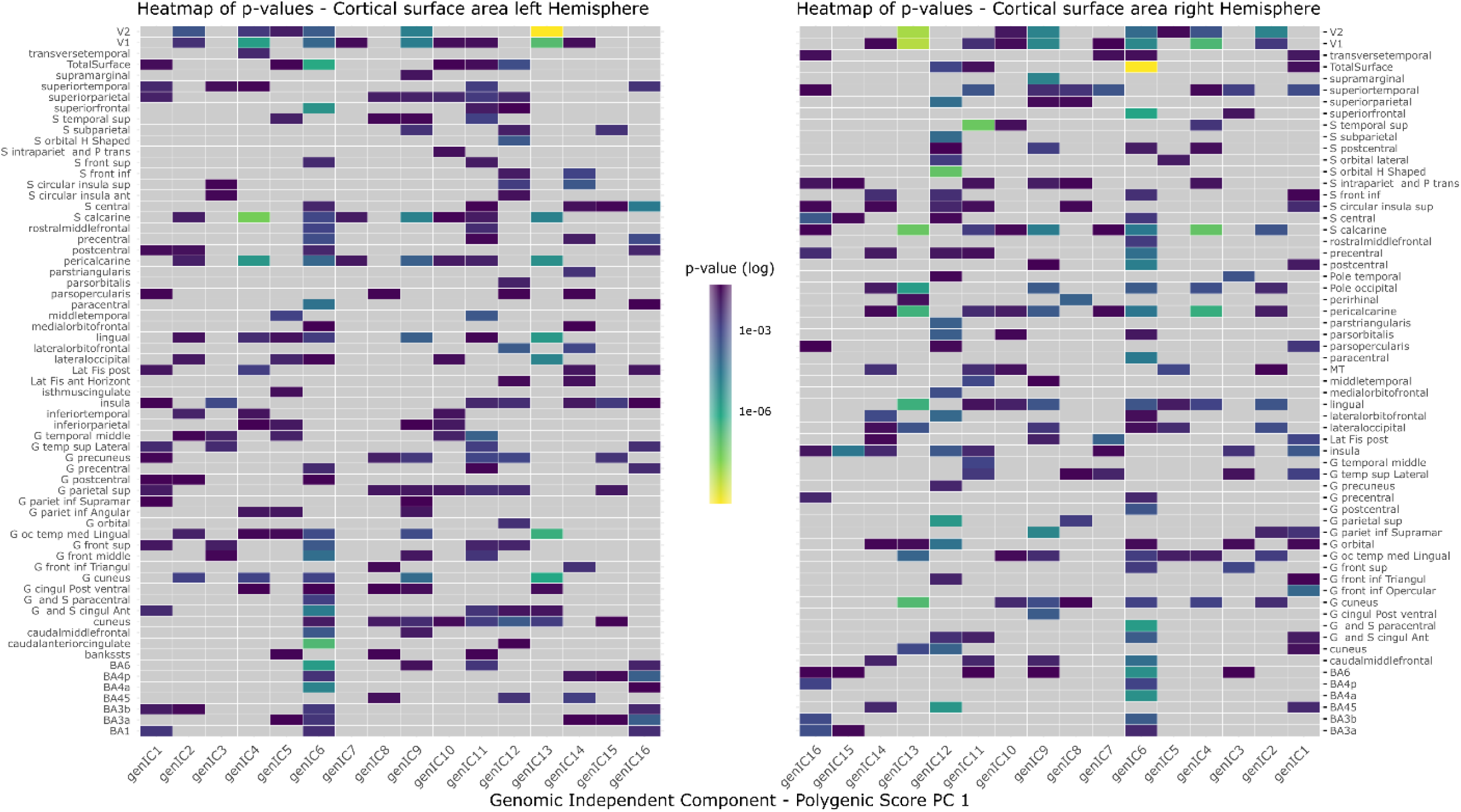
Heatmap indicating the power of association of genomic ICs with IDPs of cortical surface area. P-values are shown from high (blue) to low (yellow). Grey are non-significant associations post-MCC. Heatmaps are lateralized, with the left heatmap indicating the left hemisphere, and the right heatmap the right hemisphere, with the component order mirrored. If components are missing then they have no significant associations with any phenotype in this modality.

**Figure 78:**
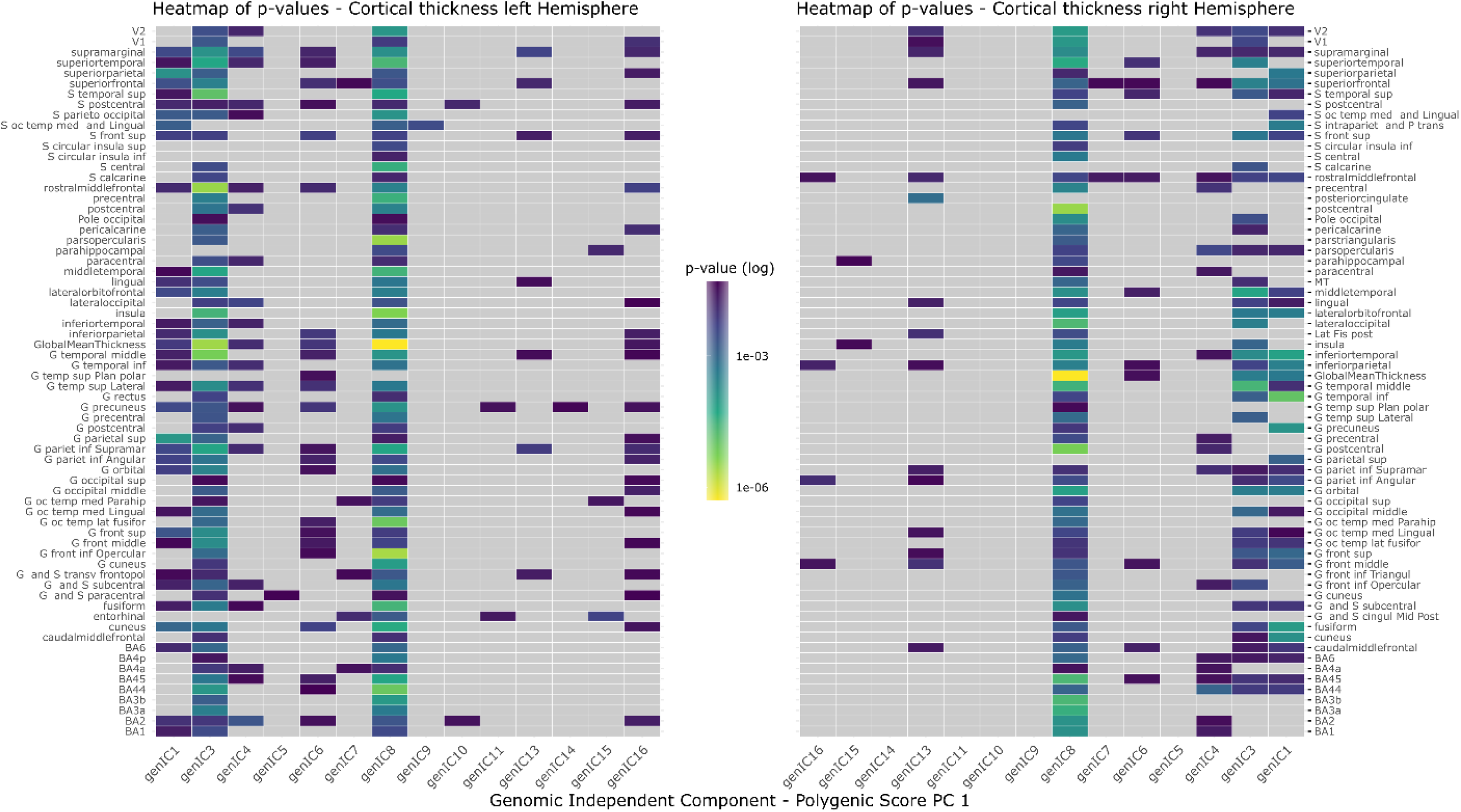
Heatmap indicating the power of association of genomic ICs with IDPs of cortical thickness. P-values are shown from high (blue) to low (yellow). Grey are non-significant associations post-MCC. Heatmaps are lateralized, with the left heatmap indicating the left hemisphere, and the right heatmap the right hemisphere, with the component order mirrored. If components are missing then they have no significant associations with any phenotype in this modality.

**Figure 79:**
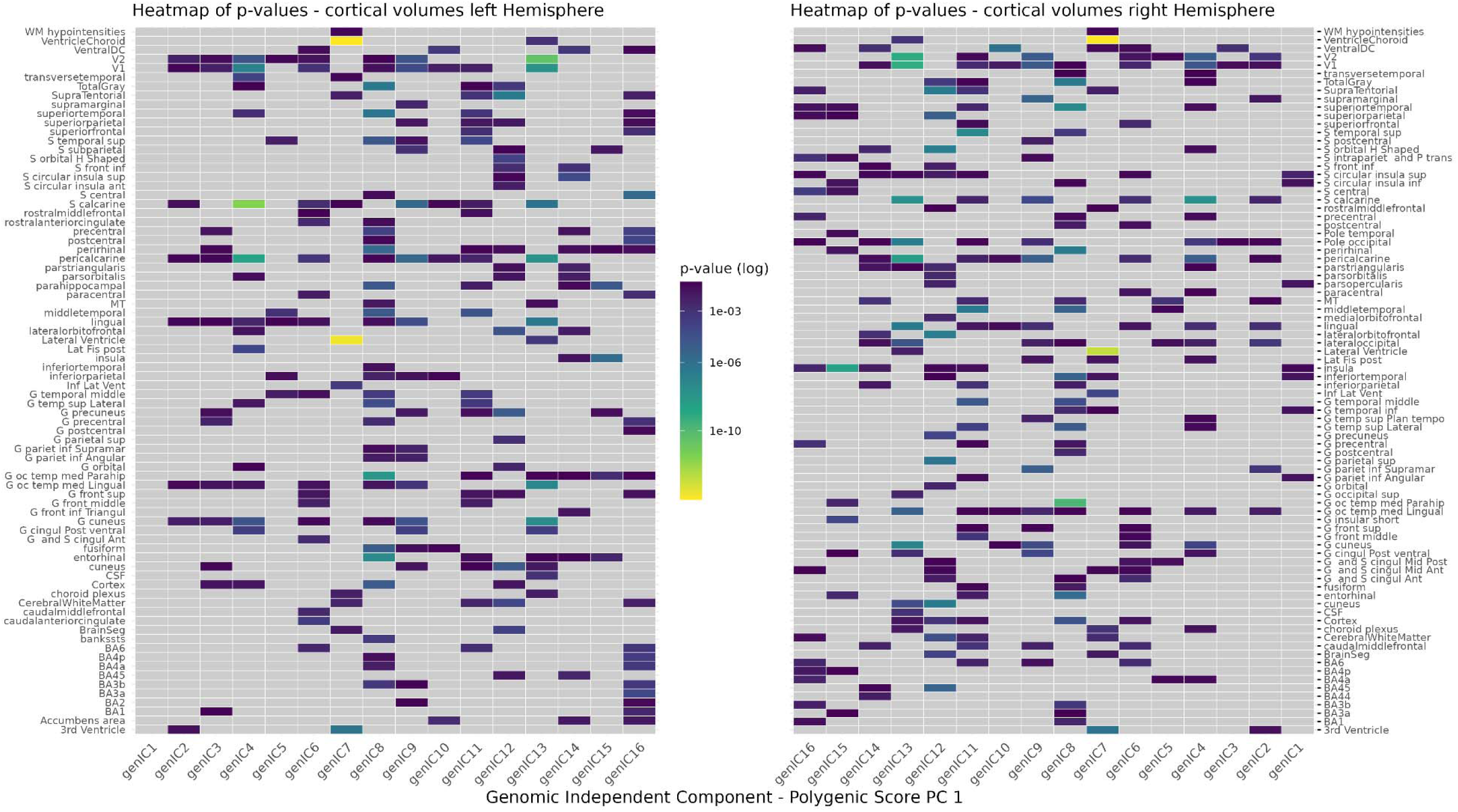
Heatmap indicating the power of association of genomic ICs with IDPs of cortical volume. P-values are shown from high (blue) to low (yellow). Grey are non-significant associations post-MCC. Heatmaps are lateralized, with the left heatmap indicating the left hemisphere, and the right heatmap the right hemisphere, with the component order mirrored. If components are missing then they have no significant associations with any phenotype in this modality.

**Figure 80:**
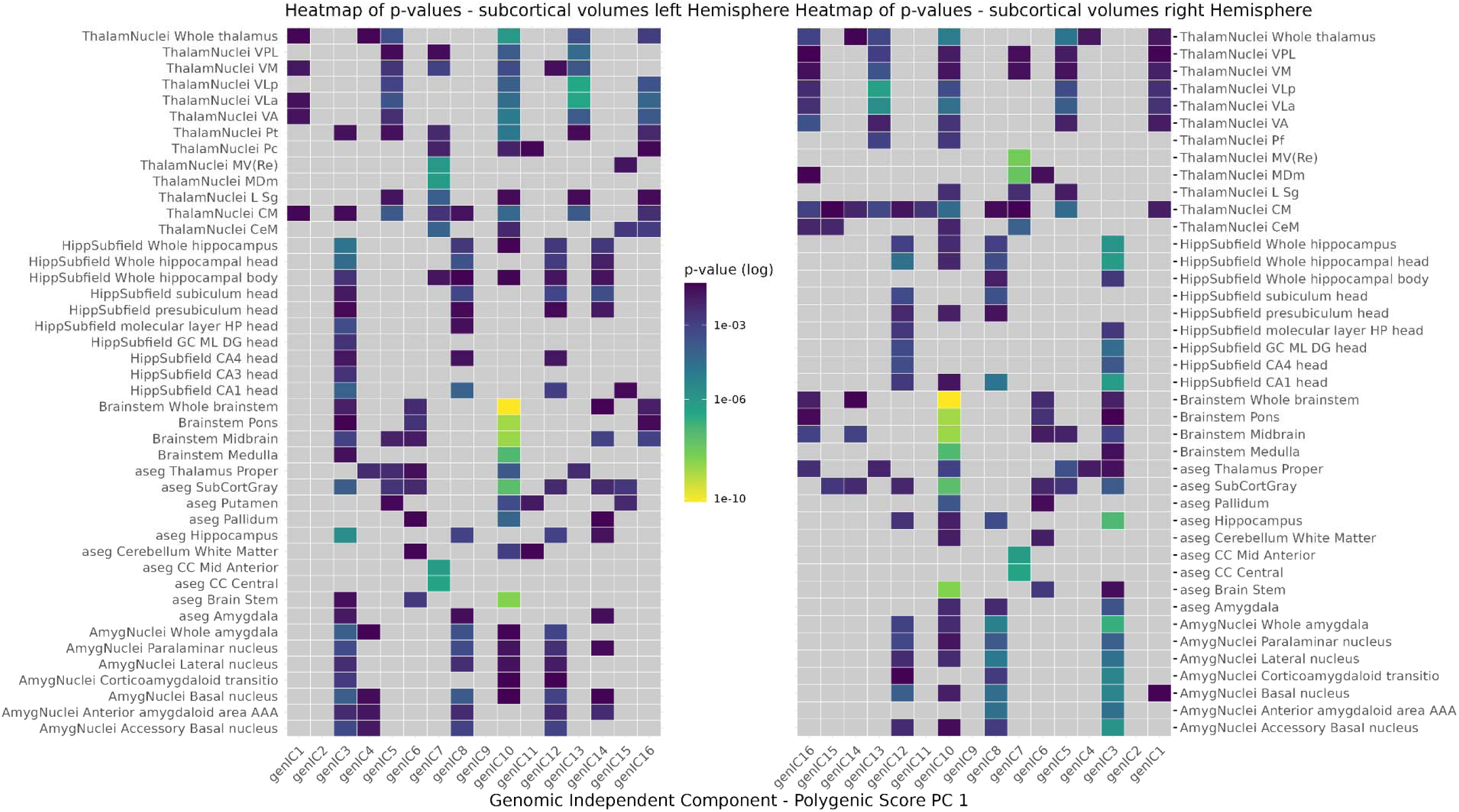
Heatmap indicating the power of association of genomic ICs with IDPs of subcortical volume. P-values are shown from high (blue) to low (yellow). Grey are non-significant associations post-MCC. Heatmaps are lateralized, with the left heatmap indicating the left hemisphere, and the right heatmap the right hemisphere, with the component order mirrored. If components are missing then they have no significant associations with any phenotype in this modality.

**Figure 81:**
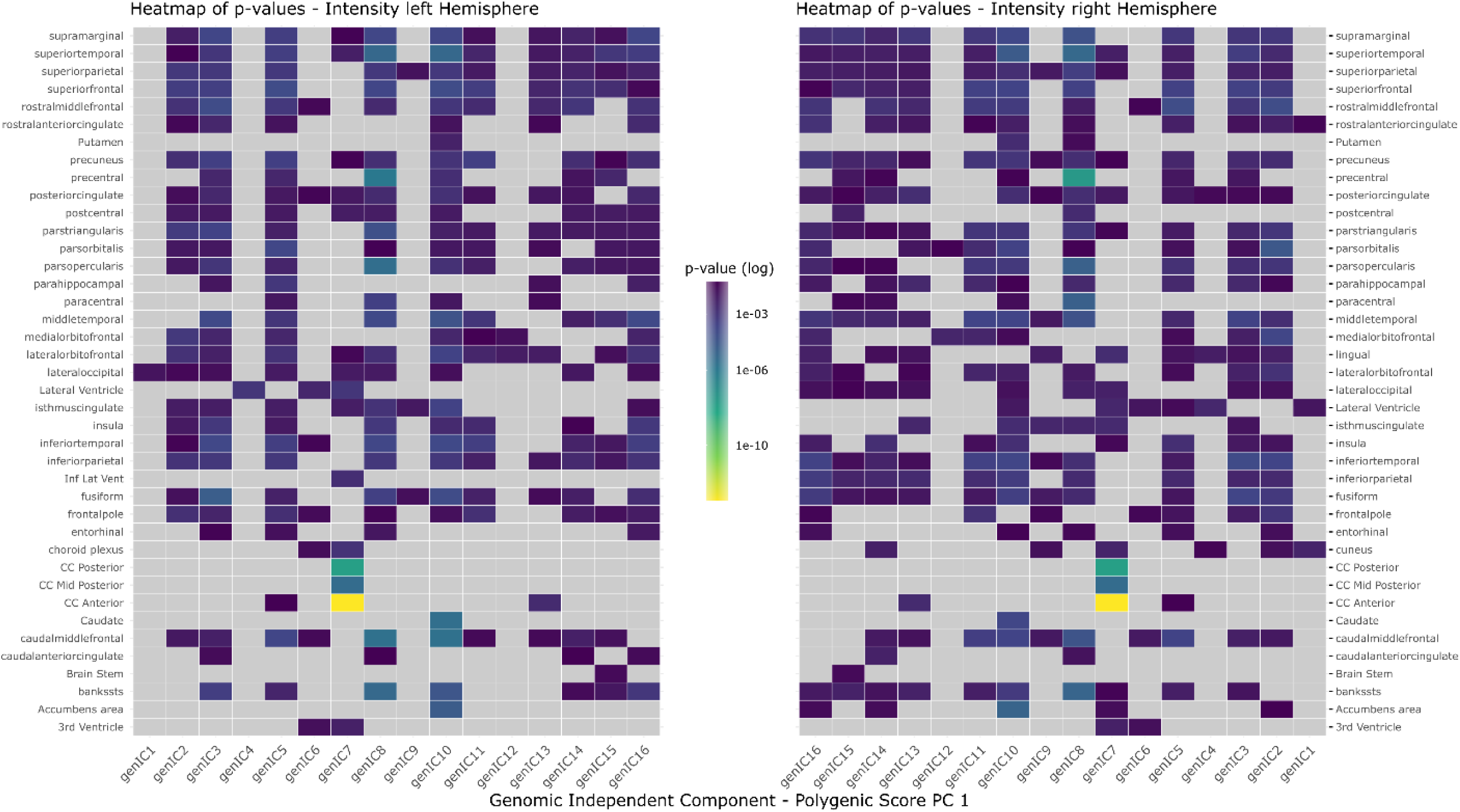
Heatmap indicating the power of association of genomic ICs with IDPs of intensity. P-values are shown from high (blue) to low (yellow). Grey are non-significant associations post-MCC. Heatmaps are lateralized, with the left heatmap indicating the left hemisphere, and the right heatmap the right hemisphere, with the component order mirrored. If components are missing then they have no significant associations with any phenotype in this modality.

**Figure 82:**
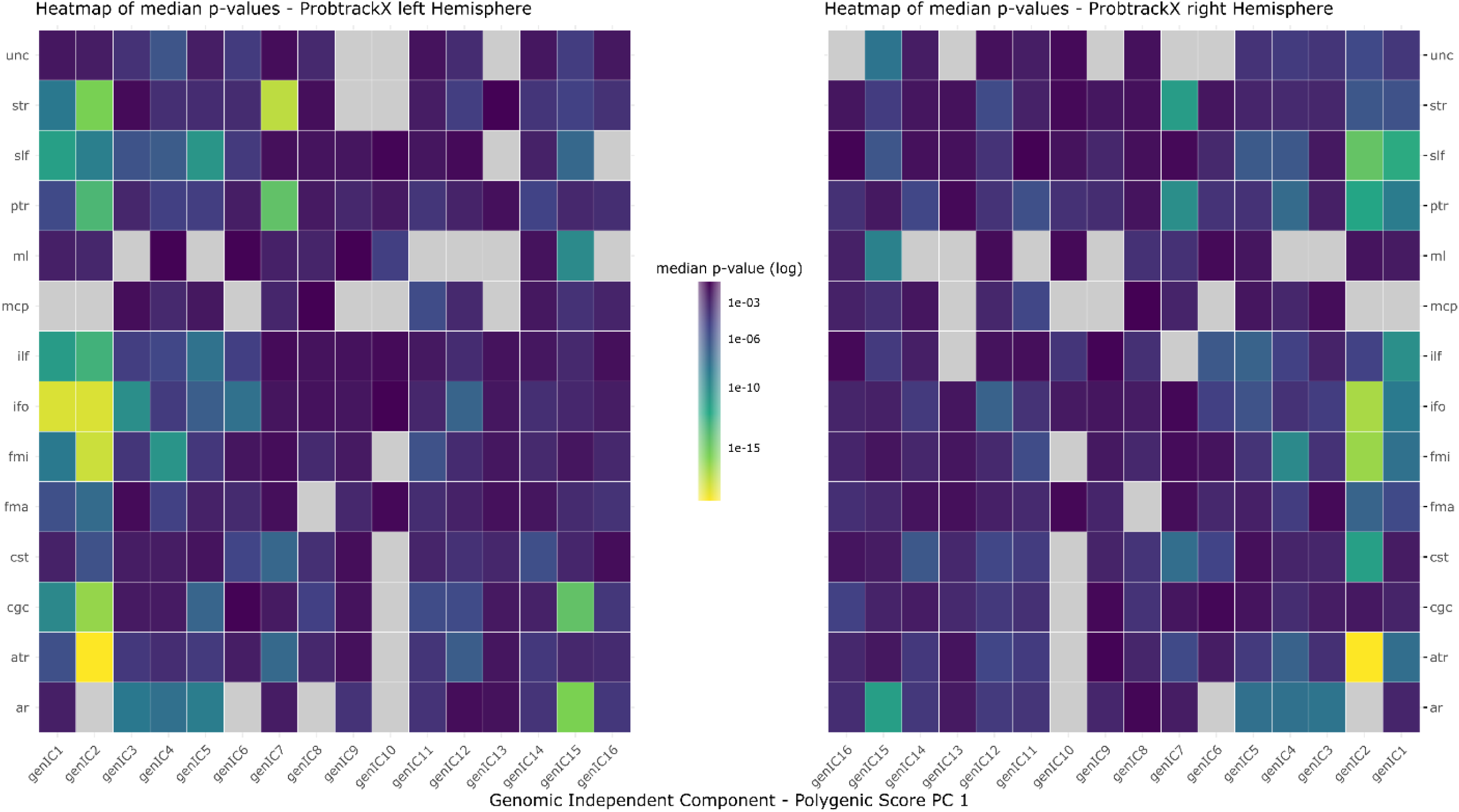
Heatmap indicating the power of association of genomic ICs with IDPs of dMRI ProbtrackX. P-values are shown from high (blue) to low (yellow). P-values have been aggregated into one median p-value across ProbtrackX modalities (e.g. MD, AD, FA, L1, etc.) to reflect the association power with white matter tracts. Grey are non-significant associations post-MCC. Heatmaps are lateralized, with the left heatmap indicating the left hemisphere, and the right heatmap the right hemisphere, with the component order mirrored. If components are missing then they have no significant associations with any phenotype in this modality.

**Figure 83:**
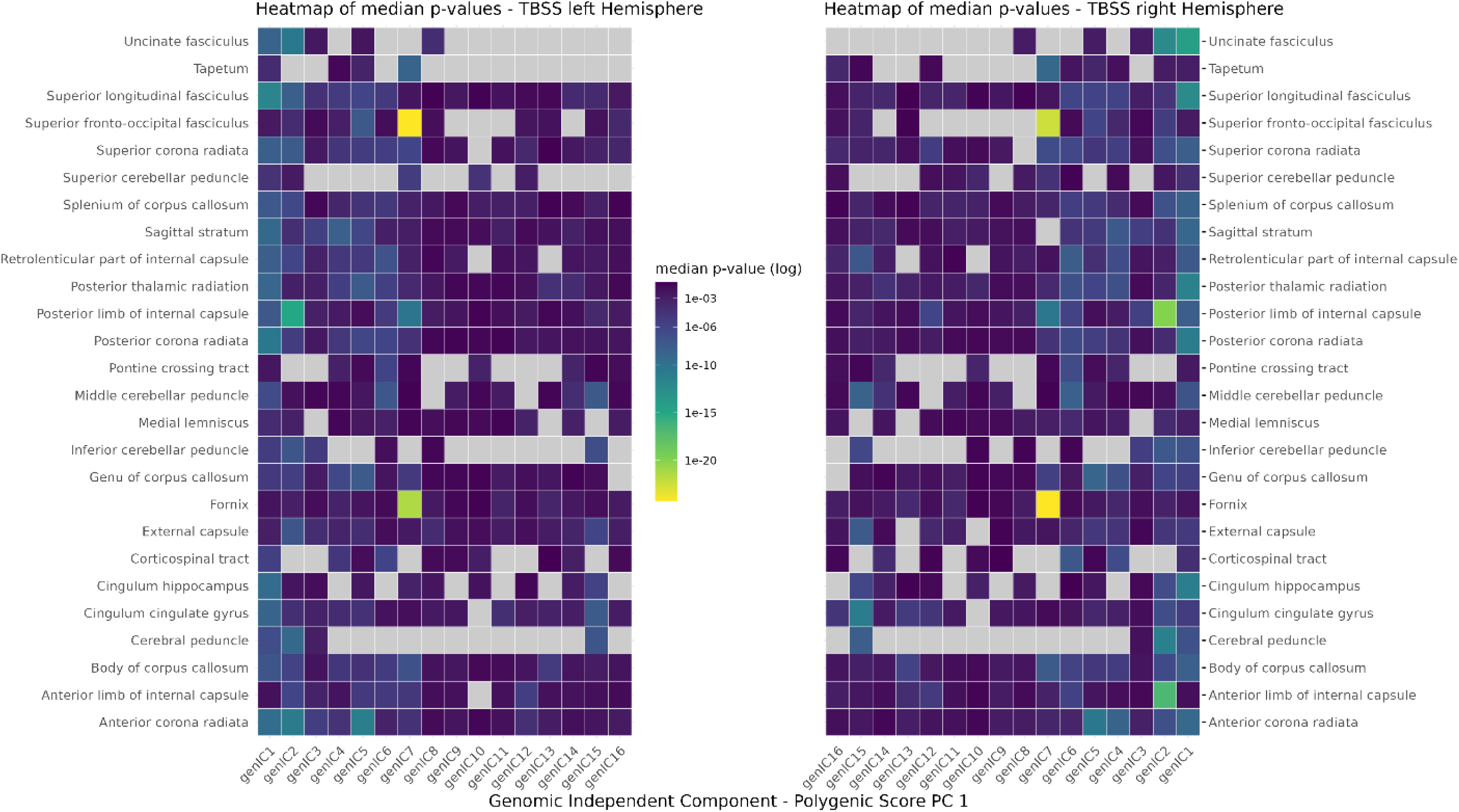
Heatmap indicating the power of association of genomic ICs with IDPs of dMRI TBSS. P-values are shown from high (blue) to low (yellow). P-values have been aggregated into one median p-value across TBSS modalities (e.g. MD, AD, FA, L1, etc.) to reflect the association power with white matter tracts. Grey are non-significant associations post-MCC. Heatmaps are lateralized, with the left heatmap indicating the left hemisphere, and the right heatmap the right hemisphere, with the component order mirrored. If components are missing then they have no significant associations with any phenotype in this modality.

**Figure 84:**
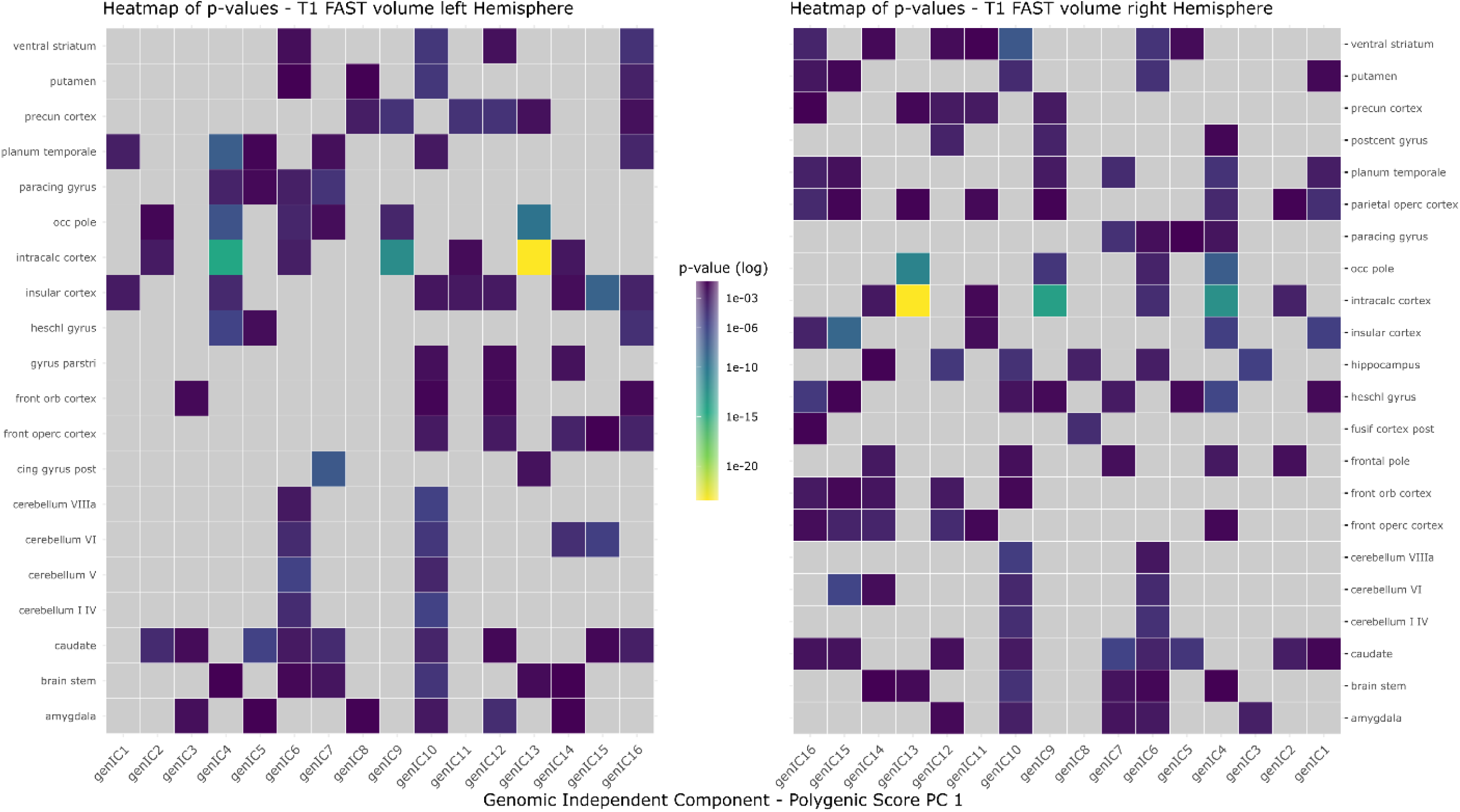
Heatmap indicating the power of association of genomic ICs with IDPs of T1-FAST volume. P-values are shown from high (blue) to low (yellow). Grey are non-significant associations post-MCC. Heatmaps are lateralized, with the left heatmap indicating the left hemisphere, and the right heatmap the right hemisphere, with the component order mirrored. If components are missing then they have no significant associations with any phenotype in this modality.

**Figure 85:**
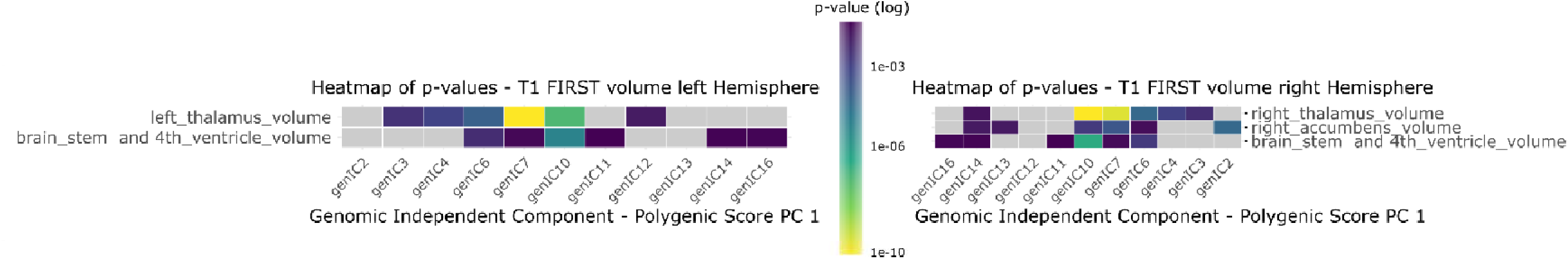
Heatmap indicating the power of association of genomic ICs with IDPs of T1-FIRST volume. P-values are shown from high (blue) to low (yellow). Grey are non-significant associations post-MCC. Heatmaps are lateralized, with the left heatmap indicating the left hemisphere, and the right heatmap the right hemisphere, with the component order mirrored. If components are missing then they have no significant associations with any phenotype in this modality.

**Figure 86:**
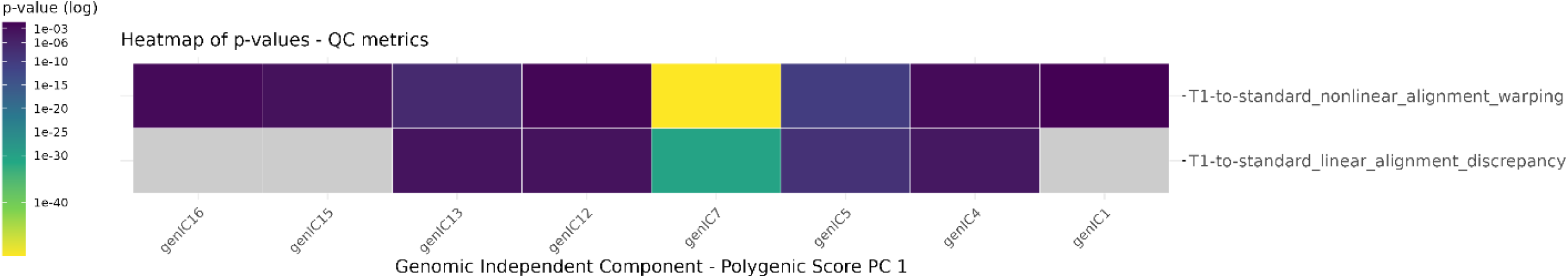
Heatmap indicating the power of association of genomic ICs with IDPs of QC metrics. P-values are shown from high (blue) to low (yellow). Grey are non-significant associations post-MCC. If components are missing then they have no significant associations with any phenotype in this modality.

### Results of Genomic Principal Component Scores in Neuroimaging Phenotype space

**Figure 87:**
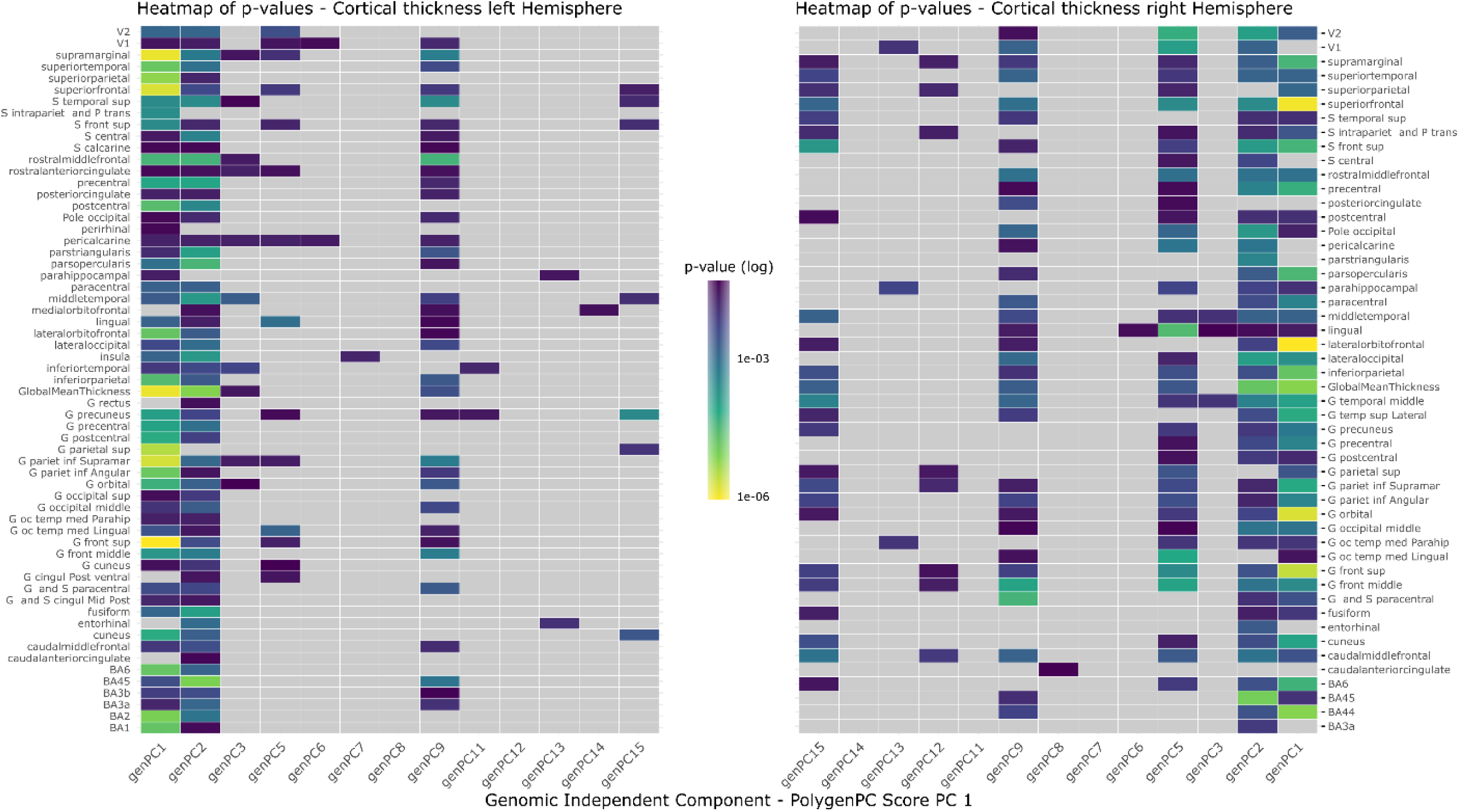
Heatmap indicating the power of association of genomic PCs with IDPs of cortical surface area. P-values are shown from high (blue) to low (yellow). Grey are non-significant associations post-MCC. Heatmaps are lateralized, with the left heatmap indicating the left hemisphere, and the right heatmap the right hemisphere, with the component order mirrored. If components are missing then they have no significant associations with any phenotype in this modality.

**Figure 88:**
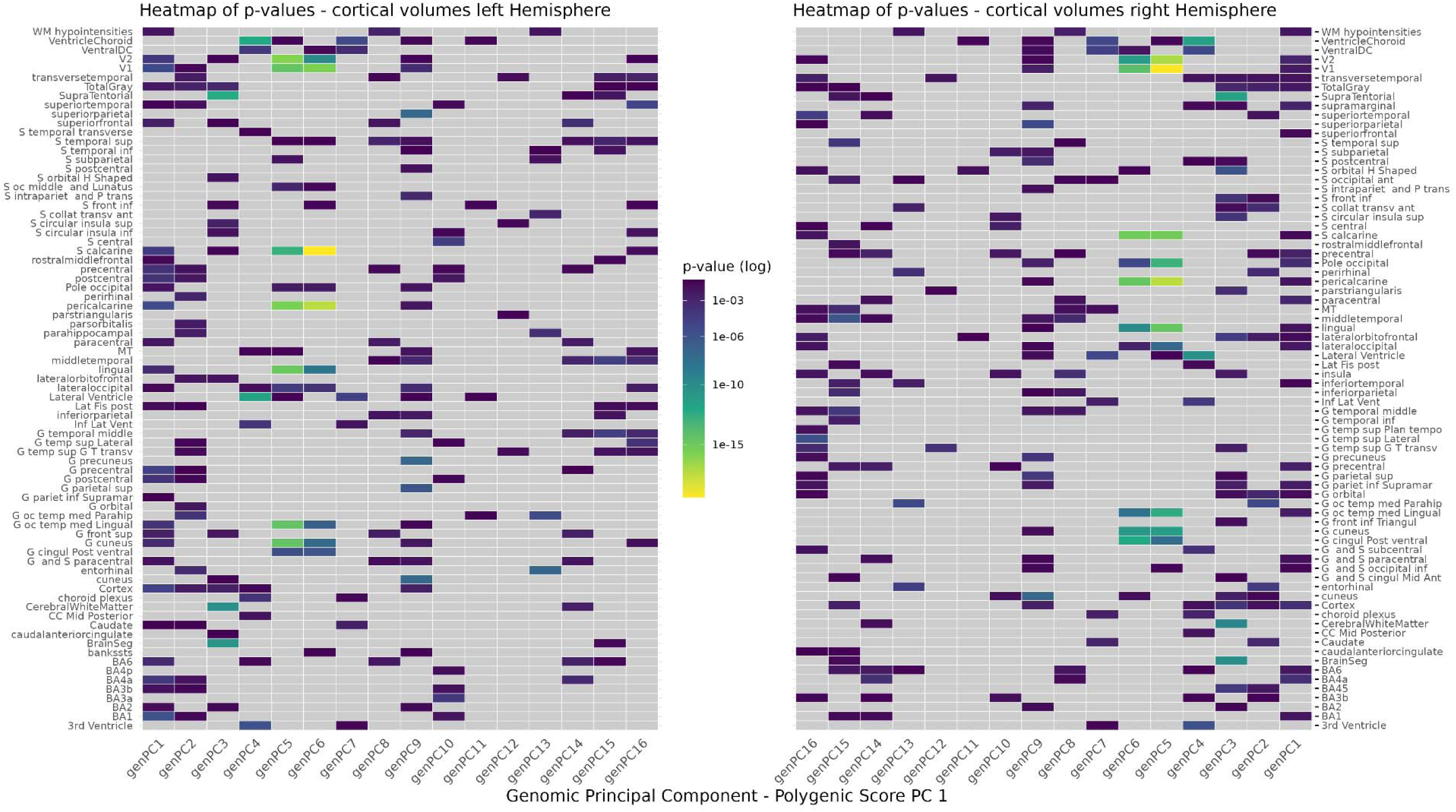
Heatmap indicating the power of association of genomic PCs with IDPs of cortical thickness. P-values are shown from high (blue) to low (yellow). Grey are non-significant associations post-MCC. Heatmaps are lateralized, with the left heatmap indicating the left hemisphere, and the right heatmap the right hemisphere, with the component order mirrored. If components are missing then they have no significant associations with any phenotype in this modality pre-MCC.

**Figure 89:**
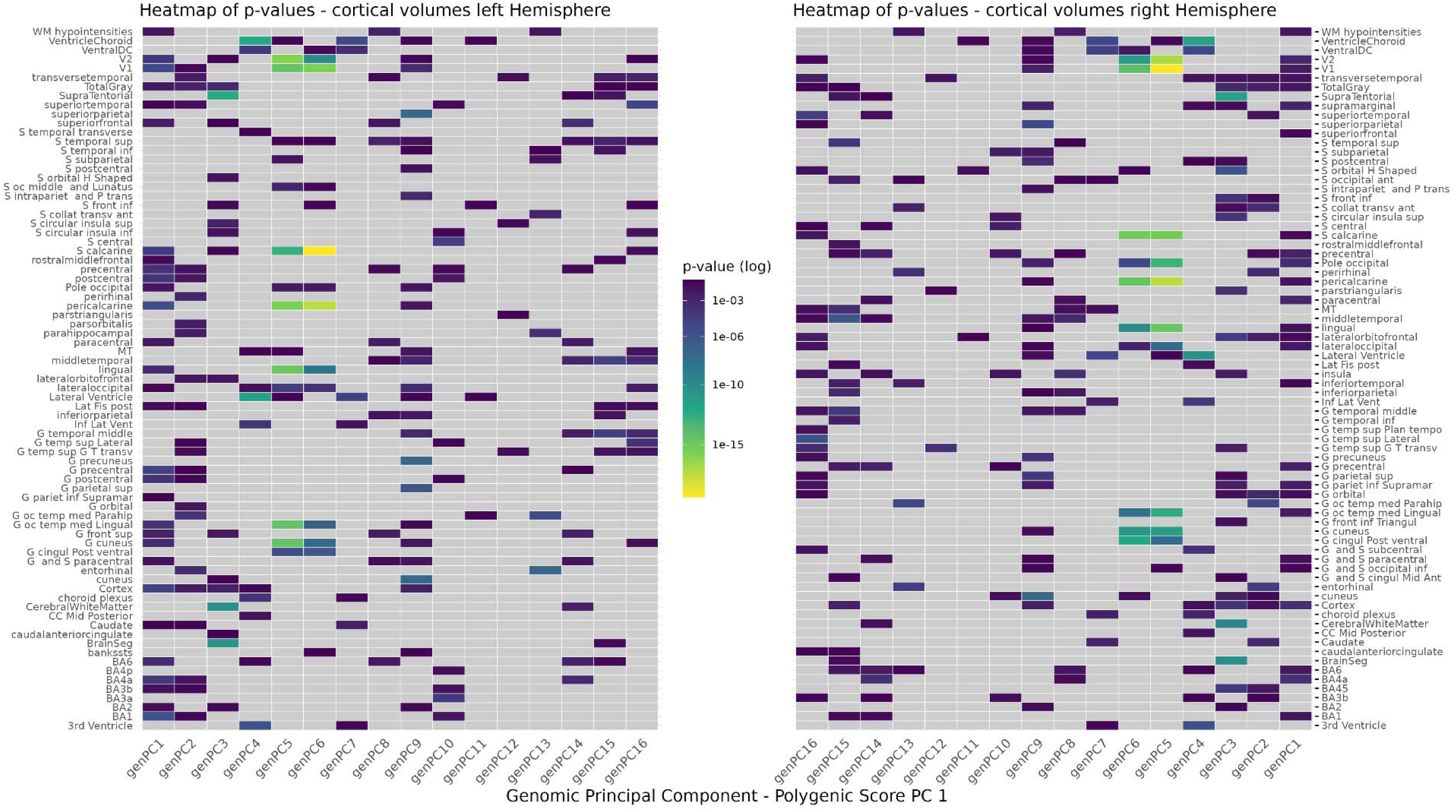
Heatmap indicating the power of association of genomic PCs with IDPs of cortical volume. P-values are shown from high (blue) to low (yellow). Grey are non-significant associations post-MCC. Heatmaps are lateralized, with the left heatmap indicating the left hemisphere, and the right heatmap the right hemisphere, with the component order mirrored. If components are missing then they have no significant associations with any phenotype in this modality.

**Figure 90:**
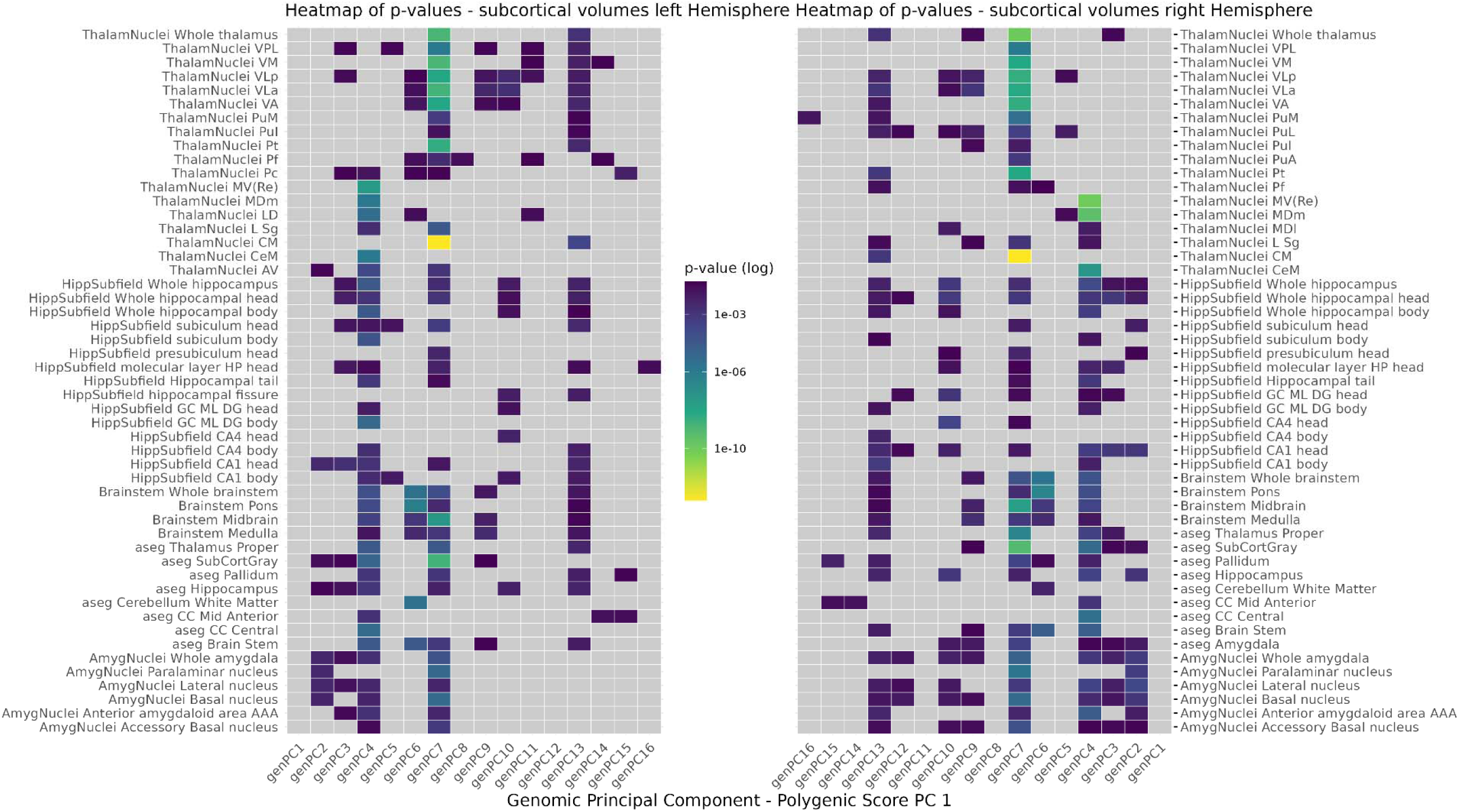
Heatmap indicating the power of association of genomic PCs with IDPs of subcortical volume. P-values are shown from high (blue) to low (yellow). Grey are non-significant associations post-MCC. Heatmaps are lateralized, with the left heatmap indicating the left hemisphere, and the right heatmap the right hemisphere, with the component order mirrored. If components are missing then they have no significant associations with any phenotype in this modality pre-MCC.

**Figure 91:**
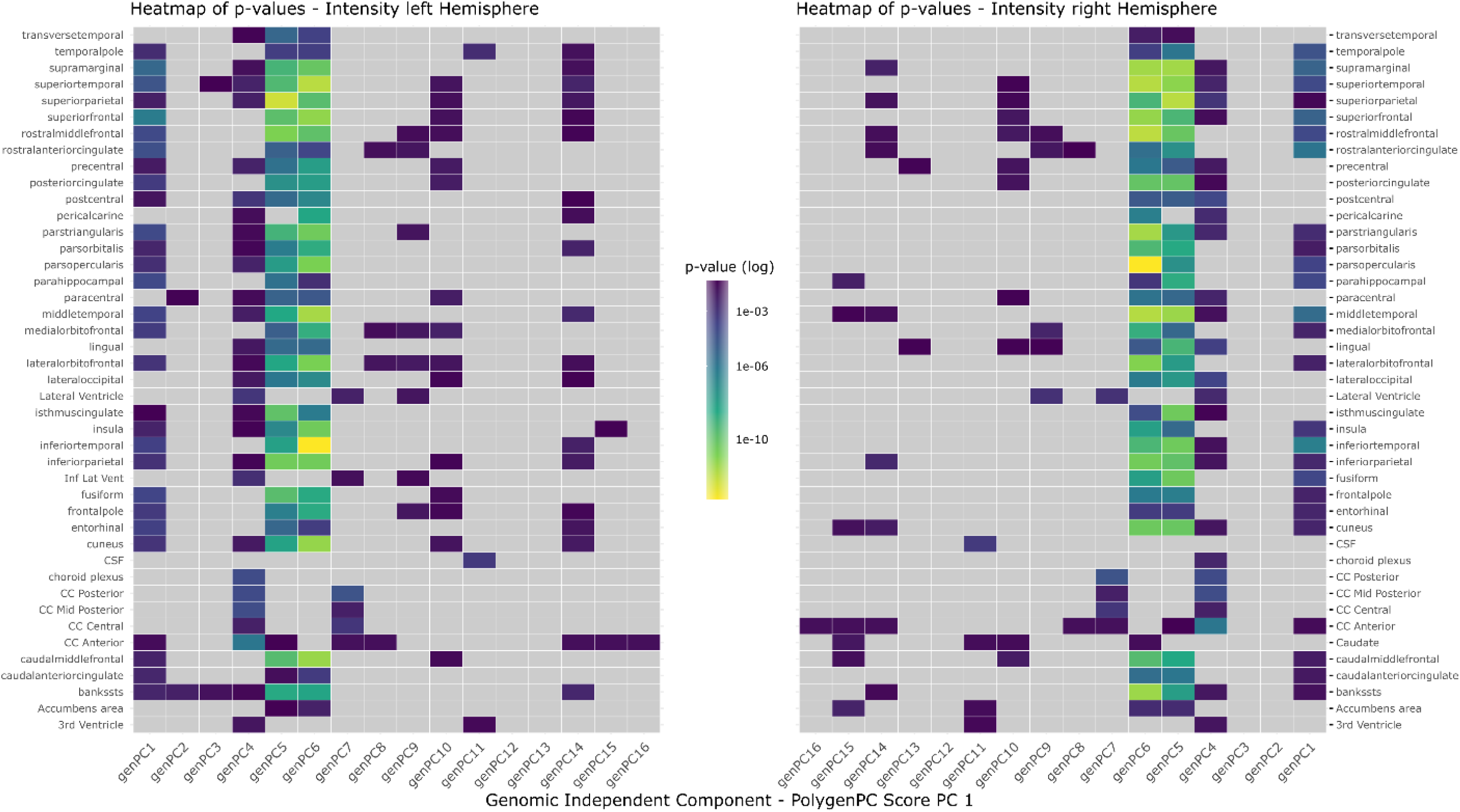
Heatmap indicating the power of association of genomic PCs with IDPs of intensity. P-values are shown from high (blue) to low (yellow). Grey are non-significant associations post-MCC. Heatmaps are lateralized, with the left heatmap indicating the left hemisphere, and the right heatmap the right hemisphere, with the component order mirrored. If components are missing then they have no significant associations with any phenotype in this modality pre-MCC.

**Figure 92:**
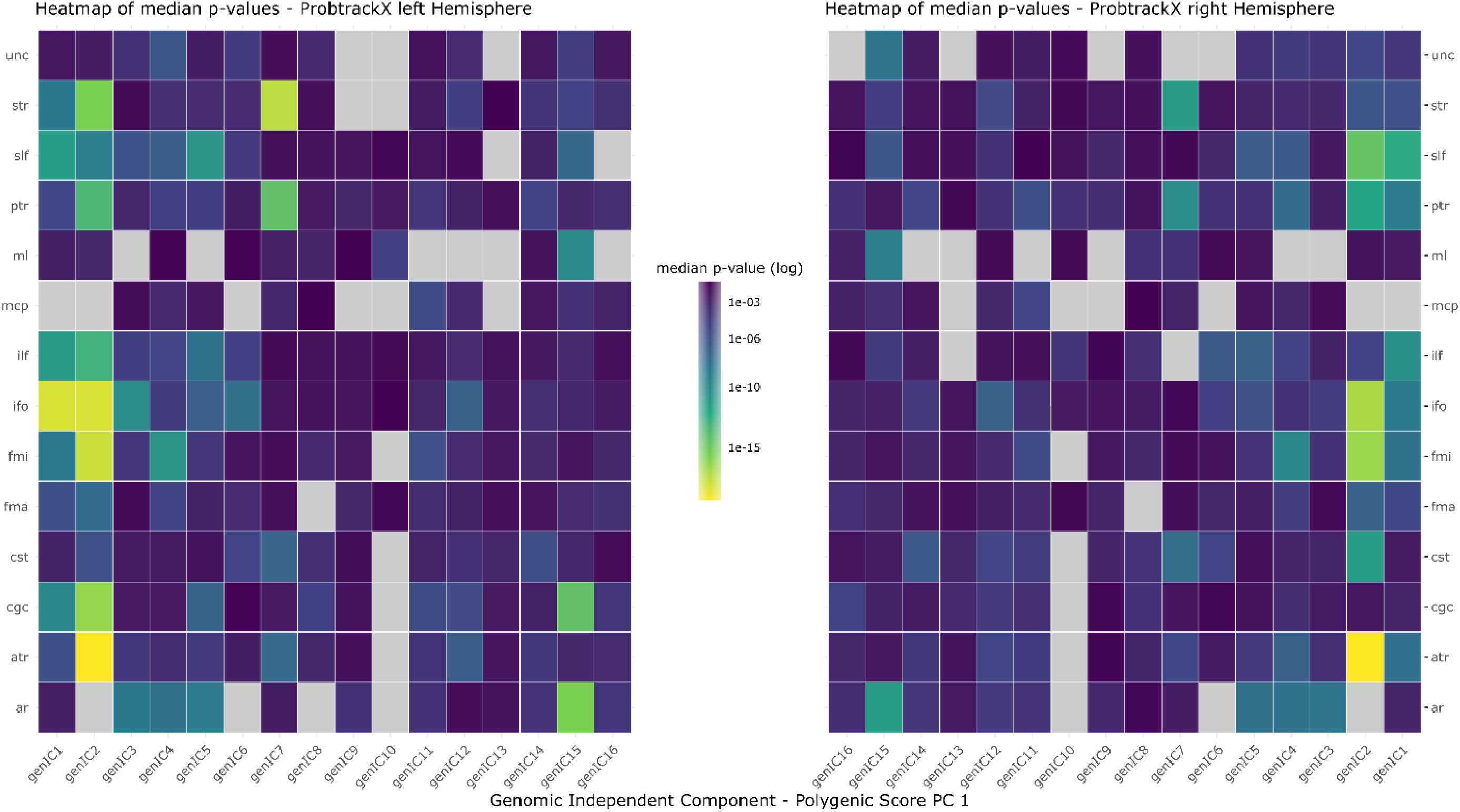
Heatmap indicating the power of association of genomic PCs with IDPs of dMRI ProbtrackX. P-values are shown from high (blue) to low (yellow). P-values have been aggregated into one median p-value across ProbtrackX modalities (e.g. MD, AD, FA, L1, etc.) to reflect the association power with white matter tracts. Grey are non-significant associations post-MCC. Heatmaps are lateralized, with the left heatmap indicating the left hemisphere, and the right heatmap the right hemisphere, with the component order mirrored. If components are missing then they have no significant associations with any phenotype in this modality.

**Figure 93:**
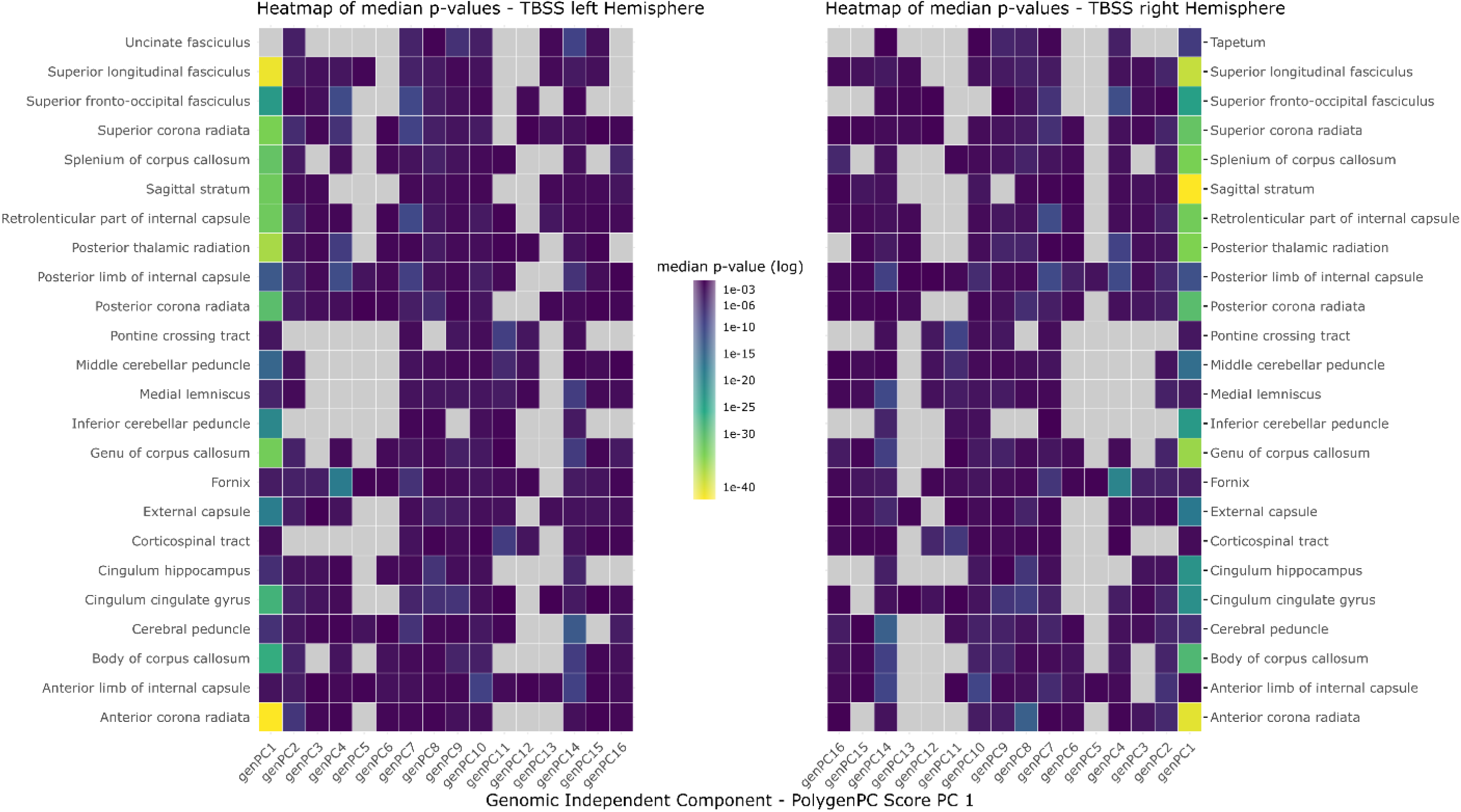
Heatmap indicating the power of association of genomic ICs with IDPs of dMRI TBSS. P-values are shown from high (blue) to low (yellow). P-values have been aggregated into one median p-value across TBSS modalities (e.g. MD, AD, FA, L1, etc.) to reflect the association power with white matter tracts. Grey are non-significant associations post-MCC. Heatmaps are lateralized, with the left heatmap indicating the left hemisphere, and the right heatmap the right hemisphere, with the component order mirrored. If components are missing then they have no significant associations with any phenotype in this modality.

**Figure 94:**
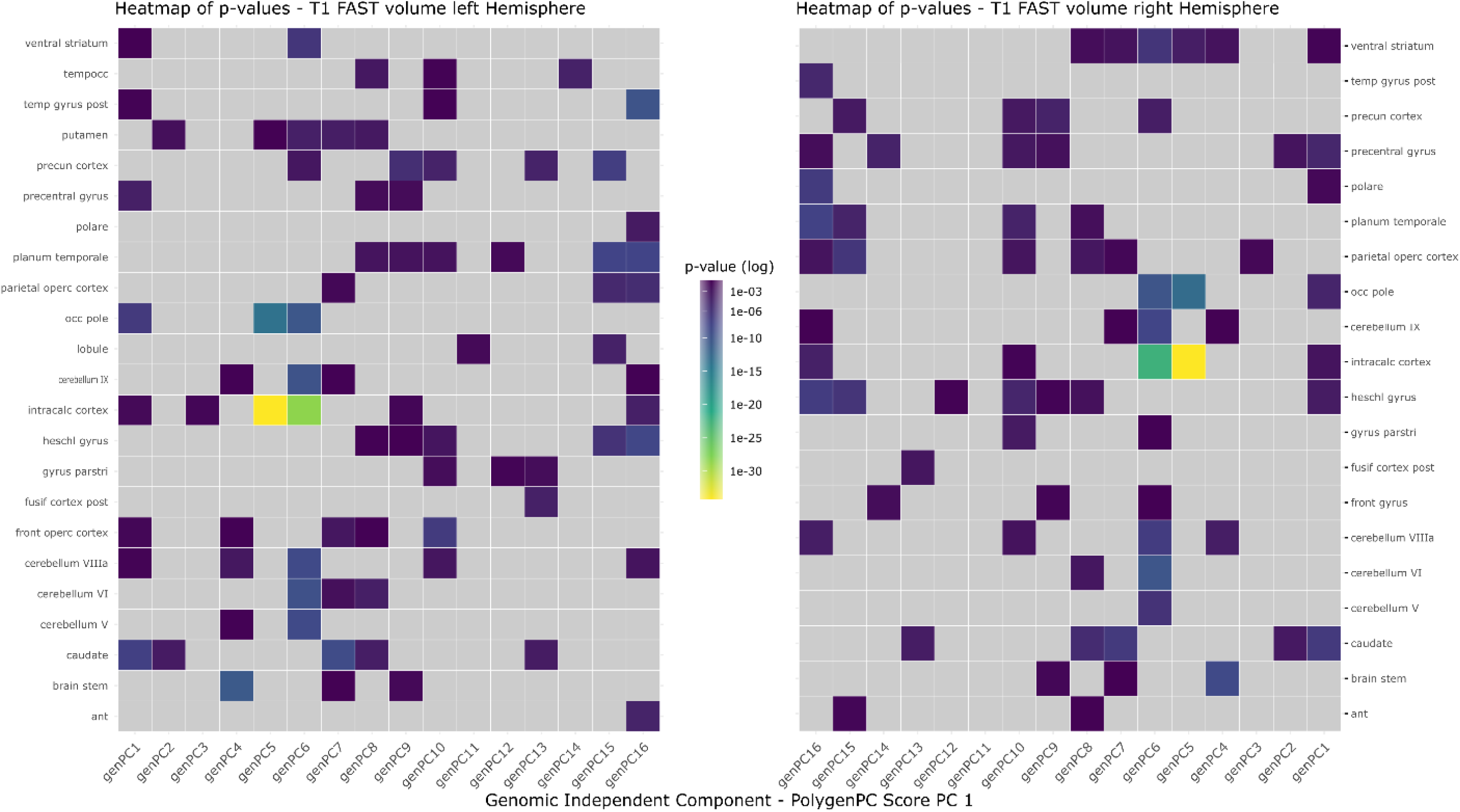
Heatmap indicating the power of association of genomic PCs with IDPs of T1-FAST volume. P-values are shown from high (blue) to low (yellow). Grey are non-significant associations post-MCC. Heatmaps are lateralized, with the left heatmap indicating the left hemisphere, and the right heatmap the right hemisphere, with the component order mirrored. If components are missing then they have no significant associations with any phenotype in this modality.

**Figure 95:**
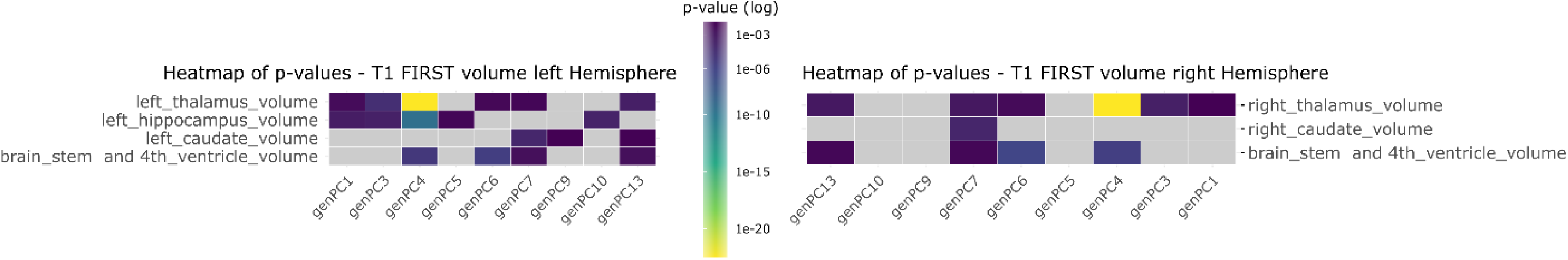
Heatmap indicating the power of association of genomic ICs with IDPs of T1-FIRST volume. P-values are shown from high (blue) to low (yellow). Grey are non-significant associations post-MCC. Heatmaps are lateralized, with the left heatmap indicating the left hemisphere, and the right heatmap the right hemisphere, with the component order mirrored. If components are missing then they have no significant associations with any phenotype in this modality.

**Figure 96:**
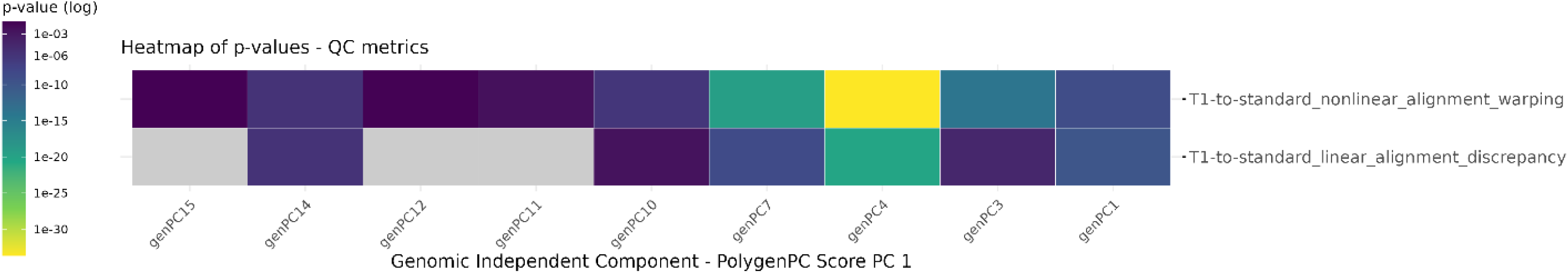
Heatmap indicating the power of association of genomic PCs with IDPs of QC metrics. P-values are shown from high (blue) to low (yellow). Grey are non-significant associations post-MCC. If components are missing then they have no significant associations with any phenotype in this modality

## References

1. Elliott, L. T. et al. Genome-wide association studies of brain imaging phenotypes in UK Biobank. Nature 562, 210–216 (2018).

2. Eyler, L. T. et al. Genetic and Environmental Contributions to Regional Cortical Surface Area in Humans: A Magnetic Resonance Imaging Twin Study. Cereb. Cortex N. Y. NY 21, 2313– 2321 (2011).

3. Glahn, D. C. et al. Genetic control over the resting brain. Proc. Natl. Acad. Sci. 107, 1223– 1228 (2010).

4. Grasby, K. L. et al. The genetic architecture of the human cerebral cortex. Science 367, eaay6690 (2020).

5. Jahanshad, N. et al. Multi-site genetic analysis of diffusion images and voxelwise heritability analysis: A pilot project of the ENIGMA–DTI working group. NeuroImage 81, 455–469 (2013).

6. McKay, D. R. et al. Influence of age, sex and genetic factors on the human brain. Brain Imaging Behav. 8, 143–152 (2014).

7. Pizzagalli, F. et al. The reliability and heritability of cortical folds and their genetic correlations across hemispheres. Commun. Biol. 3, 1–12 (2020).

8. Smith, S. M. et al. An expanded set of genome-wide association studies of brain imaging phenotypes in UK Biobank. Nat. Neurosci. 24, 737–745 (2021).

9. Sprooten, E. et al. Common genetic variants and gene expression associated with white matter microstructure in the human brain. NeuroImage 97, 252–261 (2014).

10. Watanabe, K. et al. A global overview of pleiotropy and genetic architecture in complex traits. Nat. Genet. 51, 1339–1348 (2019).

11. Winkler, A. M. et al. Cortical thickness or grey matter volume? The importance of selecting the phenotype for imaging genetics studies. NeuroImage 53, 1135–1146 (2010).

12. Hibar, D. P. et al. Novel genetic loci associated with hippocampal volume. Nat. Commun. 8, 13624 (2017).

13. Matoba, N., Love, M. I. & Stein, J. L. Evaluating brain structure traits as endophenotypes using polygenicity and discoverability. Hum. Brain Mapp. 43, 329–340 (2022).

14. Tissink, E. P. et al. Abundant pleiotropy across neuroimaging modalities identified through a multivariate genome-wide association study. Nat. Commun. 15, 2655 (2024).

15. Wendt, F. R., Pathak, G. A., Tylee, D. S., Goswami, A. & Polimanti, R. Heterogeneity and Polygenicity in Psychiatric Disorders: A Genome-Wide Perspective. Chronic Stress 4, 2470547020924844 (2020).

16. Allegrini, A. G. et al. Genomic prediction of cognitive traits in childhood and adolescence. Mol. Psychiatry 24, 819–827 (2019).

17. Demontis, D. et al. Discovery of the first genome-wide significant risk loci for attention deficit/hyperactivity disorder. Nat. Genet. 51, 63–75 (2019).

18. Karlsson Linnér, R., et al. Multivariate analysis of 1.5 million people identifies genetic associations with traits related to self-regulation and addiction. Nat. Neurosci. 24, 1367– 1376 (2021).

19. Lee, J. J., McGue, M., Iacono, W. G. & Chow, C. C. The accuracy of LD Score regression as an estimator of confounding and genetic correlations in genome-wide association studies. Genet. Epidemiol. 42, 783–795 (2018).

20. Pardiñas, A. F. et al. Common schizophrenia alleles are enriched in mutation-intolerant genes and in regions under strong background selection. Nat. Genet. 50, 381–389 (2018).

21. Savage, J. E. et al. Genome-wide association meta-analysis in 269,867 individuals identifies new genetic and functional links to intelligence. Nat. Genet. 50, 912–919 (2018).

22. Wray, N. R. et al. Genome-wide association analyses identify 44 risk variants and refine the genetic architecture of major depression. Nat. Genet. 50, 668–681 (2018).

23. Sprooten, E., Franke, B. & Greven, C. U. The P-factor and its genomic and neural equivalents: an integrated perspective. Mol. Psychiatry 27, 38–48 (2022).

24. Choi, S. W., Mak, T. S.-H. & O’Reilly, P. F. Tutorial: a guide to performing polygenic risk score analyses. Nat. Protoc. 15, 2759–2772 (2020).

25. Choi, S. W. & O’Reilly, P. F. PRSice-2: Polygenic Risk Score software for biobank-scale data. GigaScience 8, giz082 (2019).

26. Dudbridge, F. Power and Predictive Accuracy of Polygenic Risk Scores. PLoS Genet. 9, e1003348 (2013).

27. International Schizophrenia Consortium et al. Common polygenic variation contributes to risk of schizophrenia and bipolar disorder. Nature 460, 748–752 (2009).

28. Wray, N. R. et al. Research review: Polygenic methods and their application to psychiatric traits. J. Child Psychol. Psychiatry 55, 1068–1087 (2014).

29. Rodrigue, A. L. et al. Specificity of Psychiatric Polygenic Risk Scores and Their Effects on Associated Risk Phenotypes. Biol. Psychiatry Glob. Open Sci. 3, 519–529 (2023).

30. Grotzinger, A. D. et al. Genomic structural equation modelling provides insights into the multivariate genetic architecture of complex traits. Nat. Hum. Behav. 3, 513–525 (2019).

31. Fürtjes, A. E. et al. General dimensions of human brain morphometry inferred from genome-wide association data. Hum. Brain Mapp. 44, 3311–3323 (2023).

32. van der Meer, D. et al. Understanding the genetic determinants of the brain with MOSTest. Nat. Commun. 11, 3512 (2020).

33. Lazarev, D., Chau, G., Bloemendal, A., Churchhouse, C. & Neale, B. M. GUIDE deconstructs genetic architectures using association studies. 2024.05.03.592285 Preprint at 10.1101/2024.05.03.592285 (2024).

34. Oblong, L. M. et al. Principal and independent genomic components of brain structure and function. Genes Brain Behav. 23, e12876 (2024).

35. Comon, P. Independent component analysis, A new concept? Signal Process. 36, 287–314 (1994).

36. Liu, S., Smit, D. J. A., Abdellaoui, A., van Wingen, G. A. & Verweij, K. J. H. Brain Structure and Function Show Distinct Relations With Genetic Predispositions to Mental Health and Cognition. Biol. Psychiatry Cogn. Neurosci. Neuroimaging 8, 300–310 (2023).

37. Pan, X. et al. Exploring the genetic correlation between obesity-related traits and regional brain volumes: Evidence from UK Biobank cohort. NeuroImage Clin. 33, 102870 (2022).

38. Peters, S. A. E., Singhateh, Y., Mackay, D., Huxley, R. R. & Woodward, M. Total cholesterol as a risk factor for coronary heart disease and stroke in women compared with men: A systematic review and meta-analysis. Atherosclerosis 248, 123–131 (2016).

39. Yanai, H. et al. An Improvement of Cardiovascular Risk Factors by Omega-3 Polyunsaturated Fatty Acids. J. Clin. Med. Res. 10, 281–289 (2018).

40. Saher, G. et al. High cholesterol level is essential for myelin membrane growth. Nat. Neurosci. 8, 468–475 (2005).

41. Dietschy, J. M. Central nervous system: cholesterol turnover, brain development and neurodegeneration. Biol. Chem. 390, 287–293 (2009).

42. Debette, S. & Markus, H. S. The clinical importance of white matter hyperintensities on brain magnetic resonance imaging: systematic review and meta-analysis. BMJ 341, c3666 (2010).

43. Verhaaren, B. F. J. et al. High Blood Pressure and Cerebral White Matter Lesion Progression in the General Population. Hypertension 61, 1354–1359 (2013).

44. Gallucci, G., Tartarone, A., Lerose, R., Lalinga, A. V. & Capobianco, A. M. Cardiovascular risk of smoking and benefits of smoking cessation. J. Thorac. Dis. 12, 3866–3876 (2020).

45. Bonner, W. I. A. et al. Determinants of self-perceived health for Canadians aged 40 and older and policy implications. Int. J. Equity Health 16, 94 (2017).

46. Cialani, C. & Mortazavi, R. The effect of objective income and perceived economic resources on self-rated health. Int. J. Equity Health 19, 196 (2020).

47. Wang, N. et al. Perceived Health as Related to Income, Socio-economic Status, Lifestyle, and Social Support Factors in a Middle-aged Japanese. J. Epidemiol. 15, 155–162 (2005).

48. Watanabe, K., Taskesen, E., van Bochoven, A. & Posthuma, D. Functional mapping and annotation of genetic associations with FUMA. Nat. Commun. 8, 1826 (2017).

49. de Leeuw, C. A., Mooij, J. M., Heskes, T. & Posthuma, D. MAGMA: Generalized Gene-Set Analysis of GWAS Data. PLOS Comput. Biol. 11, e1004219 (2015).

50. Trevisan, N. et al. The Genetic Architecture of Brain Structure and Function: A Data-Driven Interpretation Using genomICA. 2025.04.07.25324950 Preprint at 10.1101/2025.04.07.25324950 (2025).

51. Morita, J. et al. Structure and biological function of ENPP6, a choline-specific glycerophosphodiester-phosphodiesterase. Sci. Rep. 6, 20995 (2016).

52. Dal Lin, C., Tona, F. & Osto, E. The crosstalk between the cardiovascular and the immune system. Vasc. Biol. 1, H83–H88 (2019).

53. Alfaro-Almagro, F. et al. Image processing and Quality Control for the first 10,000 brain imaging datasets from UK Biobank. NeuroImage 166, 400–424 (2018).

54. Bycroft, C. et al. The UK Biobank resource with deep phenotyping and genomic data. Nature 562, 203–209 (2018).

55. Shi, Y., Franke, B., Mota, N. R. & Sprooten, E. Genetic liability to major psychiatric disorders contributes to multi-faceted quality of life outcomes in children and adults. 2023.01.17.23284645 Preprint at 10.1101/2023.01.17.23284645 (2023).

56. Watanabe, K. et al. A global overview of pleiotropy and genetic architecture in complex traits. Nat. Genet. 51, 1339–1348 (2019).

57. Chang, C. C. et al. Second-generation PLINK: rising to the challenge of larger and richer datasets. GigaScience 4, s13742–015−0047–8 (2015).

58. Beckmann, C. F. & Smith, S. M. Probabilistic independent component analysis for functional magnetic resonance imaging. IEEE Trans. Med. Imaging 23, 137–152 (2004).

59. Jenkinson, M., Beckmann, C. F., Behrens, T. E. J., Woolrich, M. W. & Smith, S. M. FSL. NeuroImage 62, 782–790 (2012).

60. Coombes, B. J., Ploner, A., Bergen, S. E. & Biernacka, J. M. A principal component approach to improve association testing with polygenic risk scores. Genet. Epidemiol. 44, 676–686 (2020).

61. Ge, T., Chen, C.-Y., Ni, Y., Feng, Y.-C. A. & Smoller, J. W. Polygenic prediction via Bayesian regression and continuous shrinkage priors. Nat. Commun. 10, 1776 (2019).

62. Mak, T. S. H. et al. Polygenic scores via penalized regression on summary statistics. Genet. Epidemiol. 41, 469–480 (2017).

